# Burden of Non-Communicable Diseases Attributable to Polygenic Risk and Its Association with Healthcare Utilization in Taiwan

**DOI:** 10.64898/2026.10.07.26365010

**Authors:** Mu-Yi Shih, Yi-Syuan Ke, Xue-Yong Chang, Ling-Hua Wang, Yen-Chen Anne Feng

**Author notes:** Corresponding author: Yen-Chen A. Feng, Rm. 520, No. 17 Xuzhou Rd, Zhongzheng Dist, Taipei 100, Taiwan.

## Abstract

**Background:** Non-communicable diseases (NCDs) impose substantial population burden. Their polygenic architecture informs early risk stratification through polygenic risk scores (PRS). Previous studies reported elevated-PRS burden comparable to some modifiable risk factors (MRFs), yet evidence remains confined to European populations. We quantified the PRS-attributable NCD burden in Taiwan.

**Methods:** Linking Taiwan Biobank and National Health Insurance Research Database (N ≈ 140K), we analyzed 42 high-burden NCDs. Elevated genetic risk (*PRS_e_*; top 10%) was assessed using Cox models. We estimated population attributable fractions (PAFs) and disability-adjusted life years (DALYs) using Taiwan-specific estimates from Global Burden of Disease 2023, and examined associations between an integrated PRS (iPRS) and healthcare utilization.

**Findings:** Strongest *PRS_e_* associations occurred for atrial fibrillation (AFib; HR = 3.56 [95% CI: 3.23-3.92]) and gout (2.41 [2.33-2.49]), yielding PAFs of 20% and 12%, respectively, while population-level attributable DALYs were largest for type 2 diabetes (T2D), intracranial hemorrhage (ICH), and lung cancer. At the individual level, *PRS_e_* corresponded to 1.50 [1.42-1.57] healthy life-years lost for T2D and 0.57 [0.40-0.76] for ICH, approaching 2.4 years for the top 1% PRS. Polygenic attributable burden was overall greater in men. For several diseases, *PRS_e_*-associated PAF and DALYs exceeded those of individual MRFs (e.g., smoking, alcohol use). Top-decile iPRS was linked to 23.9% (15.0-33.6%) more hospital days, 19.1% (13.2-25.3%) more inpatient visits, and 9.6% (6.9-12.4%) more outpatient visits.

**Interpretation:** Polygenic risk accounts for a measurable—and for several diseases, MRF-comparable—share of NCD burden in Taiwan, providing a quantitative foundation for integrating genetic liability into preventive care and health planning.

**Funding:** National Taiwan University Academic Research-Career Development Project, Yushan Young Fellow Program from the Ministry of Education (MoE), and the Population Health Research Center from Featured Areas Research Center Program within the framework of the Higher Education Sprout Project by the MoE in Taiwan.

## Introduction

Chronic, non-communicable diseases (NCDs) impose a substantial burden on individuals and society, leading to chronic pain, disability, and even premature death.^1,2^ These conditions significantly diminish quality of life and their long-term management remains a primary driver of healthcare utilization and costs.^3–6^ Most of these high burden diseases (e.g., type 2 diabetes [T2D], ischemic heart disease, and breast cancer) are complex and polygenic, with genome-wide association studies (GWAS) identifying hundreds to thousands of genomic risk loci that influence disease susceptibility.^7–10^ These findings have enabled the development of polygenic risk scores (PRS)—which quantify an individual’s lifetime genetic liability to a disease—for early identification of high-risk individuals.^11–13^ As GWAS samples sizes grow and PRS become more powerful, studies have demonstrated promising risk-stratification utility for diseases like atrial fibrillation [AFib], T2D, and asthma (e.g., AFib: a 4.63-fold elevated risk linked to the top 1% of the PRS distribution vs. the remaining 99%; T2D: a 7-fold increased risk associated with the top 2.5% PRS vs. individuals in the IQR).^11,14^

In contrast to the extensive literature on PRS-based risk prediction, only a handful of studies have assessed the extent to which PRS contributes to disease burden at the population level.^15–18^ Such burden quantification typically follows a comparative risk assessment approach grounded in the counterfactual framework—utilizing metrics like the population attributable fraction [PAF])—which has historically been applied to modifiable risk factors (MRFs; e.g., diet, smoking).^19–22^ The most prominent example is the Global Burden of Disease (GBD) study, which estimates disease burden— typically measured in disability-adjusted life years (DALYs)—attributable to MRFs such as smoking and high body mass index across 204 countries and territories, thereby informing global and national public health strategy.^23^ Given that most high-burden chronic diseases are substantially polygenic, genetic risk represents a notable omission from this framework. While genetic risk factors, unlike MRFs, cannot be altered, transforming individual genetic liability into population-level burden estimates have the potential to inform health-system planning (e.g., resource allocation, targeted screening). A few recent studies leveraging population-scale biobank data have found that the attributable effect of common genetic variants was comparable to that of established MRFs for certain diseases.^16,24,25^ These findings, however, derive almost entirely from European-ancestry cohorts, leaving the polygenic attributable burden in other populations largely understudied.

To address this gap, the population-based Taiwan biobank (TWB; ∼140,000 individuals), linked to the longitudinal National Health Insurance Research Database (NHIRD), offers a unique opportunity to characterize Taiwan-specific polygenic associations with disease onset and their impact on overall disease burden.^26,27^ Furthermore, the detailed healthcare utilization data within the TWB-NHIRD cohort provide an additional dimension for assessing the downstream consequences of disease burden on the Taiwanese healthcare system. In this study, we investigated 42 selected chronic, high-burden NCDs in Taiwan and examined the disease burden attributable to high polygenic risk (via PAF and DALYs), its magnitude relative to that of established MRFs, and finally, the association of an integrated polygenic risk index spanning these conditions with healthcare utilization (e.g., inpatient admissions and outpatient visits)—together aiming to provide translational insights into genetically-guided preventive care planning.

## Materials and Methods

### The Taiwan Biobank

The Taiwan Biobank (TWB) is a prospective population study with phenotype and genetic data collected in the Han-Chinese-based cohort commenced in 2012.^26^ Over 150,000 cancer-free men and women, aged between 20-70 years old, were enrolled. The participants completed comprehensive questionnaires, underwent thorough physical examinations and provided urine and blood samples for genomic and other omic data generation. Whole-genome genotype data of the TWB was generated using the customized TWBv1 and TWBv2 arrays, considered as two distinct batches due to their low overlap in marker content. Quality control (QC) of genotype data was completed separately for each batch, following procedures described elsewhere (**Supplementary Methods**).^26^ Our final analytic sample consists of 27,266 individuals in batch 1 and 117,259 individuals in batch 2; variants were mapped to the GRCh37 genome build.

### The National Health Insurance Research database

The National Health Insurance Research Database (NHIRD) is a database of longitudinal claims data and electronic health records (EHR) constructed from the mandatory National Health Insurance (NHI) system that covers almost all 23 million residents in Taiwan. Established from the year of 2000, the NHIRD database contains rich, longitudinal phenotype data such as inpatient hospital care, outpatient visits, diagnosis codes, medical treatment, and drug prescriptions, providing an invaluable resource for population-based health research in Taiwan.^27^

### Linkage between TWB and NHIRD: disease definitions

To obtain detailed clinical data of TWB participants, we linked the QC’ed 140K genotyped individuals to the NHIRD, retrieving data from 2000 to 2023. We mapped the original ICD-9-CM and ICD-10-CM codes into PheCodes to define diseased individuals (i.e., cases) for each target disease, requiring at least one qualifying inpatient record or at least two outpatient records (**Table S1**).^28^ Controls for each disease were defined as individuals not meeting the case criteria, additionally limited to those without related medical conditions per target disease following the standard PheCode exclusion criteria in the PheWAS R package.^29^ We retained the PheCode naming conventions for describing diseases throughout the study.

### DALYs estimates

To measure disease burden, we obtained Taiwan-specific DALY estimates from the GBD 2023 study. One DALY represents one lost year of healthy life due to illness, disability, or premature death. It is the summation of years of life lost due to premature mortality (YLLs) and years lived with disability (YLDs).^23^ We extracted these estimates stratified by disease and sex.

### Disease selection and collecting GWAS summary statistics

We selected target diseases for analysis based on three criteria: high disease burden in Taiwan based on DALYs, a clear polygenic basis, and sufficient data availability. In total, we analyzed 42 NCDs (e.g., breast cancer, T2D, and intracranial hemorrhage [ICH]), which collectively account for 50% of the total burden of disease in Taiwan according to the GBD 2023 data. For each disease, we obtained GWAS summary statistics from large-scale published studies conducted in European (EUR), East-Asian (EAS) or multi-ancestry (MULTI) populations. In total, we collected 131 GWAS summary statistics published from 2017 to 2025. The average sample sizes are 422,942 (range: 59,945-2,535,601) for MULTI, 460,774 (range: 10,240-1,503,898) for EUR, and 185,037 (range: 29,849-349,359) for EAS. More details can be found in **Table S2**. GWAS summary statistics were harmonized to the GRCh37 build by lifting over variants from GRCh38 where necessary.

### Polygenic risk scores (PRS)

To quantify the polygenic susceptibility of each selected disease, we computed PRS for each participant in the TWB. PRS estimates an individual’s genetic liability for a given disease, calculated as a sum of risk alleles across genome-wide single nucleotide polymorphisms (SNPs) weighted by their effect sizes derived from the collected GWAS summary statistics.^13^

To compute PRS, we employed PRS-CS which utilizes a high-dimensional Bayesian regression framework.^30^ We leveraged the 1000 Genomes Project East Asian samples as the reference panel for TWB.^31^ For each disease and batch separately, PRS were then calculated as a weighted sum using the posterior effect estimates via PLINK --scoresum.^32^ Where possible, we also leveraged PRS-CSx, a cross-population method that integrates EUR and EAS GWAS summary statistics, to enhance polygenic prediction.^33^ All PRSes were calculated and subsequently standardized to have zero mean and unit variance for downstream analyses.

### Survival analyses

To assess whether high PRS is linked to an increased likelihood of high-burden diseases over time, we performed Cox proportional hazard regression analysis with the "survival" R package,^34^ using age as the time scale. For cases, time-to-event was defined as the age at first diagnosis recorded in the NHIRD. For controls, individuals were right censored at the end of study (2023) or at death, whichever occurred first. In our primary analysis, the continuous PRS was dichotomized into the top 10% (*PRS_e_*) versus the remaining 90%, representing the high- and the low-risk group. A hazard ratio (HR) of *PRS_e_* was estimated while adjusting for sex, birthyear, genotyping batch, and the first 20 principal components of genetic ancestry.

Sex-stratified analyses were performed to examine potential differences in PRS effects between males and females. Sensitivity analyses were conducted using alternative PRS cutoffs, including the top 5% vs. the bottom 95%, and the top 1% PRS vs. the bottom 99%.

To maximize statistical power for downstream analyses, we prioritized one PRS per disease yielding the strongest effect size among those with significant associations, most derived from an ancestry-matched or multi-ancestry discovery GWAS (**Table S2**). For a subset of diseases, PRS were derived from GWAS summary statistics of the Taiwan Precision Medicine Initiative (TPMI)—a multi-hospital-based genomic cohort of 565,390 participants.^35^; for these, we removed samples overlapping between TPMI and our target cohort prior to survival analyses to prevent effect inflation.^35^

### Population attributable fraction (PAF)

To evaluate the effect of PRS on a population level, we calculated PAF, which represents the proportion of disease cases that could be theoretically prevented had the exposure to a given risk factor be eliminated in the population.^36^ Using the HR estimates from the Cox regression analyses, we estimated PAF attributable to polygenic risk using the following equation:^37^

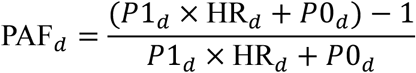

where *P*1*_d_* and *P*0*_d_* stands for the proportion of individuals in the high-risk and low-risk groups for disease *d*, here corresponding to the top 10% (*PRS_e_* ) and bottom 90% standardized PRS, respectively. *HR_d_* represents the HR for each disease *d* , estimated from the time-to-event association analyses. The 95% confidence interval of PAF were estimated using 1000-fold bootstrapping.

### Population and individual attributable DALYs

After calculating PAF, we quantified the expected loss of healthy life years attributable to polygenic risk at both the population and individual levels, following the approach proposed previously:^16^

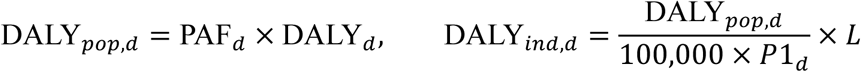

where *DALY_d_* is estimated from the GBD 2023 and represents the total DALYs per 100,000 individuals per year (annual rate) in Taiwan for each disease *d* .^23^ DALY*_pop_*_,*d*_ denotes the population-level DALYs per year attributable to the high-risk group (*PRS_e_*). DALY*_ind_*_,*d*_ represents individual-level DALYs attributable to *PRS_e_*, derived by dividing DALY*_pop_*_,*d*_ by the number of exposed individuals per 100,000 and multiplying by life expectancy (*L*) in Taiwan in 2023, thereby converting the annual rate to a lifetime estimate.^38^ The individual attributable DALYs can be interpreted as an individual’s expected reduction in healthy life years due to being in the high genetic risk group (*PRS_e_*). Attributable DALYs were additionally estimated separately by sex and across different *PRS_e_* thresholds. The 95% confidence interval of attributable DALYs were estimated using 1000-fold bootstrapping.

As TWB also records self-reported (SR) medical conditions, we repeated the Cox analysis, PAF, and DALY estimation to compare against our primary PheCode-based definitions (**Supplementary Methods**).

### Modifiable risk factors

Different from genetic risk factors, modifiable risk factors (MRFs)—such as lifestyle behaviors and environmental exposures—can be altered or controlled to reduce the risk of developing chronic diseases. To compare PAF and disease burden attributable to *PRS_e_* with those attributable to MRFs, we selected target MRFs based on their availability in the GBD 2023 study. The available MRFs included environmental (e.g., air pollution and non-optimal temperature), behavioral (tobacco and high alcohol use), and metabolic (e.g., high BMI and high systolic blood pressure) risk factors. A detailed list of available MRFs can be found in **Table S3**. Population DALY estimates for these MRFs are directly available from GBD 2023; PAF was derived as the total DALYs attributable to the risk factor for the disease divided by the total DALYs attributable to the disease.

### Joint PAF across modifiable and genetic risk factors

In addition to comparing individual PAF for each MRF against that of *PRS_e_*, we estimated a joint PAF across all available MRFs for each disease. Since simple summation of individual PAFs across risk factors can result in a total PAF exceeding 100%—as risk factors are rarely independent—we instead estimated a joint PAF by the equation below (Witte et al. 2014):

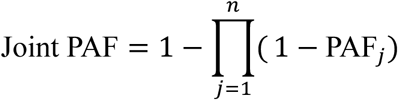

where *j* denotes each of the *n* MRFs included for a given disease. A combined joint PAF incorporating both *PRS_e_* and all MRFs was also estimated to assess their aggregate contribution to disease burden.

### Healthcare utilization and polygenic risk

Healthcare utilization reflects the real-world burden of disease on individuals’ need for medical care and the strain placed on health systems. We defined healthcare utilization using five metrics: hospitalization days, inpatient visits, outpatient visits, emergency room (ER) visits, and total visits. To relate these to overall polygenic risk, we constructed an integrated PRS (*iPR*S) across all 42 NCDs, combining their PRSs calculated from all collected GWAS summary statistics using PRSmix+, which leverages wide-spread pleiotropy to improve prediction.^39^ To estimate the mixing weights for integration, we randomly split the TWB data into training and testing sets (4:1). In the training set, we applied Elastic Net with a 5-fold cross-validation,^40^ with each healthcare utilization metric as the outcome, to learn optimal mixing weights across individual PRSs. We adjusted these weights by dividing by the standard errors of the individual PRSs estimated in the training set, then multiplied the original SNP effects by the adjusted weights to obtain final integrated per-allele effect sizes. The *iPR*S was subsequently computed in the testing set based on these effect sizes.

Within the testing set, we estimated the relative and absolute effects of the *iPR*S on each utilization metric, comparing individuals in the top 1%, 5%, 10% of the *iPR*S distribution against the remaining 99%, 95%, 90%, respectively, in separate models. As utilization counts are right-skewed and overdispersed by nature, we used negative binomial (NB) regression, with the log of the NHIRD follow-up years as an offset, allowing the modelled outcome to be interpreted as an annual rate (expected visits per year). All models were adjusted for sex, birthyear, genotyping batch and the first 20 genetic principal components. Relative effects of the top *iPRS_e_* were reported as incidence rate ratios (IRRs) with 95% CIs. From the fitted NB models, we then computed the absolute effects as the average difference in predicted annual visits between each *iPRS_e_* group and its complementary reference groups, with standard errors obtained via the delta method, using the “marginaleffects” package in R.^41^

The overall study design and analytical workflow are illustrated in **Figure S1**.

## Results

### Study cohort characteristics

Among the 141,445 individuals in the TWB-NHIRD linked cohort, age ranged from 20 to 90 years with a mean age of 49.4 years and a male-to-female ratio of 1:1.76. For the 42 selected diseases, case numbers ranged from 102 for melanomas of skin to 87,199 for back pain. The highest prevalence were observed for back pain (number of cases = 87,199, all-cohort prevalence = 61.2%), osteoarthrosis (74,180, 52.1%), hypertension (46,783, 32.8%), hearing loss (33,751, 23.7%), T2D (33,364, 23.4%), while the lowest prevalence were observed for melanomas of skin (102, 0.07%), cancer of esophagus (176, 0.12%), cervical cancer (AD; 321, 0.23%), Alzheimer’s disease (324, 0.23%), and cancer of stomach (367, 0.26%). Sex-stratified analyses revealed that males had a higher prevalence for conditions like gout, hypertension, T2D, ischemic heart disease, and chronic liver diseases and cirrhosis, whereas females had a higher prevalence of osteoarthrosis, migraine, generalized anxiety disorder, back pain, and depression (**Figure 1; Table S4**). Notably, a few diseases, such as chronic liver disease and cirrhosis (12.1%) and GERD (15.4%), showed markedly higher prevalence in TWB than observed in European-based biobanks (e.g., UK Biobank, FinnGen) and the nearby Biobank Japan (1-8%).^42–44^

**Figure 1.**
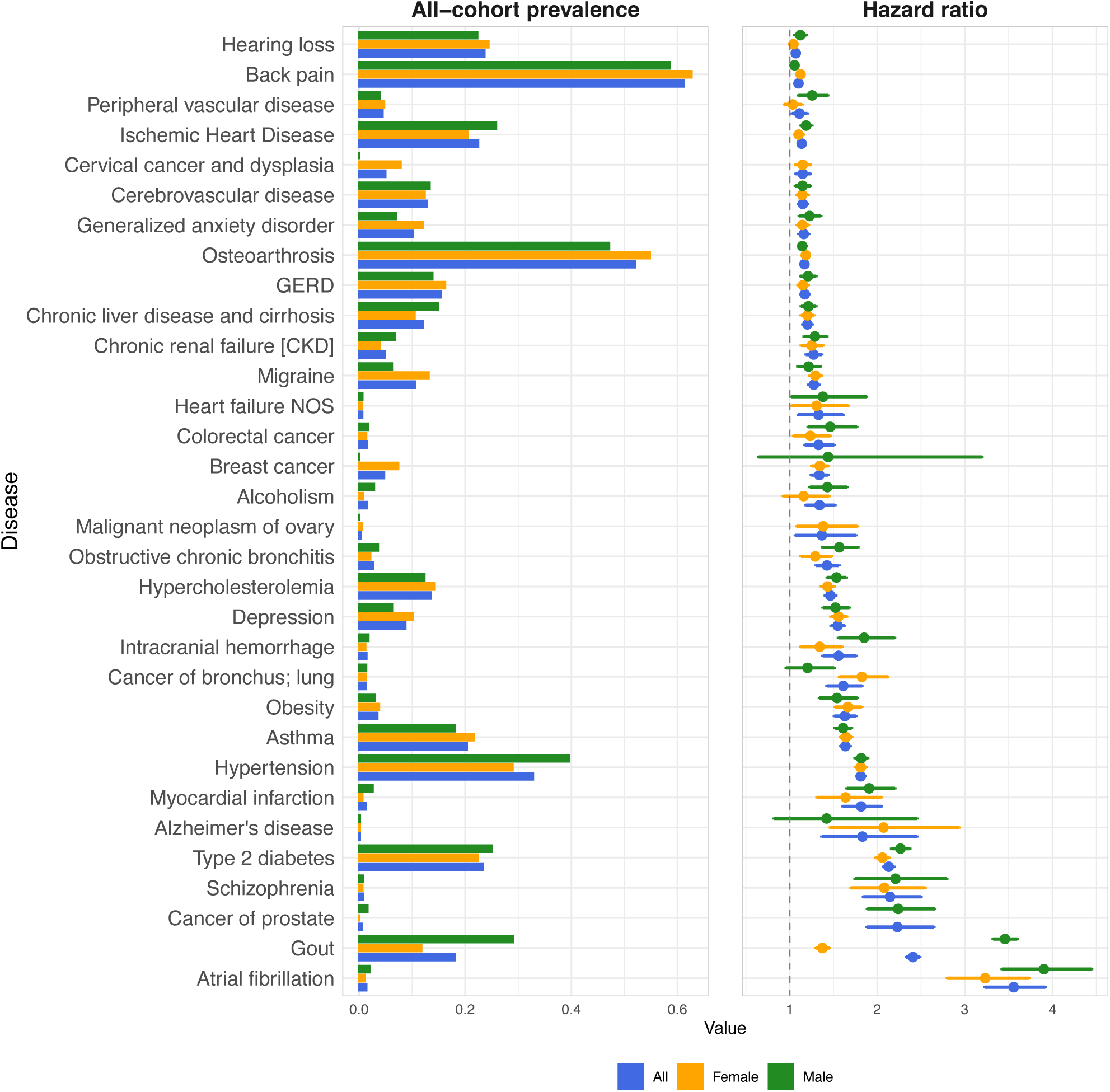
Prevalence and polygenic risk effects across selected high-burden NCDs. Disease prevalence (left) and the effect of the top 10% PRS (PRSe) on disease risk (right) are shown for the TWB-NHIRD linked cohort, overall and stratified by sex. Hazard ratios (HRs) and 95% confidence intervals (error bars) were derived from Cox proportional hazards regression with age as the time scale. Depicted are 32 of the 42 diseases with a statistically significant PRSe association (p < 0.05), ordered by the overall PRSe effect size. Disease names are labeled following the PheCode convention. Full association statistics are available in **Table S5**.

The lowest mean ages of onset was observed for schizophrenia (37.0 [SD = 12.4]), inflammatory bowel disease (39.2 [13.5]), generalized convulsive epilepsy (41.0 [14.9]), migraine (43.0 [12.2]), and back pain (43.0 [12.1]), while the highest were observed for AD (69.1 [5.58]), cancer of prostate (63.7 [8.37]), AFib (59.6 [10.6]), cancer of stomach (58.6 [10.1]), and cancer of bronchus and lung (58.1 [10.8]). Detailed descriptive epidemiology for all diseases is available in **Table S4**.

Noteworthily, the number of deaths in our analytical cohort was low (N = 2,309; 1.6%). We therefore did not estimate PAF for incidence and mortality or YLD and YLL in DALYs separately.

### Significant *PRS_e_* **effects across diseases**

To quantify the impact of the top 10% PRS (*PRS_e_*) on disease risk, we performed time-to-event analyses. Of the 42 high burden diseases, 32 exhibited a significant association with *PRS_e_* (p-value < 0.05; **Figure 1; Table S5**). The strongest *PRS_e_* effects were observed for AFib (HR = 3.56 [95% CI: 3.23-3.92]), gout (2.41 [ 2.33-2.49]), cancer of prostate (2.23 [1.88-2.65]), schizophrenia (2.15 [1.84-2.50]), and T2D (2.13 [2.07-2.19]), while the smallest effects were observed for hearing loss (1.07 [1.03-1.11]), back pain (1.10 [1.08-1.12]), peripheral vascular disease (1.11 [1.03-1.20]), ischemic heart disease (1.14 [1.10-1.18]), and cervical cancer and dysplasia (1.15 [1.07-1.24]).

As genetic architecture may differ between biological sexes, we conducted sex-specific survival analysis for all diseases. Results showed that *PRS_e_* effects were stronger in males than females for 21 of the 32 diseases. The largest sex differences were found in gout (*HR_male_* = 3.46 [3.32-3.60] vs. *HR_female_* = 1.37 [1.30-1.45]), AFib (*HR_male_* = 3.90 [3.42-4.45] vs. *HR_female_* = 3.23 [2.80-3.73]), ICH (*HR_male_* = 1.85 [1.56-2.20] vs. *HR_female_* = 1.34 [1.13-1.60]), obstructive chronic bronchitis (or chronic obstructive pulmonary disease [COPD]; *HR_male_* = 1.57 [1.38-1.78] vs. *HR_female_* = 1.29 [1.13-1.48],), and alcoholism (*HR_male_* = 1.43 [1.23-1.66] vs. *HR_female_* = 1.16 [0.93-1.45]; **Figure 1; Table S6**).

In addition to the top 10% threshold, we performed sensitivity analyses using the top 5% and top 1% PRS cutoffs (**Figure S2; Table S7**). We observed a consistent dose-response relationship, where HRs increased as the PRS thresholds became more stringent. Among individuals in the top 1% PRS, risks were elevated as high as 5.44-fold (95% CI: 4.46-6.63) for AFib, 4.62-fold (4.31-4.96) for gout, 4.44-fold (2.49-7.91) for AD, 3.27-fold (2.52-4.25) for ICH, and 2.88-fold (2.67-3.11) for asthma.

### Population attributable impact of ***PRS_e_***

To evaluate the attributable impact of elevated polygenic risk on the population level, we estimated PAF and DALYs per 100,000 individuals (DALY*_pop_*; see **Methods**). PAF quantifies the proportional contribution of polygenic risk, while DALY*_pop_* capture the full weight of these diseases in terms of years of life lost and years lived with disability due to *PRS_e_*. Due to the availability of total disease burden estimates in GBD 2023, we estimated PAF and DALY*_pop_* attributable to *PRS_e_* for 24 of the 32 diseases.

As shown in **Figure 2A**, the trend of PAF largely corresponded with that of the HR estimates. The highest PAF values for *PRS_e_* were observed for AFib (PAF = 0.20 [0.18-0.23]), gout (0.12 [0.12-0.13]), cancer of prostate (0.11 [0.08-0.14]), schizophrenia (0.10 [0.08-0.13]), and T2D (0.10 [0.10-0.11]). Similarly, for most diseases, *PRS_e_* yielded a higher PAF for male compared to female, with gout having up to 16% difference in PAF between the sexes (*PAF_male_*= 0.20 [95% CI, 0.19-0.21] vs. *PAF_female_*= 0.04 [0.03-0.04]; **Table S8**).

**Figure 2.**
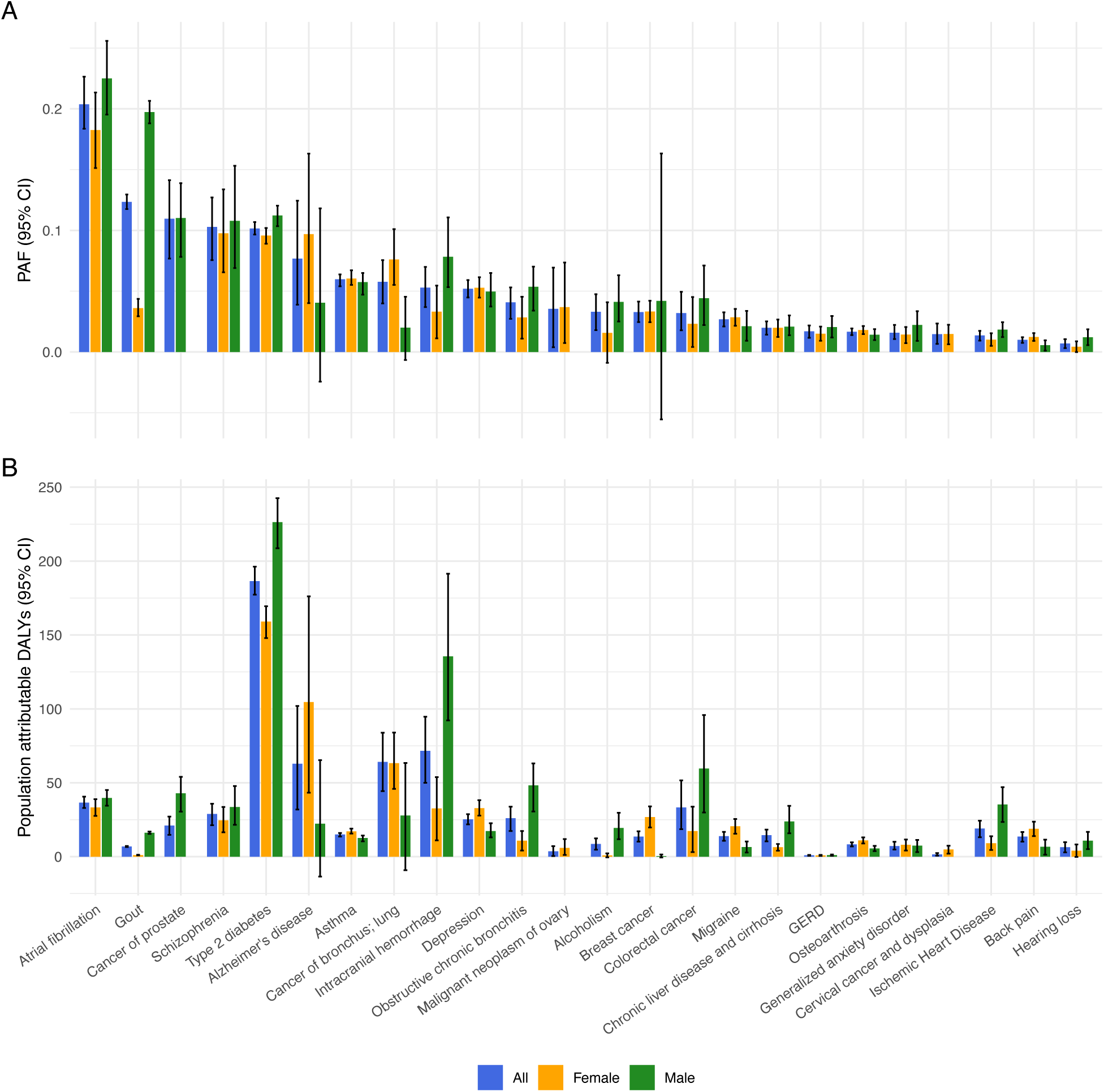
Population attributable fraction (PAF) and population attributable DALYs across selected high-burden NCDs. PAF (panel A) and population attributable DALYs (panel B) are shown for the TWB-NHIRD linked cohort, overall and stratified by sex. Estimates were derived from the HRs and DALYs estimates from the GBD 2023 study. 95% confidence intervals (error bars) were estimated using 1000-fold bootstrapping. Depicted are 24 of the 32 statistically significant PRSe-effect diseases with DALYs estimates available in the GBD 2023 study, ordered by the overall PAF estimates. Full statistics can be found in **Tables S8-S9**.

For DALY*_pop_*, *PRS_e_* contributed the most to T2D, with 186.4 [177.3-196.2] healthy life-years lost, followed by ICH (71.6 [50.0-94.7]), lung cancer (64.01 [44.4-83.9]), AD (62.9 [31.9-102.0]), and AFib (36.5 [32.9-40.6]) (**Figure 2B; Table S9**). The trend diverged from that based on HR and PAF estimates because of the incorporation of each disease’s total burden in the calculation. For example, AFib had one of the highest HR and PAF values, yet its comparatively low total DALYs resulted in modest DALY*_pop_*; in contrast, T2D did not rank highest on HR or PAF, but its substantial total burden led to the greatest DALY*_pop_*.

In sensitivity analyses using top 5% and top 1% of the PRS distribution as thresholds, population attributable impact was relatively low, reflecting the smaller exposed proportion. For example, the top 1% threshold yielded PAFs of 0.04 [0.03-0.05] for AFib and 0.02 [0.01-0.02] for T2D, with corresponding DALY*_pop_* of 7.62 [5.90-9.55] and 29.82 [26.39-33.77], respectively (**Figure S3; Tables S10-S11**).

### Individual attributable DALYs of ***PRS_e_***

Converting annual population-level DALYs to individual lifetime estimates ( DALY*_ind_* ; see **Methods**), *PRS_e_* was linked to—for the five largest contribution—1.50 [1.42-1.57] years of expected healthy life lost for T2D, 0.57 [0.40-0.76] for ICH, 0.51 [0.36-0.67] for lung cancer, 0.50 [0.26-0.82] for AD, and 0.29 [0.26-0.33] for AFib (**Figure 3A**). Sex differences also persisted: for ICH, males had notably greater DALY*_ind_* than females (1.04 [0.71-1.47] vs. 0.27 [0.09-0.45] healthy life years lost). A similarly male excess was observed for traits like T2D (male: 1.74 [1.61-1.87] vs. female: 1.33 [1.42-1.57]) and colorectal cancer (male: 0.46 [0.23-0.74] vs. female: 0.14 [0.03-0.28]). Conversely, AD (male: 0.17 [-0.10-0.50] vs. female: 0.88 [0.36-1.48]) and lung cancer (male: 0.21 [-0.07-0.49] vs. female: 0.53 [0.38-0.70]) showed a female excess.

**Figure 3.**
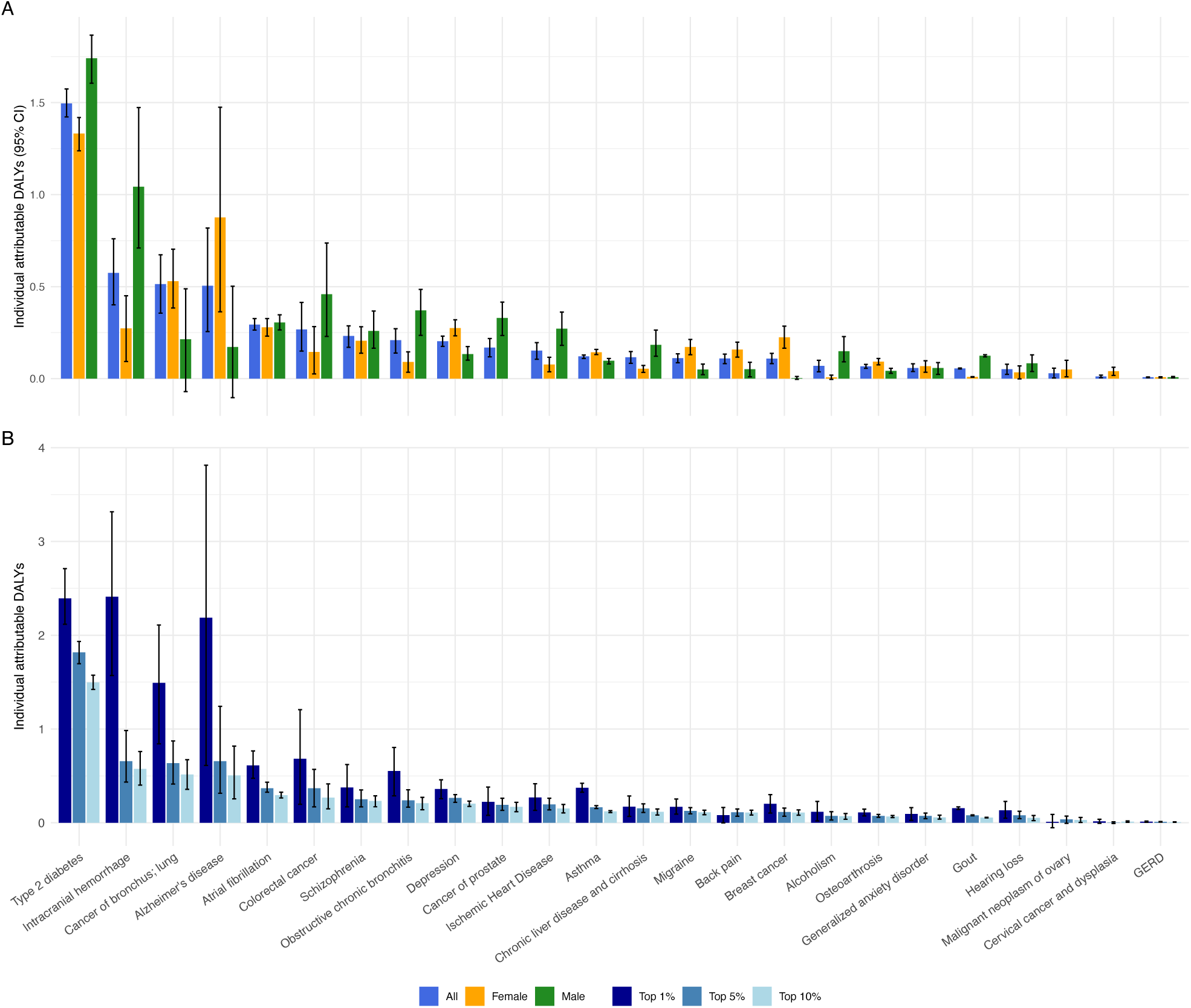
Individual attributable DALYs across selected high-burden NCDs. Inidividual attributbale DALYs are shown for the TWB-NHIRD linked cohort, overall and stratified by sex (panel A), and across different PRSe thresholds (panel B). Estimates were derived from the HRs and DALYs estimates from the GBD 2023 study. 95% confidence intervals (error bars) were estimated using 1000-fold bootstrapping. Depicted are 24 of the 32 statistically significant PRSe-effect diseases with DALYs estimates available in the GBD 2023 study, ordered by the overall individual attributable DALYs estimates. Full statistics can be found in **Tables S12-S13**.

In contrast to the population-level estimates, the top 5% and top 1% sensitivity analyses produced substantially larger values for DALY*_ind_*, consistent with the concentration of risk into a smaller group. Among individuals in the extreme genetic risk tail (top 1% PRS), the expected healthy life years lost rose to 2.41 [1.57-3.32] for ICH, 2.40 [2.12-2.71] for T2D, and 2.17 [0.61-3.81] for AD (**Figure 3B**; **Table S13**).

### Comparison between self-reported and clinical phenotyping

For 10 diseases (e.g., ICH, AFib, asthma), we compared *PRS_e_* effects from SR medical conditions against the primary PheCode-based phenotyping (**Supplementary Methods**).

Agreement was slight to moderate (Cohen’s κ 0.10-0.46; mean 0.28; **Table S14**), with the highest concordance for hypertension, and most diseases showed higher prevalence under PheCodes than SR. Interestingly, *PRS_e_* effects were generally larger under SR, particularly for gout (*HR_SR_* = 10.72 vs. *HR_P_*_ℎ*eCode*_= 2.41; *PAF_SR_* = 0.49 vs. *PAF_P_*_ℎ*eCode*_= 0.12), asthma (*HR_SR_* = 6.46 vs. 1.64; *PAF_SR_*= 0.35 vs. 0.06), and ICH (*HR_SR_* = 5.88 vs. 1.56; *PAF_SR_*= 0.33 vs. = 0.05). Conversely, AFib (*HR_SR_* = 1.45 vs. 3.56; *PAF_SR_* = 0.04 vs. 0.20) showed a larger effect under PheCode, and depression *PRS_e_* was significant only under PheCodes (**Figure S4A-4B**). Individual-level attributable DALYs diverged the most for ICH and T2D, with ∼6-fold (3.56 vs. 0.57) and ∼2-fold (2.90 vs. 1.50) more healthy life-years lost under SR (**Figure S4C**). Overall, these results indicated a non-negligible impact of phenotyping on disease burden estimates.

### Comparison between the impact of ***PRS_e_*** and MRFs

Next, we compared the impact of elevated polygenic risk with MRFs on the population level, limiting to 22 of the 24 diseases where the DALYs estimates for MRFs were available in GBD 2023. Across these diseases, the contribution of *PRS_e_* varied relative to MRFs depending on the trait. PAF and DALY*_pop_* attributable to *PRS_e_* were comparable to individual established MRFs for certain diseases (**Figure 4**; **Table S15**). For example—among the diseases with the strongest polygenic effects—the *PRS_e_* contribution for AFib (PAF = 0.20; DALY*_pop_* = 36.53) surpassed dietary risks (0.04; 7.71), high BMI (0.04; 7.34), and tobacco (0.04; 6.36), while lower than high SBP (0.28; 50.32). For cancer of prostate, a PAF of 0.11 and DALY*_pop_* of 21.06 was observed for *PRS_e_*, notably larger than tobacco (0.03; 5.56) and high alcohol use (0.01; 1.89). For T2D, *PRS_e_* (0.10; 186.44) had a lower impact than high BMI (0.50; 949.52) and dietary risks (0.26; 479.26), while on par with tobacco (0.12; 220.57) and low physical activity (0.10; 179.55) and larger than non-optimal temperature (0.03; 46.34).

**Figure 4.**
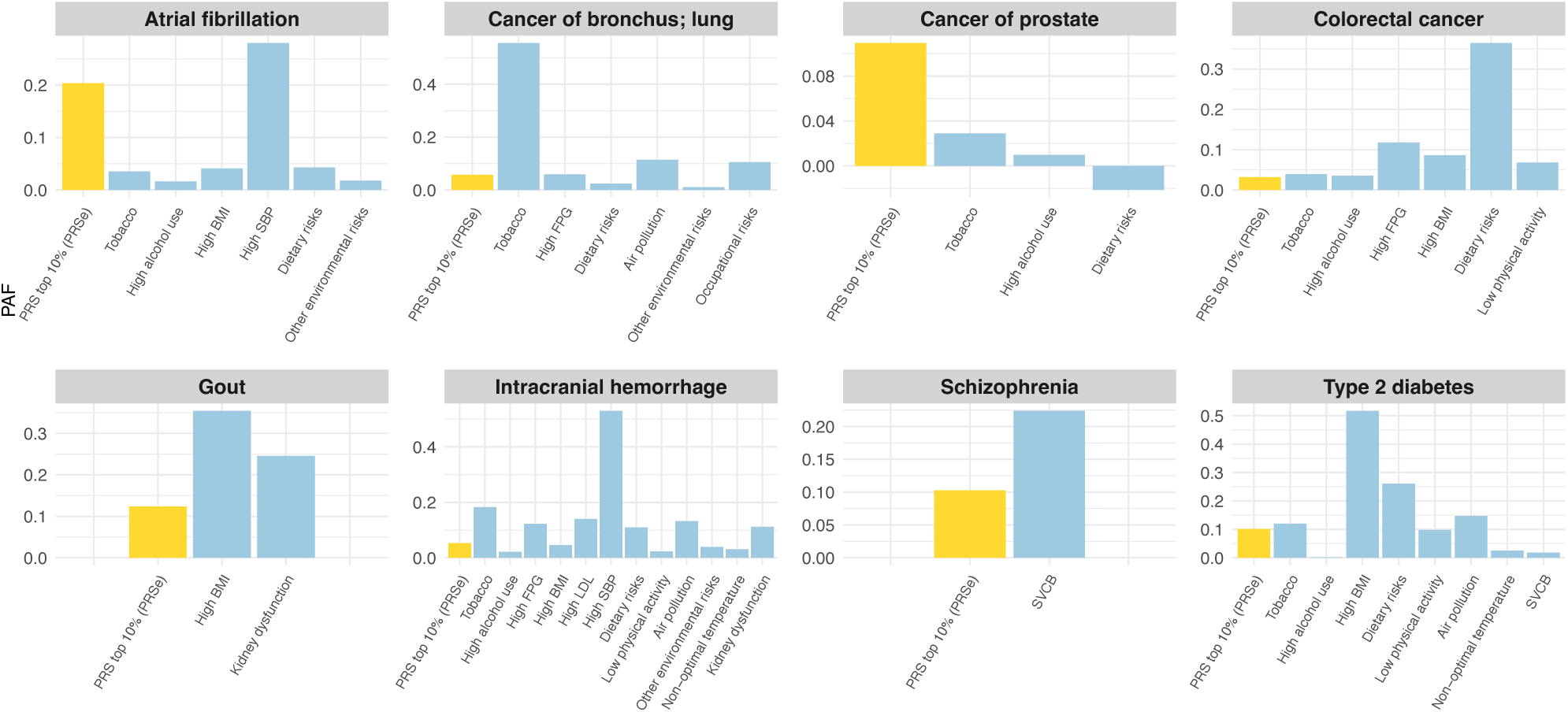
Comparison of PAF between PRSe and MRFs for selected high-burden NCDs. PAF of PRSe and MRFs are shown for the TWB-NHIRD linked cohort. Estimates of PRSe were derived from previously calculated PAFs and DALYs estimates of GBD 2023 study. For MRFs, estimates were directly used from the GBD 2023 study. Depicted are diseases with the highest PAF and/or those with the highest attributable DALYs. Full statistics across all diseases are available in **Table S15**.

Sex-specific results were overall consistent with the main analyses (**Table S15**). For AFib, the PAF of *PRS_e_* in both males and females (0.22 and 0.18 respectively) again surpassed those of dietary risks, high BMI, and tobacco (males: 0.04, 0.04 and 0.07; females: 0.04, 0.04 and 0.01), while lower than high SBP (male: 0.27, female: 0.29); for T2D, *PRS_e_* (males: 0.11, females: 0.10) maintained a lower impact than high BMI (males: 0.52, females: 0.51) and dietary risks (males: 0.28, females: 0.24), yet showing a slightly larger contribution than tobacco in females only (males: 0.16, females: 0.07).

For diseases with multiple individual MRFs, we also calculated the joint PAF of MRFs and additionally with *PRS_e_* toward a given disease. Overall, *PRS_e_* added incrementally to the joint PAF, with larger contribution for some diseases (e.g., AFib) observed than others (**Figure S5**; **Table S16**).

### Evaluation of integrated PRS on healthcare utilization

Finally, to assess the extent to which polygenic liability influences healthcare utilization patterns within the Taiwanese population, we constructed an integrated PRS ( *iPR*S ) using elastic-net regression (**Methods**). The trained mixing weights are reported in **Table S17**. As shown in **Figure 5**, individuals in the top 10% integrated PRS (*iPRS_e_*) had ∼10% more total healthcare visits compared to the reference group (IRR = 1.10 [1.07-1.13]), corresponding to an estimated 0.96 [0.67-1.25] additional visits per year. When stratified by encounter type, *iPRS_e_* was associated with ∼10% more outpatient visits (IRR = 1.10 [1.07-1.12]; +0.93 [0.65-1.21] visits/year), ∼17% more ER visits (IRR = 1.17 [1.13-1.22]; + 0.03 [0.02-0.04] visits/year), and ∼19% more inpatient admissions (IRR = 1.19 [1.13-1.25]; +0.02 [0.01-0.02] admissions/year). Most prominently, *iPRS_e_* was linked to a 23.9% increase in total days spent in hospital (IRR = 1.24 [95%CI: 1.15-1.34]), translating to an additional 0.16 [0.10-0.23] hospitalization days per year.

**Figure 5.**
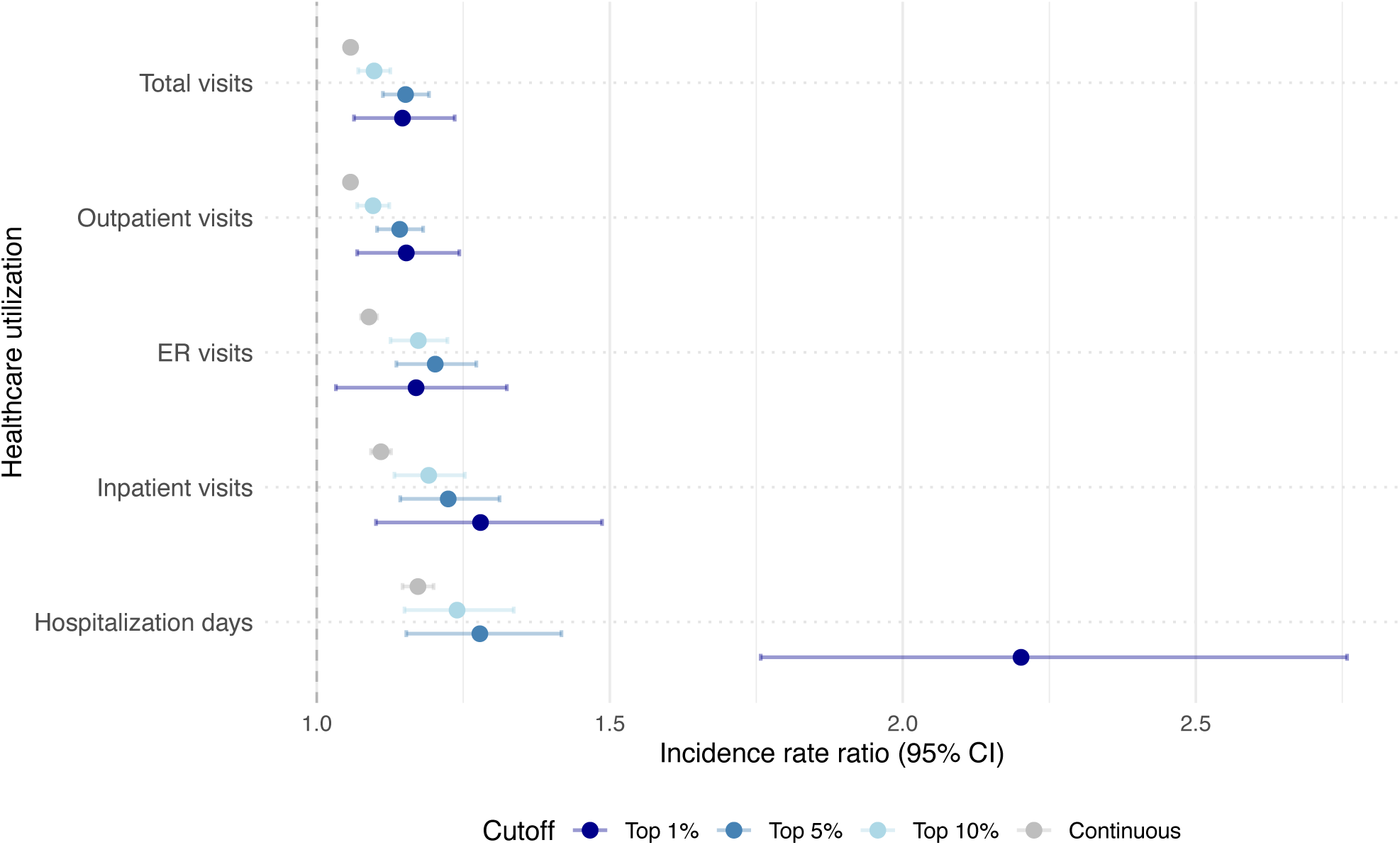
Integrated PRS effects on healthcare utilization across different integrated PRS cutoffs. The effects of continuous, the top 10%, 5% and 1% integrated PRS (iPRSe) on healthcare utilization are shown for the TWB-NHIRD linked cohort. The five healthcare utilization metrics used are hospitalization days, inpatient visits, outpatient visits, emergency room (ER) visits, and total visits. iPRS was constructed using 5-fold cross-validation Elastic Net regression with a 4:1 training set. Incidence Rate Ratios (IRRs) and 95% confidence intervals (error bars) were estimated using negative binomial regression.

As the *iPRS_e_* threshold became more stringent, the estimated effects increased, peaking at the top 1%. Being in the top 1% *iPR*S distribution was linked to a ∼15% increase in outpatient visits (IRR = 1.15 [1.07-1.24]; +1.25 [0.36-2.14] visits/year); a 28% increase in inpatient visits (1.28[1.10-1.49]; and further, a 120% increase in inpatient days (IRR = 2.20 [1.76-2.76]), translating to 0.80 [0.46-1.14] additional hospitalization days per year. Overall, these results indicate that higher polygenic burden is associated with both the frequency and the duration of healthcare use.

## Discussion

In this study, we comprehensively assessed disease burden attributable to high polygenic risk and its association with healthcare utilization for 42 common chronic diseases in the Taiwanese population. Individuals in the top 10% of the PRS distribution (*PRS_e_*) showed the most pronounced risk elevation for AFib (HR = 3.56 [95% CI: 3.23-3.92]), gout (HR = 2.41 [ 2.33-2.49]), and cancer of prostate (2.23 [1.88-2.65]), corresponding to PAFs of 0.20 [0.18-0.23], 0.12 [0.12-0.13], and 0.11 [0.08-0.14], respectively. Combined with total disease DALYs from GBD, we observed the highest population-level DALYs attributable to *PRS_e_* for T2D, ICH, and lung cancer. At the individual level, *PRS_e_* for these conditions translated to 1.50 [1.42-1.57], 0.57 [0.40-0.76], and 0.51 [0.36-0.67] healthy life-years lost, with loss approaching 2.4 years for the top 1%. Polygenic attributable burden was overall greater in men, mostly evidently for metabolic and cardiovascular conditions like T2D (1.74 reduction in health life years vs. women: 1.33) and ICH (1.04 vs. women: 0.27). Notably, for several conditions such as AFib, colorectal cancer, and T2D, the *PRS_e_*-attributable burden was of a similar magnitude to that of some well-established MRFs (e.g., smoking, low physical activity). Finally, upper percentiles of an integrated PRS were significantly associated with increased healthcare utilization— both the frequency and duration—with a stronger relative effect on inpatient days and admissions (IRR = 1.19-1.23) than on outpatient visits (IRR = 1.10).

Although our study treated *PRS_e_* and established MRFs as separate contributors for comparison, many MRFs are themselves moderately to highly heritable (e.g., SNP-heritability: 0.27 for BMI; 0.18 for smoking behavior).^45^ The attributable burdens of genomic liability and modifiable risk therefore overlap rather than partition cleanly. The non-independence may occur via mediation, where the MRF lies on the causal pathway between the genetic variants and the disease (e.g., PRS→BMI→T2D); pleiotropy, where PRS captures causal variants independently influencing both an MRF and the disease; and gene-environment interactions, where elevated polygenic risk amplifies or dampens the effect of an environmental exposure. As a result, the nontrivial genetic influence of MRFs likely overstates the fraction of their attributable burden that is genuinely modifiable, pointing to limits of what MRF-targeted interventions can avert and underscoring the value of quantifying genetic contribution to disease burden. While our comparative analyses were constrained by the MRF definitions and burden estimates provided in GBD, future investigations would benefit from formally disentangling the direct and indirect (MRF-mediated) effects of genetic factors on disease burden— for instance, leveraging mediation analysis or Mendelian randomization—in a sample with a comprehensive panel of MRFs measured alongside genotypes.

While our primary analyses relied on ICD codes mapped to PheCodes, comparison with SR medical conditions revealed low to moderate agreement (Cohen’s κ; mean = 0.28). Somewhat unexpectedly, despite PheCodes yielding higher overall prevalence, SR-based estimates—genetic effects, PAF, and DALYs—were generally larger, except for AFib. This discrepancy could arise from differential ascertainment in SR, enriching for clinically manifest cases and conflating biological susceptibility with health consciousness, or attenuation of PheCode-based estimates by false-positive cases (overreporting) from the administrative origins of ICD codes. We consider the latter mechanism less likely to dominate, as requiring at least two qualifying PheCodes yielded prevalence estimates broadly consistent with prior Taiwanese studies.^35,46^ Code-based phenotyping itself is limited in a further respect: it precludes us from distinguishing disease subtypes with at least partly distinct germline susceptibility (e.g., ER status in breast cancer, histology in lung cancer)^47,48^, most of which also lacked an adequately powered subtype-specific GWAS. Our estimates should therefore be read as disease-level averages rather than subtype-specific effects. Overall, disease definition can materially affect genetic burden estimates, warranting caution in their interpretation within the specific phenotyping context.

Beyond phenotype definitions, PRS accuracy itself may influence estimated attributable burden. Although we prioritized large-scale EAS or multi-ancestry GWAS for computing PRS, several diseases lacked an EAS-specific reference and relied on EUR GWAS (e.g., ischemic heart disease, breast cancer, cancer of prostate; **Table S2**), and EAS discovery cohorts are generally smaller than EUR ones. Given cross-population differences in MAF and LD patterns, the imperfect transferability of EUR-derived effect sizes to the Taiwanese population—along with this power imbalance—likely attenuated PRS effects and underestimated the genetically attributable burden. We indeed found that many of our estimates were lower than those from the FinnGen biobank^16^ (e.g., cancer of prostate: top 10% HR = 3.32 vs. 2.23 [TWB], individual DALYs 0.62 vs. 0.17; ischemic heart disease: HR = 2.03 vs. 1.14 [TWB], individual DALYs 2.67 vs. 0.15). While differences in disease epidemiology, DALY parameters, and methodology also contribute, our estimates may be conservative for the affected conditions in this population.

Our study has several strengths. First, we leveraged a large, population-scale biobank of 140,000 individuals linked to a national, longitudinal claims database spanning more than two decades. Because Taiwan’s single-payer system essentially capturing all healthcare encounters and utilization, this linkage provided one of the most comprehensive characterizations of polygenic disease burden and its downstream systemic consequences in a non-EUR population. Second, the genetic ancestry of the Taiwanese population is comparatively homogeneous, predominantly of Southern Han Chinese descent, which minimizes the potential for population stratification to confound our genetic effect estimates. Consequently, while absolute disease burden metrics (like DALYs and healthcare visits) are inherently country-specific and shaped by the local Taiwanese healthcare environment—such as the high utilization rates driven by the accessibility of Taiwan’s NHI—our relative genetic risk estimates may offer a useful reference for Han Chinese populations more broadly.^49,50^ Lastly, whereas MRFs are dynamic and fluctuate over the life course, an individual’s germline polygenic risk is fixed at conception and measurable long before disease symptoms appear, providing a stable, lifelong basis for risk stratification and for prioritizing screening and prevention.

In conclusion, using a large-scale population biobank linked to longitudinal medical records, we demonstrated that genetic risk is a measurable driver of Taiwan’s disease burden. This quantitative foundation can be used for integrating genetic risk into public health planning in Taiwan, bridging the gap between polygenic risk and disease burden.

## Data Availability

All data produced in the present study are available upon reasonable request to the authors

## Contributors

Study concept and design: MYS and YAF.

Data curation: MYS, YSK, XYC and LLW.

Methodology: MYS and YAF.

Formal analysis: MYS.

Visualization: MYS.

Software: MYS.

Funding acquisition: YAF.

Resources: YAF.

Study supervision: YAF.

Writing – original draft: MYS and YAF.

Writing – review and editing: all authors.

## Data sharing statement

All collected GWAS summary statistics are available for public download (**Table S2**). TWB data including individual genotypes and phenotypes is available upon application for research purposes. A detailed description of TWB data availability and the application process can be found at https://www.biobank.org.tw/.

## Acknowledgements

We thank all the Taiwan Biobank & the National Health Insurance Research Database participants and staff for making this resource available. This work is supported by the National Taiwan University Academic Research-Career Development Project (NTU-115L7813). YAF also acknowledges support from the Ministry of Education (the Yushan Young Fellow Program, MOE-111-YSFAG-0003-001-P1) and the Population Health Research Center from Featured Areas Research Center Program within the framework of the Higher Education Sprout Project (NTU-115L900402).

Access to the TWB data for this study was approved by the Ethics and Governance Council (EGC) of Taiwan Biobank (approval number: TWBR11211-02). The use of TWB and its linkage to the National Health Insurance Database (NHIRD) were approved by the Institutional Review Board of National Taiwan University Hospital (202308103RINB). The collection of TWB data was approved by its EGC and by the Department of Health and Welfare, Taiwan (Wei-Shu-I-Tzu No. 1010267471). TWB obtained informed consent from all participants for the research use of collected data.

## References

1. Vollset SE, Ababneh HS, Abate YH, et al. Burden of disease scenarios for 204 countries and territories, 2022– 2050: a forecasting analysis for the Global Burden of Disease Study 2021. The Lancet 2024; **403**(10440): 2204–2256.

2. Slama S, Kim HJ, Roglic G, et al. Care of non-communicable diseases in emergencies. The Lancet 2017; 389(10066): 326–330.

3. Barnett K, Mercer SW, Norbury M, Watt G, Wyke S, Guthrie B. Epidemiology of multimorbidity and implications for health care, research, and medical education: a cross-sectional study. The Lancet 2012; 380(9836): 37–43.

4. Muka T, Imo D, Jaspers L, et al. The global impact of non-communicable diseases on healthcare spending and national income: a systematic review. Eur J Epidemiol 2015; 30(4): 251–277.

5. Wang TD, Huang ST, Wang CY, Lin FJ, Chen HM, Hsiao FY. Nationwide Trends in Incidence, Healthcare Utilization, and Mortality in Hospitalized Heart Failure Patients in Taiwan. ESC Heart Fail 2020; 7(6): 3653–3666.

6. Wu N, Xie X, Cai M, Han Y, Wu S. Trends in health service needs, utilization, and non-communicable chronic diseases burden of older adults in China: evidence from the 1993 to 2018 National Health Service Survey. Int J Equity Health 2023; 22(1): 169.

7. Vujkovic M, Keaton JM, Lynch JA, et al. Discovery of 318 new risk loci for type 2 diabetes and related vascular outcomes among 1.4 million participants in a multi-ancestry meta-analysis. Nat Genet 2020; 52(7): 680–691.

8. Zhang H, Ahearn TU, Lecarpentier J, et al. Genome-wide association study identifies 32 novel breast cancer susceptibility loci from overall and subtype-specific analyses. Nat Genet 2020; 52(6): 572–581.

9. Aragam KG, Jiang T, Goel A, et al. Discovery and systematic characterization of risk variants and genes for coronary artery disease in over a million participants. Nat Genet 2022; 54(12): 1803– 1815.

10. Abdellaoui A, Yengo L, Verweij KJ, Visscher PM. 15 years of GWAS discovery: Realizing the promise. Am J Hum Genet 2023; 110(2): 179–194.

11. Khera AV, Chaffin M, Aragam KG, et al. Genome-wide polygenic scores for common diseases identify individuals with risk equivalent to monogenic mutations. Nat Genet 2018; 50(9): 1219– 1224.

12. Lewis CM, Vassos E. Polygenic risk scores: from research tools to clinical instruments. Genome Med 2020; 12: 44.

13. Choi SW, Mak TSH, O’Reilly PF. Tutorial: a guide to performing polygenic risk score analyses. Nat Protoc 2020; 15(9): 2759–2772.

14. Huerta-Chagoya A, Kim J, Mandla R, et al. Multi-ancestry polygenic risk scores for the prediction of type 2 diabetes and complications in diverse ancestries. The Lancet Diabetes Endocrinol 2026; 14(7): 558–569.

15. Meisner A, Kundu P, Zhang YD, et al. Combined utility of 25 disease and risk factor polygenic risk scores for stratifying risk of all-cause mortality. Am J Hum Genet 2020; 107(3): 418–431.

16. Jukarainen S, Kiiskinen T, Kuitunen S, et al. Genetic risk factors have a substantial impact on healthy life years. Nat Med 2022; 28(9): 1893–1901.

17. Widen E, Lello L, Raben TG, Tellier LCAM, Hsu SDH. Polygenic Health Index, General Health, and Pleiotropy: Sibling Analysis and Disease Risk Reduction. Sci Rep 2022; 12(1): 18173.

18. Huang Q, Lan X, Chen H, et al. Association between genetic predisposition and disease burden of stroke in China: a genetic epidemiological study. The Lancet Regional Health– Western Pacific 2023; 36.

19. Lo WC, Ku CC, Chiou ST, et al. Adult mortality of diseases and injuries attributable to selected metabolic, lifestyle, environmental, and infectious risk factors in Taiwan: a comparative risk assessment. Popul Health Metr 2017; 15(1): 17.

20. Yusuf S, Joseph P, Rangarajan S, et al. Modifiable risk factors, cardiovascular disease, and mortality in 155 722 individuals from 21 high-income, middle-income, and low-income countries (PURE): a prospective cohort study. Lancet 2020; 395(10226): 795–808.

21. Global Cardiovascular Risk Consortium, Magnussen C, Ojeda FM, et al. Global effect of modifiable risk factors on cardiovascular disease and mortality. N Engl J Med 2023; **389**(14): 1273–1285.

22. Fink H, Langselius O, Vignat J, et al. Global and regional cancer burden attributable to modifiable risk factors to inform prevention. Nat Med 2026; 32(4): 1306–1315.

23. Hay SI, Ong KL, Santomauro DF, et al. Burden of 375 diseases and injuries, risk-attributable burden of 88 risk factors, and healthy life expectancy in 204 countries and territories, including 660 subnational locations, 1990–2023: a systematic analysis for the Global Burden of Disease Study 2023. The Lancet 2025; **406**(10513): 1873–1922.

24. Liang H, Jing D, Zhu Y, et al. Association of genetic risk and lifestyle with incident adult-onset asthma in the UK Biobank cohort 2023; 9(2).

25. Bhattacharya R, Marnell CS, Cho SMJ, et al. Importance of Clinical, Laboratory, and Genetic Risk Factors for Incident CAD. Circ Genom Precis Med 2025; 18(4): e004937.

26. Feng YCA, Chen CY, Chen TT, et al. Taiwan Biobank: A rich biomedical research database of the Taiwanese population. Cell Genomics 2022; 2(11): 100197.

27. Lin LY, Warren-Gash C, Smeeth L, Chen PC. Data resource profile: the national health insurance research database (NHIRD). Epidemiol Health 2018; 40: e2018062.

28. Bastarache L. Using phecodes for research with the electronic health record: From PheWAS to PheRS. Annu Rev Biomed Data Sci 2021; 4: 1–19.

29. Carroll RJ, Bastarache L, Denny JC. R PheWAS: data analysis and plotting tools for phenome-wide association studies in the R environment. Bioinformatics 2014; 30(16): 2375–76

30. Ge T, Chen CY, Ni Y, Feng YCA, Smoller JW. Polygenic prediction via Bayesian regression and continuous shrinkage priors. Nat Commun 2019; 10(1): 1776.

31. 1000 Genomes Project Consortium, Auton A, Brooks LD, et al. A global reference for human genetic variation. Nature 2015; 526(7571): 68–74.

32. Purcell S, Neale B, Todd-Brown K, et al. PLINK: a tool set for whole-genome association and population-based linkage analyses. Am J Hum Genet 2007; 81(3): 559–575.

33. Ruan Y, Lin YF, Feng YCA, et al. Improving polygenic prediction in ancestrally diverse populations. Nat Genet 2022; 54(5): 573–580.

34. Therneau TM, Elizabeth A, Cynthia C. survival: Survival Analysis. Version 3.8–6. Jan. 16, 2026.

35. Yang HC, Kwok PY, Li LH, et al. The Taiwan Precision Medicine Initiative provides a cohort for large-scale studies. Nature 2025; 648(8092): 117–127.

36. Rowe AK, Powell KE, Flanders WD. Why population attributable fractions can sum to more than one. Am J Prev Med 2004; 26(3): 243–249.

37. Witte JS, Visscher PM, Wray NR. The contribution of genetic variants to disease depends on the ruler. Nat Rev Genet 2014; 15(11): 765–776.

38. Ministry of the Interior. Abridged Life Table of Taiwan. 2023.

39. Truong B, Hull LE, Ruan Y, et al. Integrative polygenic risk score improves the prediction accuracy of complex traits and diseases. Cell Genomics 2024; 4(4).

40. Zou H, Hastie T. Regularization and variable selection via the elastic net. J R Stat Soc Series B Stat Methodol 2005; 67(2): 301–320.

41. Arel-Bundock V, Greifer N, Heiss A. How to Interpret Statistical Models Using marginaleffects for R and Python 2024; 111: 1–32.

42. Bycroft C, Freeman C, Petkova D, et al. The UK Biobank resource with deep phenotyping and genomic data. Nature 2018; 562(7726): 203–209.

43. Kurki MI, Karjalainen J, Palta P, et al. FinnGen provides genetic insights from a well-phenotyped isolated population. Nature 2023; 613(7944): 508–518.

44. Sakaue S, Kanai M, Tanigawa Y, et al. A cross-population atlas of genetic associations for 220 human phenotypes. Nat Genet 2021; 53(10): 1415–1424.

45. Ge T, Chen CY, Neale BM, Sabuncu MR, Smoller JW. Phenome-wide heritability analysis of the UK Biobank. PLoS Genet 2017; 13(4): e1006711.

46. Liu PC, Huang TH, Wu YC, et al. Disease burden of 30 cancer groups in taiwan from 2000 to 2021. J Epidemiol Glob Health 2025; 15(1): 62.

47. Mavaddat N, Michailidou K, Dennis J, et al. Polygenic Risk Scores for Prediction of Breast Cancer and Breast Cancer Subtypes. Am J Hum Genet 2019; 104(1): 21–34.

48. Gabriel AAG, Atkins JR, Penha RCC, et al. Genetic Analysis of Lung Cancer and the Germline Impact on Somatic Mutation Burden. J Natl Cancer Inst 2022; 114(8): 1159–1166.

49. Cheng TM. Taiwan’s new national health insurance program: genesis and experience so far. Health Aff (Millwood) 2003; 22(3): 61–76.

50. Lee WC. An inconvenient truth behind high accessibility to medical services 2011; **74**(9): 383.

