## Supplementary materials for "Burden of Non-Communicable Diseases Attributable to Polygenic Risk and Its Association with Healthcare Utilization in Taiwan"

### Supplementary Material

**Supplementary Methods**

The Taiwan Biobank genotype data quality control

Comparison between self-reported and clinically defined disease endpoints

**Supplementary Figures**

Figure S1. Overall study design and analytical workflow

Figure S2. Polygenic risk effects of selected high burden diseases across different PRSe thresholds

Figure S3. PAF for selected diseases across different PRSe thresholds

Figure S4. Comparison of HRs, PAFs and individual attributable DALYs between self-reported (SR) and PheCode across overlapping diseases

Figure S5. Overall and sex-specific PAF of polygenic risk, joint PAF of MRFs and joint PAF of all risk factors

**Supplementary Tables**

Table S1. Mapping between PheCode and ICD9CM/ICD10CM coding

Table S2. GWAS summary statistics used for calculating PRS

Table S3. Definition of MRFs

Table S4. Disease prevalence, age of cases and controls and age of onset

Table S5. PRSe effects (top 10%) across all high-burden diseases

Table S6. Sex-specific PRSe effects (top 10%) across all high-burden diseases

Table S7. Sensitivity analyses of PRSe effects (top 5% and top 1%) across all high-burden diseases

Table S8. Overall and sex-specific PAF associated with PRSe (top 10%) for diseases with significant genetic effects

Table S9. Overall and sex-specific population DALYs (DALYpop) attributable to PRSe (top 10%) for significant diseases with available DALYs estimates

Table S10. Sensitivity analyses for PAF of significant diseases (top 5% and top 1%)

Table S11. Sensitivity analyses for DALYpop of significant diseases with available DALYs estimates (top 5% and top 1%)

Table S12. Overall and sex-specific individual DALYs (DALYind) attributable to PRSe (top 10%) for significant diseases with available DALYs estimates

Table S13. Sensitivity analyses for DALYind of significant diseases with available DALYs estimates (top 5% and top 1%)

Table S14. Kappa statistics for overlap between PheCode and self-reported (SR) definitions

Table S15. Overall and sex-specific comparison of PRSe and MRFs for significant diseases with available DALYs estimates

Table S16. Overall and sex-specific PAF of PRSe, joint PAF of MRFs, and joint PAF of all risk factors

Table S17. Trained mixing weights of integrated PRS for healthcare utilization indices

#### Supplementary Methods

##### The Taiwan Biobank (TWB) genotype quality control

We excluded samples with a call rate < 0.98, an outlying heterozygosity rate (> 6 standard deviations), and discordant reported and imputed sexes. Principal component analysis (PCA) was conducted with the 1000 Genomes (1KG) samples[1] as the reference panel to infer population structure, keeping only those classified as East Asians with a prediction probability > 0.8. SNPs were discarded for having a call rate < 0.95 and failing the Hardy-Weinberg Equilibrium (HWE) test (p-value < $1 \times 10^{-10}$). Phasing and imputation were then performed using Eagle v2.4[2] and Minimac4[3] with the 1KG East-Asian samples as the linkage equilibrium (LD) reference panel. After imputation, we retained SNPs with an imputation quality (INFO score) > 0.6 and a minor allele frequency (MAF) > 0.1%, yielding a final analytic sample of 27,266 individuals and 8,162,014 variants in batch 1 and 117,259 individuals and 7,553,933 variants in batch 2 of genome build GRCh37.

##### Comparison between self-reported and clinically defined disease endpoints

For self-reported (SR) medical conditions available in the TWB questionnaires, we compared their $PRS_e$ effects against those based on the clinically defined PheCode endpoints. Phenotype harmonization yielded 10 mappable disease definitions: intracranial hemorrhage (SR: apoplexia), AFib (SR: arrhythmia), asthma, breast cancer, ischemic heart disease (SR: coronary artery disease), depression, T2D, gout, hypercholesterolemia, hypertension. Cases were defined as reporting a prior diagnosis of each target disease, with an approximate age of diagnosis. Controls were defined as all participants without a self-reported diagnosis of that disease. For each mapped disease, we replicated the same analyses as previously described for the SR definitions, including Cox regression for HR estimation (using SR age of diagnosis), PAF calculation, and estimation of population-level and individual-level attributable DALYs using GBD 2023 estimates. Missing values were excluded for all analyses.

#### Supplementary Figures

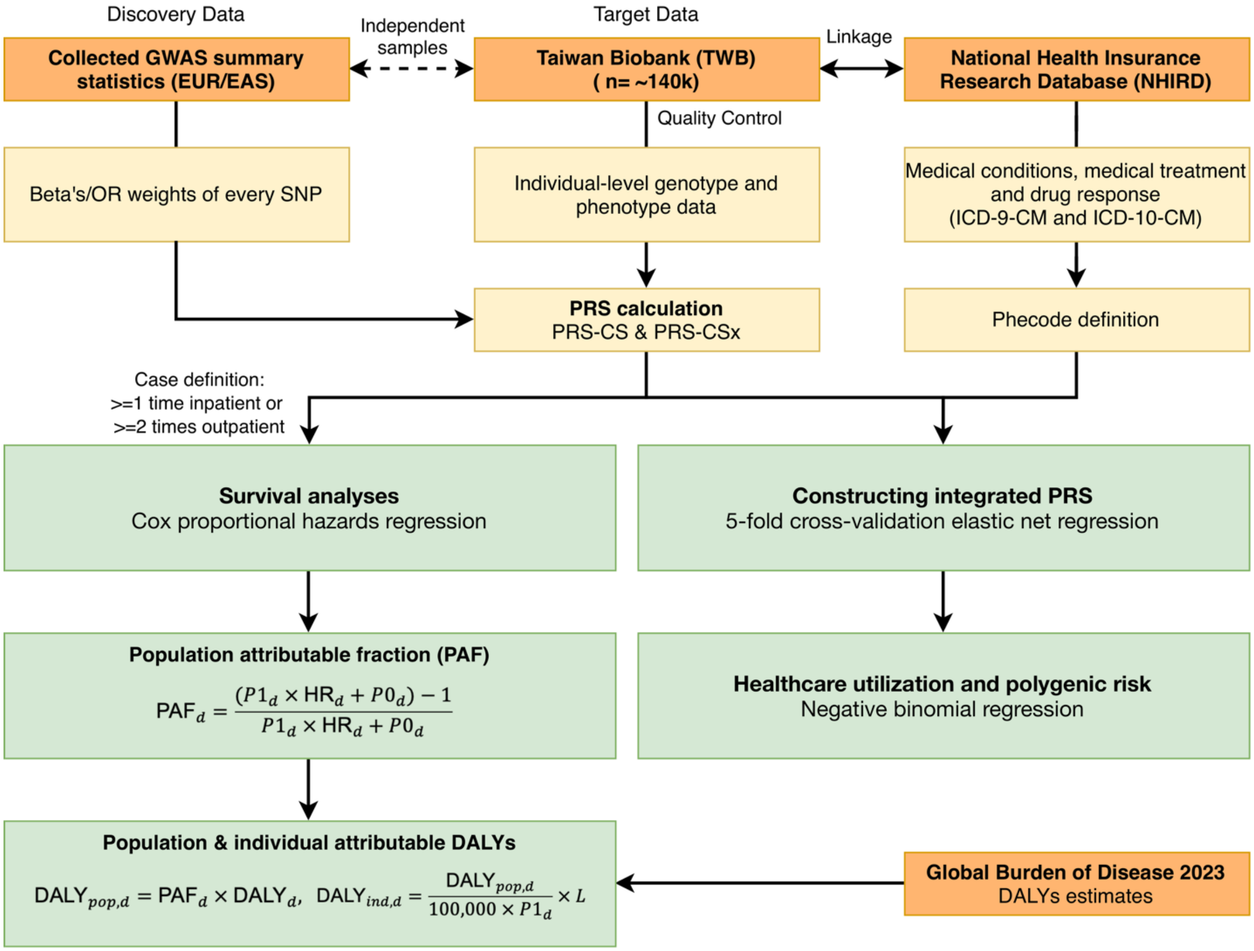

**Figure S1. Overall study design and analytical workflow**

We derived PRS weights from SNP effect sizes obtained from collected GWAS summary statistics. Leveraging linkage between the TWB and NHIRD, we then conducted survival analyses to estimate hazard ratios. Subsequently, we calculated PAFs, population-level DALYs, and individual-level DALYs using DALY estimates from the GBD 2023 study. Additionally, we constructed an integrated PRS to assess its association with healthcare utilization.

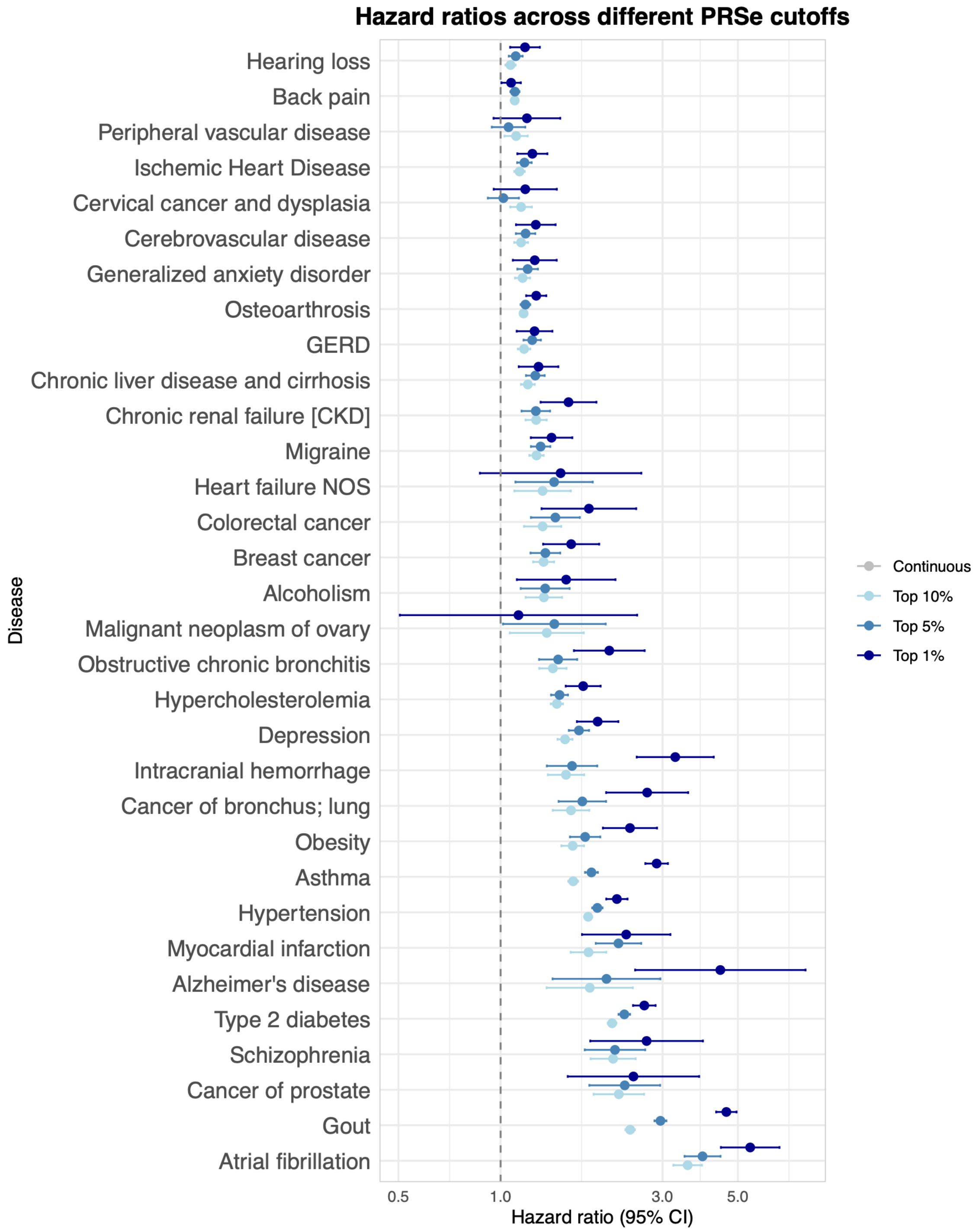

**Figure S2. Polygenic risk effects of selected high burden diseases across different PRSe thresholds**

Effects of the continuous, the top 10%, top 5% and top1% PRS (PRSe) on disease risk are shown for the TWB-NHIRD linked cohort. Hazard ratios (HRs) and 95% confidence intervals (error bars) were derived from Cox proportional hazards regression with age as the time scale. Depicted are 32 of the 42 diseases with a statistically significant top 10% PRS association ($p < 0.05$), ordered by the top 10% PRS effect size.

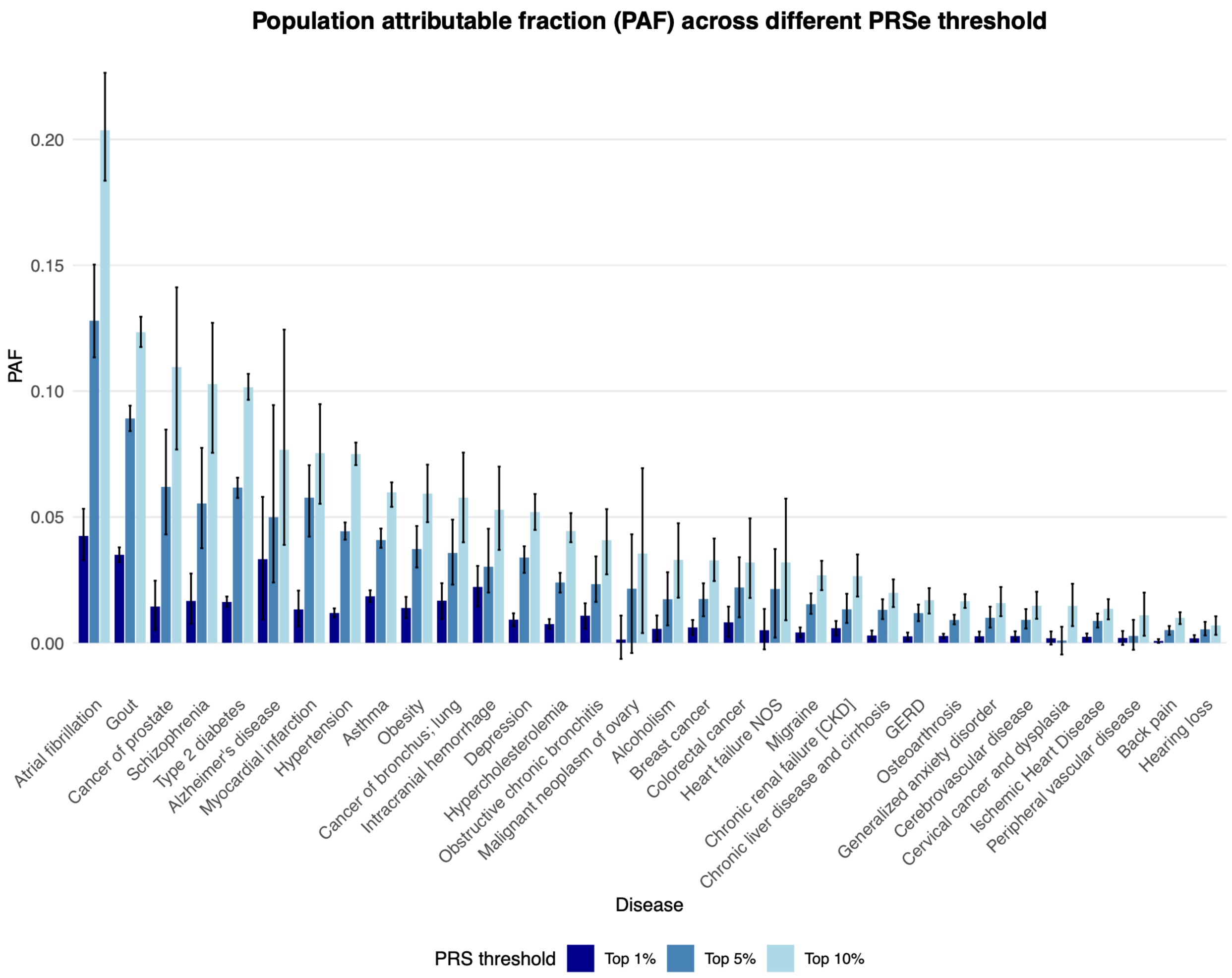

**Figure S3. PAF for selected diseases across different PRSe thresholds**

PAF of the the top 10%, top 5% and top1% PRS (PRSe) on disease risk are shown for the TWB-NHIRD linked cohort. Estimates were derived from the HRs. 95% confidence intervals (error bars) were estimated using 1000-fold bootstrapping. Depicted are 25 of the 42 diseases with a statistically significant top 1% PRS association ($p < 0.05$), ordered by the top 10% PAF.

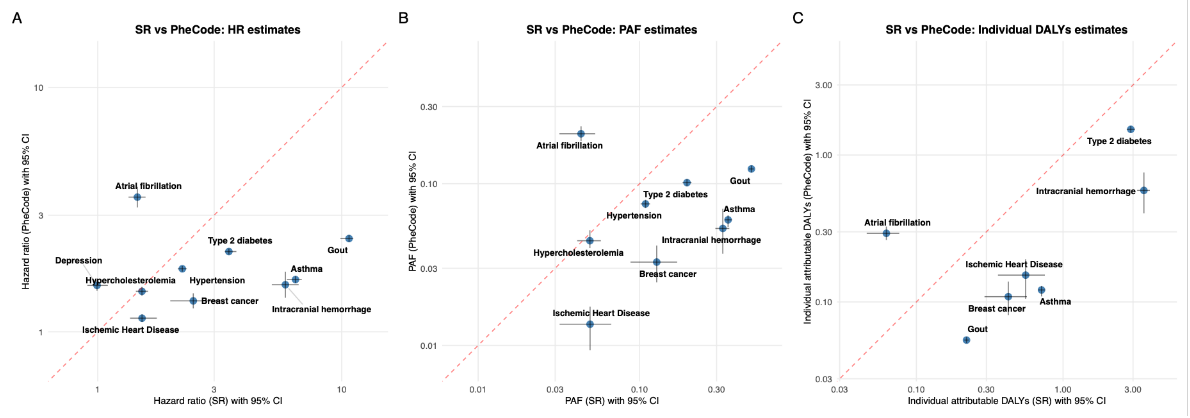

**Figure S4. Comparison of HRs, PAFs and individual attributable DALYs between self-reported (SR) and PheCode across overlapping diseases**

Estimates of HR ratios (panel A), PAF (panel B), individual attributable DALYs (panel C) are shown for the TWB-NHIRD linked cohort. The x-axis and y-axis show the estimates for SR and PheCode definitions respectively with a log10 scale. 95% confidence intervals (error bars) of HRs were derived from Cox proportional hazards regression with age as the time scale. 95% confidence intervals (error bars) of PAF and individual attributable DALYs were derived from 1000-fold bootstrapping. Depicted are the overlapping diseases between the two phenotype definitions. The

dashed red lines represent the identity line, indicating perfect agreement.

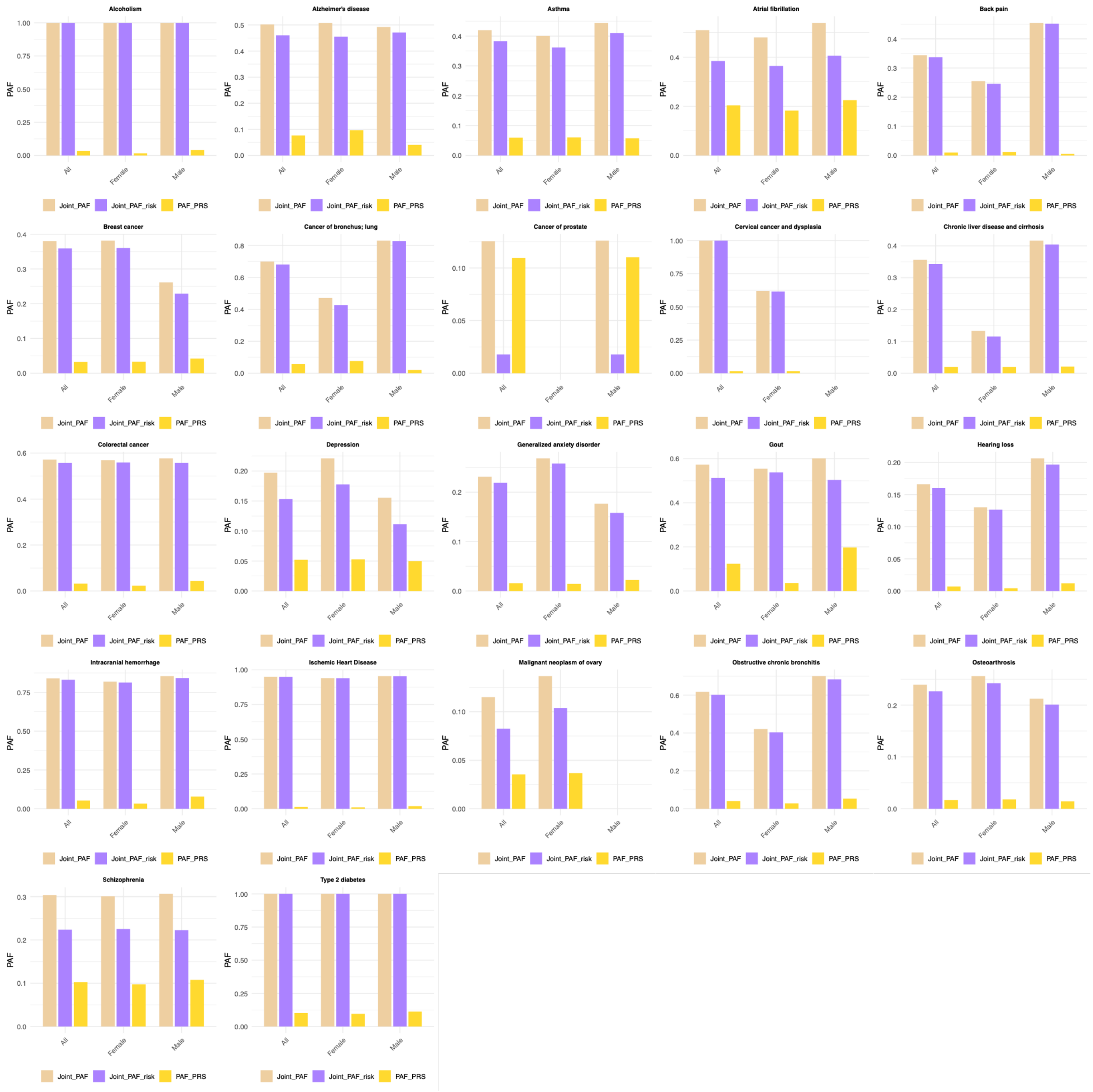

**Figure S5. Overall and sex-specific PAF of polygenic risk, joint PAF of MRFs and joint PAF of all risk factors**

This figure shows the PAF of polygenic risk (PAF_PRS), joint PAF of MRFs (Joint_PAF_risk) and joint PAF all risk factors (Joint_PAF) of all diseases with available MRFs DALYs estimates in the GBD 2023 study.

#### Supplementary Tables

#### Table S1. Mapping between PheCode and ICD9CM/ICD10CM coding

| Disease category | Disease name | PheCode | Vocabulary | ICD code | ICD description |
|---|---|---|---|---|---|
| **Neoplasms** | Cancer of esophagus | 150 | ICD9CM | 150.1 | Malignant neoplasm of thoracic esophagus |
| | Cancer of esophagus | 150 | ICD9CM | 150.2 | Malignant neoplasm of abdominal esophagus |
| | Cancer of esophagus | 150 | ICD9CM | 150 | Malignant neoplasm of esophagus |
| | Cancer of esophagus | 150 | ICD10CM | C15.9 | Malignant neoplasm of esophagus, unspecified |
| | Cancer of esophagus | 150 | ICD9CM | 150 | Malignant neoplasm of cervical esophagus |
| | Cancer of esophagus | 150 | ICD9CM | 230.1 | Carcinoma in situ of esophagus |
| | Cancer of esophagus | 150 | ICD10CM | C15 | Malignant neoplasm of esophagus |
| | Cancer of esophagus | 150 | ICD9CM | 150.9 | Malignant neoplasm of esophagus, unspecified site |
| | Cancer of esophagus | 150 | ICD10CM | C15.8 | Malignant neoplasm of overlapping sites of esophagus |
| | Cancer of esophagus | 150 | ICD10CM | C15.4 | Malignant neoplasm of middle third of esophagus |
| | Cancer of esophagus | 150 | ICD10CM | C15.5 | Malignant neoplasm of lower third of esophagus |
| | Cancer of esophagus | 150 | ICD10CM | D00.1 | Carcinoma in situ of esophagus |
| | Cancer of esophagus | 150 | ICD9CM | 150.5 | Malignant neoplasm of lower third of esophagus |
| | Cancer of esophagus | 150 | ICD9CM | V10.03 | Personal history of malignant neoplasm of esophagus |
| | Cancer of esophagus | 150 | ICD9CM | 150.4 | Malignant neoplasm of middle third of esophagus |
| | Cancer of esophagus | 150 | ICD10CM | C15.3 | Malignant neoplasm of upper third of esophagus |
| | Cancer of esophagus | 150 | ICD10CM | Z85.01 | Personal history of malignant neoplasm of esophagus |
| | Cancer of esophagus | 150 | ICD9CM | 150.8 | Malignant neoplasm of other specified part of esophagus |
| | Cancer of esophagus | 150 | ICD9CM | 150.3 | Malignant neoplasm of upper third of esophagus |
| | Cancer of stomach | 151 | ICD9CM | 151 | Malignant neoplasm of stomach |
| | Cancer of stomach | 151 | ICD9CM | 151.4 | Malignant neoplasm of body of stomach |
| | Cancer of stomach | 151 | ICD9CM | 151.3 | Malignant neoplasm of fundus of stomach |
| | Cancer of stomach | 151 | ICD10CM | C16.2 | Malignant neoplasm of body of stomach |
| | Cancer of stomach | 151 | ICD10CM | Z85.02 | Personal history of malignant neoplasm of stomach |
| | Cancer of stomach | 151 | ICD9CM | 151.8 | Malignant neoplasm of other specified sites of stomach |
| | Cancer of stomach | 151 | ICD9CM | 151.1 | Malignant neoplasm of pylorus |
| | Cancer of stomach | 151 | ICD9CM | 151.5 | Malignant neoplasm of lesser curvature of stomach, unspecified |
| | Cancer of stomach | 151 | ICD10CM | C16.4 | Malignant neoplasm of pylorus |
| | Cancer of stomach | 151 | ICD9CM | 151.6 | Malignant neoplasm of greater curvature of stomach, unspecified |
| | Cancer of stomach | 151 | ICD10CM | D00.2 | Carcinoma in situ of stomach |

| | | | | |
|---|---|---|---|---|
| Cancer of stomach | 151 | ICD9CM | 151.2 | Malignant neoplasm of pyloric antrum |
| Cancer of stomach | 151 | ICD10CM | Z85.020 | Personal history of malignant carcinoid tumor of stomach |
| Cancer of stomach | 151 | ICD9CM | 230.2 | Carcinoma in situ of stomach |
| Cancer of stomach | 151 | ICD10CM | C16.5 | Malignant neoplasm of lesser curvature of stomach, unspecified |
| Cancer of stomach | 151 | ICD10CM | C7A.092 | Malignant carcinoid tumor of the stomach |
| Cancer of stomach | 151 | ICD10CM | C16 | Malignant neoplasm of stomach |
| Cancer of stomach | 151 | ICD10CM | Z85.028 | Personal history of other malignant neoplasm of stomach |
| Cancer of stomach | 151 | ICD10CM | C16.8 | Malignant neoplasm of overlapping sites of stomach |
| Cancer of stomach | 151 | ICD9CM | 151 | Malignant neoplasm of cardia |
| Cancer of stomach | 151 | ICD9CM | 209.23 | Malignant carcinoid tumor of the stomach |
| Cancer of stomach | 151 | ICD10CM | C16.0 | Malignant neoplasm of cardia |
| Cancer of stomach | 151 | ICD10CM | C16.3 | Malignant neoplasm of pyloric antrum |
| Cancer of stomach | 151 | ICD10CM | C16.6 | Malignant neoplasm of greater curvature of stomach, unspecified |
| Cancer of stomach | 151 | ICD9CM | 151.9 | Malignant neoplasm of stomach, unspecified site |
| Cancer of stomach | 151 | ICD9CM | V10.04 | Personal history of malignant neoplasm of stomach |
| Cancer of stomach | 151 | ICD10CM | C16.1 | Malignant neoplasm of fundus of stomach |
| Cancer of stomach | 151 | ICD10CM | C16.9 | Malignant neoplasm of stomach, unspecified |
| Colorectal cancer | 153 | ICD9CM | 154.8 | Malignant neoplasm of other sites of rectum, rectosigmoid junction, and anus |
| Colorectal cancer | 153 | ICD10CM | C21.8 | Malignant neoplasm of overlapping sites of rectum, anus and anal canal |
| Colorectal cancer | 153 | ICD10CM | C21.2 | Malignant neoplasm of cloacogenic zone |
| Colorectal cancer | 153 | ICD9CM | 153.6 | Malignant neoplasm of ascending colon |
| Colorectal cancer | 153 | ICD9CM | 153.9 | Malignant neoplasm of colon, unspecified site |
| Colorectal cancer | 153 | ICD10CM | C18.0 | Malignant neoplasm of cecum |
| Colorectal cancer | 153 | ICD9CM | 209.11 | Malignant carcinoid tumor of the appendix |
| Colorectal cancer | 153 | ICD9CM | 209.1 | Malignant carcinoid tumors of the appendix, large intestine, and rectum |
| Colorectal cancer | 153 | ICD9CM | 153.8 | Malignant neoplasm of other specified sites of large intestine |
| Colorectal cancer | 153 | ICD10CM | C7A.029 | Malignant carcinoid tumor of the large intestine, unspecified portion |
| Colorectal cancer | 153 | ICD10CM | C7A.024 | Malignant carcinoid tumor of the descending colon |
| Colorectal cancer | 153 | ICD9CM | 209.12 | Malignant carcinoid tumor of the cecum |
| Colorectal cancer | 153 | ICD10CM | C7A.022 | Malignant carcinoid tumor of the ascending colon |
| Colorectal cancer | 153 | ICD10CM | D01.0 | Carcinoma in situ of colon |
| Colorectal cancer | 153 | ICD10CM | C18.7 | Malignant neoplasm of sigmoid colon |

| | | | | |
|---|---|---|---|---|
| Colorectal cancer | 153 | ICD9CM | 153.1 | Malignant neoplasm of transverse colon |
| Colorectal cancer | 153 | ICD10CM | C18.5 | Malignant neoplasm of splenic flexure |
| Colorectal cancer | 153 | ICD10CM | Z85.030 | Personal history of malignant carcinoid tumor of large intestine |
| Colorectal cancer | 153 | ICD10CM | C18.2 | Malignant neoplasm of ascending colon |
| Colorectal cancer | 153 | ICD9CM | 209.13 | Malignant carcinoid tumor of the ascending colon |
| Colorectal cancer | 153 | ICD10CM | C18.3 | Malignant neoplasm of hepatic flexure |
| Colorectal cancer | 153 | ICD9CM | 209.15 | Malignant carcinoid tumor of the descending colon |
| Colorectal cancer | 153 | ICD10CM | Z85.038 | Personal history of other malignant neoplasm of large intestine |
| Colorectal cancer | 153 | ICD9CM | 153 | Malignant neoplasm of hepatic flexure |
| Colorectal cancer | 153 | ICD9CM | 153.3 | Malignant neoplasm of sigmoid colon |
| Colorectal cancer | 153 | ICD9CM | 209.14 | Malignant carcinoid tumor of the transverse colon |
| Colorectal cancer | 153 | ICD10CM | C18.1 | Malignant neoplasm of appendix |
| Colorectal cancer | 153 | ICD9CM | 209.16 | Malignant carcinoid tumor of the sigmoid colon |
| Colorectal cancer | 153 | ICD10CM | C18 | Malignant neoplasm of colon |
| Colorectal cancer | 153 | ICD9CM | 153 | Malignant neoplasm of colon |
| Colorectal cancer | 153 | ICD10CM | C18.8 | Malignant neoplasm of overlapping sites of colon |
| Colorectal cancer | 153 | ICD9CM | 230.3 | Carcinoma in situ of colon |
| Colorectal cancer | 153 | ICD9CM | V10.05 | Personal history of malignant neoplasm of large intestine |
| Colorectal cancer | 153 | ICD9CM | 153.5 | Malignant neoplasm of appendix vermiformis |
| Colorectal cancer | 153 | ICD10CM | C18.4 | Malignant neoplasm of transverse colon |
| Colorectal cancer | 153 | ICD10CM | C18.6 | Malignant neoplasm of descending colon |
| Colorectal cancer | 153 | ICD9CM | 209.1 | Malignant carcinoid tumor of the large intestine, unspecified portion |
| Colorectal cancer | 153 | ICD10CM | C7A.025 | Malignant carcinoid tumor of the sigmoid colon |
| Colorectal cancer | 153 | ICD9CM | 153.7 | Malignant neoplasm of splenic flexure |
| Colorectal cancer | 153 | ICD10CM | C7A.020 | Malignant carcinoid tumor of the appendix |
| Colorectal cancer | 153 | ICD9CM | 159 | Malignant neoplasm of intestinal tract, part unspecified |
| Colorectal cancer | 153 | ICD9CM | 153.4 | Malignant neoplasm of cecum |
| Colorectal cancer | 153 | ICD10CM | C18.9 | Malignant neoplasm of colon, unspecified |
| Colorectal cancer | 153 | ICD10CM | C7A.023 | Malignant carcinoid tumor of the transverse colon |
| Colorectal cancer | 153 | ICD10CM | C7A.021 | Malignant carcinoid tumor of the cecum |
| Colorectal cancer | 153 | ICD10CM | C26.0 | Malignant neoplasm of intestinal tract, part unspecified |
| Colorectal cancer | 153 | ICD9CM | 153.2 | Malignant neoplasm of descending colon |
| Colorectal cancer | 153 | ICD9CM | 796.7 | Abnormal cytologic smear of anus and anal HPV |

| | | | | |
|---|---|---|---|---|
| Colorectal cancer | 153 | ICD10CM | Z85.048 | Personal history of other malignant neoplasm of rectum, rectosigmoid junction, and anus |
| Colorectal cancer | 153 | ICD10CM | C21.1 | Malignant neoplasm of anal canal |
| Colorectal cancer | 153 | ICD10CM | R85.619 | Unspecified abnormal cytological findings in specimens from anus |
| Colorectal cancer | 153 | ICD10CM | R85.614 | Cytologic evidence of malignancy on smear of anus |
| Colorectal cancer | 153 | ICD10CM | C19 | Malignant neoplasm of rectosigmoid junction |
| Colorectal cancer | 153 | ICD10CM | R85.612 | Low grade squamous intraepithelial lesion on cytologic smear of anus (LGSIL) |
| Colorectal cancer | 153 | ICD9CM | 796.71 | Papanicolaou smear of anus with atypical squamous cells of undetermined significance (ASC-US) |
| Colorectal cancer | 153 | ICD9CM | 796.7 | Abnormal glandular Papanicolaou smear of anus |
| Colorectal cancer | 153 | ICD9CM | 154 | Malignant neoplasm of rectosigmoid junction |
| Colorectal cancer | 153 | ICD9CM | 796.73 | Papanicolaou smear of anus with low grade squamous intraepithelial lesion (LGSIL) |
| Colorectal cancer | 153 | ICD10CM | R85.610 | Atypical squamous cells of undetermined significance on cytologic smear of anus (ASC-US) |
| Colorectal cancer | 153 | ICD10CM | R85.611 | Atypical squamous cells cannot exclude high grade squamous intraepithelial lesion on cytologic smear of anus (ASC-H) |
| Colorectal cancer | 153 | ICD10CM | C21.0 | Malignant neoplasm of anus, unspecified |
| Colorectal cancer | 153 | ICD9CM | 154.1 | Malignant neoplasm of rectum |
| Colorectal cancer | 153 | ICD10CM | D01.2 | Carcinoma in situ of rectum |
| Colorectal cancer | 153 | ICD9CM | 154.3 | Malignant neoplasm of anus, unspecified site |
| Colorectal cancer | 153 | ICD9CM | 796.74 | Papanicolaou smear of anus with high grade squamous intraepithelial lesion (HGSIL) |
| Colorectal cancer | 153 | ICD9CM | 796.76 | Papanicolaou smear of anus with cytologic evidence of malignancy |
| Colorectal cancer | 153 | ICD9CM | V10.06 | Personal history of malignant neoplasm of rectum, rectosigmoid junction, and anus |
| Colorectal cancer | 153 | ICD9CM | 209.17 | Malignant carcinoid tumor of the rectum |
| Colorectal cancer | 153 | ICD10CM | D01.1 | Carcinoma in situ of rectosigmoid junction |
| Colorectal cancer | 153 | ICD10CM | C20 | Malignant neoplasm of rectum |
| Colorectal cancer | 153 | ICD9CM | 796.72 | Papanicolaou smear of anus with atypical squamous cells cannot exclude high grade squamous intraepithelial lesion (ASC-H) |
| Colorectal cancer | 153 | ICD10CM | C7A.026 | Malignant carcinoid tumor of the rectum |
| Colorectal cancer | 153 | ICD9CM | 154 | Malignant neoplasm of rectum, rectosigmoid junction, and anus |
| Colorectal cancer | 153 | ICD10CM | D01.3 | Carcinoma in situ of anus and anal canal |
| Colorectal cancer | 153 | ICD9CM | 230.6 | Carcinoma in situ of anus, unspecified |
| Colorectal cancer | 153 | ICD9CM | 230.4 | Carcinoma in situ of rectum |
| Colorectal cancer | 153 | ICD9CM | 154.2 | Malignant neoplasm of anal canal |

| | | | | |
|---|---|---|---|---|
| Colorectal cancer | 153 | ICD10CM | Z85.040 | Personal history of malignant carcinoid tumor of rectum |
| Colorectal cancer | 153 | ICD10CM | R85.613 | High grade squamous intraepithelial lesion on cytologic smear of anus (HGSIL) |
| Colorectal cancer | 153 | ICD10CM | Z85.04 | Personal history of malignant neoplasm of rectum, rectosigmoid junction, and anus |
| Colorectal cancer | 153 | ICD9CM | 230.5 | Carcinoma in situ of anal canal |
| Colorectal cancer | 153 | ICD10CM | Z85.03 | Personal history of malignant neoplasm of large intestine |
| Cancer of bronchus; lung | 165.1 | ICD9CM | 162.8 | Malignant neoplasm of other parts of bronchus or lung |
| Cancer of bronchus; lung | 165.1 | ICD10CM | D02.22 | Carcinoma in situ of left bronchus and lung |
| Cancer of bronchus; lung | 165.1 | ICD10CM | C7A.090 | Malignant carcinoid tumor of the bronchus and lung |
| Cancer of bronchus; lung | 165.1 | ICD10CM | C34.3 | Malignant neoplasm of lower lobe, bronchus or lung |
| Cancer of bronchus; lung | 165.1 | ICD10CM | Z85.12 | Personal history of malignant neoplasm of trachea |
| Cancer of bronchus; lung | 165.1 | ICD10CM | C34 | Malignant neoplasm of bronchus and lung |
| Cancer of bronchus; lung | 165.1 | ICD10CM | C34.9 | Malignant neoplasm of unspecified part of bronchus or lung |
| Cancer of bronchus; lung | 165.1 | ICD10CM | C34.0 | Malignant neoplasm of main bronchus |
| Cancer of bronchus; lung | 165.1 | ICD9CM | 162.9 | Malignant neoplasm of bronchus and lung, unspecified |
| Cancer of bronchus; lung | 165.1 | ICD10CM | Z85.1 | Personal history of malignant neoplasm of trachea, bronchus and lung |
| Cancer of bronchus; lung | 165.1 | ICD10CM | C34.00 | Malignant neoplasm of unspecified main bronchus |
| Cancer of bronchus; lung | 165.1 | ICD10CM | C34.32 | Malignant neoplasm of lower lobe, left bronchus or lung |
| Cancer of bronchus; lung | 165.1 | ICD9CM | 162.2 | Malignant neoplasm of main bronchus |
| Cancer of bronchus; lung | 165.1 | ICD9CM | 162.3 | Malignant neoplasm of upper lobe, bronchus or lung |
| Cancer of bronchus; lung | 165.1 | ICD10CM | C34.30 | Malignant neoplasm of lower lobe, unspecified bronchus or lung |
| Cancer of bronchus; lung | 165.1 | ICD10CM | C34.12 | Malignant neoplasm of upper lobe, left bronchus or lung |
| Cancer of bronchus; lung | 165.1 | ICD10CM | C34.2 | Malignant neoplasm of middle lobe, bronchus or lung |
| Cancer of bronchus; lung | 165.1 | ICD10CM | C34.92 | Malignant neoplasm of unspecified part of left bronchus or lung |
| Cancer of bronchus; lung | 165.1 | ICD10CM | C34.02 | Malignant neoplasm of left main bronchus |
| Cancer of bronchus; lung | 165.1 | ICD10CM | C34.81 | Malignant neoplasm of overlapping sites of right bronchus and lung |
| Cancer of bronchus; lung | 165.1 | ICD9CM | 231.2 | Carcinoma in situ of bronchus and lung |

| | | | | |
|---|---|---|---|---|
| Cancer of bronchus; lung | 165.1 | ICD10CM | C34.1 | Malignant neoplasm of upper lobe, bronchus or lung |
| Cancer of bronchus; lung | 165.1 | ICD9CM | 162 | Malignant neoplasm of trachea |
| Cancer of bronchus; lung | 165.1 | ICD10CM | C34.8 | Malignant neoplasm of overlapping sites of bronchus and lung |
| Cancer of bronchus; lung | 165.1 | ICD9CM | 209.21 | Malignant carcinoid tumor of the bronchus and lung |
| Cancer of bronchus; lung | 165.1 | ICD10CM | Z85.11 | Personal history of malignant neoplasm of bronchus and lung |
| Cancer of bronchus; lung | 165.1 | ICD10CM | D02.21 | Carcinoma in situ of right bronchus and lung |
| Cancer of bronchus; lung | 165.1 | ICD10CM | D02.20 | Carcinoma in situ of unspecified bronchus and lung |
| Cancer of bronchus; lung | 165.1 | ICD10CM | Z85.118 | Personal history of other malignant neoplasm of bronchus and lung |
| Cancer of bronchus; lung | 165.1 | ICD10CM | C33 | Malignant neoplasm of trachea |
| Cancer of bronchus; lung | 165.1 | ICD10CM | C34.01 | Malignant neoplasm of right main bronchus |
| Cancer of bronchus; lung | 165.1 | ICD10CM | C34.80 | Malignant neoplasm of overlapping sites of unspecified bronchus and lung |
| Cancer of bronchus; lung | 165.1 | ICD10CM | C34.82 | Malignant neoplasm of overlapping sites of left bronchus and lung |
| Cancer of bronchus; lung | 165.1 | ICD10CM | C34.11 | Malignant neoplasm of upper lobe, right bronchus or lung |
| Cancer of bronchus; lung | 165.1 | ICD10CM | C34.31 | Malignant neoplasm of lower lobe, right bronchus or lung |
| Cancer of bronchus; lung | 165.1 | ICD10CM | C34.91 | Malignant neoplasm of unspecified part of right bronchus or lung |
| Cancer of bronchus; lung | 165.1 | ICD10CM | C34.90 | Malignant neoplasm of unspecified part of unspecified bronchus or lung |
| Cancer of bronchus; lung | 165.1 | ICD9CM | 162 | Malignant neoplasm of trachea, bronchus, and lung |
| Cancer of bronchus; lung | 165.1 | ICD9CM | V10.11 | Personal history of malignant neoplasm of bronchus and lung |
| Cancer of bronchus; lung | 165.1 | ICD9CM | 162.5 | Malignant neoplasm of lower lobe, bronchus or lung |
| Cancer of bronchus; lung | 165.1 | ICD10CM | D02.2 | Carcinoma in situ of bronchus and lung |
| Cancer of bronchus; lung | 165.1 | ICD10CM | C34.10 | Malignant neoplasm of upper lobe, unspecified bronchus or lung |
| Cancer of bronchus; lung | 165.1 | ICD10CM | Z85.110 | Personal history of malignant carcinoid tumor of bronchus and lung |
| Cancer of bronchus; lung | 165.1 | ICD9CM | 162.4 | Malignant neoplasm of middle lobe, bronchus or lung |
| Skin cancer | 172 | ICD10CM | Z86.007 | Personal history of in-situ neoplasm of skin |
| Skin cancer | 172 | ICD10CM | Z86.006 | Personal history of melanoma in-situ |
| Skin cancer | 172 | ICD9CM | 232 | Carcinoma in situ of skin |
| Skin cancer | 172 | ICD9CM | V10.82 | Personal history of malignant melanoma of skin |

| | | | | |
|---|---|---|---|---|
| Skin cancer | 172 | ICD10CM | Z85.820 | Personal history of malignant melanoma of skin |
| Skin cancer | 172 | ICD10CM | Z85.828 | Personal history of other malignant neoplasm of skin |
| Skin cancer | 172 | ICD10CM | Z85.821 | Personal history of Merkel cell carcinoma |
| Skin cancer | 172 | ICD10CM | C44.70 | Unspecified malignant neoplasm of skin of lower limb, including hip |
| Skin cancer | 172 | ICD10CM | C4A.59 | Merkel cell carcinoma of other part of trunk |
| Skin cancer | 172 | ICD10CM | C4A.70 | Merkel cell carcinoma of unspecified lower limb, including hip |
| Skin cancer | 172 | ICD10CM | C44.39 | Other specified malignant neoplasm of skin of other and unspecified parts of face |
| Skin cancer | 172 | ICD10CM | C43.22 | Malignant melanoma of left ear and external auricular canal |
| Skin cancer | 172 | ICD10CM | C43.11 | Malignant melanoma of right eyelid, including canthus |
| Skin cancer | 172 | ICD10CM | C43.52 | Malignant melanoma of skin of breast |
| Skin cancer | 172 | ICD10CM | D03.6 | Melanoma in situ of upper limb, including shoulder |
| Skin cancer | 172 | ICD10CM | C43.9 | Malignant melanoma of skin, unspecified |
| Skin cancer | 172 | ICD10CM | C43.51 | Malignant melanoma of anal skin |
| Skin cancer | 172 | ICD10CM | C43.72 | Malignant melanoma of left lower limb, including hip |
| Skin cancer | 172 | ICD10CM | D03.1 | Melanoma in situ of eyelid, including canthus |
| Skin cancer | 172 | ICD10CM | C43.2 | Malignant melanoma of ear and external auricular canal |
| Skin cancer | 172 | ICD10CM | C43.4 | Malignant melanoma of scalp and neck |
| Skin cancer | 172 | ICD10CM | D03.22 | Melanoma in situ of left ear and external auricular canal |
| Skin cancer | 172 | ICD10CM | C43.20 | Malignant melanoma of unspecified ear and external auricular canal |
| Skin cancer | 172 | ICD10CM | D03.59 | Melanoma in situ of other part of trunk |
| Skin cancer | 172 | ICD9CM | 172.9 | Melanoma of skin, site unspecified |
| Skin cancer | 172 | ICD10CM | C43.71 | Malignant melanoma of right lower limb, including hip |
| Skin cancer | 172 | ICD10CM | C43.12 | Malignant melanoma of left eyelid, including canthus |
| Skin cancer | 172 | ICD10CM | D03.20 | Melanoma in situ of unspecified ear and external auricular canal |
| Skin cancer | 172 | ICD10CM | D03.72 | Melanoma in situ of left lower limb, including hip |
| Skin cancer | 172 | ICD10CM | D03.4 | Melanoma in situ of scalp and neck |
| Skin cancer | 172 | ICD10CM | D03.8 | Melanoma in situ of other sites |
| Skin cancer | 172 | ICD10CM | D03.30 | Melanoma in situ of unspecified part of face |
| Skin cancer | 172 | ICD9CM | 172.6 | Malignant melanoma of skin of upper limb, including shoulder |
| Skin cancer | 172 | ICD10CM | C43.3 | Malignant melanoma of other and unspecified parts of face |

| | | | | |
|---|---|---|---|---|
| Skin cancer | 172 | ICD9CM | 172 | Malignant melanoma of skin |
| Skin cancer | 172 | ICD10CM | D03.0 | Melanoma in situ of lip |
| Skin cancer | 172 | ICD10CM | D03.2 | Melanoma in situ of ear and external auricular canal |
| Skin cancer | 172 | ICD10CM | C43.62 | Malignant melanoma of left upper limb, including shoulder |
| Skin cancer | 172 | ICD10CM | D03.51 | Melanoma in situ of anal skin |
| Skin cancer | 172 | ICD10CM | C43.8 | Malignant melanoma of overlapping sites of skin |
| Skin cancer | 172 | ICD9CM | 172.5 | Malignant melanoma of skin of trunk, except scrotum |
| Skin cancer | 172 | ICD10CM | C43.6 | Malignant melanoma of upper limb, including shoulder |
| Skin cancer | 172 | ICD10CM | D03.21 | Melanoma in situ of right ear and external auricular canal |
| Skin cancer | 172 | ICD10CM | D03.70 | Melanoma in situ of unspecified lower limb, including hip |
| Skin cancer | 172 | ICD10CM | D03.61 | Melanoma in situ of right upper limb, including shoulder |
| Skin cancer | 172 | ICD9CM | 172.2 | Malignant melanoma of skin of ear and external auditory canal |
| Skin cancer | 172 | ICD10CM | C43.7 | Malignant melanoma of lower limb, including hip |
| Skin cancer | 172 | ICD10CM | C43.61 | Malignant melanoma of right upper limb, including shoulder |
| Skin cancer | 172 | ICD10CM | C43.31 | Malignant melanoma of nose |
| Skin cancer | 172 | ICD10CM | D03.3 | Melanoma in situ of other and unspecified parts of face |
| Skin cancer | 172 | ICD10CM | C43.10 | Malignant melanoma of unspecified eyelid, including canthus |
| Skin cancer | 172 | ICD9CM | 172.8 | Malignant melanoma of other specified sites of skin |
| Skin cancer | 172 | ICD9CM | 172.3 | Malignant melanoma of skin of other and unspecified parts of face |
| Skin cancer | 172 | ICD10CM | D03.39 | Melanoma in situ of other parts of face |
| Skin cancer | 172 | ICD10CM | D03.12 | Melanoma in situ of left eyelid, including canthus |
| Skin cancer | 172 | ICD10CM | C43.21 | Malignant melanoma of right ear and external auricular canal |
| Skin cancer | 172 | ICD10CM | C43.1 | Malignant melanoma of eyelid, including canthus |
| Skin cancer | 172 | ICD10CM | D03.62 | Melanoma in situ of left upper limb, including shoulder |
| Skin cancer | 172 | ICD10CM | C43.60 | Malignant melanoma of unspecified upper limb, including shoulder |
| Skin cancer | 172 | ICD10CM | D03.71 | Melanoma in situ of right lower limb, including hip |
| Skin cancer | 172 | ICD10CM | D03.11 | Melanoma in situ of right eyelid, including canthus |
| Skin cancer | 172 | ICD10CM | D03.9 | Melanoma in situ, unspecified |
| Skin cancer | 172 | ICD10CM | C43 | Malignant melanoma of skin |

| | | | | |
|---|---|---|---|---|
| Skin cancer | 172 | ICD10CM | C43.0 | Malignant melanoma of lip |
| Skin cancer | 172 | ICD10CM | C43.70 | Malignant melanoma of unspecified lower limb, including hip |
| Skin cancer | 172 | ICD10CM | C43.59 | Malignant melanoma of other part of trunk |
| Skin cancer | 172 | ICD9CM | 172.4 | Malignant melanoma of skin of scalp and neck |
| Skin cancer | 172 | ICD10CM | C43.39 | Malignant melanoma of other parts of face |
| Skin cancer | 172 | ICD10CM | D03 | Melanoma in situ |
| Skin cancer | 172 | ICD9CM | 172 | Malignant melanoma of skin of lip |
| Skin cancer | 172 | ICD9CM | 172.1 | Malignant melanoma of skin of eyelid, including canthus |
| Skin cancer | 172 | ICD10CM | D03.52 | Melanoma in situ of breast (skin) (soft tissue) |
| Skin cancer | 172 | ICD10CM | D03.7 | Melanoma in situ of lower limb, including hip |
| Skin cancer | 172 | ICD10CM | C43.30 | Malignant melanoma of unspecified part of face |
| Skin cancer | 172 | ICD10CM | D03.5 | Melanoma in situ of trunk |
| Skin cancer | 172 | ICD10CM | D03.60 | Melanoma in situ of unspecified upper limb, including shoulder |
| Skin cancer | 172 | ICD10CM | C43.5 | Malignant melanoma of trunk |
| Skin cancer | 172 | ICD9CM | 172.7 | Malignant melanoma of skin of lower limb, including hip |
| Skin cancer | 172 | ICD10CM | D03.10 | Melanoma in situ of unspecified eyelid, including canthus |
| Skin cancer | 172 | ICD10CM | C44.310 | Basal cell carcinoma of skin of unspecified parts of face |
| Skin cancer | 172 | ICD10CM | C44.611 | Basal cell carcinoma of skin of unspecified upper limb, including shoulder |
| Skin cancer | 172 | ICD10CM | C44.519 | Basal cell carcinoma of skin of other part of trunk |
| Skin cancer | 172 | ICD9CM | 173.01 | Basal cell carcinoma of skin of lip |
| Skin cancer | 172 | ICD9CM | 173.51 | Basal cell carcinoma of skin of trunk, except scrotum |
| Skin cancer | 172 | ICD10CM | C44.211 | Basal cell carcinoma of skin of unspecified ear and external auricular canal |
| Skin cancer | 172 | ICD10CM | C44.511 | Basal cell carcinoma of skin of breast |
| Skin cancer | 172 | ICD10CM | C44.41 | Basal cell carcinoma of skin of scalp and neck |
| Skin cancer | 172 | ICD10CM | C44.212 | Basal cell carcinoma of skin of right ear and external auricular canal |
| Skin cancer | 172 | ICD9CM | 173.31 | Basal cell carcinoma of skin of other and unspecified parts of face |
| Skin cancer | 172 | ICD10CM | C44.219 | Basal cell carcinoma of skin of left ear and external auricular canal |
| Skin cancer | 172 | ICD10CM | C44.119 | Basal cell carcinoma of skin of left eyelid, including canthus |
| Skin cancer | 172 | ICD10CM | C44.112 | Basal cell carcinoma of skin of right eyelid, including canthus |
| Skin cancer | 172 | ICD9CM | 173.91 | Basal cell carcinoma of skin, site unspecified |

| | | | | |
|---|---|---|---|---|
| Skin cancer | 172 | ICD10CM | C44.719 | Basal cell carcinoma of skin of left lower limb, including hip |
| Skin cancer | 172 | ICD10CM | C44.49 | Other specified malignant neoplasm of skin of scalp and neck |
| Skin cancer | 172 | ICD10CM | C44.299 | Other specified malignant neoplasm of skin of left ear and external auricular canal |
| Skin cancer | 172 | ICD10CM | C4A.62 | Merkel cell carcinoma of left upper limb, including shoulder |
| Skin cancer | 172 | ICD10CM | C4A.12 | Merkel cell carcinoma of left eyelid, including canthus |
| Skin cancer | 172 | ICD9CM | 173.29 | Other specified malignant neoplasm of skin of ear and external auditory canal |
| Skin cancer | 172 | ICD10CM | C44.399 | Other specified malignant neoplasm of skin of other parts of face |
| Skin cancer | 172 | ICD9CM | 173.7 | Unspecified malignant neoplasm of skin of lower limb, including hip |
| Skin cancer | 172 | ICD10CM | C4A.7 | Merkel cell carcinoma of lower limb, including hip |
| Skin cancer | 172 | ICD10CM | C44.89 | Other specified malignant neoplasm of overlapping sites of skin |
| Skin cancer | 172 | ICD10CM | C44.72 | Squamous cell carcinoma of skin of lower limb, including hip |
| Skin cancer | 172 | ICD9CM | 173.2 | Other and unspecified malignant neoplasm of skin of ear and external auditory canal |
| Skin cancer | 172 | ICD9CM | 173.3 | Unspecified malignant neoplasm of skin of other and unspecified parts of face |
| Skin cancer | 172 | ICD10CM | C44.692 | Other specified malignant neoplasm of skin of right upper limb, including shoulder |
| Skin cancer | 172 | ICD10CM | C44.109 | Unspecified malignant neoplasm of skin of left eyelid, including canthus |
| Skin cancer | 172 | ICD10CM | C44.192 | Other specified malignant neoplasm of skin of right eyelid, including canthus |
| Skin cancer | 172 | ICD9CM | 173.8 | Unspecified malignant neoplasm of other specified sites of skin |
| Skin cancer | 172 | ICD10CM | C44.391 | Other specified malignant neoplasm of skin of nose |
| Skin cancer | 172 | ICD10CM | C4A.4 | Merkel cell carcinoma of scalp and neck |
| Skin cancer | 172 | ICD9CM | 173.5 | Unspecified malignant neoplasm of skin of trunk, except scrotum |
| Skin cancer | 172 | ICD10CM | C44.20 | Unspecified malignant neoplasm of skin of ear and external auricular canal |
| Skin cancer | 172 | ICD10CM | C4A.3 | Merkel cell carcinoma of other and unspecified parts of face |
| Skin cancer | 172 | ICD10CM | C44.79 | Other specified malignant neoplasm of skin of lower limb, including hip |
| Skin cancer | 172 | ICD10CM | C44.00 | Unspecified malignant neoplasm of skin of lip |
| Skin cancer | 172 | ICD9CM | 173.8 | Other and unspecified malignant neoplasm of other specified sites of skin |
| Skin cancer | 172 | ICD10CM | C44.301 | Unspecified malignant neoplasm of skin of nose |

| | | | | |
|---|---|---|---|---|
| Skin cancer | 172 | ICD10CM | C4A.61 | Merkel cell carcinoma of right upper limb, including shoulder |
| Skin cancer | 172 | ICD10CM | C44.500 | Unspecified malignant neoplasm of anal skin |
| Skin cancer | 172 | ICD10CM | C44.7 | Other and unspecified malignant neoplasm of skin of lower limb, including hip |
| Skin cancer | 172 | ICD10CM | C44.791 | Other specified malignant neoplasm of skin of unspecified lower limb, including hip |
| Skin cancer | 172 | ICD10CM | C44.4 | Other and unspecified malignant neoplasm of skin of scalp and neck |
| Skin cancer | 172 | ICD9CM | 173.3 | Other and unspecified malignant neoplasm of skin of other and unspecified parts of face |
| Skin cancer | 172 | ICD9CM | 173.89 | Other specified malignant neoplasm of other specified sites of skin |
| Skin cancer | 172 | ICD9CM | 173 | Unspecified malignant neoplasm of skin of lip |
| Skin cancer | 172 | ICD10CM | C4A.0 | Merkel cell carcinoma of lip |
| Skin cancer | 172 | ICD10CM | C44.292 | Other specified malignant neoplasm of skin of right ear and external auricular canal |
| Skin cancer | 172 | ICD10CM | C44.709 | Unspecified malignant neoplasm of skin of left lower limb, including hip |
| Skin cancer | 172 | ICD10CM | C44.601 | Unspecified malignant neoplasm of skin of unspecified upper limb, including shoulder |
| Skin cancer | 172 | ICD10CM | C4A.31 | Merkel cell carcinoma of nose |
| Skin cancer | 172 | ICD9CM | 209.36 | Merkel cell carcinoma of other sites |
| Skin cancer | 172 | ICD10CM | C4A.71 | Merkel cell carcinoma of right lower limb, including hip |
| Skin cancer | 172 | ICD10CM | C44.11 | Basal cell carcinoma of skin of eyelid, including canthus |
| Skin cancer | 172 | ICD9CM | 209.33 | Merkel cell carcinoma of the upper limb |
| Skin cancer | 172 | ICD10CM | C44.12 | Squamous cell carcinoma of skin of eyelid, including canthus |
| Skin cancer | 172 | ICD10CM | C4A.60 | Merkel cell carcinoma of unspecified upper limb, including shoulder |
| Skin cancer | 172 | ICD10CM | C4A.20 | Merkel cell carcinoma of unspecified ear and external auricular canal |
| Skin cancer | 172 | ICD9CM | 173.59 | Other specified malignant neoplasm of skin of trunk, except scrotum |
| Skin cancer | 172 | ICD10CM | C4A.52 | Merkel cell carcinoma of skin of breast |
| Skin cancer | 172 | ICD10CM | C44.71 | Basal cell carcinoma of skin of lower limb, including hip |
| Skin cancer | 172 | ICD10CM | C44.0 | Other and unspecified malignant neoplasm of skin of lip |
| Skin cancer | 172 | ICD10CM | C4A.10 | Merkel cell carcinoma of unspecified eyelid, including canthus |
| Skin cancer | 172 | ICD10CM | C4A.72 | Merkel cell carcinoma of left lower limb, including hip |
| Skin cancer | 172 | ICD10CM | C44.501 | Unspecified malignant neoplasm of skin of breast |
| Skin cancer | 172 | ICD9CM | 173.7 | Other and unspecified malignant neoplasm of skin of lower limb, including hip |

| | | | | |
|---|---|---|---|---|
| Skin cancer | 172 | ICD9CM | 173.11 | Basal cell carcinoma of eyelid, including canthus |
| Skin cancer | 172 | ICD10CM | C44.311 | Basal cell carcinoma of skin of nose |
| Skin cancer | 172 | ICD10CM | C44.612 | Basal cell carcinoma of skin of right upper limb, including shoulder |
| Skin cancer | 172 | ICD10CM | C44.91 | Basal cell carcinoma of skin, unspecified |
| Skin cancer | 172 | ICD9CM | 173.21 | Basal cell carcinoma of skin of ear and external auditory canal |
| Skin cancer | 172 | ICD10CM | C4A.39 | Merkel cell carcinoma of other parts of face |
| Skin cancer | 172 | ICD10CM | C44.29 | Other specified malignant neoplasm of skin of ear and external auricular canal |
| Skin cancer | 172 | ICD9CM | 173.2 | Unspecified malignant neoplasm of skin of ear and external auditory canal |
| Skin cancer | 172 | ICD10CM | C4A.8 | Merkel cell carcinoma of overlapping sites |
| Skin cancer | 172 | ICD10CM | C44.51 | Basal cell carcinoma of skin of trunk |
| Skin cancer | 172 | ICD10CM | C44.300 | Unspecified malignant neoplasm of skin of unspecified part of face |
| Skin cancer | 172 | ICD9CM | V10.83 | Personal history of other malignant neoplasm of skin |
| Skin cancer | 172 | ICD9CM | 173.49 | Other specified malignant neoplasm of scalp and skin of neck |
| Skin cancer | 172 | ICD10CM | C44.99 | Other specified malignant neoplasm of skin, unspecified |
| Skin cancer | 172 | ICD9CM | 209.35 | Merkel cell carcinoma of the trunk |
| Skin cancer | 172 | ICD10CM | C4A.6 | Merkel cell carcinoma of upper limb, including shoulder |
| Skin cancer | 172 | ICD10CM | C44.22 | Squamous cell carcinoma of skin of ear and external auricular canal |
| Skin cancer | 172 | ICD10CM | C44.609 | Unspecified malignant neoplasm of skin of left upper limb, including shoulder |
| Skin cancer | 172 | ICD10CM | C4A | Merkel cell carcinoma |
| Skin cancer | 172 | ICD10CM | C44.6 | Other and unspecified malignant neoplasm of skin of upper limb, including shoulder |
| Skin cancer | 172 | ICD10CM | C44.209 | Unspecified malignant neoplasm of skin of left ear and external auricular canal |
| Skin cancer | 172 | ICD9CM | 173.9 | Unspecified malignant neoplasm of skin, site unspecified |
| Skin cancer | 172 | ICD10CM | C44.309 | Unspecified malignant neoplasm of skin of other parts of face |
| Skin cancer | 172 | ICD9CM | 173.9 | Other and unspecified malignant neoplasm of skin, site unspecified |
| Skin cancer | 172 | ICD9CM | 173.6 | Other and unspecified malignant neoplasm of skin of upper limb, including shoulder |
| Skin cancer | 172 | ICD9CM | 173.6 | Unspecified malignant neoplasm of skin of upper limb, including shoulder |
| Skin cancer | 172 | ICD9CM | 173 | Other malignant neoplasm of skin of lip |
| Skin cancer | 172 | ICD10CM | C44.09 | Other specified malignant neoplasm of skin of lip |
| Skin cancer | 172 | ICD10CM | C4A.1 | Merkel cell carcinoma of eyelid, including canthus |

| | | | | |
|---|---|---|---|---|
| Skin cancer | 172 | ICD9CM | 173.81 | Basal cell carcinoma of other specified sites of skin |
| Skin cancer | 172 | ICD10CM | C44.81 | Basal cell carcinoma of overlapping sites of skin |
| Skin cancer | 172 | ICD9CM | 173.61 | Basal cell carcinoma of skin of upper limb, including shoulder |
| Skin cancer | 172 | ICD10CM | C44.111 | Basal cell carcinoma of skin of unspecified eyelid, including canthus |
| Skin cancer | 172 | ICD10CM | C44.01 | Basal cell carcinoma of skin of lip |
| Skin cancer | 172 | ICD10CM | C44.510 | Basal cell carcinoma of anal skin |
| Skin cancer | 172 | ICD10CM | C44.712 | Basal cell carcinoma of skin of right lower limb, including hip |
| Skin cancer | 172 | ICD10CM | C44.619 | Basal cell carcinoma of skin of left upper limb, including shoulder |
| Skin cancer | 172 | ICD9CM | 173.41 | Basal cell carcinoma of scalp and skin of neck |
| Skin cancer | 172 | ICD9CM | 173.71 | Basal cell carcinoma of skin of lower limb, including hip |
| Skin cancer | 172 | ICD10CM | C44.591 | Other specified malignant neoplasm of skin of breast |
| Skin cancer | 172 | ICD9CM | 173.5 | Other and unspecified malignant neoplasm of skin of trunk, except scrotum |
| Skin cancer | 172 | ICD10CM | D04.2 | Carcinoma in situ of skin of ear and external auricular canal |
| Skin cancer | 172 | ICD10CM | D04.3 | Carcinoma in situ of skin of other and unspecified parts of face |
| Skin cancer | 172 | ICD10CM | C44.711 | Basal cell carcinoma of skin of unspecified lower limb, including hip |
| Skin cancer | 172 | ICD10CM | C44.319 | Basal cell carcinoma of skin of other parts of face |
| Skin cancer | 172 | ICD10CM | C44.60 | Unspecified malignant neoplasm of skin of upper limb, including shoulder |
| Skin cancer | 172 | ICD10CM | C44.61 | Basal cell carcinoma of skin of upper limb, including shoulder |
| Skin cancer | 172 | ICD10CM | C4A.2 | Merkel cell carcinoma of ear and external auricular canal |
| Skin cancer | 172 | ICD10CM | C44.8 | Other and unspecified malignant neoplasm of overlapping sites of skin |
| Skin cancer | 172 | ICD10CM | C44.390 | Other specified malignant neoplasm of skin of unspecified parts of face |
| Skin cancer | 172 | ICD10CM | C44.701 | Unspecified malignant neoplasm of skin of unspecified lower limb, including hip |
| Skin cancer | 172 | ICD10CM | C4A.11 | Merkel cell carcinoma of right eyelid, including canthus |
| Skin cancer | 172 | ICD10CM | C4A.21 | Merkel cell carcinoma of right ear and external auricular canal |
| Skin cancer | 172 | ICD9CM | 173.1 | Other and unspecified malignant neoplasm of skin of eyelid, including canthus |
| Skin cancer | 172 | ICD10CM | C44.2 | Other and unspecified malignant neoplasm of skin of ear and external auricular canal |
| Skin cancer | 172 | ICD9CM | 173 | Other and unspecified malignant neoplasm of skin |

| | | | | |
|---|---|---|---|---|
| Skin cancer | 172 | ICD10CM | C44.21 | Basal cell carcinoma of skin of ear and external auricular canal |
| Skin cancer | 172 | ICD10CM | C44.40 | Unspecified malignant neoplasm of skin of scalp and neck |
| Skin cancer | 172 | ICD9CM | 173.69 | Other specified malignant neoplasm of skin of upper limb, including shoulder |
| Skin cancer | 172 | ICD10CM | C44.69 | Other specified malignant neoplasm of skin of upper limb, including shoulder |
| Skin cancer | 172 | ICD10CM | C44.102 | Unspecified malignant neoplasm of skin of right eyelid, including canthus |
| Skin cancer | 172 | ICD10CM | C44.101 | Unspecified malignant neoplasm of skin of unspecified eyelid, including canthus |
| Skin cancer | 172 | ICD10CM | C4A.30 | Merkel cell carcinoma of unspecified part of face |
| Skin cancer | 172 | ICD10CM | C44.799 | Other specified malignant neoplasm of skin of left lower limb, including hip |
| Skin cancer | 172 | ICD10CM | C4A.5 | Merkel cell carcinoma of trunk |
| Skin cancer | 172 | ICD9CM | 209.34 | Merkel cell carcinoma of the lower limb |
| Skin cancer | 172 | ICD10CM | C44.90 | Unspecified malignant neoplasm of skin, unspecified |
| Skin cancer | 172 | ICD10CM | C4A.22 | Merkel cell carcinoma of left ear and external auricular canal |
| Skin cancer | 172 | ICD9CM | 173.19 | Other specified malignant neoplasm of eyelid, including canthus |
| Skin cancer | 172 | ICD10CM | C44.201 | Unspecified malignant neoplasm of skin of unspecified ear and external auricular canal |
| Skin cancer | 172 | ICD10CM | C44.52 | Squamous cell carcinoma of skin of trunk |
| Skin cancer | 172 | ICD10CM | C4A.51 | Merkel cell carcinoma of anal skin |
| Skin cancer | 172 | ICD10CM | C44.62 | Squamous cell carcinoma of skin of upper limb, including shoulder |
| Skin cancer | 172 | ICD9CM | 173.4 | Unspecified malignant neoplasm of scalp and skin of neck |
| Skin cancer | 172 | ICD10CM | C44.590 | Other specified malignant neoplasm of anal skin |
| Skin cancer | 172 | ICD10CM | C44.80 | Unspecified malignant neoplasm of overlapping sites of skin |
| Skin cancer | 172 | ICD10CM | C44.10 | Unspecified malignant neoplasm of skin of eyelid, including canthus |
| Skin cancer | 172 | ICD10CM | C44.691 | Other specified malignant neoplasm of skin of unspecified upper limb, including shoulder |
| Skin cancer | 172 | ICD10CM | C44.602 | Unspecified malignant neoplasm of skin of right upper limb, including shoulder |
| Skin cancer | 172 | ICD9CM | 173.79 | Other specified malignant neoplasm of skin of lower limb, including hip |
| Skin cancer | 172 | ICD10CM | C44.9 | Other and unspecified malignant neoplasm of skin, unspecified |
| Skin cancer | 172 | ICD10CM | D04.11 | Carcinoma in situ of skin of right eyelid, including canthus |
| Skin cancer | 172 | ICD10CM | D04.10 | Carcinoma in situ of skin of unspecified eyelid, including canthus |

| | | | | |
|---|---|---|---|---|
| Skin cancer | 172 | ICD10CM | D04.22 | Carcinoma in situ of skin of left ear and external auricular canal |
| Skin cancer | 172 | ICD10CM | D04.0 | Carcinoma in situ of skin of lip |
| Skin cancer | 172 | ICD10CM | D04.1 | Carcinoma in situ of skin of eyelid, including canthus |
| Skin cancer | 172 | ICD10CM | D04.39 | Carcinoma in situ of skin of other parts of face |
| Skin cancer | 172 | ICD10CM | C4A.9 | Merkel cell carcinoma, unspecified |
| Skin cancer | 172 | ICD10CM | C44.59 | Other specified malignant neoplasm of skin of trunk |
| Skin cancer | 172 | ICD9CM | 173.39 | Other specified malignant neoplasm of skin of other and unspecified parts of face |
| Skin cancer | 172 | ICD9CM | 173.09 | Other specified malignant neoplasm of skin of lip |
| Skin cancer | 172 | ICD10CM | C44.599 | Other specified malignant neoplasm of skin of other part of trunk |
| Skin cancer | 172 | ICD10CM | C44.699 | Other specified malignant neoplasm of skin of left upper limb, including shoulder |
| Skin cancer | 172 | ICD9CM | 173.1 | Unspecified malignant neoplasm of eyelid, including canthus |
| Skin cancer | 172 | ICD10CM | C44.1 | Other and unspecified malignant neoplasm of skin of eyelid, including canthus |
| Skin cancer | 172 | ICD10CM | C44.19 | Other specified malignant neoplasm of skin of eyelid, including canthus |
| Skin cancer | 172 | ICD10CM | C44.792 | Other specified malignant neoplasm of skin of right lower limb, including hip |
| Skin cancer | 172 | ICD9CM | 209.31 | Merkel cell carcinoma of the face |
| Skin cancer | 172 | ICD10CM | D04.61 | Carcinoma in situ of skin of right upper limb, including shoulder |
| Skin cancer | 172 | ICD10CM | D04.9 | Carcinoma in situ of skin, unspecified |
| Skin cancer | 172 | ICD10CM | D04 | Carcinoma in situ of skin |
| Skin cancer | 172 | ICD10CM | D04.20 | Carcinoma in situ of skin of unspecified ear and external auricular canal |
| Skin cancer | 172 | ICD10CM | C44.509 | Unspecified malignant neoplasm of skin of other part of trunk |
| Skin cancer | 172 | ICD10CM | C44.199 | Other specified malignant neoplasm of skin of left eyelid, including canthus |
| Skin cancer | 172 | ICD9CM | 173.4 | Other and unspecified malignant neoplasm of scalp and skin of neck |
| Skin cancer | 172 | ICD10CM | C44.30 | Unspecified malignant neoplasm of skin of other and unspecified parts of face |
| Skin cancer | 172 | ICD10CM | C44.702 | Unspecified malignant neoplasm of skin of right lower limb, including hip |
| Skin cancer | 172 | ICD10CM | C44.31 | Basal cell carcinoma of skin of other and unspecified parts of face |
| Skin cancer | 172 | ICD10CM | C44.291 | Other specified malignant neoplasm of skin of unspecified ear and external auricular canal |
| Skin cancer | 172 | ICD9CM | 209.32 | Merkel cell carcinoma of the scalp and neck |
| Skin cancer | 172 | ICD9CM | 173.99 | Other specified malignant neoplasm of skin, site unspecified |

| | | | | |
|---|---|---|---|---|
| Skin cancer | 172 | ICD10CM | D04.4 | Carcinoma in situ of skin of scalp and neck |
| Skin cancer | 172 | ICD10CM | D04.62 | Carcinoma in situ of skin of left upper limb, including shoulder |
| Skin cancer | 172 | ICD9CM | 232.3 | Carcinoma in situ of skin of other and unspecified parts of face |
| Skin cancer | 172 | ICD9CM | 232 | Carcinoma in situ of skin of lip |
| Skin cancer | 172 | ICD10CM | C44.191 | Other specified malignant neoplasm of skin of unspecified eyelid, including canthus |
| Skin cancer | 172 | ICD10CM | C44.202 | Unspecified malignant neoplasm of skin of right ear and external auricular canal |
| Skin cancer | 172 | ICD9CM | 232.5 | Carcinoma in situ of skin of trunk, except scrotum |
| Skin cancer | 172 | ICD10CM | C44.722 | Squamous cell carcinoma of skin of right lower limb, including hip |
| Skin cancer | 172 | ICD10CM | C44.521 | Squamous cell carcinoma of skin of breast |
| Skin cancer | 172 | ICD10CM | D04.60 | Carcinoma in situ of skin of unspecified upper limb, including shoulder |
| Skin cancer | 172 | ICD9CM | 232.7 | Carcinoma in situ of skin of lower limb, including hip |
| Skin cancer | 172 | ICD10CM | D04.72 | Carcinoma in situ of skin of left lower limb, including hip |
| Skin cancer | 172 | ICD9CM | 232.6 | Carcinoma in situ of skin of upper limb, including shoulder |
| Skin cancer | 172 | ICD10CM | D04.12 | Carcinoma in situ of skin of left eyelid, including canthus |
| Skin cancer | 172 | ICD9CM | 232.1 | Carcinoma in situ of eyelid, including canthus |
| Skin cancer | 172 | ICD10CM | D04.6 | Carcinoma in situ of skin of upper limb, including shoulder |
| Skin cancer | 172 | ICD10CM | D04.8 | Carcinoma in situ of skin of other sites |
| Skin cancer | 172 | ICD10CM | D04.71 | Carcinoma in situ of skin of right lower limb, including hip |
| Skin cancer | 172 | ICD10CM | D04.70 | Carcinoma in situ of skin of unspecified lower limb, including hip |
| Skin cancer | 172 | ICD9CM | 173.62 | Squamous cell carcinoma of skin of upper limb, including shoulder |
| Skin cancer | 172 | ICD10CM | C44.321 | Squamous cell carcinoma of skin of nose |
| Skin cancer | 172 | ICD9CM | 173.52 | Squamous cell carcinoma of skin of trunk, except scrotum |
| Skin cancer | 172 | ICD10CM | C44.122 | Squamous cell carcinoma of skin of right eyelid, including canthus |
| Skin cancer | 172 | ICD9CM | 232.2 | Carcinoma in situ of skin of ear and external auditory canal |
| Skin cancer | 172 | ICD9CM | 173.22 | Squamous cell carcinoma of skin of ear and external auditory canal |
| Skin cancer | 172 | ICD10CM | D04.30 | Carcinoma in situ of skin of unspecified part of face |
| Skin cancer | 172 | ICD9CM | 232.4 | Carcinoma in situ of scalp and skin of neck |
| Skin cancer | 172 | ICD9CM | 232.8 | Carcinoma in situ of other specified sites of skin |

| | | | | |
|---|---|---|---|---|
| Skin cancer | 172 | ICD9CM | 173.32 | Squamous cell carcinoma of skin of other and unspecified parts of face |
| Skin cancer | 172 | ICD10CM | C44.629 | Squamous cell carcinoma of skin of left upper limb, including shoulder |
| Skin cancer | 172 | ICD10CM | C44.42 | Squamous cell carcinoma of skin of scalp and neck |
| Skin cancer | 172 | ICD10CM | D04.5 | Carcinoma in situ of skin of trunk |
| Skin cancer | 172 | ICD9CM | 232.9 | Carcinoma in situ of skin, site unspecified |
| Skin cancer | 172 | ICD10CM | C44.121 | Squamous cell carcinoma of skin of unspecified eyelid, including canthus |
| Skin cancer | 172 | ICD10CM | C44.320 | Squamous cell carcinoma of skin of unspecified parts of face |
| Skin cancer | 172 | ICD10CM | C44.229 | Squamous cell carcinoma of skin of left ear and external auricular canal |
| Skin cancer | 172 | ICD10CM | C44.621 | Squamous cell carcinoma of skin of unspecified upper limb, including shoulder |
| Skin cancer | 172 | ICD10CM | C44.329 | Squamous cell carcinoma of skin of other parts of face |
| Skin cancer | 172 | ICD10CM | C44.721 | Squamous cell carcinoma of skin of unspecified lower limb, including hip |
| Skin cancer | 172 | ICD9CM | 173.72 | Squamous cell carcinoma of skin of lower limb, including hip |
| Skin cancer | 172 | ICD10CM | D04.21 | Carcinoma in situ of skin of right ear and external auricular canal |
| Skin cancer | 172 | ICD9CM | 173.02 | Squamous cell carcinoma of skin of lip |
| Skin cancer | 172 | ICD10CM | C44.222 | Squamous cell carcinoma of skin of right ear and external auricular canal |
| Skin cancer | 172 | ICD10CM | C44.729 | Squamous cell carcinoma of skin of left lower limb, including hip |
| Skin cancer | 172 | ICD10CM | C44.520 | Squamous cell carcinoma of anal skin |
| Skin cancer | 172 | ICD10CM | C44.221 | Squamous cell carcinoma of skin of unspecified ear and external auricular canal |
| Skin cancer | 172 | ICD10CM | C44.92 | Squamous cell carcinoma of skin, unspecified |
| Skin cancer | 172 | ICD10CM | C44.02 | Squamous cell carcinoma of skin of lip |
| Skin cancer | 172 | ICD9CM | 173.82 | Squamous cell carcinoma of other specified sites of skin |
| Skin cancer | 172 | ICD10CM | C44.529 | Squamous cell carcinoma of skin of other part of trunk |
| Skin cancer | 172 | ICD9CM | 173.92 | Squamous cell carcinoma of skin, site unspecified |
| Skin cancer | 172 | ICD10CM | C44.622 | Squamous cell carcinoma of skin of right upper limb, including shoulder |
| Skin cancer | 172 | ICD10CM | D04.7 | Carcinoma in situ of skin of lower limb, including hip |
| Skin cancer | 172 | ICD10CM | C44.129 | Squamous cell carcinoma of skin of left eyelid, including canthus |
| Skin cancer | 172 | ICD9CM | 173.12 | Squamous cell carcinoma of eyelid, including canthus |

| | | | | |
|---|---|---|---|---|
| Skin cancer | 172 | ICD9CM | 173.42 | Squamous cell carcinoma of scalp and skin of neck |
| Skin cancer | 172 | ICD10CM | C44.82 | Squamous cell carcinoma of overlapping sites of skin |
| Melanomas of skin | 172.11 | ICD9CM | 172.7 | Malignant melanoma of skin of lower limb, including hip |
| Melanomas of skin | 172.11 | ICD10CM | D03.10 | Melanoma in situ of unspecified eyelid, including canthus |
| Melanomas of skin | 172.11 | ICD10CM | D03.7 | Melanoma in situ of lower limb, including hip |
| Melanomas of skin | 172.11 | ICD10CM | C43.30 | Malignant melanoma of unspecified part of face |
| Melanomas of skin | 172.11 | ICD10CM | C43.5 | Malignant melanoma of trunk |
| Melanomas of skin | 172.11 | ICD9CM | 172.8 | Malignant melanoma of other specified sites of skin |
| Melanomas of skin | 172.11 | ICD9CM | 172.3 | Malignant melanoma of skin of other and unspecified parts of face |
| Melanomas of skin | 172.11 | ICD10CM | D03.52 | Melanoma in situ of breast (skin) (soft tissue) |
| Melanomas of skin | 172.11 | ICD10CM | D03.12 | Melanoma in situ of left eyelid, including canthus |
| Melanomas of skin | 172.11 | ICD10CM | C43.21 | Malignant melanoma of right ear and external auricular canal |
| Melanomas of skin | 172.11 | ICD10CM | C43.1 | Malignant melanoma of eyelid, including canthus |
| Melanomas of skin | 172.11 | ICD10CM | D03.39 | Melanoma in situ of other parts of face |
| Melanomas of skin | 172.11 | ICD9CM | 172.1 | Malignant melanoma of skin of eyelid, including canthus |
| Melanomas of skin | 172.11 | ICD10CM | D03.0 | Melanoma in situ of lip |
| Melanomas of skin | 172.11 | ICD10CM | D03.5 | Melanoma in situ of trunk |
| Melanomas of skin | 172.11 | ICD10CM | D03.60 | Melanoma in situ of unspecified upper limb, including shoulder |
| Melanomas of skin | 172.11 | ICD10CM | C43.62 | Malignant melanoma of left upper limb, including shoulder |
| Melanomas of skin | 172.11 | ICD10CM | D03.51 | Melanoma in situ of anal skin |
| Melanomas of skin | 172.11 | ICD10CM | C43.8 | Malignant melanoma of overlapping sites of skin |
| Melanomas of skin | 172.11 | ICD9CM | 172.5 | Malignant melanoma of skin of trunk, except scrotum |
| Melanomas of skin | 172.11 | ICD10CM | C43.6 | Malignant melanoma of upper limb, including shoulder |
| Melanomas of skin | 172.11 | ICD10CM | D03.21 | Melanoma in situ of right ear and external auricular canal |
| Melanomas of skin | 172.11 | ICD10CM | D03.70 | Melanoma in situ of unspecified lower limb, including hip |
| Melanomas of skin | 172.11 | ICD10CM | D03.61 | Melanoma in situ of right upper limb, including shoulder |
| Melanomas of skin | 172.11 | ICD10CM | D03.2 | Melanoma in situ of ear and external auricular canal |
| Melanomas of skin | 172.11 | ICD10CM | C43.61 | Malignant melanoma of right upper limb, including shoulder |
| Melanomas of skin | 172.11 | ICD10CM | C43.31 | Malignant melanoma of nose |

| | | | | |
|---|---|---|---|---|
| Melanomas of skin | 172.11 | ICD10CM | D03.3 | Melanoma in situ of other and unspecified parts of face |
| Melanomas of skin | 172.11 | ICD10CM | C43.10 | Malignant melanoma of unspecified eyelid, including canthus |
| Melanomas of skin | 172.11 | ICD10CM | D03.22 | Melanoma in situ of left ear and external auricular canal |
| Melanomas of skin | 172.11 | ICD10CM | C43.20 | Malignant melanoma of unspecified ear and external auricular canal |
| Melanomas of skin | 172.11 | ICD10CM | D03.59 | Melanoma in situ of other part of trunk |
| Melanomas of skin | 172.11 | ICD10CM | D03.62 | Melanoma in situ of left upper limb, including shoulder |
| Melanomas of skin | 172.11 | ICD10CM | C43.60 | Malignant melanoma of unspecified upper limb, including shoulder |
| Melanomas of skin | 172.11 | ICD10CM | D03.71 | Melanoma in situ of right lower limb, including hip |
| Melanomas of skin | 172.11 | ICD9CM | 172.2 | Malignant melanoma of skin of ear and external auditory canal |
| Melanomas of skin | 172.11 | ICD10CM | C43.7 | Malignant melanoma of lower limb, including hip |
| Melanomas of skin | 172.11 | ICD10CM | C43 | Malignant melanoma of skin |
| Melanomas of skin | 172.11 | ICD10CM | C43.0 | Malignant melanoma of lip |
| Melanomas of skin | 172.11 | ICD10CM | C43.70 | Malignant melanoma of unspecified lower limb, including hip |
| Melanomas of skin | 172.11 | ICD10CM | C43.59 | Malignant melanoma of other part of trunk |
| Melanomas of skin | 172.11 | ICD9CM | 172.4 | Malignant melanoma of skin of scalp and neck |
| Melanomas of skin | 172.11 | ICD10CM | C43.39 | Malignant melanoma of other parts of face |
| Melanomas of skin | 172.11 | ICD10CM | D03 | Melanoma in situ |
| Melanomas of skin | 172.11 | ICD9CM | 172 | Malignant melanoma of skin of lip |
| Melanomas of skin | 172.11 | ICD10CM | C43.72 | Malignant melanoma of left lower limb, including hip |
| Melanomas of skin | 172.11 | ICD10CM | D03.1 | Melanoma in situ of eyelid, including canthus |
| Melanomas of skin | 172.11 | ICD10CM | C43.2 | Malignant melanoma of ear and external auricular canal |
| Melanomas of skin | 172.11 | ICD10CM | C43.4 | Malignant melanoma of scalp and neck |
| Melanomas of skin | 172.11 | ICD10CM | C43.22 | Malignant melanoma of left ear and external auricular canal |
| Melanomas of skin | 172.11 | ICD10CM | C43.11 | Malignant melanoma of right eyelid, including canthus |
| Melanomas of skin | 172.11 | ICD10CM | C43.52 | Malignant melanoma of skin of breast |
| Melanomas of skin | 172.11 | ICD10CM | D03.6 | Melanoma in situ of upper limb, including shoulder |
| Melanomas of skin | 172.11 | ICD10CM | C43.9 | Malignant melanoma of skin, unspecified |
| Melanomas of skin | 172.11 | ICD10CM | C43.51 | Malignant melanoma of anal skin |
| Melanomas of skin | 172.11 | ICD10CM | C43.71 | Malignant melanoma of right lower limb, including hip |
| Melanomas of skin | 172.11 | ICD10CM | C43.12 | Malignant melanoma of left eyelid, including canthus |

| | | | | |
|---|---|---|---|---|
| Melanomas of skin | 172.11 | ICD10CM | D03.11 | Melanoma in situ of right eyelid, including canthus |
| Melanomas of skin | 172.11 | ICD10CM | D03.9 | Melanoma in situ, unspecified |
| Melanomas of skin | 172.11 | ICD10CM | D03.72 | Melanoma in situ of left lower limb, including hip |
| Melanomas of skin | 172.11 | ICD10CM | D03.4 | Melanoma in situ of scalp and neck |
| Melanomas of skin | 172.11 | ICD10CM | D03.30 | Melanoma in situ of unspecified part of face |
| Melanomas of skin | 172.11 | ICD9CM | 172.6 | Malignant melanoma of skin of upper limb, including shoulder |
| Melanomas of skin | 172.11 | ICD10CM | C43.3 | Malignant melanoma of other and unspecified parts of face |
| Melanomas of skin | 172.11 | ICD9CM | 172.9 | Melanoma of skin, site unspecified |
| Melanomas of skin | 172.11 | ICD10CM | D03.20 | Melanoma in situ of unspecified ear and external auricular canal |
| Melanomas of skin | 172.11 | ICD10CM | D03.8 | Melanoma in situ of other sites |
| Melanomas of skin | 172.11 | ICD9CM | 172 | Malignant melanoma of skin |
| Breast cancer | 174 | ICD9CM | 233 | Carcinoma in situ of breast and genitourinary system |
| Breast cancer | 174 | ICD10CM | D05.10 | Intraductal carcinoma in situ of unspecified breast |
| Breast cancer | 174 | ICD10CM | D05.9 | Unspecified type of carcinoma in situ of breast |
| Breast cancer | 174 | ICD10CM | D05.00 | Lobular carcinoma in situ of unspecified breast |
| Breast cancer | 174 | ICD10CM | D05.0 | Lobular carcinoma in situ of breast |
| Breast cancer | 174 | ICD10CM | D05.1 | Intraductal carcinoma in situ of breast |
| Breast cancer | 174 | ICD10CM | D05.81 | Other specified type of carcinoma in situ of right breast |
| Breast cancer | 174 | ICD10CM | D05.02 | Lobular carcinoma in situ of left breast |
| Breast cancer | 174 | ICD10CM | D05.01 | Lobular carcinoma in situ of right breast |
| Breast cancer | 174 | ICD10CM | D05.80 | Other specified type of carcinoma in situ of unspecified breast |
| Breast cancer | 174 | ICD10CM | D05.92 | Unspecified type of carcinoma in situ of left breast |
| Breast cancer | 174 | ICD10CM | D05.91 | Unspecified type of carcinoma in situ of right breast |
| Breast cancer | 174 | ICD10CM | D05 | Carcinoma in situ of breast |
| Breast cancer | 174 | ICD10CM | D05.12 | Intraductal carcinoma in situ of left breast |
| Breast cancer | 174 | ICD10CM | D05.8 | Other specified type of carcinoma in situ of breast |
| Breast cancer | 174 | ICD10CM | D05.90 | Unspecified type of carcinoma in situ of unspecified breast |
| Breast cancer | 174 | ICD9CM | 233 | Carcinoma in situ of breast |
| Breast cancer | 174 | ICD10CM | D05.11 | Intraductal carcinoma in situ of right breast |
| Breast cancer | 174 | ICD10CM | D05.82 | Other specified type of carcinoma in situ of left breast |
| Breast cancer | 174 | ICD10CM | C50.62 | Malignant neoplasm of axillary tail of breast, male |

| | | | | |
|---|---|---|---|---|
| Breast cancer | 174 | ICD10CM | C50.22 | Malignant neoplasm of upper-inner quadrant of breast, male |
| Breast cancer | 174 | ICD10CM | C50.92 | Malignant neoplasm of breast of unspecified site, male |
| Breast cancer | 174 | ICD9CM | 175 | Malignant neoplasm of male breast |
| Breast cancer | 174 | ICD10CM | C50.829 | Malignant neoplasm of overlapping sites of unspecified male breast |
| Breast cancer | 174 | ICD10CM | C50.021 | Malignant neoplasm of nipple and areola, right male breast |
| Breast cancer | 174 | ICD9CM | 175.9 | Malignant neoplasm of other and unspecified sites of male breast |
| Breast cancer | 174 | ICD10CM | C50.822 | Malignant neoplasm of overlapping sites of left male breast |
| Breast cancer | 174 | ICD10CM | C50.32 | Malignant neoplasm of lower-inner quadrant of breast, male |
| Breast cancer | 174 | ICD10CM | C50.52 | Malignant neoplasm of lower-outer quadrant of breast, male |
| Breast cancer | 174 | ICD10CM | C50.12 | Malignant neoplasm of central portion of breast, male |
| Breast cancer | 174 | ICD10CM | C50.521 | Malignant neoplasm of lower-outer quadrant of right male breast |
| Breast cancer | 174 | ICD10CM | C50.622 | Malignant neoplasm of axillary tail of left male breast |
| Breast cancer | 174 | ICD10CM | C50.91 | Malignant neoplasm of breast of unspecified site, female |
| Breast cancer | 174 | ICD9CM | 174 | Malignant neoplasm of female breast |
| Breast cancer | 174 | ICD10CM | C50.51 | Malignant neoplasm of lower-outer quadrant of breast, female |
| Breast cancer | 174 | ICD10CM | C50.519 | Malignant neoplasm of lower-outer quadrant of unspecified female breast |
| Breast cancer | 174 | ICD10CM | C50.812 | Malignant neoplasm of overlapping sites of left female breast |
| Breast cancer | 174 | ICD10CM | C50.919 | Malignant neoplasm of unspecified site of unspecified female breast |
| Breast cancer | 174 | ICD10CM | C50.419 | Malignant neoplasm of upper-outer quadrant of unspecified female breast |
| Breast cancer | 174 | ICD10CM | C50.811 | Malignant neoplasm of overlapping sites of right female breast |
| Breast cancer | 174 | ICD10CM | C50.112 | Malignant neoplasm of central portion of left female breast |
| Breast cancer | 174 | ICD10CM | C50.012 | Malignant neoplasm of nipple and areola, left female breast |
| Breast cancer | 174 | ICD9CM | 174.6 | Malignant neoplasm of axillary tail of female breast |
| Breast cancer | 174 | ICD9CM | 174.4 | Malignant neoplasm of upper-outer quadrant of female breast |
| Breast cancer | 174 | ICD10CM | C50.211 | Malignant neoplasm of upper-inner quadrant of right female breast |
| Breast cancer | 174 | ICD10CM | C50.8 | Malignant neoplasm of overlapping sites of breast |

| | | | | |
|---|---|---|---|---|
| Breast cancer | 174 | ICD10CM | C50.41 | Malignant neoplasm of upper-outer quadrant of breast, female |
| Breast cancer | 174 | ICD10CM | C50.511 | Malignant neoplasm of lower-outer quadrant of right female breast |
| Breast cancer | 174 | ICD10CM | C50.512 | Malignant neoplasm of lower-outer quadrant of left female breast |
| Breast cancer | 174 | ICD10CM | C50.911 | Malignant neoplasm of unspecified site of right female breast |
| Breast cancer | 174 | ICD10CM | C50.212 | Malignant neoplasm of upper-inner quadrant of left female breast |
| Breast cancer | 174 | ICD10CM | C50.411 | Malignant neoplasm of upper-outer quadrant of right female breast |
| Breast cancer | 174 | ICD10CM | C50.319 | Malignant neoplasm of lower-inner quadrant of unspecified female breast |
| Breast cancer | 174 | ICD10CM | C50.612 | Malignant neoplasm of axillary tail of left female breast |
| Breast cancer | 174 | ICD10CM | C50.31 | Malignant neoplasm of lower-inner quadrant of breast, female |
| Breast cancer | 174 | ICD9CM | 174 | Malignant neoplasm of nipple and areola of female breast |
| Breast cancer | 174 | ICD9CM | 174.2 | Malignant neoplasm of upper-inner quadrant of female breast |
| Breast cancer | 174 | ICD10CM | C50.311 | Malignant neoplasm of lower-inner quadrant of right female breast |
| Breast cancer | 174 | ICD10CM | C50.011 | Malignant neoplasm of nipple and areola, right female breast |
| Breast cancer | 174 | ICD10CM | C50.21 | Malignant neoplasm of upper-inner quadrant of breast, female |
| Breast cancer | 174 | ICD10CM | C50.611 | Malignant neoplasm of axillary tail of right female breast |
| Breast cancer | 174 | ICD10CM | C50.81 | Malignant neoplasm of overlapping sites of breast, female |
| Breast cancer | 174 | ICD10CM | C50.019 | Malignant neoplasm of nipple and areola, unspecified female breast |
| Breast cancer | 174 | ICD9CM | V10.3 | Personal history of malignant neoplasm of breast |
| Breast cancer | 174 | ICD10CM | Z85.3 | Personal history of malignant neoplasm of breast |
| Breast cancer | 174 | ICD10CM | C50.61 | Malignant neoplasm of axillary tail of breast, female |
| Breast cancer | 174 | ICD10CM | C50.022 | Malignant neoplasm of nipple and areola, left male breast |
| Breast cancer | 174 | ICD10CM | C50.121 | Malignant neoplasm of central portion of right male breast |
| Breast cancer | 174 | ICD10CM | C50.82 | Malignant neoplasm of overlapping sites of breast, male |
| Breast cancer | 174 | ICD10CM | C50.42 | Malignant neoplasm of upper-outer quadrant of breast, male |
| Breast cancer | 174 | ICD10CM | C50.522 | Malignant neoplasm of lower-outer quadrant of left male breast |
| Breast cancer | 174 | ICD10CM | C50.821 | Malignant neoplasm of overlapping sites of right male breast |

| | | | | |
|---|---|---|---|---|
| Breast cancer | 174 | ICD10CM | C50.029 | Malignant neoplasm of nipple and areola, unspecified male breast |
| Breast cancer | 174 | ICD10CM | C50.629 | Malignant neoplasm of axillary tail of unspecified male breast |
| Breast cancer | 174 | ICD10CM | C50.221 | Malignant neoplasm of upper-inner quadrant of right male breast |
| Breast cancer | 174 | ICD10CM | C50.929 | Malignant neoplasm of unspecified site of unspecified male breast |
| Breast cancer | 174 | ICD10CM | C50.229 | Malignant neoplasm of upper-inner quadrant of unspecified male breast |
| Breast cancer | 174 | ICD10CM | C50.321 | Malignant neoplasm of lower-inner quadrant of right male breast |
| Breast cancer | 174 | ICD10CM | C50.912 | Malignant neoplasm of unspecified site of left female breast |
| Breast cancer | 174 | ICD9CM | 174.5 | Malignant neoplasm of lower-outer quadrant of female breast |
| Breast cancer | 174 | ICD9CM | 174.1 | Malignant neoplasm of central portion of female breast |
| Breast cancer | 174 | ICD10CM | C50 | Malignant neoplasm of breast |
| Breast cancer | 174 | ICD10CM | C50.11 | Malignant neoplasm of central portion of breast, female |
| Breast cancer | 174 | ICD10CM | C50.819 | Malignant neoplasm of overlapping sites of unspecified female breast |
| Breast cancer | 174 | ICD10CM | C50.01 | Malignant neoplasm of nipple and areola, female |
| Breast cancer | 174 | ICD9CM | 174.3 | Malignant neoplasm of lower-inner quadrant of female breast |
| Breast cancer | 174 | ICD10CM | C50.619 | Malignant neoplasm of axillary tail of unspecified female breast |
| Breast cancer | 174 | ICD9CM | 174.9 | Malignant neoplasm of breast (female), unspecified |
| Breast cancer | 174 | ICD10CM | C50.412 | Malignant neoplasm of upper-outer quadrant of left female breast |
| Breast cancer | 174 | ICD10CM | C50.111 | Malignant neoplasm of central portion of right female breast |
| Breast cancer | 174 | ICD10CM | C50.312 | Malignant neoplasm of lower-inner quadrant of left female breast |
| Breast cancer | 174 | ICD10CM | C50.219 | Malignant neoplasm of upper-inner quadrant of unspecified female breast |
| Breast cancer | 174 | ICD9CM | 174.8 | Malignant neoplasm of other specified sites of female breast |
| Breast cancer | 174 | ICD10CM | C50.119 | Malignant neoplasm of central portion of unspecified female breast |
| Breast cancer | 174 | ICD9CM | 238.3 | Neoplasm of uncertain behavior of breast |
| Breast cancer | 174 | ICD10CM | D48.60 | Neoplasm of uncertain behavior of unspecified breast |
| Breast cancer | 174 | ICD10CM | C50.422 | Malignant neoplasm of upper-outer quadrant of left male breast |
| Breast cancer | 174 | ICD10CM | C50.329 | Malignant neoplasm of lower-inner quadrant of unspecified male breast |

| | | | | |
|---|---|---|---|---|
| Breast cancer | 174 | ICD10CM | C50.421 | Malignant neoplasm of upper-outer quadrant of right male breast |
| Breast cancer | 174 | ICD9CM | 239.3 | Neoplasm of unspecified nature of breast |
| Breast cancer | 174 | ICD10CM | D49.3 | Neoplasm of unspecified behavior of breast |
| Breast cancer | 174 | ICD10CM | D48.61 | Neoplasm of uncertain behavior of right breast |
| Breast cancer | 174 | ICD10CM | D48.62 | Neoplasm of uncertain behavior of left breast |
| Breast cancer | 174 | ICD10CM | D48.6 | Neoplasm of uncertain behavior of breast |
| Breast cancer | 174 | ICD10CM | C50.129 | Malignant neoplasm of central portion of unspecified male breast |
| Breast cancer | 174 | ICD10CM | C50.02 | Malignant neoplasm of nipple and areola, male |
| Breast cancer | 174 | ICD10CM | C50.921 | Malignant neoplasm of unspecified site of right male breast |
| Breast cancer | 174 | ICD10CM | C50.529 | Malignant neoplasm of lower-outer quadrant of unspecified male breast |
| Breast cancer | 174 | ICD9CM | 175 | Malignant neoplasm of nipple and areola of male breast |
| Breast cancer | 174 | ICD10CM | C50.429 | Malignant neoplasm of upper-outer quadrant of unspecified male breast |
| Breast cancer | 174 | ICD10CM | C50.621 | Malignant neoplasm of axillary tail of right male breast |
| Breast cancer | 174 | ICD10CM | C50.222 | Malignant neoplasm of upper-inner quadrant of left male breast |
| Breast cancer | 174 | ICD10CM | C50.322 | Malignant neoplasm of lower-inner quadrant of left male breast |
| Breast cancer | 174 | ICD10CM | C50.922 | Malignant neoplasm of unspecified site of left male breast |
| Breast cancer | 174 | ICD10CM | C50.122 | Malignant neoplasm of central portion of left male breast |
| Cervical cancer and dysplasia | 180 | ICD9CM | 180 | Malignant neoplasm of cervix uteri |
| Cervical cancer and dysplasia | 180 | ICD9CM | 180.8 | Malignant neoplasm of other specified sites of cervix |
| Cervical cancer and dysplasia | 180 | ICD10CM | C53.0 | Malignant neoplasm of endocervix |
| Cervical cancer and dysplasia | 180 | ICD9CM | 180.9 | Malignant neoplasm of cervix uteri, unspecified site |
| Cervical cancer and dysplasia | 180 | ICD10CM | C53.8 | Malignant neoplasm of overlapping sites of cervix uteri |
| Cervical cancer and dysplasia | 180 | ICD10CM | C53 | Malignant neoplasm of cervix uteri |
| Cervical cancer and dysplasia | 180 | ICD9CM | 180 | Malignant neoplasm of endocervix |
| Cervical cancer and dysplasia | 180 | ICD10CM | C53.9 | Malignant neoplasm of cervix uteri, unspecified |
| Cervical cancer and dysplasia | 180 | ICD9CM | V10.41 | Personal history of malignant neoplasm of cervix uteri |
| Cervical cancer and dysplasia | 180 | ICD10CM | C53.1 | Malignant neoplasm of exocervix |

| | | | | |
|---|---|---|---|---|
| Cervical cancer and dysplasia | 180 | ICD9CM | 180.1 | Malignant neoplasm of exocervix |
| Cervical cancer and dysplasia | 180 | ICD10CM | N87.1 | Moderate cervical dysplasia |
| Cervical cancer and dysplasia | 180 | ICD9CM | 622.1 | Dysplasia of cervix (uteri) |
| Cervical cancer and dysplasia | 180 | ICD10CM | D06.9 | Carcinoma in situ of cervix, unspecified |
| Cervical cancer and dysplasia | 180 | ICD10CM | D06 | Carcinoma in situ of cervix uteri |
| Cervical cancer and dysplasia | 180 | ICD10CM | N87.0 | Mild cervical dysplasia |
| Cervical cancer and dysplasia | 180 | ICD10CM | D06.1 | Carcinoma in situ of exocervix |
| Cervical cancer and dysplasia | 180 | ICD9CM | 622.11 | Mild dysplasia of cervix |
| Cervical cancer and dysplasia | 180 | ICD10CM | D06.7 | Carcinoma in situ of other parts of cervix |
| Cervical cancer and dysplasia | 180 | ICD10CM | N87 | Dysplasia of cervix uteri |
| Cervical cancer and dysplasia | 180 | ICD9CM | 233.1 | Carcinoma in situ of cervix uteri |
| Cervical cancer and dysplasia | 180 | ICD9CM | 622.12 | Moderate dysplasia of cervix |
| Cervical cancer and dysplasia | 180 | ICD10CM | N87.9 | Dysplasia of cervix uteri, unspecified |
| Cervical cancer and dysplasia | 180 | ICD9CM | 622.1 | Dysplasia of cervix, unspecified |
| Cervical cancer and dysplasia | 180 | ICD10CM | D06.0 | Carcinoma in situ of endocervix |
| Cervical cancer | 180.1 | ICD10CM | C53.0 | Malignant neoplasm of endocervix |
| Cervical cancer | 180.1 | ICD9CM | 180.9 | Malignant neoplasm of cervix uteri, unspecified site |
| Cervical cancer | 180.1 | ICD10CM | C53.8 | Malignant neoplasm of overlapping sites of cervix uteri |
| Cervical cancer | 180.1 | ICD9CM | 180.8 | Malignant neoplasm of other specified sites of cervix |
| Cervical cancer | 180.1 | ICD10CM | C53 | Malignant neoplasm of cervix uteri |
| Cervical cancer | 180.1 | ICD9CM | 180 | Malignant neoplasm of endocervix |
| Cervical cancer | 180.1 | ICD10CM | C53.9 | Malignant neoplasm of cervix uteri, unspecified |
| Cervical cancer | 180.1 | ICD9CM | V10.41 | Personal history of malignant neoplasm of cervix uteri |
| Cervical cancer | 180.1 | ICD10CM | C53.1 | Malignant neoplasm of exocervix |
| Cervical cancer | 180.1 | ICD9CM | 180.1 | Malignant neoplasm of exocervix |
| Malignant neoplasm of ovary | 184.11 | ICD10CM | C56 | Malignant neoplasm of ovary |
| Malignant neoplasm of ovary | 184.11 | ICD10CM | C56.3 | Malignant neoplasm of bilateral ovaries |

| | | | | | |
|---|---|---|---|---|---|
| | Malignant neoplasm of ovary | 184.11 | ICD10CM | C56.2 | Malignant neoplasm of left ovary |
| | Malignant neoplasm of ovary | 184.11 | ICD9CM | V10.43 | Personal history of malignant neoplasm of ovary |
| | Malignant neoplasm of ovary | 184.11 | ICD10CM | C56.1 | Malignant neoplasm of right ovary |
| | Malignant neoplasm of ovary | 184.11 | ICD9CM | 183 | Malignant neoplasm of ovary |
| | Malignant neoplasm of ovary | 184.11 | ICD10CM | C56.9 | Malignant neoplasm of unspecified ovary |
| | Malignant neoplasm of ovary | 184.11 | ICD10CM | Z85.43 | Personal history of malignant neoplasm of ovary |
| | Cancer of prostate | 185 | ICD10CM | C61 | Malignant neoplasm of prostate |
| | Cancer of prostate | 185 | ICD9CM | V10.46 | Personal history of malignant neoplasm of prostate |
| | Cancer of prostate | 185 | ICD10CM | Z85.46 | Personal history of malignant neoplasm of prostate |
| | Cancer of prostate | 185 | ICD10CM | D07.5 | Carcinoma in situ of prostate |
| | Cancer of prostate | 185 | ICD9CM | 185 | Malignant neoplasm of prostate |
| | Cancer of prostate | 185 | ICD9CM | 233.4 | Carcinoma in situ of prostate |
| **Metabolic** | Type 2 diabetes | 250.2 | ICD10CM | E11.331 | Type 2 diabetes mellitus with moderate nonproliferative diabetic retinopathy with macular edema |
| | Type 2 diabetes | 250.2 | ICD10CM | E13.319 | Other specified diabetes mellitus with unspecified diabetic retinopathy without macular edema |
| | Type 2 diabetes | 250.2 | ICD10CM | E13.3521 | Other specified diabetes mellitus with proliferative diabetic retinopathy with traction retinal detachment involving the macula, right eye |
| | Type 2 diabetes | 250.2 | ICD10CM | E13.3492 | Other specified diabetes mellitus with severe nonproliferative diabetic retinopathy without macular edema, left eye |
| | Type 2 diabetes | 250.2 | ICD10CM | E11.3219 | Type 2 diabetes mellitus with mild nonproliferative diabetic retinopathy with macular edema, unspecified eye |
| | Type 2 diabetes | 250.2 | ICD10CM | E11.3591 | Type 2 diabetes mellitus with proliferative diabetic retinopathy without macular edema, right eye |
| | Type 2 diabetes | 250.2 | ICD10CM | E11.3553 | Type 2 diabetes mellitus with stable proliferative diabetic retinopathy, bilateral |
| | Type 2 diabetes | 250.2 | ICD10CM | E13.3319 | Other specified diabetes mellitus with moderate nonproliferative diabetic retinopathy with macular edema, unspecified eye |
| | Type 2 diabetes | 250.2 | ICD10CM | E13.628 | Other specified diabetes mellitus with other skin complications |
| | Type 2 diabetes | 250.2 | ICD10CM | E11.39 | Type 2 diabetes mellitus with other diabetic ophthalmic complication |
| | Type 2 diabetes | 250.2 | ICD10CM | E13.9 | Other specified diabetes mellitus without complications |

| | | | | |
|---|---|---|---|---|
| Type 2 diabetes | 250.2 | ICD9CM | 250.8 | Diabetes with other specified manifestations, type II or unspecified type, not stated as uncontrolled |
| Type 2 diabetes | 250.2 | ICD10CM | E13.353 | Other specified diabetes mellitus with proliferative diabetic retinopathy with traction retinal detachment not involving the macula |
| Type 2 diabetes | 250.2 | ICD10CM | E11.01 | Type 2 diabetes mellitus with hyperosmolarity with coma |
| Type 2 diabetes | 250.2 | ICD10CM | E11.3523 | Type 2 diabetes mellitus with proliferative diabetic retinopathy with traction retinal detachment involving the macula, bilateral |
| Type 2 diabetes | 250.2 | ICD10CM | E11.352 | Type 2 diabetes mellitus with proliferative diabetic retinopathy with traction retinal detachment involving the macula |
| Type 2 diabetes | 250.2 | ICD10CM | E11.3419 | Type 2 diabetes mellitus with severe nonproliferative diabetic retinopathy with macular edema, unspecified eye |
| Type 2 diabetes | 250.2 | ICD10CM | E11.3599 | Type 2 diabetes mellitus with proliferative diabetic retinopathy without macular edema, unspecified eye |
| Type 2 diabetes | 250.2 | ICD10CM | E11.638 | Type 2 diabetes mellitus with other oral complications |
| Type 2 diabetes | 250.2 | ICD10CM | E11.3539 | Type 2 diabetes mellitus with proliferative diabetic retinopathy with traction retinal detachment not involving the macula, unspecified eye |
| Type 2 diabetes | 250.2 | ICD10CM | E13.3311 | Other specified diabetes mellitus with moderate nonproliferative diabetic retinopathy with macular edema, right eye |
| Type 2 diabetes | 250.2 | ICD9CM | 250.5 | Diabetes with ophthalmic manifestations, type II or unspecified type, not stated as uncontrolled |
| Type 2 diabetes | 250.2 | ICD10CM | E13.37X9 | Other specified diabetes mellitus with diabetic macular edema, resolved following treatment, unspecified eye |
| Type 2 diabetes | 250.2 | ICD10CM | E13.3531 | Other specified diabetes mellitus with proliferative diabetic retinopathy with traction retinal detachment not involving the macula, right eye |
| Type 2 diabetes | 250.2 | ICD9CM | 250 | Diabetes mellitus without mention of complication, type II or unspecified type, not stated as uncontrolled |
| Type 2 diabetes | 250.2 | ICD10CM | E11.37 | Type 2 diabetes mellitus with diabetic macular edema, resolved following treatment |
| Type 2 diabetes | 250.2 | ICD10CM | E11.32 | Type 2 diabetes mellitus with mild nonproliferative diabetic retinopathy |
| Type 2 diabetes | 250.2 | ICD10CM | E13.3542 | Other specified diabetes mellitus with proliferative diabetic retinopathy with combined traction retinal detachment and rhegmatogenous retinal detachment, left eye |

| | | | | |
|---|---|---|---|---|
| Type 2 diabetes | 250.2 | ICD10CM | E11.3541 | Type 2 diabetes mellitus with proliferative diabetic retinopathy with combined traction retinal detachment and rhegmatogenous retinal detachment, right eye |
| Type 2 diabetes | 250.2 | ICD10CM | E13.3419 | Other specified diabetes mellitus with severe nonproliferative diabetic retinopathy with macular edema, unspecified eye |
| Type 2 diabetes | 250.2 | ICD10CM | E13.3559 | Other specified diabetes mellitus with stable proliferative diabetic retinopathy, unspecified eye |
| Type 2 diabetes | 250.2 | ICD10CM | E11.3552 | Type 2 diabetes mellitus with stable proliferative diabetic retinopathy, left eye |
| Type 2 diabetes | 250.2 | ICD10CM | E11.33 | Type 2 diabetes mellitus with moderate nonproliferative diabetic retinopathy |
| Type 2 diabetes | 250.2 | ICD10CM | E13.3532 | Other specified diabetes mellitus with proliferative diabetic retinopathy with traction retinal detachment not involving the macula, left eye |
| Type 2 diabetes | 250.2 | ICD10CM | E11.1 | Type 2 diabetes mellitus with ketoacidosis |
| Type 2 diabetes | 250.2 | ICD10CM | E11.3551 | Type 2 diabetes mellitus with stable proliferative diabetic retinopathy, right eye |
| Type 2 diabetes | 250.2 | ICD10CM | E13.11 | Other specified diabetes mellitus with ketoacidosis with coma |
| Type 2 diabetes | 250.2 | ICD10CM | E13.10 | Other specified diabetes mellitus with ketoacidosis without coma |
| Type 2 diabetes | 250.2 | ICD10CM | E13.359 | Other specified diabetes mellitus with proliferative diabetic retinopathy without macular edema |
| Type 2 diabetes | 250.2 | ICD10CM | E13.311 | Other specified diabetes mellitus with unspecified diabetic retinopathy with macular edema |
| Type 2 diabetes | 250.2 | ICD10CM | E13.3541 | Other specified diabetes mellitus with proliferative diabetic retinopathy with combined traction retinal detachment and rhegmatogenous retinal detachment, right eye |
| Type 2 diabetes | 250.2 | ICD10CM | E13.3543 | Other specified diabetes mellitus with proliferative diabetic retinopathy with combined traction retinal detachment and rhegmatogenous retinal detachment, bilateral |
| Type 2 diabetes | 250.2 | ICD10CM | E11.3492 | Type 2 diabetes mellitus with severe nonproliferative diabetic retinopathy without macular edema, left eye |
| Type 2 diabetes | 250.2 | ICD10CM | E11.11 | Type 2 diabetes mellitus with ketoacidosis with coma |
| Type 2 diabetes | 250.2 | ICD10CM | E11.3529 | Type 2 diabetes mellitus with proliferative diabetic retinopathy with traction retinal detachment involving the macula, unspecified eye |
| Type 2 diabetes | 250.2 | ICD9CM | 250.1 | Diabetes with ketoacidosis, type II or unspecified type, not stated as uncontrolled |

| | | | | |
|---|---|---|---|---|
| Type 2 diabetes | 250.2 | ICD10CM | E13.3312 | Other specified diabetes mellitus with moderate nonproliferative diabetic retinopathy with macular edema, left eye |
| Type 2 diabetes | 250.2 | ICD10CM | E13.1 | Other specified diabetes mellitus with ketoacidosis |
| Type 2 diabetes | 250.2 | ICD10CM | E13.321 | Other specified diabetes mellitus with mild nonproliferative diabetic retinopathy with macular edema |
| Type 2 diabetes | 250.2 | ICD10CM | E13.33 | Other specified diabetes mellitus with moderate nonproliferative diabetic retinopathy |
| Type 2 diabetes | 250.2 | ICD10CM | E13.32 | Other specified diabetes mellitus with mild nonproliferative diabetic retinopathy |
| Type 2 diabetes | 250.2 | ICD10CM | E13.8 | Other specified diabetes mellitus with unspecified complications |
| Type 2 diabetes | 250.2 | ICD10CM | E13.630 | Other specified diabetes mellitus with periodontal disease |
| Type 2 diabetes | 250.2 | ICD10CM | E13.34 | Other specified diabetes mellitus with severe nonproliferative diabetic retinopathy |
| Type 2 diabetes | 250.2 | ICD10CM | E11.311 | Type 2 diabetes mellitus with unspecified diabetic retinopathy with macular edema |
| Type 2 diabetes | 250.2 | ICD10CM | E13.3511 | Other specified diabetes mellitus with proliferative diabetic retinopathy with macular edema, right eye |
| Type 2 diabetes | 250.2 | ICD10CM | E13.3391 | Other specified diabetes mellitus with moderate nonproliferative diabetic retinopathy without macular edema, right eye |
| Type 2 diabetes | 250.2 | ICD10CM | E11.10 | Type 2 diabetes mellitus with ketoacidosis without coma |
| Type 2 diabetes | 250.2 | ICD10CM | E13.3533 | Other specified diabetes mellitus with proliferative diabetic retinopathy with traction retinal detachment not involving the macula, bilateral |
| Type 2 diabetes | 250.2 | ICD10CM | E11.37X1 | Type 2 diabetes mellitus with diabetic macular edema, resolved following treatment, right eye |
| Type 2 diabetes | 250.2 | ICD10CM | E11.37X9 | Type 2 diabetes mellitus with diabetic macular edema, resolved following treatment, unspecified eye |
| Type 2 diabetes | 250.2 | ICD10CM | E11.359 | Type 2 diabetes mellitus with proliferative diabetic retinopathy without macular edema |
| Type 2 diabetes | 250.2 | ICD10CM | E11.37X2 | Type 2 diabetes mellitus with diabetic macular edema, resolved following treatment, left eye |
| Type 2 diabetes | 250.2 | ICD10CM | E11.3211 | Type 2 diabetes mellitus with mild nonproliferative diabetic retinopathy with macular edema, right eye |
| Type 2 diabetes | 250.2 | ICD10CM | E11.319 | Type 2 diabetes mellitus with unspecified diabetic retinopathy without macular edema |
| Type 2 diabetes | 250.2 | ICD10CM | E11.3521 | Type 2 diabetes mellitus with proliferative diabetic retinopathy with traction retinal detachment involving the macula, right eye |

| | | | | |
|---|---|---|---|---|
| Type 2 diabetes | 250.2 | ICD10CM | E13.3549 | Other specified diabetes mellitus with proliferative diabetic retinopathy with combined traction retinal detachment and rhegmatogenous retinal detachment, unspecified eye |
| Type 2 diabetes | 250.2 | ICD10CM | E13.3 | Other specified diabetes mellitus with ophthalmic complications |
| Type 2 diabetes | 250.2 | ICD10CM | E11.41 | Type 2 diabetes mellitus with diabetic mononeuropathy |
| Type 2 diabetes | 250.2 | ICD10CM | E13.351 | Other specified diabetes mellitus with proliferative diabetic retinopathy with macular edema |
| Type 2 diabetes | 250.2 | ICD10CM | E11.63 | Type 2 diabetes mellitus with oral complications |
| Type 2 diabetes | 250.2 | ICD10CM | E11.40 | Type 2 diabetes mellitus with diabetic neuropathy, unspecified |
| Type 2 diabetes | 250.2 | ICD10CM | E11.42 | Type 2 diabetes mellitus with diabetic polyneuropathy |
| Type 2 diabetes | 250.2 | ICD9CM | 250.6 | Diabetes with neurological manifestations, type II or unspecified type, not stated as uncontrolled |
| Type 2 diabetes | 250.2 | ICD10CM | E13.43 | Other specified diabetes mellitus with diabetic autonomic (poly)neuropathy |
| Type 2 diabetes | 250.2 | ICD10CM | E11.44 | Type 2 diabetes mellitus with diabetic amyotrophy |
| Type 2 diabetes | 250.2 | ICD9CM | 250.12 | Diabetes with ketoacidosis, type II or unspecified type, uncontrolled |
| Type 2 diabetes | 250.2 | ICD10CM | E11.65 | Type 2 diabetes mellitus with hyperglycemia |
| Type 2 diabetes | 250.2 | ICD10CM | E11.628 | Type 2 diabetes mellitus with other skin complications |
| Type 2 diabetes | 250.2 | ICD10CM | E13.3392 | Other specified diabetes mellitus with moderate nonproliferative diabetic retinopathy without macular edema, left eye |
| Type 2 diabetes | 250.2 | ICD10CM | E13.3519 | Other specified diabetes mellitus with proliferative diabetic retinopathy with macular edema, unspecified eye |
| Type 2 diabetes | 250.2 | ICD10CM | E13.40 | Other specified diabetes mellitus with diabetic neuropathy, unspecified |
| Type 2 diabetes | 250.2 | ICD10CM | E13.44 | Other specified diabetes mellitus with diabetic amyotrophy |
| Type 2 diabetes | 250.2 | ICD10CM | E11.3513 | Type 2 diabetes mellitus with proliferative diabetic retinopathy with macular edema, bilateral |
| Type 2 diabetes | 250.2 | ICD10CM | E13.01 | Other specified diabetes mellitus with hyperosmolarity with coma |
| Type 2 diabetes | 250.2 | ICD10CM | E13.621 | Other specified diabetes mellitus with foot ulcer |
| Type 2 diabetes | 250.2 | ICD10CM | E13.638 | Other specified diabetes mellitus with other oral complications |
| Type 2 diabetes | 250.2 | ICD10CM | E13.620 | Other specified diabetes mellitus with diabetic dermatitis |
| Type 2 diabetes | 250.2 | ICD10CM | E11.621 | Type 2 diabetes mellitus with foot ulcer |

| | | | | |
|---|---|---|---|---|
| Type 2 diabetes | 250.2 | ICD10CM | E13.3212 | Other specified diabetes mellitus with mild nonproliferative diabetic retinopathy with macular edema, left eye |
| Type 2 diabetes | 250.2 | ICD10CM | E13.3299 | Other specified diabetes mellitus with mild nonproliferative diabetic retinopathy without macular edema, unspecified eye |
| Type 2 diabetes | 250.2 | ICD10CM | E11.3313 | Type 2 diabetes mellitus with moderate nonproliferative diabetic retinopathy with macular edema, bilateral |
| Type 2 diabetes | 250.2 | ICD10CM | E11.3412 | Type 2 diabetes mellitus with severe nonproliferative diabetic retinopathy with macular edema, left eye |
| Type 2 diabetes | 250.2 | ICD10CM | E11.61 | Type 2 diabetes mellitus with diabetic arthropathy |
| Type 2 diabetes | 250.2 | ICD10CM | E11.3533 | Type 2 diabetes mellitus with proliferative diabetic retinopathy with traction retinal detachment not involving the macula, bilateral |
| Type 2 diabetes | 250.2 | ICD10CM | E13.3529 | Other specified diabetes mellitus with proliferative diabetic retinopathy with traction retinal detachment involving the macula, unspecified eye |
| Type 2 diabetes | 250.2 | ICD9CM | 250.92 | Diabetes with unspecified complication, type II or unspecified type, uncontrolled |
| Type 2 diabetes | 250.2 | ICD10CM | E11.8 | Type 2 diabetes mellitus with unspecified complications |
| Type 2 diabetes | 250.2 | ICD10CM | E13.641 | Other specified diabetes mellitus with hypoglycemia with coma |
| Type 2 diabetes | 250.2 | ICD10CM | E11.610 | Type 2 diabetes mellitus with diabetic neuropathic arthropathy |
| Type 2 diabetes | 250.2 | ICD10CM | E13.3539 | Other specified diabetes mellitus with proliferative diabetic retinopathy with traction retinal detachment not involving the macula, unspecified eye |
| Type 2 diabetes | 250.2 | ICD10CM | E11.351 | Type 2 diabetes mellitus with proliferative diabetic retinopathy with macular edema |
| Type 2 diabetes | 250.2 | ICD10CM | E11.3292 | Type 2 diabetes mellitus with mild nonproliferative diabetic retinopathy without macular edema, left eye |
| Type 2 diabetes | 250.2 | ICD10CM | E11.3519 | Type 2 diabetes mellitus with proliferative diabetic retinopathy with macular edema, unspecified eye |
| Type 2 diabetes | 250.2 | ICD10CM | E11.36 | Type 2 diabetes mellitus with diabetic cataract |
| Type 2 diabetes | 250.2 | ICD10CM | E11.321 | Type 2 diabetes mellitus with mild nonproliferative diabetic retinopathy with macular edema |
| Type 2 diabetes | 250.2 | ICD9CM | 250.62 | Diabetes with neurological manifestations, type II or unspecified type, uncontrolled |
| Type 2 diabetes | 250.2 | ICD10CM | E13.649 | Other specified diabetes mellitus with hypoglycemia without coma |
| Type 2 diabetes | 250.2 | ICD10CM | E11.9 | Type 2 diabetes mellitus without complications |

| | | | | |
|---|---|---|---|---|
| Type 2 diabetes | 250.2 | ICD10CM | E13.622 | Other specified diabetes mellitus with other skin ulcer |
| Type 2 diabetes | 250.2 | ICD10CM | E13.00 | Other specified diabetes mellitus with hyperosmolarity without nonketotic hyperglycemic-hyperosmolar coma (NKHHC) |
| Type 2 diabetes | 250.2 | ICD10CM | E11.622 | Type 2 diabetes mellitus with other skin ulcer |
| Type 2 diabetes | 250.2 | ICD10CM | E13.0 | Other specified diabetes mellitus with hyperosmolarity |
| Type 2 diabetes | 250.2 | ICD10CM | E11.69 | Type 2 diabetes mellitus with other specified complication |
| Type 2 diabetes | 250.2 | ICD10CM | E11.630 | Type 2 diabetes mellitus with periodontal disease |
| Type 2 diabetes | 250.2 | ICD10CM | E11.3559 | Type 2 diabetes mellitus with stable proliferative diabetic retinopathy, unspecified eye |
| Type 2 diabetes | 250.2 | ICD10CM | E13.610 | Other specified diabetes mellitus with diabetic neuropathic arthropathy |
| Type 2 diabetes | 250.2 | ICD10CM | E13.42 | Other specified diabetes mellitus with diabetic polyneuropathy |
| Type 2 diabetes | 250.2 | ICD9CM | 250.02 | Diabetes mellitus without mention of complication, type II or unspecified type, uncontrolled |
| Type 2 diabetes | 250.2 | ICD9CM | 250.32 | Diabetes with other coma, type II or unspecified type, uncontrolled |
| Type 2 diabetes | 250.2 | ICD10CM | E11.37X3 | Type 2 diabetes mellitus with diabetic macular edema, resolved following treatment, bilateral |
| Type 2 diabetes | 250.2 | ICD9CM | 250.82 | Diabetes with other specified manifestations, type II or unspecified type, uncontrolled |
| Type 2 diabetes | 250.2 | ICD10CM | E11.0 | Type 2 diabetes mellitus with hyperosmolarity |
| Type 2 diabetes | 250.2 | ICD10CM | E11.649 | Type 2 diabetes mellitus with hypoglycemia without coma |
| Type 2 diabetes | 250.2 | ICD9CM | 250.2 | Diabetes with hyperosmolarity, type II or unspecified type, not stated as uncontrolled |
| Type 2 diabetes | 250.2 | ICD9CM | 250.9 | Diabetes with unspecified complication, type II or unspecified type, not stated as uncontrolled |
| Type 2 diabetes | 250.2 | ICD9CM | 250.22 | Diabetes with hyperosmolarity, type II or unspecified type, uncontrolled |
| Type 2 diabetes | 250.2 | ICD9CM | 250.3 | Diabetes with other coma, type II or unspecified type, not stated as uncontrolled |
| Type 2 diabetes | 250.2 | ICD10CM | E11.641 | Type 2 diabetes mellitus with hypoglycemia with coma |
| Type 2 diabetes | 250.2 | ICD10CM | E11.64 | Type 2 diabetes mellitus with hypoglycemia |
| Type 2 diabetes | 250.2 | ICD10CM | E11.620 | Type 2 diabetes mellitus with diabetic dermatitis |
| Type 2 diabetes | 250.2 | ICD10CM | E11.6 | Type 2 diabetes mellitus with other specified complications |
| Type 2 diabetes | 250.2 | ICD10CM | E13.3513 | Other specified diabetes mellitus with proliferative diabetic retinopathy with macular edema, bilateral |

| | | | | |
|---|---|---|---|---|
| Type 2 diabetes | 250.2 | ICD10CM | E13.37X2 | Other specified diabetes mellitus with diabetic macular edema, resolved following treatment, left eye |
| Type 2 diabetes | 250.2 | ICD10CM | E11.35 | Type 2 diabetes mellitus with proliferative diabetic retinopathy |
| Type 2 diabetes | 250.2 | ICD10CM | E11.3413 | Type 2 diabetes mellitus with severe nonproliferative diabetic retinopathy with macular edema, bilateral |
| Type 2 diabetes | 250.2 | ICD10CM | E13.3551 | Other specified diabetes mellitus with stable proliferative diabetic retinopathy, right eye |
| Type 2 diabetes | 250.2 | ICD10CM | E13.29 | Other specified diabetes mellitus with other diabetic kidney complication |
| Type 2 diabetes | 250.2 | ICD10CM | E13.21 | Other specified diabetes mellitus with diabetic nephropathy |
| Type 2 diabetes | 250.2 | ICD9CM | 250.42 | Diabetes with renal manifestations, type II or unspecified type, uncontrolled |
| Type 2 diabetes | 250.2 | ICD10CM | E11.21 | Type 2 diabetes mellitus with diabetic nephropathy |
| Type 2 diabetes | 250.2 | ICD10CM | E11.618 | Type 2 diabetes mellitus with other diabetic arthropathy |
| Type 2 diabetes | 250.2 | ICD10CM | E11 | Type 2 diabetes mellitus |
| Type 2 diabetes | 250.2 | ICD10CM | E13.3512 | Other specified diabetes mellitus with proliferative diabetic retinopathy with macular edema, left eye |
| Type 2 diabetes | 250.2 | ICD10CM | E11.3291 | Type 2 diabetes mellitus with mild nonproliferative diabetic retinopathy without macular edema, right eye |
| Type 2 diabetes | 250.2 | ICD10CM | E13.3213 | Other specified diabetes mellitus with mild nonproliferative diabetic retinopathy with macular edema, bilateral |
| Type 2 diabetes | 250.2 | ICD10CM | E13.51 | Other specified diabetes mellitus with diabetic peripheral angiopathy without gangrene |
| Type 2 diabetes | 250.2 | ICD10CM | E13.5 | Other specified diabetes mellitus with circulatory complications |
| Type 2 diabetes | 250.2 | ICD10CM | E13.3292 | Other specified diabetes mellitus with mild nonproliferative diabetic retinopathy without macular edema, left eye |
| Type 2 diabetes | 250.2 | ICD10CM | E11.3512 | Type 2 diabetes mellitus with proliferative diabetic retinopathy with macular edema, left eye |
| Type 2 diabetes | 250.2 | ICD10CM | E11.3212 | Type 2 diabetes mellitus with mild nonproliferative diabetic retinopathy with macular edema, left eye |
| Type 2 diabetes | 250.2 | ICD10CM | E13.41 | Other specified diabetes mellitus with diabetic mononeuropathy |
| Type 2 diabetes | 250.2 | ICD10CM | E11.52 | Type 2 diabetes mellitus with diabetic peripheral angiopathy with gangrene |
| Type 2 diabetes | 250.2 | ICD9CM | 250.7 | Diabetes with peripheral circulatory disorders, type II or unspecified type, not stated as uncontrolled |

| | | | | |
|---|---|---|---|---|
| Type 2 diabetes | 250.2 | ICD10CM | E13.4 | Other specified diabetes mellitus with neurological complications |
| Type 2 diabetes | 250.2 | ICD10CM | E13.49 | Other specified diabetes mellitus with other diabetic neurological complication |
| Type 2 diabetes | 250.2 | ICD10CM | E11.4 | Type 2 diabetes mellitus with neurological complications |
| Type 2 diabetes | 250.2 | ICD10CM | E11.49 | Type 2 diabetes mellitus with other diabetic neurological complication |
| Type 2 diabetes | 250.2 | ICD10CM | E11.43 | Type 2 diabetes mellitus with diabetic autonomic (poly)neuropathy |
| Type 2 diabetes | 250.2 | ICD10CM | E13.22 | Other specified diabetes mellitus with diabetic chronic kidney disease |
| Type 2 diabetes | 250.2 | ICD10CM | E13.36 | Other specified diabetes mellitus with diabetic cataract |
| Type 2 diabetes | 250.2 | ICD10CM | E13.39 | Other specified diabetes mellitus with other diabetic ophthalmic complication |
| Type 2 diabetes | 250.2 | ICD10CM | E11.353 | Type 2 diabetes mellitus with proliferative diabetic retinopathy with traction retinal detachment not involving the macula |
| Type 2 diabetes | 250.2 | ICD10CM | E13.3293 | Other specified diabetes mellitus with mild nonproliferative diabetic retinopathy without macular edema, bilateral |
| Type 2 diabetes | 250.2 | ICD10CM | E11.3392 | Type 2 diabetes mellitus with moderate nonproliferative diabetic retinopathy without macular edema, left eye |
| Type 2 diabetes | 250.2 | ICD10CM | E13.329 | Other specified diabetes mellitus with mild nonproliferative diabetic retinopathy without macular edema |
| Type 2 diabetes | 250.2 | ICD10CM | E11.3391 | Type 2 diabetes mellitus with moderate nonproliferative diabetic retinopathy without macular edema, right eye |
| Type 2 diabetes | 250.2 | ICD10CM | E13.3393 | Other specified diabetes mellitus with moderate nonproliferative diabetic retinopathy without macular edema, bilateral |
| Type 2 diabetes | 250.2 | ICD10CM | E11.3592 | Type 2 diabetes mellitus with proliferative diabetic retinopathy without macular edema, left eye |
| Type 2 diabetes | 250.2 | ICD10CM | E13.69 | Other specified diabetes mellitus with other specified complication |
| Type 2 diabetes | 250.2 | ICD10CM | E13.65 | Other specified diabetes mellitus with hyperglycemia |
| Type 2 diabetes | 250.2 | ICD10CM | E11.2 | Type 2 diabetes mellitus with kidney complications |
| Type 2 diabetes | 250.2 | ICD10CM | E11.31 | Type 2 diabetes mellitus with unspecified diabetic retinopathy |
| Type 2 diabetes | 250.2 | ICD10CM | E11.22 | Type 2 diabetes mellitus with diabetic chronic kidney disease |
| Type 2 diabetes | 250.2 | ICD10CM | E11.29 | Type 2 diabetes mellitus with other diabetic kidney complication |
| Type 2 diabetes | 250.2 | ICD9CM | 250.4 | Diabetes with renal manifestations, type II or unspecified type, not stated as uncontrolled |

| | | | | |
|---|---|---|---|---|
| Type 2 diabetes | 250.2 | ICD10CM | E13.2 | Other specified diabetes mellitus with kidney complications |
| Type 2 diabetes | 250.2 | ICD10CM | O24.11 | Pre-existing type 2 diabetes mellitus, in pregnancy |
| Type 2 diabetes | 250.2 | ICD10CM | E13.618 | Other specified diabetes mellitus with other diabetic arthropathy |
| Type 2 diabetes | 250.2 | ICD10CM | E13 | Other specified diabetes mellitus |
| Type 2 diabetes | 250.2 | ICD10CM | E11.62 | Type 2 diabetes mellitus with skin complications |
| Type 2 diabetes | 250.2 | ICD10CM | E11.341 | Type 2 diabetes mellitus with severe nonproliferative diabetic retinopathy with macular edema |
| Type 2 diabetes | 250.2 | ICD10CM | E11.34 | Type 2 diabetes mellitus with severe nonproliferative diabetic retinopathy |
| Type 2 diabetes | 250.2 | ICD10CM | E13.3523 | Other specified diabetes mellitus with proliferative diabetic retinopathy with traction retinal detachment involving the macula, bilateral |
| Type 2 diabetes | 250.2 | ICD10CM | E13.3413 | Other specified diabetes mellitus with severe nonproliferative diabetic retinopathy with macular edema, bilateral |
| Type 2 diabetes | 250.2 | ICD10CM | E11.3543 | Type 2 diabetes mellitus with proliferative diabetic retinopathy with combined traction retinal detachment and rhegmatogenous retinal detachment, bilateral |
| Type 2 diabetes | 250.2 | ICD10CM | E11.3411 | Type 2 diabetes mellitus with severe nonproliferative diabetic retinopathy with macular edema, right eye |
| Type 2 diabetes | 250.2 | ICD10CM | E13.3552 | Other specified diabetes mellitus with stable proliferative diabetic retinopathy, left eye |
| Type 2 diabetes | 250.2 | ICD10CM | E13.59 | Other specified diabetes mellitus with other circulatory complications |
| Type 2 diabetes | 250.2 | ICD10CM | E13.339 | Other specified diabetes mellitus with moderate nonproliferative diabetic retinopathy without macular edema |
| Type 2 diabetes | 250.2 | ICD10CM | E11.3531 | Type 2 diabetes mellitus with proliferative diabetic retinopathy with traction retinal detachment not involving the macula, right eye |
| Type 2 diabetes | 250.2 | ICD10CM | E11.5 | Type 2 diabetes mellitus with circulatory complications |
| Type 2 diabetes | 250.2 | ICD10CM | E13.3211 | Other specified diabetes mellitus with mild nonproliferative diabetic retinopathy with macular edema, right eye |
| Type 2 diabetes | 250.2 | ICD10CM | E11.51 | Type 2 diabetes mellitus with diabetic peripheral angiopathy without gangrene |
| Type 2 diabetes | 250.2 | ICD10CM | E11.59 | Type 2 diabetes mellitus with other circulatory complications |
| Type 2 diabetes | 250.2 | ICD10CM | E13.354 | Other specified diabetes mellitus with proliferative diabetic retinopathy with combined traction retinal detachment and rhegmatogenous retinal detachment |

| | | | | |
|---|---|---|---|---|
| Type 2 diabetes | 250.2 | ICD10CM | E11.3549 | Type 2 diabetes mellitus with proliferative diabetic retinopathy with combined traction retinal detachment and rhegmatogenous retinal detachment, unspecified eye |
| Type 2 diabetes | 250.2 | ICD10CM | E11.3312 | Type 2 diabetes mellitus with moderate nonproliferative diabetic retinopathy with macular edema, left eye |
| Type 2 diabetes | 250.2 | ICD10CM | E13.31 | Other specified diabetes mellitus with unspecified diabetic retinopathy |
| Type 2 diabetes | 250.2 | ICD10CM | E11.3319 | Type 2 diabetes mellitus with moderate nonproliferative diabetic retinopathy with macular edema, unspecified eye |
| Type 2 diabetes | 250.2 | ICD10CM | E11.3532 | Type 2 diabetes mellitus with proliferative diabetic retinopathy with traction retinal detachment not involving the macula, left eye |
| Type 2 diabetes | 250.2 | ICD10CM | E13.3599 | Other specified diabetes mellitus with proliferative diabetic retinopathy without macular edema, unspecified eye |
| Type 2 diabetes | 250.2 | ICD10CM | E11.3 | Type 2 diabetes mellitus with ophthalmic complications |
| Type 2 diabetes | 250.2 | ICD10CM | E13.37 | Other specified diabetes mellitus with diabetic macular edema, resolved following treatment |
| Type 2 diabetes | 250.2 | ICD10CM | E13.352 | Other specified diabetes mellitus with proliferative diabetic retinopathy with traction retinal detachment involving the macula |
| Type 2 diabetes | 250.2 | ICD10CM | E13.3291 | Other specified diabetes mellitus with mild nonproliferative diabetic retinopathy without macular edema, right eye |
| Type 2 diabetes | 250.2 | ICD10CM | E11.3522 | Type 2 diabetes mellitus with proliferative diabetic retinopathy with traction retinal detachment involving the macula, left eye |
| Type 2 diabetes | 250.2 | ICD10CM | E13.3591 | Other specified diabetes mellitus with proliferative diabetic retinopathy without macular edema, right eye |
| Type 2 diabetes | 250.2 | ICD10CM | E11.3299 | Type 2 diabetes mellitus with mild nonproliferative diabetic retinopathy without macular edema, unspecified eye |
| Type 2 diabetes | 250.2 | ICD10CM | E11.3293 | Type 2 diabetes mellitus with mild nonproliferative diabetic retinopathy without macular edema, bilateral |
| Type 2 diabetes | 250.2 | ICD10CM | E13.3313 | Other specified diabetes mellitus with moderate nonproliferative diabetic retinopathy with macular edema, bilateral |
| Type 2 diabetes | 250.2 | ICD10CM | E11.3213 | Type 2 diabetes mellitus with mild nonproliferative diabetic retinopathy with macular edema, bilateral |
| Type 2 diabetes | 250.2 | ICD10CM | E13.35 | Other specified diabetes mellitus with proliferative diabetic retinopathy |
| Type 2 diabetes | 250.2 | ICD10CM | E13.37X1 | Other specified diabetes mellitus with diabetic macular edema, resolved following treatment, right eye |

| | | | | |
|---|---|---|---|---|
| Type 2 diabetes | 250.2 | ICD10CM | E13.3491 | Other specified diabetes mellitus with severe nonproliferative diabetic retinopathy without macular edema, right eye |
| Type 2 diabetes | 250.2 | ICD10CM | E11.00 | Type 2 diabetes mellitus with hyperosmolarity without nonketotic hyperglycemic-hyperosmolar coma (NKHHC) |
| Type 2 diabetes | 250.2 | ICD9CM | 250.52 | Diabetes with ophthalmic manifestations, type II or unspecified type, uncontrolled |
| Type 2 diabetes | 250.2 | ICD10CM | E13.37X3 | Other specified diabetes mellitus with diabetic macular edema, resolved following treatment, bilateral |
| Type 2 diabetes | 250.2 | ICD10CM | E13.331 | Other specified diabetes mellitus with moderate nonproliferative diabetic retinopathy with macular edema |
| Type 2 diabetes | 250.2 | ICD10CM | E11.3399 | Type 2 diabetes mellitus with moderate nonproliferative diabetic retinopathy without macular edema, unspecified eye |
| Type 2 diabetes | 250.2 | ICD10CM | E11.3542 | Type 2 diabetes mellitus with proliferative diabetic retinopathy with combined traction retinal detachment and rhegmatogenous retinal detachment, left eye |
| Type 2 diabetes | 250.2 | ICD10CM | E11.329 | Type 2 diabetes mellitus with mild nonproliferative diabetic retinopathy without macular edema |
| Type 2 diabetes | 250.2 | ICD10CM | E11.355 | Type 2 diabetes mellitus with stable proliferative diabetic retinopathy |
| Type 2 diabetes | 250.2 | ICD10CM | E13.349 | Other specified diabetes mellitus with severe nonproliferative diabetic retinopathy without macular edema |
| Type 2 diabetes | 250.2 | ICD10CM | E13.3493 | Other specified diabetes mellitus with severe nonproliferative diabetic retinopathy without macular edema, bilateral |
| Type 2 diabetes | 250.2 | ICD10CM | E13.341 | Other specified diabetes mellitus with severe nonproliferative diabetic retinopathy with macular edema |
| Type 2 diabetes | 250.2 | ICD10CM | E13.3593 | Other specified diabetes mellitus with proliferative diabetic retinopathy without macular edema, bilateral |
| Type 2 diabetes | 250.2 | ICD10CM | E11.354 | Type 2 diabetes mellitus with proliferative diabetic retinopathy with combined traction retinal detachment and rhegmatogenous retinal detachment |
| Type 2 diabetes | 250.2 | ICD10CM | E13.52 | Other specified diabetes mellitus with diabetic peripheral angiopathy with gangrene |
| Type 2 diabetes | 250.2 | ICD9CM | 250.72 | Diabetes with peripheral circulatory disorders, type II or unspecified type, uncontrolled |
| Type 2 diabetes | 250.2 | ICD10CM | E11.3393 | Type 2 diabetes mellitus with moderate nonproliferative diabetic retinopathy without macular edema, bilateral |
| Type 2 diabetes | 250.2 | ICD10CM | E13.3499 | Other specified diabetes mellitus with severe nonproliferative diabetic retinopathy without macular edema, unspecified eye |

| | | | | |
|---|---|---|---|---|
| Type 2 diabetes | 250.2 | ICD10CM | E13.3522 | Other specified diabetes mellitus with proliferative diabetic retinopathy with traction retinal detachment involving the macula, left eye |
| Type 2 diabetes | 250.2 | ICD10CM | E13.3219 | Other specified diabetes mellitus with mild nonproliferative diabetic retinopathy with macular edema, unspecified eye |
| Type 2 diabetes | 250.2 | ICD10CM | E13.355 | Other specified diabetes mellitus with stable proliferative diabetic retinopathy |
| Type 2 diabetes | 250.2 | ICD10CM | E13.3399 | Other specified diabetes mellitus with moderate nonproliferative diabetic retinopathy without macular edema, unspecified eye |
| Type 2 diabetes | 250.2 | ICD10CM | E11.3491 | Type 2 diabetes mellitus with severe nonproliferative diabetic retinopathy without macular edema, right eye |
| Type 2 diabetes | 250.2 | ICD10CM | E13.3411 | Other specified diabetes mellitus with severe nonproliferative diabetic retinopathy with macular edema, right eye |
| Type 2 diabetes | 250.2 | ICD10CM | E13.3412 | Other specified diabetes mellitus with severe nonproliferative diabetic retinopathy with macular edema, left eye |
| Type 2 diabetes | 250.2 | ICD10CM | E11.3511 | Type 2 diabetes mellitus with proliferative diabetic retinopathy with macular edema, right eye |
| Type 2 diabetes | 250.2 | ICD10CM | E13.3553 | Other specified diabetes mellitus with stable proliferative diabetic retinopathy, bilateral |
| Type 2 diabetes | 250.2 | ICD10CM | E11.339 | Type 2 diabetes mellitus with moderate nonproliferative diabetic retinopathy without macular edema |
| Type 2 diabetes | 250.2 | ICD10CM | E11.3493 | Type 2 diabetes mellitus with severe nonproliferative diabetic retinopathy without macular edema, bilateral |
| Type 2 diabetes | 250.2 | ICD10CM | E11.3593 | Type 2 diabetes mellitus with proliferative diabetic retinopathy without macular edema, bilateral |
| Type 2 diabetes | 250.2 | ICD10CM | E11.349 | Type 2 diabetes mellitus with severe nonproliferative diabetic retinopathy without macular edema |
| Type 2 diabetes | 250.2 | ICD10CM | E11.3499 | Type 2 diabetes mellitus with severe nonproliferative diabetic retinopathy without macular edema, unspecified eye |
| Type 2 diabetes | 250.2 | ICD10CM | E13.3592 | Other specified diabetes mellitus with proliferative diabetic retinopathy without macular edema, left eye |
| Type 2 diabetes | 250.2 | ICD10CM | E11.3311 | Type 2 diabetes mellitus with moderate nonproliferative diabetic retinopathy with macular edema, right eye |
| Hypercholesterolemia | 272.11 | ICD10CM | E78.01 | Familial hypercholesterolemia |
| Hypercholesterolemia | 272.11 | ICD10CM | E78.0 | Pure hypercholesterolemia |
| Hypercholesterolemia | 272.11 | ICD9CM | 272 | Pure hypercholesterolemia |
| Hypercholesterolemia | 272.11 | ICD10CM | E78.00 | Pure hypercholesterolemia, unspecified |

| | | | | |
|---|---|---|---|---|
| Gout | 274.1 | ICD10CM | M10.42 | Other secondary gout, elbow |
| Gout | 274.1 | ICD9CM | 274.03 | Chronic gouty arthropathy with tophus (tophi) |
| Gout | 274.1 | ICD10CM | M1A.02 | Idiopathic chronic gout, elbow |
| Gout | 274.1 | ICD10CM | M1A.9XX1 | Chronic gout, unspecified, with tophus (tophi) |
| Gout | 274.1 | ICD10CM | M10.472 | Other secondary gout, left ankle and foot |
| Gout | 274.1 | ICD10CM | M10.44 | Other secondary gout, hand |
| Gout | 274.1 | ICD9CM | 274.81 | Gouty tophi of ear |
| Gout | 274.1 | ICD10CM | M10.412 | Other secondary gout, left shoulder |
| Gout | 274.1 | ICD10CM | M10.459 | Other secondary gout, unspecified hip |
| Gout | 274.1 | ICD10CM | M10.9 | Gout, unspecified |
| Gout | 274.1 | ICD10CM | M1A.9 | Chronic gout, unspecified |
| Gout | 274.1 | ICD10CM | M10.439 | Other secondary gout, unspecified wrist |
| Gout | 274.1 | ICD10CM | M10.462 | Other secondary gout, left knee |
| Gout | 274.1 | ICD10CM | M10.449 | Other secondary gout, unspecified hand |
| Gout | 274.1 | ICD9CM | 274 | Gout |
| Gout | 274.1 | ICD10CM | M10.41 | Other secondary gout, shoulder |
| Gout | 274.1 | ICD10CM | M10.48 | Other secondary gout, vertebrae |
| Gout | 274.1 | ICD10CM | M1A.3791 | Chronic gout due to renal impairment, unspecified ankle and foot, with tophus (tophi) |
| Gout | 274.1 | ICD10CM | M1A.239 | Drug-induced chronic gout, unspecified wrist |
| Gout | 274.1 | ICD10CM | M1A.2521 | Drug-induced chronic gout, left hip, with tophus (tophi) |
| Gout | 274.1 | ICD10CM | M1A.40X0 | Other secondary chronic gout, unspecified site, without tophus (tophi) |
| Gout | 274.1 | ICD10CM | M10.37 | Gout due to renal impairment, ankle and foot |
| Gout | 274.1 | ICD10CM | M10.432 | Other secondary gout, left wrist |
| Gout | 274.1 | ICD9CM | 274.1 | Gouty nephropathy |
| Gout | 274.1 | ICD10CM | M10.47 | Other secondary gout, ankle and foot |
| Gout | 274.1 | ICD9CM | 274.9 | Gout, unspecified |
| Gout | 274.1 | ICD10CM | M10.411 | Other secondary gout, right shoulder |
| Gout | 274.1 | ICD10CM | M10.461 | Other secondary gout, right knee |
| Gout | 274.1 | ICD10CM | M10.431 | Other secondary gout, right wrist |
| Gout | 274.1 | ICD10CM | M10.442 | Other secondary gout, left hand |
| Gout | 274.1 | ICD10CM | M10.452 | Other secondary gout, left hip |
| Gout | 274.1 | ICD10CM | M10.419 | Other secondary gout, unspecified shoulder |
| Gout | 274.1 | ICD10CM | M10.43 | Other secondary gout, wrist |
| Gout | 274.1 | ICD10CM | M10.421 | Other secondary gout, right elbow |
| Gout | 274.1 | ICD10CM | M10.46 | Other secondary gout, knee |
| Gout | 274.1 | ICD10CM | M10.40 | Other secondary gout, unspecified site |
| Gout | 274.1 | ICD9CM | 274.89 | Gout with other specified manifestations |
| Gout | 274.1 | ICD10CM | M10.441 | Other secondary gout, right hand |

| | | | | |
|---|---|---|---|---|
| Gout | 274.1 | ICD10CM | M10.451 | Other secondary gout, right hip |
| Gout | 274.1 | ICD10CM | M10 | Gout |
| Gout | 274.1 | ICD10CM | M10.45 | Other secondary gout, hip |
| Gout | 274.1 | ICD10CM | M10.49 | Other secondary gout, multiple sites |
| Gout | 274.1 | ICD10CM | M10.429 | Other secondary gout, unspecified elbow |
| Gout | 274.1 | ICD9CM | 274.82 | Gouty tophi of other sites, except ear |
| Gout | 274.1 | ICD10CM | M10.471 | Other secondary gout, right ankle and foot |
| Gout | 274.1 | ICD10CM | M10.4 | Other secondary gout |
| Gout | 274.1 | ICD10CM | M10.422 | Other secondary gout, left elbow |
| Gout | 274.1 | ICD10CM | M10.479 | Other secondary gout, unspecified ankle and foot |
| Gout | 274.1 | ICD10CM | M1A.4311 | Other secondary chronic gout, right wrist, with tophus (tophi) |
| Gout | 274.1 | ICD10CM | M1A.3290 | Chronic gout due to renal impairment, unspecified elbow, without tophus (tophi) |
| Gout | 274.1 | ICD10CM | M1A.3211 | Chronic gout due to renal impairment, right elbow, with tophus (tophi) |
| Gout | 274.1 | ICD10CM | M1A.00X0 | Idiopathic chronic gout, unspecified site, without tophus (tophi) |
| Gout | 274.1 | ICD10CM | M1A.2110 | Drug-induced chronic gout, right shoulder, without tophus (tophi) |
| Gout | 274.1 | ICD10CM | M1A.21 | Drug-induced chronic gout, shoulder |
| Gout | 274.1 | ICD10CM | M1A.20X1 | Drug-induced chronic gout, unspecified site, with tophus (tophi) |
| Gout | 274.1 | ICD10CM | M1A.0310 | Idiopathic chronic gout, right wrist, without tophus (tophi) |
| Gout | 274.1 | ICD10CM | M10.362 | Gout due to renal impairment, left knee |
| Gout | 274.1 | ICD10CM | M1A.4720 | Other secondary chronic gout, left ankle and foot, without tophus (tophi) |
| Gout | 274.1 | ICD10CM | M1A.40X1 | Other secondary chronic gout, unspecified site, with tophus (tophi) |
| Gout | 274.1 | ICD10CM | M1A.452 | Other secondary chronic gout, left hip |
| Gout | 274.1 | ICD10CM | M1A.4190 | Other secondary chronic gout, unspecified shoulder, without tophus (tophi) |
| Gout | 274.1 | ICD10CM | M1A.261 | Drug-induced chronic gout, right knee |
| Gout | 274.1 | ICD10CM | M1A.419 | Other secondary chronic gout, unspecified shoulder |
| Gout | 274.1 | ICD10CM | M1A.2211 | Drug-induced chronic gout, right elbow, with tophus (tophi) |
| Gout | 274.1 | ICD10CM | M1A.4111 | Other secondary chronic gout, right shoulder, with tophus (tophi) |
| Gout | 274.1 | ICD10CM | M1A.3120 | Chronic gout due to renal impairment, left shoulder, without tophus (tophi) |
| Gout | 274.1 | ICD10CM | M1A.052 | Idiopathic chronic gout, left hip |
| Gout | 274.1 | ICD10CM | M10.379 | Gout due to renal impairment, unspecified ankle and foot |
| Gout | 274.1 | ICD10CM | M1A.359 | Chronic gout due to renal impairment, unspecified hip |

| | | | | |
|---|---|---|---|---|
| Gout | 274.1 | ICD10CM | M1A.3491 | Chronic gout due to renal impairment, unspecified hand, with tophus (tophi) |
| Gout | 274.1 | ICD10CM | M1A.9XX0 | Chronic gout, unspecified, without tophus (tophi) |
| Gout | 274.1 | ICD9CM | 274.8 | Gout with other specified manifestations |
| Gout | 274.1 | ICD10CM | M10.469 | Other secondary gout, unspecified knee |
| Gout | 274.1 | ICD10CM | M1A.23 | Drug-induced chronic gout, wrist |
| Gout | 274.1 | ICD10CM | M10.0 | Idiopathic gout |
| Gout | 274.1 | ICD10CM | M10.049 | Idiopathic gout, unspecified hand |
| Gout | 274.1 | ICD10CM | M1A.2391 | Drug-induced chronic gout, unspecified wrist, with tophus (tophi) |
| Gout | 274.1 | ICD10CM | M10.32 | Gout due to renal impairment, elbow |
| Gout | 274.1 | ICD10CM | M10.04 | Idiopathic gout, hand |
| Gout | 274.1 | ICD10CM | M1A.0391 | Idiopathic chronic gout, unspecified wrist, with tophus (tophi) |
| Gout | 274.1 | ICD10CM | M1A.2791 | Drug-induced chronic gout, unspecified ankle and foot, with tophus (tophi) |
| Gout | 274.1 | ICD9CM | 274.11 | Uric acid nephrolithiasis |
| Gout | 274.1 | ICD10CM | M1A.38X0 | Chronic gout due to renal impairment, vertebrae, without tophus (tophi) |
| Gout | 274.1 | ICD10CM | M1A.3721 | Chronic gout due to renal impairment, left ankle and foot, with tophus (tophi) |
| Gout | 274.1 | ICD10CM | M1A.0120 | Idiopathic chronic gout, left shoulder, without tophus (tophi) |
| Gout | 274.1 | ICD10CM | M1A.079 | Idiopathic chronic gout, unspecified ankle and foot |
| Gout | 274.1 | ICD10CM | M1A.39X0 | Chronic gout due to renal impairment, multiple sites, without tophus (tophi) |
| Gout | 274.1 | ICD10CM | M10.20 | Drug-induced gout, unspecified site |
| Gout | 274.1 | ICD10CM | M1A.2490 | Drug-induced chronic gout, unspecified hand, without tophus (tophi) |
| Gout | 274.1 | ICD10CM | M1A.2291 | Drug-induced chronic gout, unspecified elbow, with tophus (tophi) |
| Gout | 274.1 | ICD10CM | M1A.272 | Drug-induced chronic gout, left ankle and foot |
| Gout | 274.1 | ICD10CM | M1A.411 | Other secondary chronic gout, right shoulder |
| Gout | 274.1 | ICD10CM | M1A.3490 | Chronic gout due to renal impairment, unspecified hand, without tophus (tophi) |
| Gout | 274.1 | ICD10CM | M1A.062 | Idiopathic chronic gout, left knee |
| Gout | 274.1 | ICD10CM | M1A.0191 | Idiopathic chronic gout, unspecified shoulder, with tophus (tophi) |
| Gout | 274.1 | ICD10CM | M1A.0321 | Idiopathic chronic gout, left wrist, with tophus (tophi) |
| Gout | 274.1 | ICD10CM | M1A.231 | Drug-induced chronic gout, right wrist |
| Gout | 274.1 | ICD10CM | M1A.2411 | Drug-induced chronic gout, right hand, with tophus (tophi) |
| Gout | 274.1 | ICD10CM | M1A.369 | Chronic gout due to renal impairment, unspecified knee |
| Gout | 274.1 | ICD10CM | M10.251 | Drug-induced gout, right hip |

| | | | | |
|---|---|---|---|---|
| Gout | 274.1 | ICD10CM | M1A.0721 | Idiopathic chronic gout, left ankle and foot, with tophus (tophi) |
| Gout | 274.1 | ICD10CM | M1A.4121 | Other secondary chronic gout, left shoulder, with tophus (tophi) |
| Gout | 274.1 | ICD10CM | M1A.08X0 | Idiopathic chronic gout, vertebrae, without tophus (tophi) |
| Gout | 274.1 | ICD10CM | M1A.06 | Idiopathic chronic gout, knee |
| Gout | 274.1 | ICD10CM | M1A.42 | Other secondary chronic gout, elbow |
| Gout | 274.1 | ICD10CM | M10.319 | Gout due to renal impairment, unspecified shoulder |
| Gout | 274.1 | ICD10CM | M1A.4220 | Other secondary chronic gout, left elbow, without tophus (tophi) |
| Gout | 274.1 | ICD9CM | 274.1 | Gouty nephropathy, unspecified |
| Gout | 274.1 | ICD10CM | M1A.09 | Idiopathic chronic gout, multiple sites |
| Gout | 274.1 | ICD10CM | M1A.459 | Other secondary chronic gout, unspecified hip |
| Gout | 274.1 | ICD10CM | M10.069 | Idiopathic gout, unspecified knee |
| Gout | 274.1 | ICD10CM | M1A.429 | Other secondary chronic gout, unspecified elbow |
| Gout | 274.1 | ICD10CM | M1A.249 | Drug-induced chronic gout, unspecified hand |
| Gout | 274.1 | ICD10CM | M1A.2510 | Drug-induced chronic gout, right hip, without tophus (tophi) |
| Gout | 274.1 | ICD10CM | M1A.4211 | Other secondary chronic gout, right elbow, with tophus (tophi) |
| Gout | 274.1 | ICD10CM | M10.19 | Lead-induced gout, multiple sites |
| Gout | 274.1 | ICD10CM | M1A.251 | Drug-induced chronic gout, right hip |
| Gout | 274.1 | ICD10CM | M1A.0221 | Idiopathic chronic gout, left elbow, with tophus (tophi) |
| Gout | 274.1 | ICD10CM | M10.151 | Lead-induced gout, right hip |
| Gout | 274.1 | ICD10CM | M10.39 | Gout due to renal impairment, multiple sites |
| Gout | 274.1 | ICD10CM | M1A.3510 | Chronic gout due to renal impairment, right hip, without tophus (tophi) |
| Gout | 274.1 | ICD9CM | 274 | Gouty arthropathy, unspecified |
| Gout | 274.1 | ICD10CM | M10.29 | Drug-induced gout, multiple sites |
| Gout | 274.1 | ICD10CM | M10.30 | Gout due to renal impairment, unspecified site |
| Gout | 274.1 | ICD10CM | M1A.3720 | Chronic gout due to renal impairment, left ankle and foot, without tophus (tophi) |
| Gout | 274.1 | ICD10CM | M1A.2711 | Drug-induced chronic gout, right ankle and foot, with tophus (tophi) |
| Gout | 274.1 | ICD10CM | M1A.3690 | Chronic gout due to renal impairment, unspecified knee, without tophus (tophi) |
| Gout | 274.1 | ICD10CM | M1A.0711 | Idiopathic chronic gout, right ankle and foot, with tophus (tophi) |
| Gout | 274.1 | ICD10CM | M1A.0620 | Idiopathic chronic gout, left knee, without tophus (tophi) |
| Gout | 274.1 | ICD10CM | M1A.48X0 | Other secondary chronic gout, vertebrae, without tophus (tophi) |

| | | | | |
|---|---|---|---|---|
| Gout | 274.1 | ICD10CM | M1A.0320 | Idiopathic chronic gout, left wrist, without tophus (tophi) |
| Gout | 274.1 | ICD10CM | M1A.4711 | Other secondary chronic gout, right ankle and foot, with tophus (tophi) |
| Gout | 274.1 | ICD10CM | M10.241 | Drug-induced gout, right hand |
| Gout | 274.1 | ICD10CM | M10.021 | Idiopathic gout, right elbow |
| Gout | 274.1 | ICD10CM | M10.119 | Lead-induced gout, unspecified shoulder |
| Gout | 274.1 | ICD10CM | M1A.412 | Other secondary chronic gout, left shoulder |
| Gout | 274.1 | ICD10CM | M10.35 | Gout due to renal impairment, hip |
| Gout | 274.1 | ICD10CM | M1A.4320 | Other secondary chronic gout, left wrist, without tophus (tophi) |
| Gout | 274.1 | ICD10CM | M10.01 | Idiopathic gout, shoulder |
| Gout | 274.1 | ICD10CM | M10.042 | Idiopathic gout, left hand |
| Gout | 274.1 | ICD10CM | M1A.421 | Other secondary chronic gout, right elbow |
| Gout | 274.1 | ICD10CM | M1A.0720 | Idiopathic chronic gout, left ankle and foot, without tophus (tophi) |
| Gout | 274.1 | ICD10CM | M1A.2721 | Drug-induced chronic gout, left ankle and foot, with tophus (tophi) |
| Gout | 274.1 | ICD10CM | M1A.28 | Drug-induced chronic gout, vertebrae |
| Gout | 274.1 | ICD10CM | M1A.04 | Idiopathic chronic gout, hand |
| Gout | 274.1 | ICD10CM | M1A.3390 | Chronic gout due to renal impairment, unspecified wrist, without tophus (tophi) |
| Gout | 274.1 | ICD10CM | M1A.47 | Other secondary chronic gout, ankle and foot |
| Gout | 274.1 | ICD10CM | M1A.441 | Other secondary chronic gout, right hand |
| Gout | 274.1 | ICD10CM | M1A.042 | Idiopathic chronic gout, left hand |
| Gout | 274.1 | ICD10CM | M1A.342 | Chronic gout due to renal impairment, left hand |
| Gout | 274.1 | ICD10CM | M1A.051 | Idiopathic chronic gout, right hip |
| Gout | 274.1 | ICD10CM | M1A.4491 | Other secondary chronic gout, unspecified hand, with tophus (tophi) |
| Gout | 274.1 | ICD10CM | M10.259 | Drug-induced gout, unspecified hip |
| Gout | 274.1 | ICD10CM | M1A.3520 | Chronic gout due to renal impairment, left hip, without tophus (tophi) |
| Gout | 274.1 | ICD10CM | M10.27 | Drug-induced gout, ankle and foot |
| Gout | 274.1 | ICD10CM | M1A.00 | Idiopathic chronic gout, unspecified site |
| Gout | 274.1 | ICD10CM | M1A.259 | Drug-induced chronic gout, unspecified hip |
| Gout | 274.1 | ICD10CM | M1A.4420 | Other secondary chronic gout, left hand, without tophus (tophi) |
| Gout | 274.1 | ICD10CM | M1A.032 | Idiopathic chronic gout, left wrist |
| Gout | 274.1 | ICD10CM | M1A.4710 | Other secondary chronic gout, right ankle and foot, without tophus (tophi) |
| Gout | 274.1 | ICD10CM | M1A.3320 | Chronic gout due to renal impairment, left wrist, without tophus (tophi) |
| Gout | 274.1 | ICD10CM | M1A.3221 | Chronic gout due to renal impairment, left elbow, with tophus (tophi) |

| | | | | |
|---|---|---|---|---|
| Gout | 274.1 | ICD10CM | M1A.0420 | Idiopathic chronic gout, left hand, without tophus (tophi) |
| Gout | 274.1 | ICD10CM | M1A.0510 | Idiopathic chronic gout, right hip, without tophus (tophi) |
| Gout | 274.1 | ICD10CM | M1A.4590 | Other secondary chronic gout, unspecified hip, without tophus (tophi) |
| Gout | 274.1 | ICD10CM | M1A.461 | Other secondary chronic gout, right knee |
| Gout | 274.1 | ICD10CM | M1A.0110 | Idiopathic chronic gout, right shoulder, without tophus (tophi) |
| Gout | 274.1 | ICD10CM | M10.219 | Drug-induced gout, unspecified shoulder |
| Gout | 274.1 | ICD10CM | M1A.45 | Other secondary chronic gout, hip |
| Gout | 274.1 | ICD10CM | M1A.30X1 | Chronic gout due to renal impairment, unspecified site, with tophus (tophi) |
| Gout | 274.1 | ICD10CM | M1A.0290 | Idiopathic chronic gout, unspecified elbow, without tophus (tophi) |
| Gout | 274.1 | ICD10CM | M10.23 | Drug-induced gout, wrist |
| Gout | 274.1 | ICD10CM | M1A.0390 | Idiopathic chronic gout, unspecified wrist, without tophus (tophi) |
| Gout | 274.1 | ICD10CM | M1A.33 | Chronic gout due to renal impairment, wrist |
| Gout | 274.1 | ICD10CM | M1A.4791 | Other secondary chronic gout, unspecified ankle and foot, with tophus (tophi) |
| Gout | 274.1 | ICD10CM | M1A.331 | Chronic gout due to renal impairment, right wrist |
| Gout | 274.1 | ICD10CM | M1A.341 | Chronic gout due to renal impairment, right hand |
| Gout | 274.1 | ICD10CM | M10.149 | Lead-induced gout, unspecified hand |
| Gout | 274.1 | ICD10CM | M1A.49X0 | Other secondary chronic gout, multiple sites, without tophus (tophi) |
| Gout | 274.1 | ICD10CM | M1A.3511 | Chronic gout due to renal impairment, right hip, with tophus (tophi) |
| Gout | 274.1 | ICD10CM | M10.152 | Lead-induced gout, left hip |
| Gout | 274.1 | ICD10CM | M1A.4410 | Other secondary chronic gout, right hand, without tophus (tophi) |
| Gout | 274.1 | ICD10CM | M1A.3410 | Chronic gout due to renal impairment, right hand, without tophus (tophi) |
| Gout | 274.1 | ICD10CM | M1A.4511 | Other secondary chronic gout, right hip, with tophus (tophi) |
| Gout | 274.1 | ICD10CM | M1A.3421 | Chronic gout due to renal impairment, left hand, with tophus (tophi) |
| Gout | 274.1 | ICD10CM | M1A.029 | Idiopathic chronic gout, unspecified elbow |
| Gout | 274.1 | ICD10CM | M1A.3291 | Chronic gout due to renal impairment, unspecified elbow, with tophus (tophi) |
| Gout | 274.1 | ICD10CM | M1A.2191 | Drug-induced chronic gout, unspecified shoulder, with tophus (tophi) |
| Gout | 274.1 | ICD10CM | M10.352 | Gout due to renal impairment, left hip |
| Gout | 274.1 | ICD10CM | M10.212 | Drug-induced gout, left shoulder |
| Gout | 274.1 | ICD10CM | M1A.2210 | Drug-induced chronic gout, right elbow, without tophus (tophi) |
| Gout | 274.1 | ICD10CM | M1A.20 | Drug-induced chronic gout, unspecified site |

| | | | | |
|---|---|---|---|---|
| Gout | 274.1 | ICD10CM | M10.269 | Drug-induced gout, unspecified knee |
| Gout | 274.1 | ICD10CM | M1A.0291 | Idiopathic chronic gout, unspecified elbow, with tophus (tophi) |
| Gout | 274.1 | ICD10CM | M1A.0410 | Idiopathic chronic gout, right hand, without tophus (tophi) |
| Gout | 274.1 | ICD10CM | M1A.3391 | Chronic gout due to renal impairment, unspecified wrist, with tophus (tophi) |
| Gout | 274.1 | ICD10CM | M1A.211 | Drug-induced chronic gout, right shoulder |
| Gout | 274.1 | ICD10CM | M1A.3420 | Chronic gout due to renal impairment, left hand, without tophus (tophi) |
| Gout | 274.1 | ICD10CM | M1A.351 | Chronic gout due to renal impairment, right hip |
| Gout | 274.1 | ICD10CM | M10.129 | Lead-induced gout, unspecified elbow |
| Gout | 274.1 | ICD10CM | M1A.0491 | Idiopathic chronic gout, unspecified hand, with tophus (tophi) |
| Gout | 274.1 | ICD10CM | M1A.4721 | Other secondary chronic gout, left ankle and foot, with tophus (tophi) |
| Gout | 274.1 | ICD10CM | M1A.0190 | Idiopathic chronic gout, unspecified shoulder, without tophus (tophi) |
| Gout | 274.1 | ICD10CM | M1A.4591 | Other secondary chronic gout, unspecified hip, with tophus (tophi) |
| Gout | 274.1 | ICD10CM | M10.052 | Idiopathic gout, left hip |
| Gout | 274.1 | ICD10CM | M10.15 | Lead-induced gout, hip |
| Gout | 274.1 | ICD10CM | M1A.222 | Drug-induced chronic gout, left elbow |
| Gout | 274.1 | ICD10CM | M1A.4120 | Other secondary chronic gout, left shoulder, without tophus (tophi) |
| Gout | 274.1 | ICD10CM | M1A.3521 | Chronic gout due to renal impairment, left hip, with tophus (tophi) |
| Gout | 274.1 | ICD10CM | M1A.0511 | Idiopathic chronic gout, right hip, with tophus (tophi) |
| Gout | 274.1 | ICD10CM | M10.131 | Lead-induced gout, right wrist |
| Gout | 274.1 | ICD10CM | M1A.449 | Other secondary chronic gout, unspecified hand |
| Gout | 274.1 | ICD10CM | M1A.25 | Drug-induced chronic gout, hip |
| Gout | 274.1 | ICD10CM | M10.159 | Lead-induced gout, unspecified hip |
| Gout | 274.1 | ICD10CM | M1A.4520 | Other secondary chronic gout, left hip, without tophus (tophi) |
| Gout | 274.1 | ICD10CM | M1A.0211 | Idiopathic chronic gout, right elbow, with tophus (tophi) |
| Gout | 274.1 | ICD10CM | M1A.3110 | Chronic gout due to renal impairment, right shoulder, without tophus (tophi) |
| Gout | 274.1 | ICD10CM | M10.262 | Drug-induced gout, left knee |
| Gout | 274.1 | ICD10CM | M1A.3621 | Chronic gout due to renal impairment, left knee, with tophus (tophi) |
| Gout | 274.1 | ICD10CM | M1A.012 | Idiopathic chronic gout, left shoulder |
| Gout | 274.1 | ICD10CM | M10.179 | Lead-induced gout, unspecified ankle and foot |
| Gout | 274.1 | ICD10CM | M1A.041 | Idiopathic chronic gout, right hand |
| Gout | 274.1 | ICD10CM | M10.07 | Idiopathic gout, ankle and foot |

| | | | | |
|---|---|---|---|---|
| Gout | 274.1 | ICD10CM | M1A.3310 | Chronic gout due to renal impairment, right wrist, without tophus (tophi) |
| Gout | 274.1 | ICD10CM | M1A.339 | Chronic gout due to renal impairment, unspecified wrist |
| Gout | 274.1 | ICD10CM | M1A.322 | Chronic gout due to renal impairment, left elbow |
| Gout | 274.1 | ICD10CM | M1A.34 | Chronic gout due to renal impairment, hand |
| Gout | 274.1 | ICD10CM | M10.18 | Lead-induced gout, vertebrae |
| Gout | 274.1 | ICD10CM | M1A.09X1 | Idiopathic chronic gout, multiple sites, with tophus (tophi) |
| Gout | 274.1 | ICD10CM | M1A.3710 | Chronic gout due to renal impairment, right ankle and foot, without tophus (tophi) |
| Gout | 274.1 | ICD10CM | M10.09 | Idiopathic gout, multiple sites |
| Gout | 274.1 | ICD10CM | M1A.3411 | Chronic gout due to renal impairment, right hand, with tophus (tophi) |
| Gout | 274.1 | ICD10CM | M1A.00X1 | Idiopathic chronic gout, unspecified site, with tophus (tophi) |
| Gout | 274.1 | ICD10CM | M10.16 | Lead-induced gout, knee |
| Gout | 274.1 | ICD10CM | M1A.4191 | Other secondary chronic gout, unspecified shoulder, with tophus (tophi) |
| Gout | 274.1 | ICD10CM | M10.132 | Lead-induced gout, left wrist |
| Gout | 274.1 | ICD10CM | M1A.229 | Drug-induced chronic gout, unspecified elbow |
| Gout | 274.1 | ICD10CM | M10.122 | Lead-induced gout, left elbow |
| Gout | 274.1 | ICD10CM | M1A.0591 | Idiopathic chronic gout, unspecified hip, with tophus (tophi) |
| Gout | 274.1 | ICD10CM | M1A.472 | Other secondary chronic gout, left ankle and foot |
| Gout | 274.1 | ICD10CM | M1A.30 | Chronic gout due to renal impairment, unspecified site |
| Gout | 274.1 | ICD10CM | M10.111 | Lead-induced gout, right shoulder |
| Gout | 274.1 | ICD10CM | M10.032 | Idiopathic gout, left wrist |
| Gout | 274.1 | ICD10CM | M1A.2591 | Drug-induced chronic gout, unspecified hip, with tophus (tophi) |
| Gout | 274.1 | ICD10CM | M1A.3590 | Chronic gout due to renal impairment, unspecified hip, without tophus (tophi) |
| Gout | 274.1 | ICD10CM | M10.03 | Idiopathic gout, wrist |
| Gout | 274.1 | ICD10CM | M10.221 | Drug-induced gout, right elbow |
| Gout | 274.1 | ICD10CM | M1A.2621 | Drug-induced chronic gout, left knee, with tophus (tophi) |
| Gout | 274.1 | ICD10CM | M10.242 | Drug-induced gout, left hand |
| Gout | 274.1 | ICD10CM | M1A.0210 | Idiopathic chronic gout, right elbow, without tophus (tophi) |
| Gout | 274.1 | ICD10CM | M1A.4611 | Other secondary chronic gout, right knee, with tophus (tophi) |
| Gout | 274.1 | ICD10CM | M1A.43 | Other secondary chronic gout, wrist |
| Gout | 274.1 | ICD10CM | M10.229 | Drug-induced gout, unspecified elbow |
| Gout | 274.1 | ICD10CM | M1A.0521 | Idiopathic chronic gout, left hip, with tophus (tophi) |

| | | | | |
|---|---|---|---|---|
| Gout | 274.1 | ICD10CM | M10.22 | Drug-induced gout, elbow |
| Gout | 274.1 | ICD10CM | M1A.3121 | Chronic gout due to renal impairment, left shoulder, with tophus (tophi) |
| Gout | 274.1 | ICD10CM | M1A.4421 | Other secondary chronic gout, left hand, with tophus (tophi) |
| Gout | 274.1 | ICD10CM | M1A.08X1 | Idiopathic chronic gout, vertebrae, with tophus (tophi) |
| Gout | 274.1 | ICD10CM | M1A.019 | Idiopathic chronic gout, unspecified shoulder |
| Gout | 274.1 | ICD10CM | M1A.29 | Drug-induced chronic gout, multiple sites |
| Gout | 274.1 | ICD10CM | M10.041 | Idiopathic gout, right hand |
| Gout | 274.1 | ICD10CM | M1A.36 | Chronic gout due to renal impairment, knee |
| Gout | 274.1 | ICD10CM | M10.031 | Idiopathic gout, right wrist |
| Gout | 274.1 | ICD10CM | M1A.0311 | Idiopathic chronic gout, right wrist, with tophus (tophi) |
| Gout | 274.1 | ICD10CM | M1A.0111 | Idiopathic chronic gout, right shoulder, with tophus (tophi) |
| Gout | 274.1 | ICD10CM | M10.21 | Drug-induced gout, shoulder |
| Gout | 274.1 | ICD10CM | M10.05 | Idiopathic gout, hip |
| Gout | 274.1 | ICD10CM | M1A.05 | Idiopathic chronic gout, hip |
| Gout | 274.1 | ICD10CM | M1A.022 | Idiopathic chronic gout, left elbow |
| Gout | 274.1 | ICD10CM | M10.022 | Idiopathic gout, left elbow |
| Gout | 274.1 | ICD10CM | M1A.32 | Chronic gout due to renal impairment, elbow |
| Gout | 274.1 | ICD10CM | M1A.3311 | Chronic gout due to renal impairment, right wrist, with tophus (tophi) |
| Gout | 274.1 | ICD10CM | M1A.29X0 | Drug-induced chronic gout, multiple sites, without tophus (tophi) |
| Gout | 274.1 | ICD10CM | M1A.471 | Other secondary chronic gout, right ankle and foot |
| Gout | 274.1 | ICD10CM | M1A.0691 | Idiopathic chronic gout, unspecified knee, with tophus (tophi) |
| Gout | 274.1 | ICD10CM | M10.2 | Drug-induced gout |
| Gout | 274.1 | ICD10CM | M1A.30X0 | Chronic gout due to renal impairment, unspecified site, without tophus (tophi) |
| Gout | 274.1 | ICD10CM | M1A.0610 | Idiopathic chronic gout, right knee, without tophus (tophi) |
| Gout | 274.1 | ICD10CM | M1A.432 | Other secondary chronic gout, left wrist |
| Gout | 274.1 | ICD10CM | M1A.479 | Other secondary chronic gout, unspecified ankle and foot |
| Gout | 274.1 | ICD10CM | M1A.03 | Idiopathic chronic gout, wrist |
| Gout | 274.1 | ICD10CM | M1A.332 | Chronic gout due to renal impairment, left wrist |
| Gout | 274.1 | ICD10CM | M1A.352 | Chronic gout due to renal impairment, left hip |
| Gout | 274.1 | ICD10CM | M1A.4390 | Other secondary chronic gout, unspecified wrist, without tophus (tophi) |
| Gout | 274.1 | ICD10CM | M1A.3591 | Chronic gout due to renal impairment, unspecified hip, with tophus (tophi) |

| | | | | |
|---|---|---|---|---|
| Gout | 274.1 | ICD10CM | M1A.372 | Chronic gout due to renal impairment, left ankle and foot |
| Gout | 274.1 | ICD10CM | M1A.4310 | Other secondary chronic gout, right wrist, without tophus (tophi) |
| Gout | 274.1 | ICD10CM | M1A.379 | Chronic gout due to renal impairment, unspecified ankle and foot |
| Gout | 274.1 | ICD10CM | M1A.362 | Chronic gout due to renal impairment, left knee |
| Gout | 274.1 | ICD10CM | M10.071 | Idiopathic gout, right ankle and foot |
| Gout | 274.1 | ICD10CM | M1A.4690 | Other secondary chronic gout, unspecified knee, without tophus (tophi) |
| Gout | 274.1 | ICD10CM | M1A.0690 | Idiopathic chronic gout, unspecified knee, without tophus (tophi) |
| Gout | 274.1 | ICD10CM | M1A.3691 | Chronic gout due to renal impairment, unspecified knee, with tophus (tophi) |
| Gout | 274.1 | ICD10CM | M10.169 | Lead-induced gout, unspecified knee |
| Gout | 274.1 | ICD10CM | M1A.2310 | Drug-induced chronic gout, right wrist, without tophus (tophi) |
| Gout | 274.1 | ICD10CM | M1A.39X1 | Chronic gout due to renal impairment, multiple sites, with tophus (tophi) |
| Gout | 274.1 | ICD10CM | M10.321 | Gout due to renal impairment, right elbow |
| Gout | 274.1 | ICD10CM | M1A.2710 | Drug-induced chronic gout, right ankle and foot, without tophus (tophi) |
| Gout | 274.1 | ICD10CM | M10.051 | Idiopathic gout, right hip |
| Gout | 274.1 | ICD10CM | M1A.3790 | Chronic gout due to renal impairment, unspecified ankle and foot, without tophus (tophi) |
| Gout | 274.1 | ICD10CM | M10.339 | Gout due to renal impairment, unspecified wrist |
| Gout | 274.1 | ICD10CM | M1A.2321 | Drug-induced chronic gout, left wrist, with tophus (tophi) |
| Gout | 274.1 | ICD10CM | M1A.28X0 | Drug-induced chronic gout, vertebrae, without tophus (tophi) |
| Gout | 274.1 | ICD10CM | M10.329 | Gout due to renal impairment, unspecified elbow |
| Gout | 274.1 | ICD10CM | M10.331 | Gout due to renal impairment, right wrist |
| Gout | 274.1 | ICD10CM | M10.31 | Gout due to renal impairment, shoulder |
| Gout | 274.1 | ICD10CM | M1A.069 | Idiopathic chronic gout, unspecified knee |
| Gout | 274.1 | ICD10CM | M1A.3220 | Chronic gout due to renal impairment, left elbow, without tophus (tophi) |
| Gout | 274.1 | ICD10CM | M10.121 | Lead-induced gout, right elbow |
| Gout | 274.1 | ICD10CM | M1A.422 | Other secondary chronic gout, left elbow |
| Gout | 274.1 | ICD10CM | M1A.2111 | Drug-induced chronic gout, right shoulder, with tophus (tophi) |
| Gout | 274.1 | ICD10CM | M10.261 | Drug-induced gout, right knee |
| Gout | 274.1 | ICD10CM | M1A.0611 | Idiopathic chronic gout, right knee, with tophus (tophi) |
| Gout | 274.1 | ICD10CM | M10.141 | Lead-induced gout, right hand |
| Gout | 274.1 | ICD10CM | M10.26 | Drug-induced gout, knee |
| Gout | 274.1 | ICD10CM | M10.249 | Drug-induced gout, unspecified hand |

| | | | | |
|---|---|---|---|---|
| Gout | 274.1 | ICD10CM | M1A.4110 | Other secondary chronic gout, right shoulder, without tophus (tophi) |
| Gout | 274.1 | ICD10CM | M1A.241 | Drug-induced chronic gout, right hand |
| Gout | 274.1 | ICD10CM | M10.341 | Gout due to renal impairment, right hand |
| Gout | 274.1 | ICD10CM | M1A.232 | Drug-induced chronic gout, left wrist |
| Gout | 274.1 | ICD10CM | M1A.2590 | Drug-induced chronic gout, unspecified hip, without tophus (tophi) |
| Gout | 274.1 | ICD10CM | M10.00 | Idiopathic gout, unspecified site |
| Gout | 274.1 | ICD10CM | M1A.4290 | Other secondary chronic gout, unspecified elbow, without tophus (tophi) |
| Gout | 274.1 | ICD10CM | M10.36 | Gout due to renal impairment, knee |
| Gout | 274.1 | ICD10CM | M10.112 | Lead-induced gout, left shoulder |
| Gout | 274.1 | ICD10CM | M1A.38X1 | Chronic gout due to renal impairment, vertebrae, with tophus (tophi) |
| Gout | 274.1 | ICD10CM | M1A.269 | Drug-induced chronic gout, unspecified knee |
| Gout | 274.1 | ICD10CM | M1A.262 | Drug-induced chronic gout, left knee |
| Gout | 274.1 | ICD10CM | M1A.2620 | Drug-induced chronic gout, left knee, without tophus (tophi) |
| Gout | 274.1 | ICD10CM | M10.38 | Gout due to renal impairment, vertebrae |
| Gout | 274.1 | ICD10CM | M10.372 | Gout due to renal impairment, left ankle and foot |
| Gout | 274.1 | ICD10CM | M1A.4 | Other secondary chronic gout |
| Gout | 274.1 | ICD10CM | M10.271 | Drug-induced gout, right ankle and foot |
| Gout | 274.1 | ICD10CM | M1A.2390 | Drug-induced chronic gout, unspecified wrist, without tophus (tophi) |
| Gout | 274.1 | ICD10CM | M1A.0621 | Idiopathic chronic gout, left knee, with tophus (tophi) |
| Gout | 274.1 | ICD10CM | M1A.09X0 | Idiopathic chronic gout, multiple sites, without tophus (tophi) |
| Gout | 274.1 | ICD10CM | M1A.071 | Idiopathic chronic gout, right ankle and foot |
| Gout | 274.1 | ICD10CM | M1A.27 | Drug-induced chronic gout, ankle and foot |
| Gout | 274.1 | ICD10CM | M1A.2690 | Drug-induced chronic gout, unspecified knee, without tophus (tophi) |
| Gout | 274.1 | ICD10CM | M1A.3620 | Chronic gout due to renal impairment, left knee, without tophus (tophi) |
| Gout | 274.1 | ICD10CM | M1A.072 | Idiopathic chronic gout, left ankle and foot |
| Gout | 274.1 | ICD10CM | M1A.279 | Drug-induced chronic gout, unspecified ankle and foot |
| Gout | 274.1 | ICD10CM | M1A.26 | Drug-induced chronic gout, knee |
| Gout | 274.1 | ICD10CM | M1A.2790 | Drug-induced chronic gout, unspecified ankle and foot, without tophus (tophi) |
| Gout | 274.1 | ICD10CM | M1A.38 | Chronic gout due to renal impairment, vertebrae |
| Gout | 274.1 | ICD10CM | M10.342 | Gout due to renal impairment, left hand |
| Gout | 274.1 | ICD10CM | M1A.4620 | Other secondary chronic gout, left knee, without tophus (tophi) |
| Gout | 274.1 | ICD10CM | M1A.4691 | Other secondary chronic gout, unspecified knee, with tophus (tophi) |

| | | | | |
|---|---|---|---|---|
| Gout | 274.1 | ICD10CM | M10.312 | Gout due to renal impairment, left shoulder |
| Gout | 274.1 | ICD10CM | M10.10 | Lead-induced gout, unspecified site |
| Gout | 274.1 | ICD10CM | M10.11 | Lead-induced gout, shoulder |
| Gout | 274.1 | ICD10CM | M1A.329 | Chronic gout due to renal impairment, unspecified elbow |
| Gout | 274.1 | ICD10CM | M1A.4621 | Other secondary chronic gout, left knee, with tophus (tophi) |
| Gout | 274.1 | ICD10CM | M1A.469 | Other secondary chronic gout, unspecified knee |
| Gout | 274.1 | ICD10CM | M1A.039 | Idiopathic chronic gout, unspecified wrist |
| Gout | 274.1 | ICD10CM | M1A.312 | Chronic gout due to renal impairment, left shoulder |
| Gout | 274.1 | ICD10CM | M10.24 | Drug-induced gout, hand |
| Gout | 274.1 | ICD10CM | M1A.3111 | Chronic gout due to renal impairment, right shoulder, with tophus (tophi) |
| Gout | 274.1 | ICD10CM | M1A.0 | Idiopathic chronic gout |
| Gout | 274.1 | ICD10CM | M1A.4490 | Other secondary chronic gout, unspecified hand, without tophus (tophi) |
| Gout | 274.1 | ICD10CM | M1A.2611 | Drug-induced chronic gout, right knee, with tophus (tophi) |
| Gout | 274.1 | ICD10CM | M1A.349 | Chronic gout due to renal impairment, unspecified hand |
| Gout | 274.1 | ICD10CM | M1A.2190 | Drug-induced chronic gout, unspecified shoulder, without tophus (tophi) |
| Gout | 274.1 | ICD10CM | M1A.462 | Other secondary chronic gout, left knee |
| Gout | 274.1 | ICD10CM | M1A.221 | Drug-induced chronic gout, right elbow |
| Gout | 274.1 | ICD10CM | M1A.4521 | Other secondary chronic gout, left hip, with tophus (tophi) |
| Gout | 274.1 | ICD10CM | M10.252 | Drug-induced gout, left hip |
| Gout | 274.1 | ICD10CM | M1A.2221 | Drug-induced chronic gout, left elbow, with tophus (tophi) |
| Gout | 274.1 | ICD10CM | M1A.0121 | Idiopathic chronic gout, left shoulder, with tophus (tophi) |
| Gout | 274.1 | ICD10CM | M1A.3191 | Chronic gout due to renal impairment, unspecified shoulder, with tophus (tophi) |
| Gout | 274.1 | ICD10CM | M1A.3210 | Chronic gout due to renal impairment, right elbow, without tophus (tophi) |
| Gout | 274.1 | ICD10CM | M1A.4510 | Other secondary chronic gout, right hip, without tophus (tophi) |
| Gout | 274.1 | ICD10CM | M10.222 | Drug-induced gout, left elbow |
| Gout | 274.1 | ICD10CM | M10.239 | Drug-induced gout, unspecified wrist |
| Gout | 274.1 | ICD10CM | M1A.2121 | Drug-induced chronic gout, left shoulder, with tophus (tophi) |
| Gout | 274.1 | ICD10CM | M1A.01 | Idiopathic chronic gout, shoulder |
| Gout | 274.1 | ICD10CM | M1A.0421 | Idiopathic chronic gout, left hand, with tophus (tophi) |
| Gout | 274.1 | ICD10CM | M1A.49 | Other secondary chronic gout, multiple sites |
| Gout | 274.1 | ICD10CM | M10.142 | Lead-induced gout, left hand |

| | | | | |
|---|---|---|---|---|
| Gout | 274.1 | ICD10CM | M1A.321 | Chronic gout due to renal impairment, right elbow |
| Gout | 274.1 | ICD10CM | M1A.46 | Other secondary chronic gout, knee |
| Gout | 274.1 | ICD10CM | M10.14 | Lead-induced gout, hand |
| Gout | 274.1 | ICD10CM | M10.231 | Drug-induced gout, right wrist |
| Gout | 274.1 | ICD10CM | M10.232 | Drug-induced gout, left wrist |
| Gout | 274.1 | ICD10CM | M1A.49X1 | Other secondary chronic gout, multiple sites, with tophus (tophi) |
| Gout | 274.1 | ICD10CM | M1A.35 | Chronic gout due to renal impairment, hip |
| Gout | 274.1 | ICD10CM | M10.361 | Gout due to renal impairment, right knee |
| Gout | 274.1 | ICD10CM | M10.1 | Lead-induced gout |
| Gout | 274.1 | ICD10CM | M1A.4610 | Other secondary chronic gout, right knee, without tophus (tophi) |
| Gout | 274.1 | ICD10CM | M1A.08 | Idiopathic chronic gout, vertebrae |
| Gout | 274.1 | ICD10CM | M1A.07 | Idiopathic chronic gout, ankle and foot |
| Gout | 274.1 | ICD10CM | M10.012 | Idiopathic gout, left shoulder |
| Gout | 274.1 | ICD10CM | M1A.0710 | Idiopathic chronic gout, right ankle and foot, without tophus (tophi) |
| Gout | 274.1 | ICD10CM | M1A.37 | Chronic gout due to renal impairment, ankle and foot |
| Gout | 274.1 | ICD10CM | M1A.2120 | Drug-induced chronic gout, left shoulder, without tophus (tophi) |
| Gout | 274.1 | ICD10CM | M1A.40 | Other secondary chronic gout, unspecified site |
| Gout | 274.1 | ICD10CM | M1A.4391 | Other secondary chronic gout, unspecified wrist, with tophus (tophi) |
| Gout | 274.1 | ICD10CM | M1A.212 | Drug-induced chronic gout, left shoulder |
| Gout | 274.1 | ICD10CM | M1A.41 | Other secondary chronic gout, shoulder |
| Gout | 274.1 | ICD10CM | M1A.2520 | Drug-induced chronic gout, left hip, without tophus (tophi) |
| Gout | 274.1 | ICD10CM | M10.039 | Idiopathic gout, unspecified wrist |
| Gout | 274.1 | ICD10CM | M1A.4321 | Other secondary chronic gout, left wrist, with tophus (tophi) |
| Gout | 274.1 | ICD10CM | M1A.3611 | Chronic gout due to renal impairment, right knee, with tophus (tophi) |
| Gout | 274.1 | ICD10CM | M1A.219 | Drug-induced chronic gout, unspecified shoulder |
| Gout | 274.1 | ICD10CM | M1A.2320 | Drug-induced chronic gout, left wrist, without tophus (tophi) |
| Gout | 274.1 | ICD10CM | M10.019 | Idiopathic gout, unspecified shoulder |
| Gout | 274.1 | ICD10CM | M10.349 | Gout due to renal impairment, unspecified hand |
| Gout | 274.1 | ICD10CM | M1A.2691 | Drug-induced chronic gout, unspecified knee, with tophus (tophi) |
| Gout | 274.1 | ICD10CM | M1A.2220 | Drug-induced chronic gout, left elbow, without tophus (tophi) |
| Gout | 274.1 | ICD10CM | M1A.061 | Idiopathic chronic gout, right knee |
| Gout | 274.1 | ICD10CM | M10.272 | Drug-induced gout, left ankle and foot |

| | | | | |
|---|---|---|---|---|
| Gout | 274.1 | ICD10CM | M1A.0590 | Idiopathic chronic gout, unspecified hip, without tophus (tophi) |
| Gout | 274.1 | ICD10CM | M10.28 | Drug-induced gout, vertebrae |
| Gout | 274.1 | ICD10CM | M1A.451 | Other secondary chronic gout, right hip |
| Gout | 274.1 | ICD10CM | M1A.3190 | Chronic gout due to renal impairment, unspecified shoulder, without tophus (tophi) |
| Gout | 274.1 | ICD10CM | M10.161 | Lead-induced gout, right knee |
| Gout | 274.1 | ICD10CM | M1A.3610 | Chronic gout due to renal impairment, right knee, without tophus (tophi) |
| Gout | 274.1 | ICD10CM | M1A.4291 | Other secondary chronic gout, unspecified elbow, with tophus (tophi) |
| Gout | 274.1 | ICD10CM | M1A.021 | Idiopathic chronic gout, right elbow |
| Gout | 274.1 | ICD10CM | M1A.431 | Other secondary chronic gout, right wrist |
| Gout | 274.1 | ICD10CM | M10.08 | Idiopathic gout, vertebrae |
| Gout | 274.1 | ICD10CM | M1A.28X1 | Drug-induced chronic gout, vertebrae, with tophus (tophi) |
| Gout | 274.1 | ICD10CM | M10.33 | Gout due to renal impairment, wrist |
| Gout | 274.1 | ICD10CM | M10.072 | Idiopathic gout, left ankle and foot |
| Gout | 274.1 | ICD10CM | M10.211 | Drug-induced gout, right shoulder |
| Gout | 274.1 | ICD10CM | M1A.361 | Chronic gout due to renal impairment, right knee |
| Gout | 274.1 | ICD10CM | M1A.031 | Idiopathic chronic gout, right wrist |
| Gout | 274.1 | ICD9CM | 274.19 | Other gouty nephropathy |
| Gout | 274.1 | ICD10CM | M1A.439 | Other secondary chronic gout, unspecified wrist |
| Gout | 274.1 | ICD10CM | M1A.48X1 | Other secondary chronic gout, vertebrae, with tophus (tophi) |
| Gout | 274.1 | ICD10CM | M1A.2 | Drug-induced chronic gout |
| Gout | 274.1 | ICD10CM | M1A.20X0 | Drug-induced chronic gout, unspecified site, without tophus (tophi) |
| Gout | 274.1 | ICD10CM | M1A.0411 | Idiopathic chronic gout, right hand, with tophus (tophi) |
| Gout | 274.1 | ICD10CM | M10.12 | Lead-induced gout, elbow |
| Gout | 274.1 | ICD10CM | M10.25 | Drug-induced gout, hip |
| Gout | 274.1 | ICD10CM | M10.371 | Gout due to renal impairment, right ankle and foot |
| Gout | 274.1 | ICD10CM | M1A | Chronic gout |
| Gout | 274.1 | ICD10CM | M1A.2311 | Drug-induced chronic gout, right wrist, with tophus (tophi) |
| Gout | 274.1 | ICD10CM | M1A.371 | Chronic gout due to renal impairment, right ankle and foot |
| Gout | 274.1 | ICD10CM | M10.02 | Idiopathic gout, elbow |
| Gout | 274.1 | ICD10CM | M1A.4221 | Other secondary chronic gout, left elbow, with tophus (tophi) |
| Gout | 274.1 | ICD10CM | M10.059 | Idiopathic gout, unspecified hip |
| Gout | 274.1 | ICD10CM | M1A.29X1 | Drug-induced chronic gout, multiple sites, with tophus (tophi) |

| | | | | |
|---|---|---|---|---|
| Gout | 274.1 | ICD10CM | M10.162 | Lead-induced gout, left knee |
| Gout | 274.1 | ICD10CM | M1A.0490 | Idiopathic chronic gout, unspecified hand, without tophus (tophi) |
| Gout | 274.1 | ICD10CM | M10.332 | Gout due to renal impairment, left wrist |
| Gout | 274.1 | ICD10CM | M1A.44 | Other secondary chronic gout, hand |
| Gout | 274.1 | ICD10CM | M1A.2290 | Drug-induced chronic gout, unspecified elbow, without tophus (tophi) |
| Gout | 274.1 | ICD10CM | M10.13 | Lead-induced gout, wrist |
| Gout | 274.1 | ICD10CM | M1A.059 | Idiopathic chronic gout, unspecified hip |
| Gout | 274.1 | ICD10CM | M1A.442 | Other secondary chronic gout, left hand |
| Gout | 274.1 | ICD10CM | M1A.22 | Drug-induced chronic gout, elbow |
| Gout | 274.1 | ICD10CM | M1A.4411 | Other secondary chronic gout, right hand, with tophus (tophi) |
| Gout | 274.1 | ICD10CM | M1A.252 | Drug-induced chronic gout, left hip |
| Gout | 274.1 | ICD10CM | M10.17 | Lead-induced gout, ankle and foot |
| Gout | 274.1 | ICD10CM | M1A.2421 | Drug-induced chronic gout, left hand, with tophus (tophi) |
| Gout | 274.1 | ICD10CM | M1A.0220 | Idiopathic chronic gout, left elbow, without tophus (tophi) |
| Gout | 274.1 | ICD10CM | M10.06 | Idiopathic gout, knee |
| Gout | 274.1 | ICD10CM | M1A.3711 | Chronic gout due to renal impairment, right ankle and foot, with tophus (tophi) |
| Gout | 274.1 | ICD10CM | M10.079 | Idiopathic gout, unspecified ankle and foot |
| Gout | 274.1 | ICD10CM | M10.139 | Lead-induced gout, unspecified wrist |
| Gout | 274.1 | ICD10CM | M1A.2410 | Drug-induced chronic gout, right hand, without tophus (tophi) |
| Gout | 274.1 | ICD10CM | M1A.4790 | Other secondary chronic gout, unspecified ankle and foot, without tophus (tophi) |
| Gout | 274.1 | ICD10CM | M10.171 | Lead-induced gout, right ankle and foot |
| Gout | 274.1 | ICD10CM | M1A.48 | Other secondary chronic gout, vertebrae |
| Gout | 274.1 | ICD10CM | M1A.3 | Chronic gout due to renal impairment |
| Gout | 274.1 | ICD10CM | M1A.049 | Idiopathic chronic gout, unspecified hand |
| Gout | 274.1 | ICD10CM | M1A.311 | Chronic gout due to renal impairment, right shoulder |
| Gout | 274.1 | ICD10CM | M1A.011 | Idiopathic chronic gout, right shoulder |
| Gout | 274.1 | ICD10CM | M10.172 | Lead-induced gout, left ankle and foot |
| Gout | 274.1 | ICD10CM | M1A.31 | Chronic gout due to renal impairment, shoulder |
| Gout | 274.1 | ICD10CM | M10.062 | Idiopathic gout, left knee |
| Gout | 274.1 | ICD10CM | M10.369 | Gout due to renal impairment, unspecified knee |
| Gout | 274.1 | ICD10CM | M1A.2420 | Drug-induced chronic gout, left hand, without tophus (tophi) |
| Gout | 274.1 | ICD10CM | M1A.4210 | Other secondary chronic gout, right elbow, without tophus (tophi) |
| Gout | 274.1 | ICD10CM | M10.34 | Gout due to renal impairment, hand |
| Gout | 274.1 | ICD9CM | 274.01 | Acute gouty arthropathy |

| | | | | |
|---|---|---|---|---|
| Gout | 274.1 | ICD10CM | M1A.2610 | Drug-induced chronic gout, right knee, without tophus (tophi) |
| Gout | 274.1 | ICD10CM | M10.322 | Gout due to renal impairment, left elbow |
| Gout | 274.1 | ICD10CM | M1A.3321 | Chronic gout due to renal impairment, left wrist, with tophus (tophi) |
| Gout | 274.1 | ICD10CM | M10.029 | Idiopathic gout, unspecified elbow |
| Gout | 274.1 | ICD10CM | M1A.24 | Drug-induced chronic gout, hand |
| Gout | 274.1 | ICD10CM | M10.011 | Idiopathic gout, right shoulder |
| Gout | 274.1 | ICD10CM | M10.061 | Idiopathic gout, right knee |
| Gout | 274.1 | ICD10CM | M10.351 | Gout due to renal impairment, right hip |
| Gout | 274.1 | ICD9CM | 274 | Gouty arthropathy |
| Gout | 274.1 | ICD9CM | 274.02 | Chronic gouty arthropathy without mention of tophus (tophi) |
| Gout | 274.1 | ICD10CM | M1A.2491 | Drug-induced chronic gout, unspecified hand, with tophus (tophi) |
| Gout | 274.1 | ICD10CM | M1A.2720 | Drug-induced chronic gout, left ankle and foot, without tophus (tophi) |
| Gout | 274.1 | ICD10CM | M1A.271 | Drug-induced chronic gout, right ankle and foot |
| Gout | 274.1 | ICD10CM | M1A.242 | Drug-induced chronic gout, left hand |
| Gout | 274.1 | ICD10CM | M10.279 | Drug-induced gout, unspecified ankle and foot |
| Gout | 274.1 | ICD10CM | M1A.2511 | Drug-induced chronic gout, right hip, with tophus (tophi) |
| Gout | 274.1 | ICD10CM | M10.359 | Gout due to renal impairment, unspecified hip |
| Gout | 274.1 | ICD10CM | M10.311 | Gout due to renal impairment, right shoulder |
| Gout | 274.1 | ICD10CM | M1A.0790 | Idiopathic chronic gout, unspecified ankle and foot, without tophus (tophi) |
| Gout | 274.1 | ICD10CM | M1A.0791 | Idiopathic chronic gout, unspecified ankle and foot, with tophus (tophi) |
| Gout | 274.1 | ICD10CM | M1A.319 | Chronic gout due to renal impairment, unspecified shoulder |
| Gout | 274.1 | ICD10CM | M1A.0520 | Idiopathic chronic gout, left hip, without tophus (tophi) |
| Gout | 274.1 | ICD10CM | M1A.39 | Chronic gout due to renal impairment, multiple sites |
| Obesity | 278.1 | ICD10CM | Z68.34 | Body mass index [BMI] 34.0-34.9, adult |
| Obesity | 278.1 | ICD10CM | E66.0 | Obesity due to excess calories |
| Obesity | 278.1 | ICD10CM | E66.1 | Drug-induced obesity |
| Obesity | 278.1 | ICD10CM | Z68.33 | Body mass index [BMI] 33.0-33.9, adult |
| Obesity | 278.1 | ICD9CM | V85.30 | Body Mass Index 30.0-30.9, adult |
| Obesity | 278.1 | ICD10CM | E66.8 | Other obesity |
| Obesity | 278.1 | ICD9CM | V85.34 | Body Mass Index 34.0-34.9, adult |
| Obesity | 278.1 | ICD10CM | E66.09 | Other obesity due to excess calories |
| Obesity | 278.1 | ICD9CM | V85.3 | Body Mass Index between 30-39, adult |
| Obesity | 278.1 | ICD9CM | V85.33 | Body Mass Index 33.0-33.9, adult |

| | | | | | |
|---|---|---|---|---|---|
| | Obesity | 278.1 | ICD10CM | R93.9 | Diagnostic imaging inconclusive due to excess body fat of patient |
| | Obesity | 278.1 | ICD10CM | E66.9 | Obesity, unspecified |
| | Obesity | 278.1 | ICD9CM | 278 | Obesity, unspecified |
| | Obesity | 278.1 | ICD9CM | V85.32 | Body Mass Index 32.0-32.9, adult |
| | Obesity | 278.1 | ICD10CM | Z68.32 | Body mass index [BMI] 32.0-32.9, adult |
| | Obesity | 278.1 | ICD9CM | V85.41 | Body Mass Index 40.0-44.9, adult |
| | Obesity | 278.1 | ICD10CM | Z68.3 | Body mass index [BMI] 30-39, adult |
| | Obesity | 278.1 | ICD9CM | V85.31 | Body Mass Index 31.0-31.9, adult |
| | Obesity | 278.1 | ICD9CM | 278.01 | Morbid obesity |
| | Obesity | 278.1 | ICD10CM | Z68.42 | Body mass index [BMI] 45.0-49.9, adult |
| | Obesity | 278.1 | ICD9CM | V85.43 | Body Mass Index 50.0-59.9, adult |
| | Obesity | 278.1 | ICD9CM | V85.35 | Body Mass Index 35.0-35.9, adult |
| | Obesity | 278.1 | ICD9CM | 793.91 | Image test inconclusive due to excess body fat |
| | Obesity | 278.1 | ICD10CM | Z68.30 | Body mass index [BMI] 30.0-30.9, adult |
| | Obesity | 278.1 | ICD9CM | V85.54 | Body Mass Index, pediatric, greater than or equal to 95th percentile for age |
| | Obesity | 278.1 | ICD9CM | V85.4 | Body Mass Index 40 and over, adult |
| | Obesity | 278.1 | ICD10CM | Z68.43 | Body mass index [BMI] 50.0-59.9, adult |
| | Obesity | 278.1 | ICD10CM | Z68.54 | Body mass index [BMI] pediatric, greater than or equal to 95th percentile for age |
| | Obesity | 278.1 | ICD10CM | Z68.31 | Body mass index [BMI] 31.0-31.9, adult |
| | Obesity | 278.1 | ICD9CM | V85.39 | Body Mass Index 39.0-39.9, adult |
| | Obesity | 278.1 | ICD9CM | V85.38 | Body Mass Index 38.0-38.9, adult |
| | Obesity | 278.1 | ICD10CM | Z68.44 | Body mass index [BMI] 60.0-69.9, adult |
| | Obesity | 278.1 | ICD10CM | Z68.37 | Body mass index [BMI] 37.0-37.9, adult |
| | Obesity | 278.1 | ICD10CM | Z68.36 | Body mass index [BMI] 36.0-36.9, adult |
| | Obesity | 278.1 | ICD10CM | Z68.35 | Body mass index [BMI] 35.0-35.9, adult |
| | Obesity | 278.1 | ICD10CM | Z68.39 | Body mass index [BMI] 39.0-39.9, adult |
| | Obesity | 278.1 | ICD9CM | V85.36 | Body Mass Index 36.0-36.9, adult |
| | Obesity | 278.1 | ICD9CM | V85.42 | Body Mass Index 45.0-49.9, adult |
| | Obesity | 278.1 | ICD10CM | Z68.4 | Body mass index [BMI] 40 or greater, adult |
| | Obesity | 278.1 | ICD10CM | Z68.41 | Body mass index [BMI] 40.0-44.9, adult |
| | Obesity | 278.1 | ICD10CM | Z68.45 | Body mass index [BMI] 70 or greater, adult |
| | Obesity | 278.1 | ICD9CM | V85.45 | Body Mass Index 70 and over, adult |
| | Obesity | 278.1 | ICD9CM | V85.37 | Body Mass Index 37.0-37.9, adult |
| | Obesity | 278.1 | ICD10CM | Z68.38 | Body mass index [BMI] 38.0-38.9, adult |
| | Obesity | 278.1 | ICD9CM | V85.44 | Body Mass Index 60.0-69.9, adult |
| | Obesity | 278.1 | ICD10CM | E66.01 | Morbid (severe) obesity due to excess calories |
| **Mental Disorders** | Alzheimer's disease | 290.11 | ICD10CM | G30.8 | Other Alzheimer's disease |
| | Alzheimer's disease | 290.11 | ICD10CM | G30.0 | Alzheimer's disease with early onset |

| | | | | |
|---|---|---|---|---|
| Alzheimer's disease | 290.11 | ICD10CM | G30.9 | Alzheimer's disease, unspecified |
| Alzheimer's disease | 290.11 | ICD10CM | G30 | Alzheimer's disease |
| Alzheimer's disease | 290.11 | ICD9CM | 331 | Alzheimer's disease * |
| Alzheimer's disease | 290.11 | ICD9CM | 331 | Alzheimer's disease |
| Alzheimer's disease | 290.11 | ICD10CM | G30.1 | Alzheimer's disease with late onset |
| Schizophrenia | 295.1 | ICD9CM | 295.52 | Latent schizophrenia, chronic |
| Schizophrenia | 295.1 | ICD10CM | F20.89 | Other schizophrenia |
| Schizophrenia | 295.1 | ICD9CM | 295.71 | Schizoaffective disorder, subchronic |
| Schizophrenia | 295.1 | ICD10CM | F20 | Schizophrenia |
| Schizophrenia | 295.1 | ICD9CM | 295.05 | Simple type schizophrenia, in remission |
| Schizophrenia | 295.1 | ICD9CM | 295.92 | Unspecified schizophrenia, chronic |
| Schizophrenia | 295.1 | ICD9CM | 295.33 | Paranoid type schizophrenia, subchronic with acute exacerbation |
| Schizophrenia | 295.1 | ICD9CM | 295.63 | Schizophrenic disorders, residual type, subchronic with acute exacerbation |
| Schizophrenia | 295.1 | ICD9CM | 295.23 | Catatonic type schizophrenia, subchronic with acute exacerbation |
| Schizophrenia | 295.1 | ICD9CM | 295.9 | Unspecified schizophrenia |
| Schizophrenia | 295.1 | ICD9CM | 295.15 | Disorganized type schizophrenia, in remission |
| Schizophrenia | 295.1 | ICD9CM | 295.72 | Schizoaffective disorder, chronic |
| Schizophrenia | 295.1 | ICD9CM | 295.51 | Latent schizophrenia, subchronic |
| Schizophrenia | 295.1 | ICD9CM | 295.7 | Schizoaffective disorder |
| Schizophrenia | 295.1 | ICD10CM | F20.3 | Undifferentiated schizophrenia |
| Schizophrenia | 295.1 | ICD9CM | 295.64 | Schizophrenic disorders, residual type, chronic with acute exacerbation |
| Schizophrenia | 295.1 | ICD9CM | 295.75 | Schizoaffective disorder, in remission |
| Schizophrenia | 295.1 | ICD10CM | F20.9 | Schizophrenia, unspecified |
| Schizophrenia | 295.1 | ICD9CM | 295.91 | Unspecified schizophrenia, subchronic |
| Schizophrenia | 295.1 | ICD9CM | 295.54 | Latent schizophrenia, chronic with acute exacerbation |
| Schizophrenia | 295.1 | ICD9CM | 295.25 | Catatonic type schizophrenia, in remission |
| Schizophrenia | 295.1 | ICD9CM | 295.62 | Schizophrenic disorders, residual type, chronic |
| Schizophrenia | 295.1 | ICD9CM | 295.3 | Paranoid type schizophrenia, unspecified |
| Schizophrenia | 295.1 | ICD10CM | F20.81 | Schizophreniform disorder |
| Schizophrenia | 295.1 | ICD9CM | 295.5 | Latent schizophrenia, unspecified |
| Schizophrenia | 295.1 | ICD10CM | F25.1 | Schizoaffective disorder, depressive type |
| Schizophrenia | 295.1 | ICD10CM | F25.8 | Other schizoaffective disorders |
| Schizophrenia | 295.1 | ICD9CM | 295.6 | Residual type schizophrenic disorders |
| Schizophrenia | 295.1 | ICD9CM | 295.81 | Other specified types of schizophrenia, subchronic |
| Schizophrenia | 295.1 | ICD9CM | 295.8 | Other specified types of schizophrenia |
| Schizophrenia | 295.1 | ICD9CM | 295.14 | Disorganized type schizophrenia, chronic with acute exacerbation |

| | | | | |
|---|---|---|---|---|
| Schizophrenia | 295.1 | ICD10CM | F25.9 | Schizoaffective disorder, unspecified |
| Schizophrenia | 295.1 | ICD9CM | 295.6 | Schizophrenic disorders, residual type, unspecified |
| Schizophrenia | 295.1 | ICD9CM | 295.45 | Schizophreniform disorder, in remission |
| Schizophrenia | 295.1 | ICD9CM | 295.82 | Other specified types of schizophrenia, chronic |
| Schizophrenia | 295.1 | ICD9CM | 295.22 | Catatonic type schizophrenia, chronic |
| Schizophrenia | 295.1 | ICD9CM | 295.11 | Disorganized type schizophrenia, subchronic |
| Schizophrenia | 295.1 | ICD9CM | 295.41 | Schizophreniform disorder, subchronic |
| Schizophrenia | 295.1 | ICD9CM | 295.13 | Disorganized type schizophrenia, subchronic with acute exacerbation |
| Schizophrenia | 295.1 | ICD9CM | 295.9 | Unspecified schizophrenia, unspecified |
| Schizophrenia | 295.1 | ICD9CM | 295.5 | Latent schizophrenia |
| Schizophrenia | 295.1 | ICD9CM | 295.01 | Simple type schizophrenia, subchronic |
| Schizophrenia | 295.1 | ICD9CM | 295.2 | Catatonic type schizophrenia, unspecified |
| Schizophrenia | 295.1 | ICD9CM | 295.4 | Schizophreniform disorder, unspecified |
| Schizophrenia | 295.1 | ICD9CM | 295.12 | Disorganized type schizophrenia, chronic |
| Schizophrenia | 295.1 | ICD9CM | 295.35 | Paranoid type schizophrenia, in remission |
| Schizophrenia | 295.1 | ICD9CM | 295.53 | Latent schizophrenia, subchronic with acute exacerbation |
| Schizophrenia | 295.1 | ICD9CM | 295.44 | Schizophreniform disorder, chronic with acute exacerbation |
| Schizophrenia | 295.1 | ICD9CM | V11.0 | Personal history of schizophrenia |
| Schizophrenia | 295.1 | ICD9CM | 295.85 | Other specified types of schizophrenia, in remission |
| Schizophrenia | 295.1 | ICD10CM | F25.0 | Schizoaffective disorder, bipolar type |
| Schizophrenia | 295.1 | ICD9CM | 295.42 | Schizophreniform disorder, chronic |
| Schizophrenia | 295.1 | ICD10CM | F20.5 | Residual schizophrenia |
| Schizophrenia | 295.1 | ICD9CM | 295.93 | Unspecified schizophrenia, subchronic with acute exacerbation |
| Schizophrenia | 295.1 | ICD9CM | 295.03 | Simple type schizophrenia, subchronic with acute exacerbation |
| Schizophrenia | 295.1 | ICD9CM | 295.32 | Paranoid type schizophrenia, chronic |
| Schizophrenia | 295.1 | ICD9CM | 295.94 | Unspecified schizophrenia, chronic with acute exacerbation |
| Schizophrenia | 295.1 | ICD9CM | 295.31 | Paranoid type schizophrenia, subchronic |
| Schizophrenia | 295.1 | ICD9CM | 295.84 | Other specified types of schizophrenia, chronic with acute exacerbation |
| Schizophrenia | 295.1 | ICD9CM | 295.83 | Other specified types of schizophrenia, subchronic with acute exacerbation |
| Schizophrenia | 295.1 | ICD10CM | F20.8 | Other schizophrenia |
| Schizophrenia | 295.1 | ICD9CM | 295.1 | Disorganized type schizophrenia, unspecified |
| Schizophrenia | 295.1 | ICD9CM | 295.34 | Paranoid type schizophrenia, chronic with acute exacerbation |
| Schizophrenia | 295.1 | ICD9CM | 295.74 | Schizoaffective disorder, chronic with acute exacerbation |

| | | | | |
|---|---|---|---|---|
| Schizophrenia | 295.1 | ICD9CM | 295.3 | Paranoid type schizophrenia |
| Schizophrenia | 295.1 | ICD9CM | 295.73 | Schizoaffective disorder, subchronic with acute exacerbation |
| Schizophrenia | 295.1 | ICD9CM | 295.2 | Catatonic type schizophrenia |
| Schizophrenia | 295.1 | ICD10CM | F25 | Schizoaffective disorders |
| Schizophrenia | 295.1 | ICD9CM | 295.8 | Other specified types of schizophrenia, unspecified |
| Schizophrenia | 295.1 | ICD10CM | F20.0 | Paranoid schizophrenia |
| Schizophrenia | 295.1 | ICD9CM | 295.21 | Catatonic type schizophrenia, subchronic |
| Schizophrenia | 295.1 | ICD9CM | 295.95 | Unspecified schizophrenia, in remission |
| Schizophrenia | 295.1 | ICD10CM | F20.2 | Catatonic schizophrenia |
| Schizophrenia | 295.1 | ICD9CM | 295.02 | Simple type schizophrenia, chronic |
| Schizophrenia | 295.1 | ICD10CM | F20.1 | Disorganized schizophrenia |
| Schizophrenia | 295.1 | ICD9CM | 295.61 | Schizophrenic disorders, residual type, subchronic |
| Schizophrenia | 295.1 | ICD9CM | 295.65 | Schizophrenic disorders, residual type, in remission |
| Schizophrenia | 295.1 | ICD9CM | 295.7 | Schizoaffective disorder, unspecified |
| Schizophrenia | 295.1 | ICD9CM | 295.04 | Simple type schizophrenia, chronic with acute exacerbation |
| Schizophrenia | 295.1 | ICD9CM | 295.55 | Latent schizophrenia, in remission |
| Schizophrenia | 295.1 | ICD9CM | 295 | Simple type schizophrenia |
| Schizophrenia | 295.1 | ICD9CM | 295.24 | Catatonic type schizophrenia, chronic with acute exacerbation |
| Schizophrenia | 295.1 | ICD9CM | 295.43 | Schizophreniform disorder, subchronic with acute exacerbation |
| Schizophrenia | 295.1 | ICD9CM | 295.1 | Disorganized type schizophrenia |
| Schizophrenia | 295.1 | ICD9CM | 295 | Simple type schizophrenia, unspecified |
| Depression | 296.2 | ICD9CM | 296.21 | Major depressive affective disorder, single episode, mild |
| Depression | 296.2 | ICD9CM | 311 | Depressive disorder, not elsewhere classified |
| Depression | 296.2 | ICD9CM | 296.31 | Major depressive affective disorder, recurrent episode, mild |
| Depression | 296.2 | ICD9CM | 296.26 | Major depressive affective disorder, single episode, in full remission |
| Depression | 296.2 | ICD10CM | F33.42 | Major depressive disorder, recurrent, in full remission |
| Depression | 296.2 | ICD9CM | 296.34 | Major depressive affective disorder, recurrent episode, severe, specified as with psychotic behavior |
| Depression | 296.2 | ICD9CM | 296.25 | Major depressive affective disorder, single episode, in partial or unspecified remission |
| Depression | 296.2 | ICD10CM | F32.8 | Other depressive episodes |
| Depression | 296.2 | ICD9CM | 296.22 | Major depressive affective disorder, single episode, moderate |
| Depression | 296.2 | ICD10CM | F32.0 | Major depressive disorder, single episode, mild |

| | | | | |
|---|---|---|---|---|
| Depression | 296.2 | ICD10CM | F32.89 | Other specified depressive episodes |
| Depression | 296.2 | ICD9CM | 296.23 | Major depressive affective disorder, single episode, severe, without mention of psychotic behavior |
| Depression | 296.2 | ICD9CM | 296.32 | Major depressive affective disorder, recurrent episode, moderate |
| Depression | 296.2 | ICD10CM | F33.41 | Major depressive disorder, recurrent, in partial remission |
| Depression | 296.2 | ICD10CM | F32 | Depressive episode |
| Depression | 296.2 | ICD9CM | 296.35 | Major depressive affective disorder, recurrent episode, in partial or unspecified remission |
| Depression | 296.2 | ICD10CM | F32.2 | Major depressive disorder, single episode, severe without psychotic features |
| Depression | 296.2 | ICD10CM | F33.3 | Major depressive disorder, recurrent, severe with psychotic symptoms |
| Depression | 296.2 | ICD10CM | F33.40 | Major depressive disorder, recurrent, in remission, unspecified |
| Depression | 296.2 | ICD9CM | 296.2 | Major depressive disorder, single episode |
| Depression | 296.2 | ICD9CM | 296.24 | Major depressive affective disorder, single episode, severe, specified as with psychotic behavior |
| Depression | 296.2 | ICD10CM | F33 | Major depressive disorder, recurrent |
| Depression | 296.2 | ICD10CM | F32.1 | Major depressive disorder, single episode, moderate |
| Depression | 296.2 | ICD9CM | 296.3 | Major depressive disorder, recurrent episode |
| Depression | 296.2 | ICD10CM | F32.9 | Major depressive disorder, single episode, unspecified |
| Depression | 296.2 | ICD10CM | F32.81 | Premenstrual dysphoric disorder |
| Depression | 296.2 | ICD9CM | 296.2 | Major depressive affective disorder, single episode, unspecified |
| Depression | 296.2 | ICD10CM | F33.0 | Major depressive disorder, recurrent, mild |
| Depression | 296.2 | ICD10CM | F33.4 | Major depressive disorder, recurrent, in remission |
| Depression | 296.2 | ICD10CM | F32.5 | Major depressive disorder, single episode, in full remission |
| Depression | 296.2 | ICD10CM | F33.2 | Major depressive disorder, recurrent severe without psychotic features |
| Depression | 296.2 | ICD10CM | F33.8 | Other recurrent depressive disorders |
| Depression | 296.2 | ICD9CM | 296.33 | Major depressive affective disorder, recurrent episode, severe, without mention of psychotic behavior |
| Depression | 296.2 | ICD9CM | 296.3 | Major depressive affective disorder, recurrent episode, unspecified |
| Depression | 296.2 | ICD10CM | F33.1 | Major depressive disorder, recurrent, moderate |
| Depression | 296.2 | ICD10CM | F32.A | Depression, unspecified |
| Depression | 296.2 | ICD10CM | F33.9 | Major depressive disorder, recurrent, unspecified |

| | | | | |
|---|---|---|---|---|
| Depression | 296.2 | ICD10CM | F32.3 | Major depressive disorder, single episode, severe with psychotic features |
| Depression | 296.2 | ICD9CM | 296.36 | Major depressive affective disorder, recurrent episode, in full remission |
| Depression | 296.2 | ICD10CM | F32.4 | Major depressive disorder, single episode, in partial remission |
| Generalized anxiety disorder | 300.11 | ICD10CM | F41.1 | Generalized anxiety disorder |
| Generalized anxiety disorder | 300.11 | ICD9CM | 300.02 | Generalized anxiety disorder |
| Alcoholism | 317.1 | ICD9CM | 535.31 | Alcoholic gastritis, with hemorrhage |
| Alcoholism | 317.1 | ICD9CM | 291.81 | Alcohol withdrawal |
| Alcoholism | 317.1 | ICD9CM | 357.5 | Alcoholic polyneuropathy |
| Alcoholism | 317.1 | ICD10CM | F10.230 | Alcohol dependence with withdrawal, uncomplicated |
| Alcoholism | 317.1 | ICD9CM | 305 | Alcohol abuse |
| Alcoholism | 317.1 | ICD10CM | F10.151 | Alcohol abuse with alcohol-induced psychotic disorder with hallucinations |
| Alcoholism | 317.1 | ICD9CM | 535.3 | Alcoholic gastritis |
| Alcoholism | 317.1 | ICD10CM | F10.120 | Alcohol abuse with intoxication, uncomplicated |
| Alcoholism | 317.1 | ICD10CM | F10.232 | Alcohol dependence with withdrawal with perceptual disturbance |
| Alcoholism | 317.1 | ICD9CM | 305.03 | Alcohol abuse, in remission |
| Alcoholism | 317.1 | ICD10CM | F10.251 | Alcohol dependence with alcohol-induced psychotic disorder with hallucinations |
| Alcoholism | 317.1 | ICD10CM | F10.10 | Alcohol abuse, uncomplicated |
| Alcoholism | 317.1 | ICD10CM | F10.27 | Alcohol dependence with alcohol-induced persisting dementia |
| Alcoholism | 317.1 | ICD9CM | 291.1 | Alcohol-induced persisting amnestic disorder |
| Alcoholism | 317.1 | ICD9CM | 305.01 | Alcohol abuse, continuous |
| Alcoholism | 317.1 | ICD10CM | F10.1 | Alcohol abuse |
| Alcoholism | 317.1 | ICD9CM | V11.3 | Personal history of alcoholism |
| Alcoholism | 317.1 | ICD10CM | F10.26 | Alcohol dependence with alcohol-induced persisting amnestic disorder |
| Alcoholism | 317.1 | ICD9CM | 291.3 | Alcohol-induced psychotic disorder with hallucinations |
| Alcoholism | 317.1 | ICD10CM | F10.96 | Alcohol use, unspecified with alcohol-induced persisting amnestic disorder |
| Alcoholism | 317.1 | ICD10CM | K29.20 | Alcoholic gastritis without bleeding |
| Alcoholism | 317.1 | ICD9CM | 305.02 | Alcohol abuse, episodic |
| Alcoholism | 317.1 | ICD10CM | F10.132 | Alcohol abuse with withdrawal with perceptual disturbance |
| Alcoholism | 317.1 | ICD10CM | G62.1 | Alcoholic polyneuropathy |
| Alcoholism | 317.1 | ICD10CM | K29.21 | Alcoholic gastritis with bleeding |
| Alcoholism | 317.1 | ICD9CM | 305 | Alcohol abuse, unspecified |
| Alcoholism | 317.1 | ICD10CM | F10.139 | Alcohol abuse with withdrawal, unspecified |

| | | | | | |
|---|---|---|---|---|---|
| | Alcoholism | 317.1 | ICD10CM | F10.130 | Alcohol abuse with withdrawal, uncomplicated |
| | Alcoholism | 317.1 | ICD10CM | F10.131 | Alcohol abuse with withdrawal delirium |
| | Alcoholism | 317.1 | ICD10CM | F10.97 | Alcohol use, unspecified with alcohol-induced persisting dementia |
| | Alcoholism | 317.1 | ICD10CM | F10.129 | Alcohol abuse with intoxication, unspecified |
| | Alcoholism | 317.1 | ICD10CM | K29.2 | Alcoholic gastritis |
| | Alcoholism | 317.1 | ICD10CM | F10.951 | Alcohol use, unspecified with alcohol-induced psychotic disorder with hallucinations |
| | Alcoholism | 317.1 | ICD9CM | 535.3 | Alcoholic gastritis, without mention of hemorrhage |
| | Alcoholism | 317.1 | ICD9CM | 291.2 | Alcohol-induced persisting dementia |
| | Alcoholism | 317.1 | ICD10CM | F10.239 | Alcohol dependence with withdrawal, unspecified |
| | Alcoholism | 317.1 | ICD10CM | K70.40 | Alcoholic hepatic failure without coma |
| | Alcoholism | 317.1 | ICD10CM | K70 | Alcoholic liver disease |
| | Alcoholism | 317.1 | ICD10CM | K70.11 | Alcoholic hepatitis with ascites |
| | Alcoholism | 317.1 | ICD10CM | K70.3 | Alcoholic cirrhosis of liver |
| | Alcoholism | 317.1 | ICD9CM | 571 | Alcoholic fatty liver |
| | Alcoholism | 317.1 | ICD10CM | K70.30 | Alcoholic cirrhosis of liver without ascites |
| | Alcoholism | 317.1 | ICD9CM | 571.1 | Acute alcoholic hepatitis |
| | Alcoholism | 317.1 | ICD10CM | K70.31 | Alcoholic cirrhosis of liver with ascites |
| | Alcoholism | 317.1 | ICD9CM | 571.2 | Alcoholic cirrhosis of liver |
| | Alcoholism | 317.1 | ICD10CM | K70.9 | Alcoholic liver disease, unspecified |
| | Alcoholism | 317.1 | ICD9CM | 571.3 | Alcoholic liver damage, unspecified |
| | Alcoholism | 317.1 | ICD10CM | K70.1 | Alcoholic hepatitis |
| | Alcoholism | 317.1 | ICD10CM | K70.41 | Alcoholic hepatic failure with coma |
| | Alcoholism | 317.1 | ICD10CM | K70.0 | Alcoholic fatty liver |
| | Alcoholism | 317.1 | ICD10CM | K70.10 | Alcoholic hepatitis without ascites |
| | Alcoholism | 317.1 | ICD10CM | K70.2 | Alcoholic fibrosis and sclerosis of liver |
| **Neurological** | Migraine | 340 | ICD10CM | G43.0 | Migraine without aura |
| | Migraine | 340 | ICD9CM | 346.21 | Variants of migraine, not elsewhere classified, with intractable migraine, so stated, without mention of status migrainosus |
| | Migraine | 340 | ICD10CM | G43.411 | Hemiplegic migraine, intractable, with status migrainosus |
| | Migraine | 340 | ICD9CM | 346.3 | Hemiplegic migraine, without mention of intractable migraine without mention of status migrainosus |
| | Migraine | 340 | ICD9CM | 346.43 | Menstrual migraine, with intractable migraine, so stated, with status migrainosus |
| | Migraine | 340 | ICD9CM | 339.03 | Episodic paroxysmal hemicrania |
| | Migraine | 340 | ICD10CM | G43.819 | Other migraine, intractable, without status migrainosus |
| | Migraine | 340 | ICD10CM | G43.82 | Menstrual migraine, not intractable |

| | | | | |
|---|---|---|---|---|
| Migraine | 340 | ICD10CM | G43.829 | Menstrual migraine, not intractable, without status migrainosus |
| Migraine | 340 | ICD10CM | G43.B0 | Ophthalmoplegic migraine, not intractable |
| Migraine | 340 | ICD9CM | 346.9 | Migraine, unspecified |
| Migraine | 340 | ICD9CM | 346.92 | Migraine, unspecified, without mention of intractable migraine with status migrainosus |
| Migraine | 340 | ICD9CM | 339.01 | Episodic cluster headache |
| Migraine | 340 | ICD10CM | G43.C | Periodic headache syndromes in child or adult |
| Migraine | 340 | ICD9CM | 346.22 | Variants of migraine, not elsewhere classified, without mention of intractable migraine with status migrainosus |
| Migraine | 340 | ICD10CM | G44.021 | Chronic cluster headache, intractable |
| Migraine | 340 | ICD10CM | G44.03 | Episodic paroxysmal hemicrania |
| Migraine | 340 | ICD10CM | G43.401 | Hemiplegic migraine, not intractable, with status migrainosus |
| Migraine | 340 | ICD10CM | G44.031 | Episodic paroxysmal hemicrania, intractable |
| Migraine | 340 | ICD9CM | 346.83 | Other forms of migraine, with intractable migraine, so stated, with status migrainosus |
| Migraine | 340 | ICD9CM | 346.93 | Migraine, unspecified, with intractable migraine, so stated, with status migrainosus |
| Migraine | 340 | ICD9CM | 339.02 | Chronic cluster headache |
| Migraine | 340 | ICD10CM | G43.501 | Persistent migraine aura without cerebral infarction, not intractable, with status migrainosus |
| Migraine | 340 | ICD10CM | G43.41 | Hemiplegic migraine, intractable |
| Migraine | 340 | ICD10CM | G43 | Migraine |
| Migraine | 340 | ICD9CM | 346.1 | Migraine without aura, without mention of intractable migraine without mention of status migrainosus |
| Migraine | 340 | ICD10CM | G43.009 | Migraine without aura, not intractable, without status migrainosus |
| Migraine | 340 | ICD10CM | G44.02 | Chronic cluster headache |
| Migraine | 340 | ICD10CM | G43.70 | Chronic migraine without aura, not intractable |
| Migraine | 340 | ICD10CM | G43.E19 | Chronic migraine with aura, intractable, without status migrainosus |
| Migraine | 340 | ICD10CM | G43.839 | Menstrual migraine, intractable, without status migrainosus |
| Migraine | 340 | ICD10CM | G44.001 | Cluster headache syndrome, unspecified, intractable |
| Migraine | 340 | ICD10CM | G43.A1 | Cyclical vomiting, in migraine, intractable |
| Migraine | 340 | ICD10CM | G44.01 | Episodic cluster headache |
| Migraine | 340 | ICD10CM | G43.919 | Migraine, unspecified, intractable, without status migrainosus |
| Migraine | 340 | ICD10CM | G43.001 | Migraine without aura, not intractable, with status migrainosus |
| Migraine | 340 | ICD9CM | 346.2 | Variants of migraine, not elsewhere classified |

| | | | | |
|---|---|---|---|---|
| Migraine | 340 | ICD10CM | G43.801 | Other migraine, not intractable, with status migrainosus |
| Migraine | 340 | ICD9CM | 346.91 | Migraine, unspecified, with intractable migraine, so stated, without mention of status migrainosus |
| Migraine | 340 | ICD9CM | 339 | Cluster headache syndrome, unspecified |
| Migraine | 340 | ICD10CM | G43.E09 | Chronic migraine with aura, not intractable, without status migrainosus |
| Migraine | 340 | ICD10CM | G43.E11 | Chronic migraine with aura, intractable, with status migrainosus |
| Migraine | 340 | ICD10CM | G43.D0 | Abdominal migraine, not intractable |
| Migraine | 340 | ICD9CM | 346.82 | Other forms of migraine, without mention of intractable migraine with status migrainosus |
| Migraine | 340 | ICD9CM | 346.33 | Hemiplegic migraine, with intractable migraine, so stated, with status migrainosus |
| Migraine | 340 | ICD10CM | G44.049 | Chronic paroxysmal hemicrania, not intractable |
| Migraine | 340 | ICD9CM | 346.32 | Hemiplegic migraine, without mention of intractable migraine with status migrainosus |
| Migraine | 340 | ICD10CM | G43.83 | Menstrual migraine, intractable |
| Migraine | 340 | ICD9CM | 346.1 | Migraine without aura |
| Migraine | 340 | ICD10CM | G43.E01 | Chronic migraine with aura, not intractable, with status migrainosus |
| Migraine | 340 | ICD10CM | G43.701 | Chronic migraine without aura, not intractable, with status migrainosus |
| Migraine | 340 | ICD10CM | G43.7 | Chronic migraine without aura |
| Migraine | 340 | ICD10CM | G44.04 | Chronic paroxysmal hemicrania |
| Migraine | 340 | ICD9CM | 346.9 | Migraine, unspecified, without mention of intractable migraine without mention of status migrainosus |
| Migraine | 340 | ICD9CM | 346.23 | Variants of migraine, not elsewhere classified, with intractable migraine, so stated, with status migrainosus |
| Migraine | 340 | ICD10CM | G44.00 | Cluster headache syndrome, unspecified |
| Migraine | 340 | ICD9CM | 346.42 | Menstrual migraine, without mention of intractable migraine with status migrainosus |
| Migraine | 340 | ICD10CM | G43.419 | Hemiplegic migraine, intractable, without status migrainosus |
| Migraine | 340 | ICD10CM | G43.B | Ophthalmoplegic migraine |
| Migraine | 340 | ICD10CM | G43.011 | Migraine without aura, intractable, with status migrainosus |
| Migraine | 340 | ICD9CM | 346.13 | Migraine without aura, with intractable migraine, so stated, with status migrainosus |
| Migraine | 340 | ICD9CM | 346.2 | Variants of migraine, not elsewhere classified, without mention of intractable migraine without mention of status migrainosus |
| Migraine | 340 | ICD10CM | G43.809 | Other migraine, not intractable, without status migrainosus |
| Migraine | 340 | ICD10CM | G44.009 | Cluster headache syndrome, unspecified, not intractable |

| | | | | |
|---|---|---|---|---|
| Migraine | 340 | ICD10CM | G44.041 | Chronic paroxysmal hemicrania, intractable |
| Migraine | 340 | ICD9CM | 346.4 | Menstrual migraine, without mention of intractable migraine without mention of status migrainosus |
| Migraine | 340 | ICD9CM | 346.81 | Other forms of migraine, with intractable migraine, so stated, without mention of status migrainosus |
| Migraine | 340 | ICD9CM | 346.8 | Other forms of migraine, without mention of intractable migraine without mention of status migrainosus |
| Migraine | 340 | ICD9CM | 346.52 | Persistent migraine aura without cerebral infarction, without mention of intractable migraine with status migrainosus |
| Migraine | 340 | ICD10CM | G43.711 | Chronic migraine without aura, intractable, with status migrainosus |
| Migraine | 340 | ICD9CM | 346.8 | Other forms of migraine |
| Migraine | 340 | ICD9CM | 346.72 | Chronic migraine without aura, without mention of intractable migraine with status migrainosus |
| Migraine | 340 | ICD9CM | 346.7 | Chronic migraine without aura, without mention of intractable migraine without mention of status migrainosus |
| Migraine | 340 | ICD9CM | 346.73 | Chronic migraine without aura, with intractable migraine, so stated, with status migrainosus |
| Migraine | 340 | ICD10CM | G43.D | Abdominal migraine |
| Migraine | 340 | ICD10CM | G43.D1 | Abdominal migraine, intractable |
| Migraine | 340 | ICD10CM | G43.91 | Migraine, unspecified, intractable |
| Migraine | 340 | ICD10CM | G43.901 | Migraine, unspecified, not intractable, with status migrainosus |
| Migraine | 340 | ICD10CM | G43.8 | Other migraine |
| Migraine | 340 | ICD10CM | G43.831 | Menstrual migraine, intractable, with status migrainosus |
| Migraine | 340 | ICD10CM | G43.909 | Migraine, unspecified, not intractable, without status migrainosus |
| Migraine | 340 | ICD9CM | 346.3 | Hemiplegic migraine |
| Migraine | 340 | ICD9CM | 346.71 | Chronic migraine without aura, with intractable migraine, so stated, without mention of status migrainosus |
| Migraine | 340 | ICD10CM | G43.9 | Migraine, unspecified |
| Migraine | 340 | ICD9CM | 346.31 | Hemiplegic migraine, with intractable migraine, so stated, without mention of status migrainosus |
| Migraine | 340 | ICD9CM | 339 | Cluster headaches and other trigeminal autonomic cephalgias |
| Migraine | 340 | ICD9CM | 346 | Migraine |
| Migraine | 340 | ICD10CM | G43.90 | Migraine, unspecified, not intractable |
| Migraine | 340 | ICD10CM | G43.4 | Hemiplegic migraine |
| Migraine | 340 | ICD10CM | G43.C1 | Periodic headache syndromes in child or adult, intractable |

| | | | | |
|---|---|---|---|---|
| Migraine | 340 | ICD10CM | G43.719 | Chronic migraine without aura, intractable, without status migrainosus |
| Migraine | 340 | ICD10CM | G43.811 | Other migraine, intractable, with status migrainosus |
| Migraine | 340 | ICD10CM | G43.911 | Migraine, unspecified, intractable, with status migrainosus |
| Migraine | 340 | ICD10CM | G43.409 | Hemiplegic migraine, not intractable, without status migrainosus |
| Migraine | 340 | ICD9CM | 339.04 | Chronic paroxysmal hemicrania |
| Migraine | 340 | ICD9CM | 346.41 | Menstrual migraine, with intractable migraine, so stated, without mention of status migrainosus |
| Migraine | 340 | ICD9CM | 346.11 | Migraine without aura, with intractable migraine, so stated, without mention of status migrainosus |
| Migraine | 340 | ICD10CM | G43.019 | Migraine without aura, intractable, without status migrainosus |
| Migraine | 340 | ICD10CM | G44.011 | Episodic cluster headache, intractable |
| Migraine | 340 | ICD10CM | G44.019 | Episodic cluster headache, not intractable |
| Migraine | 340 | ICD10CM | G43.A0 | Cyclical vomiting, in migraine, not intractable |
| Migraine | 340 | ICD10CM | G43.B1 | Ophthalmoplegic migraine, intractable |
| Migraine | 340 | ICD10CM | G44.039 | Episodic paroxysmal hemicrania, not intractable |
| Migraine | 340 | ICD10CM | G43.821 | Menstrual migraine, not intractable, with status migrainosus |
| Migraine | 340 | ICD9CM | 346.7 | Chronic migraine without aura |
| Migraine | 340 | ICD10CM | G43.71 | Chronic migraine without aura, intractable |
| Migraine | 340 | ICD10CM | G43.80 | Other migraine, not intractable |
| Migraine | 340 | ICD9CM | 346.12 | Migraine without aura, without mention of intractable migraine with status migrainosus |
| Migraine | 340 | ICD10CM | G43.40 | Hemiplegic migraine, not intractable |
| Migraine | 340 | ICD9CM | 346.4 | Menstrual migraine |
| Migraine | 340 | ICD10CM | G43.81 | Other migraine, intractable |
| Migraine | 340 | ICD10CM | G43.C0 | Periodic headache syndromes in child or adult, not intractable |
| Migraine | 340 | ICD10CM | G44.029 | Chronic cluster headache, not intractable |
| Migraine | 340 | ICD10CM | G43.00 | Migraine without aura, not intractable |
| Migraine | 340 | ICD10CM | G43.01 | Migraine without aura, intractable |
| Migraine | 340 | ICD10CM | G43.709 | Chronic migraine without aura, not intractable, without status migrainosus |
| Migraine | 340 | ICD9CM | 346.5 | Persistent migraine aura without cerebral infarction, without mention of intractable migraine without mention of status migrainosus |
| Migraine | 340 | ICD10CM | G43.11 | Migraine with aura, intractable |
| Migraine | 340 | ICD9CM | 346 | Migraine with aura |
| Migraine | 340 | ICD9CM | 346.02 | Migraine with aura, without mention of intractable migraine with status migrainosus |

| | | | | |
|---|---|---|---|---|
| Migraine | 340 | ICD10CM | G43.509 | Persistent migraine aura without cerebral infarction, not intractable, without status migrainosus |
| Migraine | 340 | ICD10CM | G43.1 | Migraine with aura |
| Migraine | 340 | ICD10CM | G43.10 | Migraine with aura, not intractable |
| Migraine | 340 | ICD9CM | 346.5 | Persistent migraine aura without cerebral infarction |
| Migraine | 340 | ICD9CM | 346.53 | Persistent migraine aura without cerebral infarction, with intractable migraine, so stated, with status migrainosus |
| Migraine | 340 | ICD10CM | G43.119 | Migraine with aura, intractable, without status migrainosus |
| Migraine | 340 | ICD10CM | G43.519 | Persistent migraine aura without cerebral infarction, intractable, without status migrainosus |
| Migraine | 340 | ICD10CM | G43.111 | Migraine with aura, intractable, with status migrainosus |
| Migraine | 340 | ICD10CM | G43.511 | Persistent migraine aura without cerebral infarction, intractable, with status migrainosus |
| Migraine | 340 | ICD10CM | G43.101 | Migraine with aura, not intractable, with status migrainosus |
| Migraine | 340 | ICD9CM | 346.01 | Migraine with aura, with intractable migraine, so stated, without mention of status migrainosus |
| Migraine | 340 | ICD9CM | 346.51 | Persistent migraine aura without cerebral infarction, with intractable migraine, so stated, without mention of status migrainosus |
| Migraine | 340 | ICD9CM | 346.03 | Migraine with aura, with intractable migraine, so stated, with status migrainosus |
| Migraine | 340 | ICD10CM | G43.109 | Migraine with aura, not intractable, without status migrainosus |
| Migraine | 340 | ICD9CM | 346 | Migraine with aura, without mention of intractable migraine without mention of status migrainosus |
| Epilepsy | 345.1 | ICD9CM | 345.7 | Epilepsia partialis continua |
| Epilepsy | 345.1 | ICD10CM | G40.311 | Generalized idiopathic epilepsy and epileptic syndromes, intractable, with status epilepticus |
| Epilepsy | 345.1 | ICD10CM | G40.B1 | Juvenile myoclonic epilepsy, intractable |
| Epilepsy | 345.1 | ICD10CM | G40.834 | Dravet syndrome, intractable, without status epilepticus |
| Epilepsy | 345.1 | ICD10CM | G40.A | Absence epileptic syndrome |
| Epilepsy | 345.1 | ICD9CM | 345.3 | Grand mal status |
| Epilepsy | 345.1 | ICD10CM | G40.301 | Generalized idiopathic epilepsy and epileptic syndromes, not intractable, with status epilepticus |
| Epilepsy | 345.1 | ICD10CM | G40.C09 | Lafora progressive myoclonus epilepsy, not intractable, without status epilepticus |
| Epilepsy | 345.1 | ICD10CM | G40.822 | Epileptic spasms, not intractable, without status epilepticus |
| Epilepsy | 345.1 | ICD10CM | G40.82 | Epileptic spasms |

| | | | | |
|---|---|---|---|---|
| Epilepsy | 345.1 | ICD10CM | G40.50 | Epileptic seizures related to external causes, not intractable |
| Epilepsy | 345.1 | ICD10CM | G40.B0 | Juvenile myoclonic epilepsy, not intractable |
| Epilepsy | 345.1 | ICD10CM | G40.A01 | Absence epileptic syndrome, not intractable, with status epilepticus |
| Epilepsy | 345.1 | ICD9CM | 345 | Generalized nonconvulsive epilepsy, without mention of intractable epilepsy |
| Epilepsy | 345.1 | ICD9CM | 345.61 | Infantile spasms, with intractable epilepsy |
| Epilepsy | 345.1 | ICD10CM | G40.A1 | Absence epileptic syndrome, intractable |
| Epilepsy | 345.1 | ICD10CM | G40.814 | Lennox-Gastaut syndrome, intractable, without status epilepticus |
| Epilepsy | 345.1 | ICD10CM | G40.B09 | Juvenile myoclonic epilepsy, not intractable, without status epilepticus |
| Epilepsy | 345.1 | ICD10CM | G40.A09 | Absence epileptic syndrome, not intractable, without status epilepticus |
| Epilepsy | 345.1 | ICD9CM | 345.8 | Other forms of epilepsy and recurrent seizures, without mention of intractable epilepsy |
| Epilepsy | 345.1 | ICD9CM | 345 | Generalized nonconvulsive epilepsy |
| Epilepsy | 345.1 | ICD10CM | G40.813 | Lennox-Gastaut syndrome, intractable, with status epilepticus |
| Epilepsy | 345.1 | ICD10CM | G40.812 | Lennox-Gastaut syndrome, not intractable, without status epilepticus |
| Epilepsy | 345.1 | ICD10CM | G40.A11 | Absence epileptic syndrome, intractable, with status epilepticus |
| Epilepsy | 345.1 | ICD10CM | G40.B01 | Juvenile myoclonic epilepsy, not intractable, with status epilepticus |
| Epilepsy | 345.1 | ICD10CM | G40.1 | Localization-related (focal) (partial) symptomatic epilepsy and epileptic syndromes with simple partial seizures |
| Epilepsy | 345.1 | ICD9CM | 345.11 | Genetic generalized epilepsy, with intractable epilepsy |
| Epilepsy | 345.1 | ICD10CM | G40.42 | Cyclin-Dependent Kinase-Like 5 Deficiency Disorder |
| Epilepsy | 345.1 | ICD10CM | G40.B19 | Juvenile myoclonic epilepsy, intractable, without status epilepticus |
| Epilepsy | 345.1 | ICD9CM | 345.2 | Petit mal status |
| Epilepsy | 345.1 | ICD10CM | G40.319 | Generalized idiopathic epilepsy and epileptic syndromes, intractable, without status epilepticus |
| Epilepsy | 345.1 | ICD9CM | 345.6 | Infantile spasms |
| Epilepsy | 345.1 | ICD10CM | G40.A0 | Absence epileptic syndrome, not intractable |
| Epilepsy | 345.1 | ICD9CM | 345.7 | Epilepsia partialis continua, without mention of intractable epilepsy |
| Epilepsy | 345.1 | ICD9CM | 345.1 | Genetic generalized epilepsy, without mention of intractable epilepsy |
| Epilepsy | 345.1 | ICD10CM | G40.409 | Other generalized epilepsy and epileptic syndromes, not intractable, without status epilepticus |

| | | | | |
|---|---|---|---|---|
| Epilepsy | 345.1 | ICD9CM | 345.4 | Localization-related (focal) (partial) epilepsy and epileptic syndromes with complex partial seizures |
| Epilepsy | 345.1 | ICD9CM | 345.1 | Genetic generalized epilepsy |
| Epilepsy | 345.1 | ICD10CM | G40.4 | Other generalized epilepsy and epileptic syndromes |
| Epilepsy | 345.1 | ICD10CM | G40.3 | Generalized idiopathic epilepsy and epileptic syndromes |
| Epilepsy | 345.1 | ICD10CM | G40.309 | Generalized idiopathic epilepsy and epileptic syndromes, not intractable, without status epilepticus |
| Epilepsy | 345.1 | ICD10CM | G40.211 | Localization-related (focal) (partial) symptomatic epilepsy and epileptic syndromes with complex partial seizures, intractable, with status epilepticus |
| Epilepsy | 345.1 | ICD10CM | G40.20 | Localization-related (focal) (partial) symptomatic epilepsy and epileptic syndromes with complex partial seizures, not intractable |
| Epilepsy | 345.1 | ICD10CM | G40.209 | Localization-related (focal) (partial) symptomatic epilepsy and epileptic syndromes with complex partial seizures, not intractable, without status epilepticus |
| Epilepsy | 345.1 | ICD10CM | G40.01 | Localization-related (focal) (partial) idiopathic epilepsy and epileptic syndromes with seizures of localized onset, intractable |
| Epilepsy | 345.1 | ICD10CM | G40.101 | Localization-related (focal) (partial) symptomatic epilepsy and epileptic syndromes with simple partial seizures, not intractable, with status epilepticus |
| Epilepsy | 345.1 | ICD9CM | 345.5 | Localization-related (focal) (partial) epilepsy and epileptic syndromes with simple partial seizures |
| Epilepsy | 345.1 | ICD9CM | 345.91 | Epilepsy, unspecified, with intractable epilepsy |
| Epilepsy | 345.1 | ICD10CM | G40.009 | Localization-related (focal) (partial) idiopathic epilepsy and epileptic syndromes with seizures of localized onset, not intractable, without status epilepticus |
| Epilepsy | 345.1 | ICD9CM | 345.41 | Localization-related (focal) (partial) epilepsy and epileptic syndromes with complex partial seizures, with intractable epilepsy |
| Epilepsy | 345.1 | ICD10CM | G40.119 | Localization-related (focal) (partial) symptomatic epilepsy and epileptic syndromes with simple partial seizures, intractable, without status epilepticus |
| Epilepsy | 345.1 | ICD10CM | G40.011 | Localization-related (focal) (partial) idiopathic epilepsy and epileptic syndromes with seizures of localized onset, intractable, with status epilepticus |
| Epilepsy | 345.1 | ICD10CM | G40.2 | Localization-related (focal) (partial) symptomatic epilepsy and epileptic syndromes with complex partial seizures |

| | | | | |
|---|---|---|---|---|
| Epilepsy | 345.1 | ICD10CM | G40.219 | Localization-related (focal) (partial) symptomatic epilepsy and epileptic syndromes with complex partial seizures, intractable, without status epilepticus |
| Epilepsy | 345.1 | ICD10CM | G40.A19 | Absence epileptic syndrome, intractable, without status epilepticus |
| Epilepsy | 345.1 | ICD10CM | G40.C11 | Lafora progressive myoclonus epilepsy, intractable, with status epilepticus |
| Epilepsy | 345.1 | ICD9CM | 345.8 | Other forms of epilepsy and recurrent seizures |
| Epilepsy | 345.1 | ICD10CM | G40.801 | Other epilepsy, not intractable, with status epilepticus |
| Epilepsy | 345.1 | ICD9CM | 345.6 | Infantile spasms, without mention of intractable epilepsy |
| Epilepsy | 345.1 | ICD10CM | G40.802 | Other epilepsy, not intractable, without status epilepticus |
| Epilepsy | 345.1 | ICD10CM | G40.811 | Lennox-Gastaut syndrome, not intractable, with status epilepticus |
| Epilepsy | 345.1 | ICD10CM | G40.833 | Dravet syndrome, intractable, with status epilepticus |
| Epilepsy | 345.1 | ICD10CM | G40.411 | Other generalized epilepsy and epileptic syndromes, intractable, with status epilepticus |
| Epilepsy | 345.1 | ICD10CM | G40.B11 | Juvenile myoclonic epilepsy, intractable, with status epilepticus |
| Epilepsy | 345.1 | ICD10CM | G40.509 | Epileptic seizures related to external causes, not intractable, without status epilepticus |
| Epilepsy | 345.1 | ICD10CM | G40.804 | Other epilepsy, intractable, without status epilepticus |
| Epilepsy | 345.1 | ICD9CM | 345.81 | Other forms of epilepsy and recurrent seizures, with intractable epilepsy |
| Epilepsy | 345.1 | ICD10CM | G40.C19 | Lafora progressive myoclonus epilepsy, intractable, without status epilepticus |
| Epilepsy | 345.1 | ICD10CM | G40.401 | Other generalized epilepsy and epileptic syndromes, not intractable, with status epilepticus |
| Epilepsy | 345.1 | ICD10CM | G40.824 | Epileptic spasms, intractable, without status epilepticus |
| Epilepsy | 345.1 | ICD10CM | G40.0 | Localization-related (focal) (partial) idiopathic epilepsy and epileptic syndromes with seizures of localized onset |
| Epilepsy | 345.1 | ICD9CM | 345.4 | Localization-related (focal) (partial) epilepsy and epileptic syndromes with complex partial seizures, without mention of intractable epilepsy |
| Epilepsy | 345.1 | ICD10CM | G40.001 | Localization-related (focal) (partial) idiopathic epilepsy and epileptic syndromes with seizures of localized onset, not intractable, with status epilepticus |
| Epilepsy | 345.1 | ICD10CM | G40.501 | Epileptic seizures related to external causes, not intractable, with status epilepticus |
| Epilepsy | 345.1 | ICD10CM | G40.89 | Other seizures |
| Epilepsy | 345.1 | ICD10CM | G40.5 | Epileptic seizures related to external causes |

| | | | | |
|---|---|---|---|---|
| Epilepsy | 345.1 | ICD10CM | G40.B | Juvenile myoclonic epilepsy [impulsive petit mal] |
| Epilepsy | 345.1 | ICD10CM | G40.C01 | Lafora progressive myoclonus epilepsy, not intractable, with status epilepticus |
| Epilepsy | 345.1 | ICD10CM | G40.823 | Epileptic spasms, intractable, with status epilepticus |
| Epilepsy | 345.1 | ICD10CM | G40.821 | Epileptic spasms, not intractable, with status epilepticus |
| Epilepsy | 345.1 | ICD9CM | 345.01 | Generalized nonconvulsive epilepsy, with intractable epilepsy |
| Epilepsy | 345.1 | ICD10CM | G40.911 | Epilepsy, unspecified, intractable, with status epilepticus |
| Epilepsy | 345.1 | ICD10CM | G40.419 | Other generalized epilepsy and epileptic syndromes, intractable, without status epilepticus |
| Epilepsy | 345.1 | ICD10CM | G40.803 | Other epilepsy, intractable, with status epilepticus |
| Epilepsy | 345.1 | ICD9CM | 345.71 | Epilepsia partialis continua, with intractable epilepsy |
| Epilepsy | 345.1 | ICD10CM | G40.109 | Localization-related (focal) (partial) symptomatic epilepsy and epileptic syndromes with simple partial seizures, not intractable, without status epilepticus |
| Epilepsy | 345.1 | ICD10CM | G40.21 | Localization-related (focal) (partial) symptomatic epilepsy and epileptic syndromes with complex partial seizures, intractable |
| Epilepsy | 345.1 | ICD9CM | 345.51 | Localization-related (focal) (partial) epilepsy and epileptic syndromes with simple partial seizures, with intractable epilepsy |
| Epilepsy | 345.1 | ICD9CM | 345.5 | Localization-related (focal) (partial) epilepsy and epileptic syndromes with simple partial seizures, without mention of intractable epilepsy |
| Epilepsy | 345.1 | ICD10CM | G40.201 | Localization-related (focal) (partial) symptomatic epilepsy and epileptic syndromes with complex partial seizures, not intractable, with status epilepticus |
| Epilepsy | 345.1 | ICD10CM | G40.919 | Epilepsy, unspecified, intractable, without status epilepticus |
| Epilepsy | 345.1 | ICD10CM | G40.111 | Localization-related (focal) (partial) symptomatic epilepsy and epileptic syndromes with simple partial seizures, intractable, with status epilepticus |
| Epilepsy | 345.1 | ICD10CM | G40.019 | Localization-related (focal) (partial) idiopathic epilepsy and epileptic syndromes with seizures of localized onset, intractable, without status epilepticus |
| Epilepsy | 345.1 | ICD10CM | G40.00 | Localization-related (focal) (partial) idiopathic epilepsy and epileptic syndromes with seizures of localized onset, not intractable |
| Genetic generalized epilepsy | 345.11 | ICD10CM | G40.42 | Cyclin-Dependent Kinase-Like 5 Deficiency Disorder |
| Genetic generalized epilepsy | 345.11 | ICD9CM | 345.1 | Genetic generalized epilepsy, without mention of intractable epilepsy |

| | | | | | |
|---|---|---|---|---|---|
| | Genetic generalized epilepsy | 345.11 | ICD9CM | 345.11 | Genetic generalized epilepsy, with intractable epilepsy |
| | Genetic generalized epilepsy | 345.11 | ICD10CM | G40.409 | Other generalized epilepsy and epileptic syndromes, not intractable, without status epilepticus |
| | Genetic generalized epilepsy | 345.11 | ICD10CM | G40.3 | Generalized idiopathic epilepsy and epileptic syndromes |
| | Genetic generalized epilepsy | 345.11 | ICD10CM | G40.309 | Generalized idiopathic epilepsy and epileptic syndromes, not intractable, without status epilepticus |
| | Genetic generalized epilepsy | 345.11 | ICD9CM | 345.1 | Genetic generalized epilepsy |
| | Genetic generalized epilepsy | 345.11 | ICD10CM | G40.4 | Other generalized epilepsy and epileptic syndromes |
| **Sense Organs** | Hearing loss | 389 | ICD10CM | H91.92 | Unspecified hearing loss, left ear |
| | Hearing loss | 389 | ICD10CM | H91.93 | Unspecified hearing loss, bilateral |
| | Hearing loss | 389 | ICD10CM | H91.9 | Unspecified hearing loss |
| | Hearing loss | 389 | ICD10CM | Z97.4 | Presence of external hearing-aid |
| | Hearing loss | 389 | ICD9CM | 389.2 | Mixed conductive and sensorineural hearing loss |
| | Hearing loss | 389 | ICD10CM | H91.09 | Ototoxic hearing loss, unspecified ear |
| | Hearing loss | 389 | ICD9CM | 389.8 | Other specified forms of hearing loss |
| | Hearing loss | 389 | ICD9CM | V53.2 | Fitting and adjustment of hearing aid |
| | Hearing loss | 389 | ICD10CM | H90.8 | Mixed conductive and sensorineural hearing loss, unspecified |
| | Hearing loss | 389 | ICD10CM | H90.7 | Mixed conductive and sensorineural hearing loss, unilateral with unrestricted hearing on the contralateral side |
| | Hearing loss | 389 | ICD10CM | H90.6 | Mixed conductive and sensorineural hearing loss, bilateral |
| | Hearing loss | 389 | ICD10CM | H91.20 | Sudden idiopathic hearing loss, unspecified ear |
| | Hearing loss | 389 | ICD10CM | H91.91 | Unspecified hearing loss, right ear |
| | Hearing loss | 389 | ICD10CM | H90.A3 | Mixed conductive and sensorineural hearing loss, unilateral with restricted hearing on the contralateral side |
| | Hearing loss | 389 | ICD10CM | H90.A | Conductive and sensorineural hearing loss with restricted hearing on the contralateral side |
| | Hearing loss | 389 | ICD10CM | H91.90 | Unspecified hearing loss, unspecified ear |
| | Hearing loss | 389 | ICD9CM | 389.7 | Deaf, nonspeaking, not elsewhere classifiable |
| | Hearing loss | 389 | ICD10CM | H90.72 | Mixed conductive and sensorineural hearing loss, unilateral, left ear, with unrestricted hearing on the contralateral side |
| | Hearing loss | 389 | ICD10CM | H90 | Conductive and sensorineural hearing loss |
| | Hearing loss | 389 | ICD10CM | Z96.29 | Presence of other otological and audiological implants |
| | Hearing loss | 389 | ICD9CM | 389.21 | Mixed hearing loss, unilateral |
| | Hearing loss | 389 | ICD10CM | Z46.1 | Encounter for fitting and adjustment of hearing aid |

| | | | | |
|---|---|---|---|---|
| Hearing loss | 389 | ICD10CM | H90.A32 | Mixed conductive and sensorineural hearing loss, unilateral, left ear with restricted hearing on the contralateral side |
| Hearing loss | 389 | ICD10CM | H91.03 | Ototoxic hearing loss, bilateral |
| Hearing loss | 389 | ICD9CM | V41.2 | Problems with hearing |
| Hearing loss | 389 | ICD9CM | 389.22 | Mixed hearing loss, bilateral |
| Hearing loss | 389 | ICD10CM | H91.8X | Other specified hearing loss |
| Hearing loss | 389 | ICD10CM | H91.8X3 | Other specified hearing loss, bilateral |
| Hearing loss | 389 | ICD10CM | Z96.21 | Cochlear implant status |
| Hearing loss | 389 | ICD10CM | H91.01 | Ototoxic hearing loss, right ear |
| Hearing loss | 389 | ICD9CM | 389.2 | Mixed hearing loss, unspecified |
| Hearing loss | 389 | ICD10CM | H91.23 | Sudden idiopathic hearing loss, bilateral |
| Hearing loss | 389 | ICD10CM | Z96.2 | Presence of otological and audiological implants |
| Hearing loss | 389 | ICD9CM | 389.9 | Unspecified hearing loss |
| Hearing loss | 389 | ICD10CM | H90.71 | Mixed conductive and sensorineural hearing loss, unilateral, right ear, with unrestricted hearing on the contralateral side |
| Hearing loss | 389 | ICD10CM | H91.2 | Sudden idiopathic hearing loss |
| Hearing loss | 389 | ICD10CM | H91 | Other and unspecified hearing loss |
| Hearing loss | 389 | ICD10CM | H91.8X1 | Other specified hearing loss, right ear |
| Hearing loss | 389 | ICD10CM | H91.8 | Other specified hearing loss |
| Hearing loss | 389 | ICD10CM | Z96.22 | Myringotomy tube(s) status |
| Hearing loss | 389 | ICD10CM | H91.3 | Deaf nonspeaking, not elsewhere classified |
| Hearing loss | 389 | ICD10CM | Z96.20 | Presence of otological and audiological implant, unspecified |
| Hearing loss | 389 | ICD10CM | H91.21 | Sudden idiopathic hearing loss, right ear |
| Hearing loss | 389 | ICD9CM | 389 | Hearing loss |
| Hearing loss | 389 | ICD10CM | H91.22 | Sudden idiopathic hearing loss, left ear |
| Hearing loss | 389 | ICD10CM | H91.8X2 | Other specified hearing loss, left ear |
| Hearing loss | 389 | ICD10CM | H91.0 | Ototoxic hearing loss |
| Hearing loss | 389 | ICD10CM | H90.A31 | Mixed conductive and sensorineural hearing loss, unilateral, right ear with restricted hearing on the contralateral side |
| Hearing loss | 389 | ICD9CM | V49.85 | Dual sensory impairment |
| Hearing loss | 389 | ICD10CM | H91.02 | Ototoxic hearing loss, left ear |
| Hearing loss | 389 | ICD10CM | H91.8X9 | Other specified hearing loss, unspecified ear |
| Hearing loss | 389 | ICD9CM | 388.2 | Sudden hearing loss, unspecified |
| Hearing loss | 389 | ICD10CM | H90.4 | Sensorineural hearing loss, unilateral with unrestricted hearing on the contralateral side |
| Hearing loss | 389 | ICD9CM | 389.1 | Sensorineural hearing loss |
| Hearing loss | 389 | ICD9CM | 389.16 | Sensorineural hearing loss, asymmetrical |

| | | | | |
|---|---|---|---|---|
| Hearing loss | 389 | ICD10CM | H90.41 | Sensorineural hearing loss, unilateral, right ear, with unrestricted hearing on the contralateral side |
| Hearing loss | 389 | ICD9CM | 389.14 | Central hearing loss |
| Hearing loss | 389 | ICD10CM | H90.A2 | Sensorineural hearing loss, unilateral, with restricted hearing on the contralateral side |
| Hearing loss | 389 | ICD10CM | H90.3 | Sensorineural hearing loss, bilateral |
| Hearing loss | 389 | ICD9CM | 389.15 | Sensorineural hearing loss, unilateral |
| Hearing loss | 389 | ICD10CM | H90.A22 | Sensorineural hearing loss, unilateral, left ear, with restricted hearing on the contralateral side |
| Hearing loss | 389 | ICD10CM | H90.42 | Sensorineural hearing loss, unilateral, left ear, with unrestricted hearing on the contralateral side |
| Hearing loss | 389 | ICD9CM | 389.12 | Neural hearing loss, bilateral |
| Hearing loss | 389 | ICD9CM | 389.11 | Sensory hearing loss, bilateral |
| Hearing loss | 389 | ICD9CM | 389.1 | Sensorineural hearing loss, unspecified |
| Hearing loss | 389 | ICD10CM | H90.A21 | Sensorineural hearing loss, unilateral, right ear, with restricted hearing on the contralateral side |
| Hearing loss | 389 | ICD10CM | H90.5 | Unspecified sensorineural hearing loss |
| Hearing loss | 389 | ICD9CM | 389.13 | Neural hearing loss, unilateral |
| Hearing loss | 389 | ICD9CM | 389.17 | Sensory hearing loss, unilateral |
| Hearing loss | 389 | ICD9CM | 389.18 | Sensorineural hearing loss, bilateral |
| Hearing loss | 389 | ICD10CM | H93.292 | Other abnormal auditory perceptions, left ear |
| Hearing loss | 389 | ICD9CM | 388.4 | Other abnormal auditory perception |
| Hearing loss | 389 | ICD10CM | H93.3X | Disorders of acoustic nerve |
| Hearing loss | 389 | ICD10CM | H93.3X2 | Disorders of left acoustic nerve |
| Hearing loss | 389 | ICD10CM | H90.A1 | Conductive hearing loss, unilateral, with restricted hearing on the contralateral side |
| Hearing loss | 389 | ICD9CM | 389.08 | Conductive hearing loss of combined types |
| Hearing loss | 389 | ICD9CM | 389 | Conductive hearing loss |
| Hearing loss | 389 | ICD9CM | 389.01 | Conductive hearing loss, external ear |
| Hearing loss | 389 | ICD10CM | H90.12 | Conductive hearing loss, unilateral, left ear, with unrestricted hearing on the contralateral side |
| Hearing loss | 389 | ICD9CM | 388.02 | Transient ischemic deafness |
| Hearing loss | 389 | ICD10CM | H83.3X3 | Noise effects on inner ear, bilateral |
| Hearing loss | 389 | ICD10CM | H93.0 | Degenerative and vascular disorders of ear |
| Hearing loss | 389 | ICD10CM | H83.3 | Noise effects on inner ear |
| Hearing loss | 389 | ICD10CM | H93.019 | Transient ischemic deafness, unspecified ear |
| Hearing loss | 389 | ICD9CM | 388 | Degenerative and vascular disorders, unspecified |
| Hearing loss | 389 | ICD9CM | 388.1 | Noise effects on inner ear |
| Hearing loss | 389 | ICD10CM | H91.1 | Presbycusis |
| Hearing loss | 389 | ICD9CM | 388.3 | Tinnitus |

| | | | | |
|---|---|---|---|---|
| Hearing loss | 389 | ICD10CM | H93.A3 | Pulsatile tinnitus, bilateral |
| Hearing loss | 389 | ICD10CM | H93.19 | Tinnitus, unspecified ear |
| Hearing loss | 389 | ICD10CM | H93.A9 | Pulsatile tinnitus, unspecified ear |
| Hearing loss | 389 | ICD10CM | H93.13 | Tinnitus, bilateral |
| Hearing loss | 389 | ICD10CM | H93.A1 | Pulsatile tinnitus, right ear |
| Hearing loss | 389 | ICD10CM | H93.12 | Tinnitus, left ear |
| Hearing loss | 389 | ICD10CM | H93.A2 | Pulsatile tinnitus, left ear |
| Hearing loss | 389 | ICD10CM | H90.2 | Conductive hearing loss, unspecified |
| Hearing loss | 389 | ICD9CM | 389.04 | Conductive hearing loss, inner ear |
| Hearing loss | 389 | ICD9CM | 389.06 | Conductive hearing loss, bilateral |
| Hearing loss | 389 | ICD9CM | 389 | Conductive hearing loss, unspecified |
| Hearing loss | 389 | ICD9CM | 389.03 | Conductive hearing loss, middle ear |
| Hearing loss | 389 | ICD9CM | 389.05 | Conductive hearing loss, unilateral |
| Hearing loss | 389 | ICD10CM | H90.1 | Conductive hearing loss, unilateral with unrestricted hearing on the contralateral side |
| Hearing loss | 389 | ICD10CM | H90.11 | Conductive hearing loss, unilateral, right ear, with unrestricted hearing on the contralateral side |
| Hearing loss | 389 | ICD10CM | H90.0 | Conductive hearing loss, bilateral |
| Hearing loss | 389 | ICD9CM | 389.02 | Conductive hearing loss, tympanic membrane |
| Hearing loss | 389 | ICD10CM | H90.A12 | Conductive hearing loss, unilateral, left ear with restricted hearing on the contralateral side |
| Hearing loss | 389 | ICD10CM | H90.A11 | Conductive hearing loss, unilateral, right ear with restricted hearing on the contralateral side |
| Hearing loss | 389 | ICD10CM | H93.093 | Unspecified degenerative and vascular disorders of ear, bilateral |
| Hearing loss | 389 | ICD10CM | H93.091 | Unspecified degenerative and vascular disorders of right ear |
| Hearing loss | 389 | ICD10CM | H91.11 | Presbycusis, right ear |
| Hearing loss | 389 | ICD9CM | 388.12 | Noise-induced hearing loss |
| Hearing loss | 389 | ICD10CM | H93.09 | Unspecified degenerative and vascular disorders of ear |
| Hearing loss | 389 | ICD10CM | H93.29 | Other abnormal auditory perceptions |
| Hearing loss | 389 | ICD10CM | H93.213 | Auditory recruitment, bilateral |
| Hearing loss | 389 | ICD10CM | H93.223 | Diplacusis, bilateral |
| Hearing loss | 389 | ICD9CM | 794.15 | Nonspecific abnormal auditory function studies |
| Hearing loss | 389 | ICD10CM | H93.241 | Temporary auditory threshold shift, right ear |
| Hearing loss | 389 | ICD10CM | H93.21 | Auditory recruitment |
| Hearing loss | 389 | ICD10CM | H93.231 | Hyperacusis, right ear |
| Hearing loss | 389 | ICD10CM | H93.011 | Transient ischemic deafness, right ear |
| Hearing loss | 389 | ICD10CM | H93.013 | Transient ischemic deafness, bilateral |
| Hearing loss | 389 | ICD9CM | 388.4 | Abnormal auditory perception, unspecified |

| | | | | |
|---|---|---|---|---|
| Hearing loss | 389 | ICD10CM | H93.242 | Temporary auditory threshold shift, left ear |
| Hearing loss | 389 | ICD9CM | 388.5 | Disorders of acoustic nerve |
| Hearing loss | 389 | ICD10CM | H93.2 | Other abnormal auditory perceptions |
| Hearing loss | 389 | ICD9CM | 388.1 | Noise effects on inner ear, unspecified |
| Hearing loss | 389 | ICD10CM | H93.092 | Unspecified degenerative and vascular disorders of left ear |
| Hearing loss | 389 | ICD9CM | 388.01 | Presbyacusis |
| Hearing loss | 389 | ICD10CM | H91.10 | Presbycusis, unspecified ear |
| Hearing loss | 389 | ICD10CM | H91.13 | Presbycusis, bilateral |
| Hearing loss | 389 | ICD10CM | H93.099 | Unspecified degenerative and vascular disorders of unspecified ear |
| Hearing loss | 389 | ICD10CM | H83.3X9 | Noise effects on inner ear, unspecified ear |
| Hearing loss | 389 | ICD10CM | H93.012 | Transient ischemic deafness, left ear |
| Hearing loss | 389 | ICD10CM | H83.3X1 | Noise effects on right inner ear |
| Hearing loss | 389 | ICD10CM | H91.12 | Presbycusis, left ear |
| Hearing loss | 389 | ICD9CM | 388 | Degenerative and vascular disorders of ear |
| Hearing loss | 389 | ICD10CM | H93.249 | Temporary auditory threshold shift, unspecified ear |
| Hearing loss | 389 | ICD10CM | H93.24 | Temporary auditory threshold shift |
| Hearing loss | 389 | ICD10CM | H93.3 | Disorders of acoustic nerve |
| Hearing loss | 389 | ICD10CM | H93.219 | Auditory recruitment, unspecified ear |
| Hearing loss | 389 | ICD9CM | 388.44 | Auditory recruitment |
| Hearing loss | 389 | ICD10CM | H83.3X2 | Noise effects on left inner ear |
| Hearing loss | 389 | ICD10CM | H93.01 | Transient ischemic deafness |
| Hearing loss | 389 | ICD10CM | H93.3X9 | Disorders of unspecified acoustic nerve |
| Hearing loss | 389 | ICD10CM | H93.293 | Other abnormal auditory perceptions, bilateral |
| Hearing loss | 389 | ICD9CM | 388.43 | Impairment of auditory discrimination |
| Hearing loss | 389 | ICD10CM | H94.03 | Acoustic neuritis in infectious and parasitic diseases classified elsewhere, bilateral |
| Hearing loss | 389 | ICD10CM | H93.3X3 | Disorders of bilateral acoustic nerves |
| Hearing loss | 389 | ICD10CM | H94.01 | Acoustic neuritis in infectious and parasitic diseases classified elsewhere, right ear |
| Hearing loss | 389 | ICD10CM | R94.120 | Abnormal auditory function study |
| Hearing loss | 389 | ICD10CM | H93.212 | Auditory recruitment, left ear |
| Hearing loss | 389 | ICD10CM | H93.233 | Hyperacusis, bilateral |
| Hearing loss | 389 | ICD10CM | H83.3X | Noise effects on inner ear |
| Hearing loss | 389 | ICD9CM | 388.45 | Acquired auditory processing disorder |
| Hearing loss | 389 | ICD10CM | H93.232 | Hyperacusis, left ear |
| Hearing loss | 389 | ICD10CM | H94.0 | Acoustic neuritis in infectious and parasitic diseases classified elsewhere |
| Hearing loss | 389 | ICD10CM | H93.299 | Other abnormal auditory perceptions, unspecified ear |

| | | | | | |
|---|---|---|---|---|---|
| | Hearing loss | 389 | ICD10CM | H94.02 | Acoustic neuritis in infectious and parasitic diseases classified elsewhere, left ear |
| | Hearing loss | 389 | ICD10CM | H93.222 | Diplacusis, left ear |
| | Hearing loss | 389 | ICD10CM | H93.243 | Temporary auditory threshold shift, bilateral |
| | Hearing loss | 389 | ICD10CM | H93.239 | Hyperacusis, unspecified ear |
| | Hearing loss | 389 | ICD10CM | H93.22 | Diplacusis |
| | Hearing loss | 389 | ICD10CM | H93.291 | Other abnormal auditory perceptions, right ear |
| | Hearing loss | 389 | ICD10CM | H93.211 | Auditory recruitment, right ear |
| | Hearing loss | 389 | ICD10CM | H93.221 | Diplacusis, right ear |
| | Hearing loss | 389 | ICD9CM | 388.41 | Diplacusis |
| | Hearing loss | 389 | ICD10CM | H93.A | Pulsatile tinnitus |
| | Hearing loss | 389 | ICD9CM | 388.3 | Tinnitus, unspecified |
| | Hearing loss | 389 | ICD9CM | 388.32 | Objective tinnitus |
| | Hearing loss | 389 | ICD10CM | H93.1 | Tinnitus |
| | Hearing loss | 389 | ICD9CM | 388.31 | Subjective tinnitus |
| | Hearing loss | 389 | ICD10CM | H93.11 | Tinnitus, right ear |
| | Hearing loss | 389 | ICD10CM | H94.00 | Acoustic neuritis in infectious and parasitic diseases classified elsewhere, unspecified ear |
| | Hearing loss | 389 | ICD10CM | H93.229 | Diplacusis, unspecified ear |
| | Hearing loss | 389 | ICD9CM | 388.42 | Hyperacusis |
| | Hearing loss | 389 | ICD10CM | H93.3X1 | Disorders of right acoustic nerve |
| | Hearing loss | 389 | ICD10CM | H93.23 | Hyperacusis |
| **Circulatory System** | Hypertension | 401 | ICD10CM | I97.3 | Postprocedural hypertension |
| | Hypertension | 401 | ICD9CM | 997.91 | Complications affecting other specified body systems, not elsewhere classified, hypertension |
| | Hypertension | 401 | ICD10CM | I1A.0 | Resistant hypertension |
| | Hypertension | 401 | ICD9CM | 401.9 | Unspecified essential hypertension |
| | Hypertension | 401 | ICD9CM | 401.1 | Benign essential hypertension |
| | Hypertension | 401 | ICD9CM | 401 | Essential hypertension |
| | Hypertension | 401 | ICD10CM | I10 | Essential (primary) hypertension |
| | Hypertension | 401 | ICD9CM | 403.1 | Hypertensive renal disease, benign |
| | Hypertension | 401 | ICD9CM | 403.91 | Hypertensive chronic kidney disease, unspecified, with chronic kidney disease stage V or end stage renal disease |
| | Hypertension | 401 | ICD9CM | 403.11 | Hypertensive chronic kidney disease, benign, with chronic kidney disease stage V or end stage renal disease |
| | Hypertension | 401 | ICD10CM | I12.9 | Hypertensive chronic kidney disease with stage 1 through stage 4 chronic kidney disease, or unspecified chronic kidney disease |
| | Hypertension | 401 | ICD9CM | 403.9 | Hypertensive renal disease, unspecified |

| | | | | |
|---|---|---|---|---|
| Hypertension | 401 | ICD9CM | 404.92 | Hypertensive heart and chronic kidney disease, unspecified, without heart failure and with chronic kidney disease stage V or end stage renal disease |
| Hypertension | 401 | ICD9CM | 404 | Hypertensive heart and chronic kidney disease, malignant |
| Hypertension | 401 | ICD9CM | 404.9 | Hypertensive heart and renal disease, unspecified |
| Hypertension | 401 | ICD9CM | 404.11 | Hypertensive heart and chronic kidney disease, benign, with heart failure and with chronic kidney disease stage I through stage IV, or unspecified |
| Hypertension | 401 | ICD9CM | 404.03 | Hypertensive heart and chronic kidney disease, malignant, with heart failure and with chronic kidney disease stage V or end stage renal disease |
| Hypertension | 401 | ICD9CM | 404 | Hypertensive heart and chronic kidney disease |
| Hypertension | 401 | ICD10CM | I13.2 | Hypertensive heart and chronic kidney disease with heart failure and with stage 5 chronic kidney disease, or end stage renal disease |
| Hypertension | 401 | ICD10CM | I13.0 | Hypertensive heart and chronic kidney disease with heart failure and stage 1 through stage 4 chronic kidney disease, or unspecified chronic kidney disease |
| Hypertension | 401 | ICD9CM | 404.01 | Hypertensive heart and chronic kidney disease, malignant, with heart failure and with chronic kidney disease stage I through stage IV, or unspecified |
| Hypertension | 401 | ICD9CM | 404.1 | Hypertensive heart and renal disease, benign |
| Hypertension | 401 | ICD10CM | I13.11 | Hypertensive heart and chronic kidney disease without heart failure, with stage 5 chronic kidney disease, or end stage renal disease |
| Hypertension | 401 | ICD10CM | I13.10 | Hypertensive heart and chronic kidney disease without heart failure, with stage 1 through stage 4 chronic kidney disease, or unspecified chronic kidney disease |
| Hypertension | 401 | ICD10CM | I13.1 | Hypertensive heart and chronic kidney disease without heart failure |
| Hypertension | 401 | ICD9CM | 404.91 | Hypertensive heart and chronic kidney disease, unspecified, with heart failure and with chronic kidney disease stage I through stage IV, or unspecified |
| Hypertension | 401 | ICD9CM | 404.13 | Hypertensive heart and chronic kidney disease, benign, with heart failure and chronic kidney disease stage V or end stage renal disease |
| Hypertension | 401 | ICD10CM | I13 | Hypertensive heart and chronic kidney disease |
| Hypertension | 401 | ICD9CM | 404.02 | Hypertensive heart and chronic kidney disease, malignant, without heart failure and with chronic kidney disease stage V or end stage renal disease |
| Hypertension | 401 | ICD9CM | 404.93 | Hypertensive heart and chronic kidney disease, unspecified, with heart failure and chronic kidney disease stage V or end stage renal disease |

| | | | | |
|---|---|---|---|---|
| Hypertension | 401 | ICD9CM | 404 | Hypertensive heart and chronic kidney disease, malignant, without heart failure and with chronic kidney disease stage I through stage IV, or unspecified |
| Hypertension | 401 | ICD9CM | 404.1 | Hypertensive heart and chronic kidney disease, benign, without heart failure and with chronic kidney disease stage I through stage IV, or unspecified |
| Hypertension | 401 | ICD9CM | 404.12 | Hypertensive heart and chronic kidney disease, benign, without heart failure and with chronic kidney disease stage V or end stage renal disease |
| Hypertension | 401 | ICD9CM | 404.9 | Hypertensive heart and chronic kidney disease, unspecified, without heart failure and with chronic kidney disease stage I through stage IV, or unspecified |
| Hypertension | 401 | ICD9CM | 402 | Malignant hypertensive heart disease |
| Hypertension | 401 | ICD9CM | 402.9 | Unspecified hypertensive heart disease |
| Hypertension | 401 | ICD10CM | I11 | Hypertensive heart disease |
| Hypertension | 401 | ICD9CM | 402.01 | Malignant hypertensive heart disease with heart failure |
| Hypertension | 401 | ICD10CM | I16.1 | Hypertensive emergency |
| Hypertension | 401 | ICD9CM | 405.99 | Other unspecified secondary hypertension |
| Hypertension | 401 | ICD9CM | 405.01 | Malignant renovascular hypertension |
| Hypertension | 401 | ICD9CM | 437.2 | Hypertensive encephalopathy |
| Hypertension | 401 | ICD9CM | 403.01 | Hypertensive chronic kidney disease, malignant, with chronic kidney disease stage V or end stage renal disease |
| Hypertension | 401 | ICD9CM | 403.1 | Hypertensive chronic kidney disease, benign, with chronic kidney disease stage I through stage IV, or unspecified |
| Hypertension | 401 | ICD10CM | N26.2 | Page kidney |
| Hypertension | 401 | ICD10CM | I16 | Hypertensive crisis |
| Hypertension | 401 | ICD9CM | 405.91 | Unspecified renovascular hypertension |
| Hypertension | 401 | ICD10CM | I67.4 | Hypertensive encephalopathy |
| Hypertension | 401 | ICD10CM | I15.8 | Other secondary hypertension |
| Hypertension | 401 | ICD10CM | I15.1 | Hypertension secondary to other renal disorders |
| Hypertension | 401 | ICD10CM | I15.9 | Secondary hypertension, unspecified |
| Hypertension | 401 | ICD9CM | 405 | Malignant secondary hypertension |
| Hypertension | 401 | ICD9CM | 405.09 | Other malignant secondary hypertension |
| Hypertension | 401 | ICD9CM | 405.9 | Unspecified secondary hypertension |
| Hypertension | 401 | ICD9CM | 402 | Malignant hypertensive heart disease without heart failure |
| Hypertension | 401 | ICD9CM | 402.1 | Benign hypertensive heart disease |
| Hypertension | 401 | ICD10CM | I11.9 | Hypertensive heart disease without heart failure |
| Hypertension | 401 | ICD9CM | 402.9 | Unspecified hypertensive heart disease without heart failure |

| | | | | |
|---|---|---|---|---|
| Hypertension | 401 | ICD9CM | 402.1 | Benign hypertensive heart disease without heart failure |
| Hypertension | 401 | ICD9CM | 402 | Hypertensive heart disease |
| Hypertension | 401 | ICD9CM | 402.11 | Benign hypertensive heart disease with heart failure |
| Hypertension | 401 | ICD9CM | 402.91 | Unspecified hypertensive heart disease with heart failure |
| Hypertension | 401 | ICD10CM | I11.0 | Hypertensive heart disease with heart failure |
| Hypertension | 401 | ICD9CM | 403 | Hypertensive chronic kidney disease, malignant, with chronic kidney disease stage I through stage IV, or unspecified |
| Hypertension | 401 | ICD9CM | 403 | Hypertensive chronic kidney disease |
| Hypertension | 401 | ICD10CM | I12 | Hypertensive chronic kidney disease |
| Hypertension | 401 | ICD9CM | 401 | Malignant essential hypertension |
| Hypertension | 401 | ICD10CM | I16.9 | Hypertensive crisis, unspecified |
| Hypertension | 401 | ICD10CM | I16.0 | Hypertensive urgency |
| Hypertension | 401 | ICD10CM | I15 | Secondary hypertension |
| Hypertension | 401 | ICD9CM | 405.19 | Other benign secondary hypertension |
| Hypertension | 401 | ICD10CM | I15.0 | Renovascular hypertension |
| Hypertension | 401 | ICD9CM | 405.1 | Benign secondary hypertension |
| Hypertension | 401 | ICD9CM | 403.9 | Hypertensive chronic kidney disease, unspecified, with chronic kidney disease stage I through stage IV, or unspecified |
| Hypertension | 401 | ICD9CM | 403 | Hypertensive chronic kidney disease, malignant |
| Hypertension | 401 | ICD10CM | I12.0 | Hypertensive chronic kidney disease with stage 5 chronic kidney disease or end stage renal disease |
| Hypertension | 401 | ICD10CM | I15.2 | Hypertension secondary to endocrine disorders |
| Hypertension | 401 | ICD9CM | 405 | Secondary hypertension |
| Hypertension | 401 | ICD9CM | 405.11 | Benign renovascular hypertension |
| Ischemic Heart Disease | 411 | ICD9CM | 411 | Other acute and subacute forms of ischemic heart disease |
| Ischemic Heart Disease | 411 | ICD10CM | I25.110 | Atherosclerotic heart disease of native coronary artery with unstable angina pectoris |
| Ischemic Heart Disease | 411 | ICD10CM | I25.790 | Atherosclerosis of other coronary artery bypass graft(s) with unstable angina pectoris |
| Ischemic Heart Disease | 411 | ICD9CM | 411.1 | Intermediate coronary syndrome |
| Ischemic Heart Disease | 411 | ICD10CM | I25.760 | Atherosclerosis of bypass graft of coronary artery of transplanted heart with unstable angina |
| Ischemic Heart Disease | 411 | ICD10CM | I25.750 | Atherosclerosis of native coronary artery of transplanted heart with unstable angina |
| Ischemic Heart Disease | 411 | ICD10CM | I25.730 | Atherosclerosis of nonautologous biological coronary artery bypass graft(s) with unstable angina pectoris |
| Ischemic Heart Disease | 411 | ICD10CM | I20.0 | Unstable angina |

| | | | | |
|---|---|---|---|---|
| Ischemic Heart Disease | 411 | ICD10CM | I25.700 | Atherosclerosis of coronary artery bypass graft(s), unspecified, with unstable angina pectoris |
| Ischemic Heart Disease | 411 | ICD10CM | I25.710 | Atherosclerosis of autologous vein coronary artery bypass graft(s) with unstable angina pectoris |
| Ischemic Heart Disease | 411 | ICD10CM | I25.720 | Atherosclerosis of autologous artery coronary artery bypass graft(s) with unstable angina pectoris |
| Ischemic Heart Disease | 411 | ICD10CM | I25.719 | Atherosclerosis of autologous vein coronary artery bypass graft(s) with unspecified angina pectoris |
| Ischemic Heart Disease | 411 | ICD10CM | I20 | Angina pectoris |
| Ischemic Heart Disease | 411 | ICD10CM | I20.81 | Angina pectoris with coronary microvascular dysfunction |
| Ischemic Heart Disease | 411 | ICD10CM | I20.89 | Other forms of angina pectoris |
| Ischemic Heart Disease | 411 | ICD10CM | I25.731 | Atherosclerosis of nonautologous biological coronary artery bypass graft(s) with angina pectoris with documented spasm |
| Ischemic Heart Disease | 411 | ICD10CM | I20.1 | Angina pectoris with documented spasm |
| Ischemic Heart Disease | 411 | ICD9CM | 413 | Angina pectoris |
| Ischemic Heart Disease | 411 | ICD10CM | I25.751 | Atherosclerosis of native coronary artery of transplanted heart with angina pectoris with documented spasm |
| Ischemic Heart Disease | 411 | ICD9CM | 413.9 | Other and unspecified angina pectoris |
| Ischemic Heart Disease | 411 | ICD10CM | I25.708 | Atherosclerosis of coronary artery bypass graft(s), unspecified, with other forms of angina pectoris |
| Ischemic Heart Disease | 411 | ICD10CM | I24.8 | Other forms of acute ischemic heart disease |
| Ischemic Heart Disease | 411 | ICD9CM | 411.89 | Other acute and subacute forms of ischemic heart disease, other |
| Ischemic Heart Disease | 411 | ICD9CM | 410.12 | Acute myocardial infarction of other anterior wall, subsequent episode of care |
| Ischemic Heart Disease | 411 | ICD9CM | 410.8 | Acute myocardial infarction, of other specified sites |
| Ischemic Heart Disease | 411 | ICD9CM | 410.01 | Acute myocardial infarction of anterolateral wall, initial episode of care |
| Ischemic Heart Disease | 411 | ICD10CM | I21.19 | ST elevation (STEMI) myocardial infarction involving other coronary artery of inferior wall |
| Ischemic Heart Disease | 411 | ICD9CM | 410.5 | Acute myocardial infarction, of other lateral wall |
| Ischemic Heart Disease | 411 | ICD9CM | 410.02 | Acute myocardial infarction of anterolateral wall, subsequent episode of care |
| Ischemic Heart Disease | 411 | ICD10CM | I21.01 | ST elevation (STEMI) myocardial infarction involving left main coronary artery |

| | | | | |
|---|---|---|---|---|
| Ischemic Heart Disease | 411 | ICD9CM | 410.82 | Acute myocardial infarction of other specified sites, subsequent episode of care |
| Ischemic Heart Disease | 411 | ICD10CM | I22.2 | Subsequent non-ST elevation (NSTEMI) myocardial infarction |
| Ischemic Heart Disease | 411 | ICD9CM | 410.3 | Acute myocardial infarction, of inferoposterior wall |
| Ischemic Heart Disease | 411 | ICD9CM | 410 | Acute myocardial infarction, of anterolateral wall |
| Ischemic Heart Disease | 411 | ICD9CM | 410.5 | Acute myocardial infarction of other lateral wall, episode of care unspecified |
| Ischemic Heart Disease | 411 | ICD10CM | I22 | Subsequent ST elevation (STEMI) and non-ST elevation (NSTEMI) myocardial infarction |
| Ischemic Heart Disease | 411 | ICD10CM | I21.B | Myocardial infarction with coronary microvascular dysfunction |
| Ischemic Heart Disease | 411 | ICD9CM | 410.6 | True posterior wall infarction, episode of care unspecified |
| Ischemic Heart Disease | 411 | ICD10CM | I51.3 | Intracardiac thrombosis, not elsewhere classified |
| Ischemic Heart Disease | 411 | ICD9CM | 410 | Acute myocardial infarction |
| Ischemic Heart Disease | 411 | ICD10CM | I23.8 | Other current complications following acute myocardial infarction |
| Ischemic Heart Disease | 411 | ICD10CM | I22.1 | Subsequent ST elevation (STEMI) myocardial infarction of inferior wall |
| Ischemic Heart Disease | 411 | ICD9CM | 410.91 | Acute myocardial infarction of unspecified site, initial episode of care |
| Ischemic Heart Disease | 411 | ICD9CM | 410.41 | Acute myocardial infarction of other inferior wall, initial episode of care |
| Ischemic Heart Disease | 411 | ICD9CM | 410 | Acute myocardial infarction of anterolateral wall, episode of care unspecified |
| Ischemic Heart Disease | 411 | ICD10CM | I23.7 | Postinfarction angina |
| Ischemic Heart Disease | 411 | ICD9CM | 410.71 | Subendocardial infarction, initial episode of care |
| Ischemic Heart Disease | 411 | ICD9CM | 429.79 | Certain sequelae of myocardial infarction, not elsewhere classified, other |
| Ischemic Heart Disease | 411 | ICD9CM | 410.62 | True posterior wall infarction, subsequent episode of care |
| Ischemic Heart Disease | 411 | ICD10CM | I51.0 | Cardiac septal defect, acquired |
| Ischemic Heart Disease | 411 | ICD9CM | 410.2 | Acute myocardial infarction, of inferolateral wall |
| Ischemic Heart Disease | 411 | ICD10CM | I23.2 | Ventricular septal defect as current complication following acute myocardial infarction |
| Ischemic Heart Disease | 411 | ICD10CM | I23.0 | Hemopericardium as current complication following acute myocardial infarction |
| Ischemic Heart Disease | 411 | ICD10CM | I21 | Acute myocardial infarction |
| Ischemic Heart Disease | 411 | ICD9CM | 410.32 | Acute myocardial infarction of inferoposterior wall, subsequent episode of care |

| | | | | |
|---|---|---|---|---|
| Ischemic Heart Disease | 411 | ICD9CM | 410.7 | Subendocardial infarction, episode of care unspecified |
| Ischemic Heart Disease | 411 | ICD10CM | I22.8 | Subsequent ST elevation (STEMI) myocardial infarction of other sites |
| Ischemic Heart Disease | 411 | ICD10CM | I22.9 | Subsequent ST elevation (STEMI) myocardial infarction of unspecified site |
| Ischemic Heart Disease | 411 | ICD9CM | 410.72 | Subendocardial infarction, subsequent episode of care |
| Ischemic Heart Disease | 411 | ICD9CM | 410.8 | Acute myocardial infarction of other specified sites, episode of care unspecified |
| Ischemic Heart Disease | 411 | ICD9CM | 410.52 | Acute myocardial infarction of other lateral wall, subsequent episode of care |
| Ischemic Heart Disease | 411 | ICD9CM | 411 | Postmyocardial infarction syndrome |
| Ischemic Heart Disease | 411 | ICD10CM | I21.0 | ST elevation (STEMI) myocardial infarction of anterior wall |
| Ischemic Heart Disease | 411 | ICD10CM | I21.9 | Acute myocardial infarction, unspecified |
| Ischemic Heart Disease | 411 | ICD10CM | I21.A | Other type of myocardial infarction |
| Ischemic Heart Disease | 411 | ICD9CM | 410.6 | Acute myocardial infarction, true posterior wall infarction |
| Ischemic Heart Disease | 411 | ICD9CM | 429.7 | Certain sequelae of myocardial infarction, not elsewhere classified |
| Ischemic Heart Disease | 411 | ICD10CM | I23.3 | Rupture of cardiac wall without hemopericardium as current complication following acute myocardial infarction |
| Ischemic Heart Disease | 411 | ICD10CM | I25.2 | Old myocardial infarction |
| Ischemic Heart Disease | 411 | ICD9CM | 410.9 | Acute myocardial infarction, unspecified site |
| Ischemic Heart Disease | 411 | ICD10CM | I21.A1 | Myocardial infarction type 2 |
| Ischemic Heart Disease | 411 | ICD10CM | I23.1 | Atrial septal defect as current complication following acute myocardial infarction |
| Ischemic Heart Disease | 411 | ICD9CM | 410.61 | True posterior wall infarction, initial episode of care |
| Ischemic Heart Disease | 411 | ICD10CM | I23.6 | Thrombosis of atrium, auricular appendage, and ventricle as current complications following acute myocardial infarction |
| Ischemic Heart Disease | 411 | ICD9CM | 410.11 | Acute myocardial infarction of other anterior wall, initial episode of care |
| Ischemic Heart Disease | 411 | ICD9CM | 410.9 | Acute myocardial infarction of unspecified site, episode of care unspecified |
| Ischemic Heart Disease | 411 | ICD10CM | I22.0 | Subsequent ST elevation (STEMI) myocardial infarction of anterior wall |
| Ischemic Heart Disease | 411 | ICD9CM | 410.22 | Acute myocardial infarction of inferolateral wall, subsequent episode of care |
| Ischemic Heart Disease | 411 | ICD9CM | 410.42 | Acute myocardial infarction of other inferior wall, subsequent episode of care |

| | | | | |
|---|---|---|---|---|
| Ischemic Heart Disease | 411 | ICD10CM | I21.02 | ST elevation (STEMI) myocardial infarction involving left anterior descending coronary artery |
| Ischemic Heart Disease | 411 | ICD9CM | 410.31 | Acute myocardial infarction of inferoposterior wall, initial episode of care |
| Ischemic Heart Disease | 411 | ICD9CM | 410.4 | Acute myocardial infarction, of other inferior wall |
| Ischemic Heart Disease | 411 | ICD10CM | I21.11 | ST elevation (STEMI) myocardial infarction involving right coronary artery |
| Ischemic Heart Disease | 411 | ICD10CM | I21.3 | ST elevation (STEMI) myocardial infarction of unspecified site |
| Ischemic Heart Disease | 411 | ICD9CM | 410.1 | Acute myocardial infarction, of other anterior wall |
| Ischemic Heart Disease | 411 | ICD10CM | I21.2 | ST elevation (STEMI) myocardial infarction of other sites |
| Ischemic Heart Disease | 411 | ICD10CM | I21.1 | ST elevation (STEMI) myocardial infarction of inferior wall |
| Ischemic Heart Disease | 411 | ICD9CM | 410.3 | Acute myocardial infarction of inferoposterior wall, episode of care unspecified |
| Ischemic Heart Disease | 411 | ICD9CM | 429.71 | Acquired cardiac septal defect |
| Ischemic Heart Disease | 411 | ICD9CM | 410.51 | Acute myocardial infarction of other lateral wall, initial episode of care |
| Ischemic Heart Disease | 411 | ICD9CM | 410.21 | Acute myocardial infarction of inferolateral wall, initial episode of care |
| Ischemic Heart Disease | 411 | ICD10CM | I21.4 | Non-ST elevation (NSTEMI) myocardial infarction |
| Ischemic Heart Disease | 411 | ICD9CM | 410.81 | Acute myocardial infarction of other specified sites, initial episode of care |
| Ischemic Heart Disease | 411 | ICD9CM | 410.7 | Acute myocardial infarction, subendocardial infarction |
| Ischemic Heart Disease | 411 | ICD10CM | I21.A9 | Other myocardial infarction type |
| Ischemic Heart Disease | 411 | ICD9CM | 410.92 | Acute myocardial infarction of unspecified site, subsequent episode of care |
| Ischemic Heart Disease | 411 | ICD9CM | 410.4 | Acute myocardial infarction of other inferior wall, episode of care unspecified |
| Ischemic Heart Disease | 411 | ICD10CM | I21.29 | ST elevation (STEMI) myocardial infarction involving other sites |
| Ischemic Heart Disease | 411 | ICD9CM | 410.1 | Acute myocardial infarction of other anterior wall, episode of care unspecified |
| Ischemic Heart Disease | 411 | ICD9CM | 410.2 | Acute myocardial infarction of inferolateral wall, episode of care unspecified |
| Ischemic Heart Disease | 411 | ICD9CM | 412 | Old myocardial infarction |
| Ischemic Heart Disease | 411 | ICD10CM | I21.21 | ST elevation (STEMI) myocardial infarction involving left circumflex coronary artery |
| Ischemic Heart Disease | 411 | ICD10CM | I21.09 | ST elevation (STEMI) myocardial infarction involving other coronary artery of anterior wall |
| Ischemic Heart Disease | 411 | ICD10CM | I24.1 | Dressler's syndrome |

| | | | | |
|---|---|---|---|---|
| Ischemic Heart Disease | 411 | ICD10CM | I25.739 | Atherosclerosis of nonautologous biological coronary artery bypass graft(s) with unspecified angina pectoris |
| Ischemic Heart Disease | 411 | ICD10CM | I25.711 | Atherosclerosis of autologous vein coronary artery bypass graft(s) with angina pectoris with documented spasm |
| Ischemic Heart Disease | 411 | ICD9CM | 413 | Angina decubitus |
| Ischemic Heart Disease | 411 | ICD10CM | I20.8 | Other forms of angina pectoris |
| Ischemic Heart Disease | 411 | ICD10CM | I25.791 | Atherosclerosis of other coronary artery bypass graft(s) with angina pectoris with documented spasm |
| Ischemic Heart Disease | 411 | ICD10CM | I25.84 | Coronary atherosclerosis due to calcified coronary lesion |
| Ischemic Heart Disease | 411 | ICD9CM | 414.3 | Coronary atherosclerosis due to lipid rich plaque |
| Ischemic Heart Disease | 411 | ICD10CM | I25.83 | Coronary atherosclerosis due to lipid rich plaque |
| Ischemic Heart Disease | 411 | ICD10CM | I25.111 | Atherosclerotic heart disease of native coronary artery with angina pectoris with documented spasm |
| Ischemic Heart Disease | 411 | ICD10CM | I25.728 | Atherosclerosis of autologous artery coronary artery bypass graft(s) with other forms of angina pectoris |
| Ischemic Heart Disease | 411 | ICD10CM | I24.0 | Acute coronary thrombosis not resulting in myocardial infarction |
| Ischemic Heart Disease | 411 | ICD10CM | I25.759 | Atherosclerosis of native coronary artery of transplanted heart with unspecified angina pectoris |
| Ischemic Heart Disease | 411 | ICD10CM | I24.9 | Acute ischemic heart disease, unspecified |
| Ischemic Heart Disease | 411 | ICD9CM | 411.8 | Other acute and subacute forms of ischemic heart disease |
| Ischemic Heart Disease | 411 | ICD10CM | I25.758 | Atherosclerosis of native coronary artery of transplanted heart with other forms of angina pectoris |
| Ischemic Heart Disease | 411 | ICD10CM | I25.761 | Atherosclerosis of bypass graft of coronary artery of transplanted heart with angina pectoris with documented spasm |
| Ischemic Heart Disease | 411 | ICD10CM | I25.82 | Chronic total occlusion of coronary artery |
| Ischemic Heart Disease | 411 | ICD10CM | Z95.1 | Presence of aortocoronary bypass graft |
| Ischemic Heart Disease | 411 | ICD9CM | 414.04 | Coronary atherosclerosis of artery bypass graft |
| Ischemic Heart Disease | 411 | ICD10CM | I25.709 | Atherosclerosis of coronary artery bypass graft(s), unspecified, with unspecified angina pectoris |
| Ischemic Heart Disease | 411 | ICD10CM | Z98.61 | Coronary angioplasty status |

| | | | | |
|---|---|---|---|---|
| Ischemic Heart Disease | 411 | ICD9CM | 414.2 | Chronic total occlusion of coronary artery |
| Ischemic Heart Disease | 411 | ICD10CM | I25.738 | Atherosclerosis of nonautologous biological coronary artery bypass graft(s) with other forms of angina pectoris |
| Ischemic Heart Disease | 411 | ICD9CM | 414 | Coronary atherosclerosis |
| Ischemic Heart Disease | 411 | ICD9CM | V45.82 | Percutaneous transluminal coronary angioplasty status |
| Ischemic Heart Disease | 411 | ICD9CM | 414 | Coronary atherosclerosis of unspecified type of vessel, native or graft |
| Ischemic Heart Disease | 411 | ICD10CM | I25.119 | Atherosclerotic heart disease of native coronary artery with unspecified angina pectoris |
| Ischemic Heart Disease | 411 | ICD10CM | I25.1 | Atherosclerotic heart disease of native coronary artery |
| Ischemic Heart Disease | 411 | ICD10CM | I25.810 | Atherosclerosis of coronary artery bypass graft(s) without angina pectoris |
| Ischemic Heart Disease | 411 | ICD10CM | Z95.5 | Presence of coronary angioplasty implant and graft |
| Ischemic Heart Disease | 411 | ICD10CM | I25.118 | Atherosclerotic heart disease of native coronary artery with other forms of angina pectoris |
| Ischemic Heart Disease | 411 | ICD10CM | I25.729 | Atherosclerosis of autologous artery coronary artery bypass graft(s) with unspecified angina pectoris |
| Ischemic Heart Disease | 411 | ICD10CM | I25.42 | Coronary artery dissection |
| Ischemic Heart Disease | 411 | ICD9CM | 414.11 | Aneurysm of coronary vessels |
| Ischemic Heart Disease | 411 | ICD10CM | I25.3 | Aneurysm of heart |
| Ischemic Heart Disease | 411 | ICD9CM | 414.02 | Coronary atherosclerosis of autologous vein bypass graft |
| Ischemic Heart Disease | 411 | ICD10CM | I25.721 | Atherosclerosis of autologous artery coronary artery bypass graft(s) with angina pectoris with documented spasm |
| Ischemic Heart Disease | 411 | ICD10CM | I25.112 | Atherosclerotic heart disease of native coronary artery with refractory angina pectoris |
| Ischemic Heart Disease | 411 | ICD10CM | I25.4 | Coronary artery aneurysm and dissection |
| Ischemic Heart Disease | 411 | ICD10CM | I25.798 | Atherosclerosis of other coronary artery bypass graft(s) with other forms of angina pectoris |
| Ischemic Heart Disease | 411 | ICD10CM | I25.799 | Atherosclerosis of other coronary artery bypass graft(s) with unspecified angina pectoris |
| Ischemic Heart Disease | 411 | ICD10CM | I25.718 | Atherosclerosis of autologous vein coronary artery bypass graft(s) with other forms of angina pectoris |
| Ischemic Heart Disease | 411 | ICD10CM | I20.9 | Angina pectoris, unspecified |
| Ischemic Heart Disease | 411 | ICD10CM | I25.769 | Atherosclerosis of bypass graft of coronary artery of transplanted heart with unspecified angina pectoris |

| | | | | |
|---|---|---|---|---|
| Ischemic Heart Disease | 411 | ICD10CM | I25.701 | Atherosclerosis of coronary artery bypass graft(s), unspecified, with angina pectoris with documented spasm |
| Ischemic Heart Disease | 411 | ICD10CM | I25.768 | Atherosclerosis of bypass graft of coronary artery of transplanted heart with other forms of angina pectoris |
| Ischemic Heart Disease | 411 | ICD9CM | 411.81 | Acute coronary occlusion without myocardial infarction |
| Ischemic Heart Disease | 411 | ICD9CM | 414.01 | Coronary atherosclerosis of native coronary artery |
| Ischemic Heart Disease | 411 | ICD9CM | 414.19 | Other aneurysm of heart |
| Ischemic Heart Disease | 411 | ICD10CM | I25.41 | Coronary artery aneurysm |
| Ischemic Heart Disease | 411 | ICD9CM | 414.1 | Aneurysm of heart (wall) |
| Ischemic Heart Disease | 411 | ICD9CM | 414.1 | Aneurysm and dissection of heart |
| Ischemic Heart Disease | 411 | ICD9CM | 996.03 | Mechanical complication due to coronary bypass graft |
| Ischemic Heart Disease | 411 | ICD9CM | 414.4 | Coronary atherosclerosis due to calcified coronary lesion |
| Ischemic Heart Disease | 411 | ICD10CM | I25.10 | Atherosclerotic heart disease of native coronary artery without angina pectoris |
| Ischemic Heart Disease | 411 | ICD9CM | 414.03 | Coronary atherosclerosis of nonautologous biological bypass graft |
| Ischemic Heart Disease | 411 | ICD10CM | Z98.6 | Angioplasty status |
| Ischemic Heart Disease | 411 | ICD10CM | Z98.62 | Peripheral vascular angioplasty status |
| Ischemic Heart Disease | 411 | ICD9CM | 414.12 | Dissection of coronary artery |
| Ischemic Heart Disease | 411 | ICD10CM | I25 | Chronic ischemic heart disease |
| Ischemic Heart Disease | 411 | ICD10CM | I20.2 | Refractory angina pectoris |
| Ischemic Heart Disease | 411 | ICD9CM | 413.1 | Prinzmetal angina |
| Ischemic Heart Disease | 411 | ICD10CM | I25.752 | Atherosclerosis of native coronary artery of transplanted heart with refractory angina pectoris |
| Ischemic Heart Disease | 411 | ICD10CM | I25.702 | Atherosclerosis of coronary artery bypass graft(s), unspecified, with refractory angina pectoris |
| Ischemic Heart Disease | 411 | ICD9CM | 414.8 | Other specified forms of chronic ischemic heart disease |
| Ischemic Heart Disease | 411 | ICD9CM | V45.81 | Aortocoronary bypass status |
| Ischemic Heart Disease | 411 | ICD10CM | I25.792 | Atherosclerosis of other coronary artery bypass graft(s) with refractory angina pectoris |

| | | | | |
|---|---|---|---|---|
| Ischemic Heart Disease | 411 | ICD10CM | I25.9 | Chronic ischemic heart disease, unspecified |
| Ischemic Heart Disease | 411 | ICD9CM | 414.05 | Coronary atherosclerosis of unspecified bypass graft |
| Ischemic Heart Disease | 411 | ICD10CM | I25.712 | Atherosclerosis of autologous vein coronary artery bypass graft(s) with refractory angina pectoris |
| Ischemic Heart Disease | 411 | ICD10CM | I25.89 | Other forms of chronic ischemic heart disease |
| Ischemic Heart Disease | 411 | ICD10CM | I25.722 | Atherosclerosis of autologous artery coronary artery bypass graft(s) with refractory angina pectoris |
| Ischemic Heart Disease | 411 | ICD10CM | I25.8 | Other forms of chronic ischemic heart disease |
| Ischemic Heart Disease | 411 | ICD10CM | I25.85 | Chronic coronary microvascular dysfunction |
| Ischemic Heart Disease | 411 | ICD10CM | I25.5 | Ischemic cardiomyopathy |
| Ischemic Heart Disease | 411 | ICD10CM | I25.6 | Silent myocardial ischemia |
| Ischemic Heart Disease | 411 | ICD9CM | 414.9 | Chronic ischemic heart disease, unspecified |
| Ischemic Heart Disease | 411 | ICD10CM | I25.732 | Atherosclerosis of nonautologous biological coronary artery bypass graft(s) with refractory angina pectoris |
| Ischemic Heart Disease | 411 | ICD9CM | 414 | Other forms of chronic ischemic heart disease |
| Ischemic Heart Disease | 411 | ICD10CM | I25.762 | Atherosclerosis of bypass graft of coronary artery of transplanted heart with refractory angina pectoris |
| Myocardial infarction | 411.2 | ICD9CM | 410.7 | Subendocardial infarction, episode of care unspecified |
| Myocardial infarction | 411.2 | ICD10CM | I22.8 | Subsequent ST elevation (STEMI) myocardial infarction of other sites |
| Myocardial infarction | 411.2 | ICD10CM | I21.A | Other type of myocardial infarction |
| Myocardial infarction | 411.2 | ICD9CM | 410.32 | Acute myocardial infarction of inferoposterior wall, subsequent episode of care |
| Myocardial infarction | 411.2 | ICD10CM | I21.0 | ST elevation (STEMI) myocardial infarction of anterior wall |
| Myocardial infarction | 411.2 | ICD10CM | I21.9 | Acute myocardial infarction, unspecified |
| Myocardial infarction | 411.2 | ICD9CM | 410.3 | Acute myocardial infarction, of inferoposterior wall |
| Myocardial infarction | 411.2 | ICD9CM | 411 | Postmyocardial infarction syndrome |
| Myocardial infarction | 411.2 | ICD9CM | 410.12 | Acute myocardial infarction of other anterior wall, subsequent episode of care |
| Myocardial infarction | 411.2 | ICD9CM | 410.8 | Acute myocardial infarction, of other specified sites |
| Myocardial infarction | 411.2 | ICD9CM | 410.72 | Subendocardial infarction, subsequent episode of care |

| | | | | |
|---|---|---|---|---|
| Myocardial infarction | 411.2 | ICD9CM | 410.8 | Acute myocardial infarction of other specified sites, episode of care unspecified |
| Myocardial infarction | 411.2 | ICD9CM | 410.52 | Acute myocardial infarction of other lateral wall, subsequent episode of care |
| Myocardial infarction | 411.2 | ICD9CM | 410.82 | Acute myocardial infarction of other specified sites, subsequent episode of care |
| Myocardial infarction | 411.2 | ICD10CM | I22.2 | Subsequent non-ST elevation (NSTEMI) myocardial infarction |
| Myocardial infarction | 411.2 | ICD10CM | I23.0 | Hemopericardium as current complication following acute myocardial infarction |
| Myocardial infarction | 411.2 | ICD10CM | I21 | Acute myocardial infarction |
| Myocardial infarction | 411.2 | ICD9CM | 410 | Acute myocardial infarction, of anterolateral wall |
| Myocardial infarction | 411.2 | ICD9CM | 410.5 | Acute myocardial infarction of other lateral wall, episode of care unspecified |
| Myocardial infarction | 411.2 | ICD9CM | 410.01 | Acute myocardial infarction of anterolateral wall, initial episode of care |
| Myocardial infarction | 411.2 | ICD10CM | I22.9 | Subsequent ST elevation (STEMI) myocardial infarction of unspecified site |
| Myocardial infarction | 411.2 | ICD9CM | 410.5 | Acute myocardial infarction, of other lateral wall |
| Myocardial infarction | 411.2 | ICD9CM | 410.02 | Acute myocardial infarction of anterolateral wall, subsequent episode of care |
| Myocardial infarction | 411.2 | ICD10CM | I21.01 | ST elevation (STEMI) myocardial infarction involving left main coronary artery |
| Myocardial infarction | 411.2 | ICD9CM | 410.51 | Acute myocardial infarction of other lateral wall, initial episode of care |
| Myocardial infarction | 411.2 | ICD9CM | 410.21 | Acute myocardial infarction of inferolateral wall, initial episode of care |
| Myocardial infarction | 411.2 | ICD10CM | I21.4 | Non-ST elevation (NSTEMI) myocardial infarction |
| Myocardial infarction | 411.2 | ICD9CM | 410.81 | Acute myocardial infarction of other specified sites, initial episode of care |
| Myocardial infarction | 411.2 | ICD10CM | I21.3 | ST elevation (STEMI) myocardial infarction of unspecified site |
| Myocardial infarction | 411.2 | ICD9CM | 410.1 | Acute myocardial infarction, of other anterior wall |
| Myocardial infarction | 411.2 | ICD10CM | I21.19 | ST elevation (STEMI) myocardial infarction involving other coronary artery of inferior wall |
| Myocardial infarction | 411.2 | ICD10CM | I21.1 | ST elevation (STEMI) myocardial infarction of inferior wall |
| Myocardial infarction | 411.2 | ICD9CM | 410.3 | Acute myocardial infarction of inferoposterior wall, episode of care unspecified |
| Myocardial infarction | 411.2 | ICD9CM | 429.71 | Acquired cardiac septal defect |
| Myocardial infarction | 411.2 | ICD10CM | I51.3 | Intracardiac thrombosis, not elsewhere classified |
| Myocardial infarction | 411.2 | ICD10CM | I23.8 | Other current complications following acute myocardial infarction |
| Myocardial infarction | 411.2 | ICD10CM | I22.1 | Subsequent ST elevation (STEMI) myocardial infarction of inferior wall |

| | | | | |
|---|---|---|---|---|
| Myocardial infarction | 411.2 | ICD9CM | 410.91 | Acute myocardial infarction of unspecified site, initial episode of care |
| Myocardial infarction | 411.2 | ICD9CM | 410.41 | Acute myocardial infarction of other inferior wall, initial episode of care |
| Myocardial infarction | 411.2 | ICD9CM | 410.7 | Acute myocardial infarction, subendocardial infarction |
| Myocardial infarction | 411.2 | ICD10CM | I21.A9 | Other myocardial infarction type |
| Myocardial infarction | 411.2 | ICD9CM | 410.92 | Acute myocardial infarction of unspecified site, subsequent episode of care |
| Myocardial infarction | 411.2 | ICD10CM | I21.2 | ST elevation (STEMI) myocardial infarction of other sites |
| Myocardial infarction | 411.2 | ICD9CM | 429.79 | Certain sequelae of myocardial infarction, not elsewhere classified, other |
| Myocardial infarction | 411.2 | ICD9CM | 410.62 | True posterior wall infarction, subsequent episode of care |
| Myocardial infarction | 411.2 | ICD10CM | I51.0 | Cardiac septal defect, acquired |
| Myocardial infarction | 411.2 | ICD9CM | 410.2 | Acute myocardial infarction, of inferolateral wall |
| Myocardial infarction | 411.2 | ICD10CM | I23.2 | Ventricular septal defect as current complication following acute myocardial infarction |
| Myocardial infarction | 411.2 | ICD9CM | 410.4 | Acute myocardial infarction, of other inferior wall |
| Myocardial infarction | 411.2 | ICD10CM | I21.11 | ST elevation (STEMI) myocardial infarction involving right coronary artery |
| Myocardial infarction | 411.2 | ICD9CM | 410 | Acute myocardial infarction of anterolateral wall, episode of care unspecified |
| Myocardial infarction | 411.2 | ICD10CM | I23.7 | Postinfarction angina |
| Myocardial infarction | 411.2 | ICD9CM | 410.71 | Subendocardial infarction, initial episode of care |
| Myocardial infarction | 411.2 | ICD9CM | 410.6 | Acute myocardial infarction, true posterior wall infarction |
| Myocardial infarction | 411.2 | ICD9CM | 429.7 | Certain sequelae of myocardial infarction, not elsewhere classified |
| Myocardial infarction | 411.2 | ICD10CM | I23.3 | Rupture of cardiac wall without hemopericardium as current complication following acute myocardial infarction |
| Myocardial infarction | 411.2 | ICD10CM | I25.2 | Old myocardial infarction |
| Myocardial infarction | 411.2 | ICD9CM | 410 | Acute myocardial infarction |
| Myocardial infarction | 411.2 | ICD9CM | 410.9 | Acute myocardial infarction, unspecified site |
| Myocardial infarction | 411.2 | ICD10CM | I21.A1 | Myocardial infarction type 2 |
| Myocardial infarction | 411.2 | ICD10CM | I23.1 | Atrial septal defect as current complication following acute myocardial infarction |
| Myocardial infarction | 411.2 | ICD10CM | I24.1 | Dressler's syndrome |
| Myocardial infarction | 411.2 | ICD9CM | 410.61 | True posterior wall infarction, initial episode of care |
| Myocardial infarction | 411.2 | ICD10CM | I23.6 | Thrombosis of atrium, auricular appendage, and ventricle as current complications following acute myocardial infarction |

| | | | | |
|---|---|---|---|---|
| Myocardial infarction | 411.2 | ICD9CM | 410.9 | Acute myocardial infarction of unspecified site, episode of care unspecified |
| Myocardial infarction | 411.2 | ICD10CM | I22.0 | Subsequent ST elevation (STEMI) myocardial infarction of anterior wall |
| Myocardial infarction | 411.2 | ICD9CM | 410.22 | Acute myocardial infarction of inferolateral wall, subsequent episode of care |
| Myocardial infarction | 411.2 | ICD9CM | 410.42 | Acute myocardial infarction of other inferior wall, subsequent episode of care |
| Myocardial infarction | 411.2 | ICD10CM | I21.02 | ST elevation (STEMI) myocardial infarction involving left anterior descending coronary artery |
| Myocardial infarction | 411.2 | ICD9CM | 410.31 | Acute myocardial infarction of inferoposterior wall, initial episode of care |
| Myocardial infarction | 411.2 | ICD10CM | I21.09 | ST elevation (STEMI) myocardial infarction involving other coronary artery of anterior wall |
| Myocardial infarction | 411.2 | ICD9CM | 410.11 | Acute myocardial infarction of other anterior wall, initial episode of care |
| Myocardial infarction | 411.2 | ICD9CM | 410.4 | Acute myocardial infarction of other inferior wall, episode of care unspecified |
| Myocardial infarction | 411.2 | ICD10CM | I22 | Subsequent ST elevation (STEMI) and non-ST elevation (NSTEMI) myocardial infarction |
| Myocardial infarction | 411.2 | ICD10CM | I21.B | Myocardial infarction with coronary microvascular dysfunction |
| Myocardial infarction | 411.2 | ICD9CM | 410.6 | True posterior wall infarction, episode of care unspecified |
| Myocardial infarction | 411.2 | ICD10CM | I21.21 | ST elevation (STEMI) myocardial infarction involving left circumflex coronary artery |
| Myocardial infarction | 411.2 | ICD10CM | I21.29 | ST elevation (STEMI) myocardial infarction involving other sites |
| Myocardial infarction | 411.2 | ICD9CM | 410.2 | Acute myocardial infarction of inferolateral wall, episode of care unspecified |
| Myocardial infarction | 411.2 | ICD9CM | 412 | Old myocardial infarction |
| Myocardial infarction | 411.2 | ICD9CM | 410.1 | Acute myocardial infarction of other anterior wall, episode of care unspecified |
| Atrial fibrillation | 427.21 | ICD10CM | I48.0 | Paroxysmal atrial fibrillation |
| Atrial fibrillation | 427.21 | ICD9CM | 427.31 | Atrial fibrillation |
| Atrial fibrillation | 427.21 | ICD10CM | I48.21 | Permanent atrial fibrillation |
| Atrial fibrillation | 427.21 | ICD10CM | I48.2 | Chronic atrial fibrillation |
| Atrial fibrillation | 427.21 | ICD10CM | I48.91 | Unspecified atrial fibrillation |
| Atrial fibrillation | 427.21 | ICD10CM | I48.20 | Chronic atrial fibrillation, unspecified |
| Heart failure NOS | 428.2 | ICD9CM | 428.9 | Heart failure, unspecified |
| Heart failure NOS | 428.2 | ICD10CM | I50.1 | Left ventricular failure, unspecified |
| Heart failure NOS | 428.2 | ICD10CM | I50.811 | Acute right heart failure |
| Heart failure NOS | 428.2 | ICD10CM | I50.814 | Right heart failure due to left heart failure |
| Heart failure NOS | 428.2 | ICD10CM | I50.810 | Right heart failure, unspecified |
| Heart failure NOS | 428.2 | ICD10CM | I50.82 | Biventricular heart failure |

| | | | | |
|---|---|---|---|---|
| Heart failure NOS | 428.2 | ICD10CM | I50.813 | Acute on chronic right heart failure |
| Heart failure NOS | 428.2 | ICD9CM | 428.1 | Left heart failure |
| Heart failure NOS | 428.2 | ICD10CM | I50.89 | Other heart failure |
| Heart failure NOS | 428.2 | ICD10CM | I50 | Heart failure |
| Heart failure NOS | 428.2 | ICD10CM | I50.83 | High output heart failure |
| Heart failure NOS | 428.2 | ICD10CM | I50.812 | Chronic right heart failure |
| Heart failure NOS | 428.2 | ICD10CM | I50.8 | Other heart failure |
| Heart failure NOS | 428.2 | ICD10CM | I50.84 | End stage heart failure |
| Heart failure NOS | 428.2 | ICD10CM | I50.81 | Right heart failure |
| Intracranial hemorrhage | 430 | ICD9CM | 432 | Nontraumatic extradural hemorrhage |
| Intracranial hemorrhage | 430 | ICD10CM | I62.1 | Nontraumatic extradural hemorrhage |
| Intracranial hemorrhage | 430 | ICD9CM | 432.9 | Unspecified intracranial hemorrhage |
| Intracranial hemorrhage | 430 | ICD9CM | 432 | Other and unspecified intracranial hemorrhage |
| Intracranial hemorrhage | 430 | ICD10CM | I62.9 | Nontraumatic intracranial hemorrhage, unspecified |
| Intracranial hemorrhage | 430 | ICD10CM | I60.1 | Nontraumatic subarachnoid hemorrhage from middle cerebral artery |
| Intracranial hemorrhage | 430 | ICD10CM | I60.0 | Nontraumatic subarachnoid hemorrhage from carotid siphon and bifurcation |
| Intracranial hemorrhage | 430 | ICD10CM | I60.01 | Nontraumatic subarachnoid hemorrhage from right carotid siphon and bifurcation |
| Intracranial hemorrhage | 430 | ICD10CM | I60.11 | Nontraumatic subarachnoid hemorrhage from right middle cerebral artery |
| Intracranial hemorrhage | 430 | ICD10CM | I60.31 | Nontraumatic subarachnoid hemorrhage from right posterior communicating artery |
| Intracranial hemorrhage | 430 | ICD10CM | I60.50 | Nontraumatic subarachnoid hemorrhage from unspecified vertebral artery |
| Intracranial hemorrhage | 430 | ICD9CM | 430 | Subarachnoid hemorrhage * |
| Intracranial hemorrhage | 430 | ICD10CM | I60.7 | Nontraumatic subarachnoid hemorrhage from unspecified intracranial artery |
| Intracranial hemorrhage | 430 | ICD9CM | 430 | Subarachnoid hemorrhage |
| Intracranial hemorrhage | 430 | ICD10CM | I60.30 | Nontraumatic subarachnoid hemorrhage from unspecified posterior communicating artery |
| Intracranial hemorrhage | 430 | ICD10CM | I60.12 | Nontraumatic subarachnoid hemorrhage from left middle cerebral artery |
| Intracranial hemorrhage | 430 | ICD10CM | I60.10 | Nontraumatic subarachnoid hemorrhage from unspecified middle cerebral artery |
| Intracranial hemorrhage | 430 | ICD10CM | I60.9 | Nontraumatic subarachnoid hemorrhage, unspecified |
| Intracranial hemorrhage | 430 | ICD10CM | I60.2 | Nontraumatic subarachnoid hemorrhage from anterior communicating artery |
| Intracranial hemorrhage | 430 | ICD10CM | I60.52 | Nontraumatic subarachnoid hemorrhage from left vertebral artery |

| | | | | |
|---|---|---|---|---|
| Intracranial hemorrhage | 430 | ICD10CM | I60.5 | Nontraumatic subarachnoid hemorrhage from vertebral artery |
| Intracranial hemorrhage | 430 | ICD10CM | I60.32 | Nontraumatic subarachnoid hemorrhage from left posterior communicating artery |
| Intracranial hemorrhage | 430 | ICD10CM | I60.4 | Nontraumatic subarachnoid hemorrhage from basilar artery |
| Intracranial hemorrhage | 430 | ICD10CM | I60.00 | Nontraumatic subarachnoid hemorrhage from unspecified carotid siphon and bifurcation |
| Intracranial hemorrhage | 430 | ICD10CM | I60.51 | Nontraumatic subarachnoid hemorrhage from right vertebral artery |
| Intracranial hemorrhage | 430 | ICD10CM | I60.6 | Nontraumatic subarachnoid hemorrhage from other intracranial arteries |
| Intracranial hemorrhage | 430 | ICD10CM | I60.8 | Other nontraumatic subarachnoid hemorrhage |
| Intracranial hemorrhage | 430 | ICD10CM | I60.02 | Nontraumatic subarachnoid hemorrhage from left carotid siphon and bifurcation |
| Intracranial hemorrhage | 430 | ICD10CM | I60.3 | Nontraumatic subarachnoid hemorrhage from posterior communicating artery |
| Intracranial hemorrhage | 430 | ICD9CM | 431 | Intracerebral hemorrhage * |
| Intracranial hemorrhage | 430 | ICD10CM | I61.9 | Nontraumatic intracerebral hemorrhage, unspecified |
| Intracranial hemorrhage | 430 | ICD10CM | I62.0 | Nontraumatic subdural hemorrhage |
| Intracranial hemorrhage | 430 | ICD10CM | I62.00 | Nontraumatic subdural hemorrhage, unspecified |
| Intracranial hemorrhage | 430 | ICD10CM | I62.01 | Nontraumatic acute subdural hemorrhage |
| Intracranial hemorrhage | 430 | ICD9CM | 432.1 | Subdural hemorrhage |
| Intracranial hemorrhage | 430 | ICD10CM | I62.03 | Nontraumatic chronic subdural hemorrhage |
| Intracranial hemorrhage | 430 | ICD10CM | I62.02 | Nontraumatic subacute subdural hemorrhage |
| Intracranial hemorrhage | 430 | ICD10CM | I61.4 | Nontraumatic intracerebral hemorrhage in cerebellum |
| Intracranial hemorrhage | 430 | ICD10CM | I61.2 | Nontraumatic intracerebral hemorrhage in hemisphere, unspecified |
| Intracranial hemorrhage | 430 | ICD10CM | I61.0 | Nontraumatic intracerebral hemorrhage in hemisphere, subcortical |
| Intracranial hemorrhage | 430 | ICD10CM | I61 | Nontraumatic intracerebral hemorrhage |
| Intracranial hemorrhage | 430 | ICD10CM | I61.3 | Nontraumatic intracerebral hemorrhage in brain stem |
| Intracranial hemorrhage | 430 | ICD10CM | I61.5 | Nontraumatic intracerebral hemorrhage, intraventricular |
| Intracranial hemorrhage | 430 | ICD10CM | I61.6 | Nontraumatic intracerebral hemorrhage, multiple localized |
| Intracranial hemorrhage | 430 | ICD10CM | I61.1 | Nontraumatic intracerebral hemorrhage in hemisphere, cortical |

| | | | | |
|---|---|---|---|---|
| Intracranial hemorrhage | 430 | ICD9CM | 431 | Intracerebral hemorrhage |
| Intracranial hemorrhage | 430 | ICD10CM | I61.8 | Other nontraumatic intracerebral hemorrhage |
| Subarachnoid hemorrhage | 430.1 | ICD10CM | I60.0 | Nontraumatic subarachnoid hemorrhage from carotid siphon and bifurcation |
| Subarachnoid hemorrhage | 430.1 | ICD10CM | I60.12 | Nontraumatic subarachnoid hemorrhage from left middle cerebral artery |
| Subarachnoid hemorrhage | 430.1 | ICD10CM | I60.10 | Nontraumatic subarachnoid hemorrhage from unspecified middle cerebral artery |
| Subarachnoid hemorrhage | 430.1 | ICD10CM | I60.9 | Nontraumatic subarachnoid hemorrhage, unspecified |
| Subarachnoid hemorrhage | 430.1 | ICD10CM | I60.2 | Nontraumatic subarachnoid hemorrhage from anterior communicating artery |
| Subarachnoid hemorrhage | 430.1 | ICD10CM | I60.01 | Nontraumatic subarachnoid hemorrhage from right carotid siphon and bifurcation |
| Subarachnoid hemorrhage | 430.1 | ICD10CM | I60.11 | Nontraumatic subarachnoid hemorrhage from right middle cerebral artery |
| Subarachnoid hemorrhage | 430.1 | ICD10CM | I60.31 | Nontraumatic subarachnoid hemorrhage from right posterior communicating artery |
| Subarachnoid hemorrhage | 430.1 | ICD10CM | I60.50 | Nontraumatic subarachnoid hemorrhage from unspecified vertebral artery |
| Subarachnoid hemorrhage | 430.1 | ICD9CM | 430 | Subarachnoid hemorrhage * |
| Subarachnoid hemorrhage | 430.1 | ICD10CM | I60.7 | Nontraumatic subarachnoid hemorrhage from unspecified intracranial artery |
| Subarachnoid hemorrhage | 430.1 | ICD9CM | 430 | Subarachnoid hemorrhage |
| Subarachnoid hemorrhage | 430.1 | ICD10CM | I60.30 | Nontraumatic subarachnoid hemorrhage from unspecified posterior communicating artery |
| Subarachnoid hemorrhage | 430.1 | ICD10CM | I60.8 | Other nontraumatic subarachnoid hemorrhage |
| Subarachnoid hemorrhage | 430.1 | ICD10CM | I60.02 | Nontraumatic subarachnoid hemorrhage from left carotid siphon and bifurcation |
| Subarachnoid hemorrhage | 430.1 | ICD10CM | I60.3 | Nontraumatic subarachnoid hemorrhage from posterior communicating artery |
| Subarachnoid hemorrhage | 430.1 | ICD10CM | I60.1 | Nontraumatic subarachnoid hemorrhage from middle cerebral artery |
| Subarachnoid hemorrhage | 430.1 | ICD10CM | I60.32 | Nontraumatic subarachnoid hemorrhage from left posterior communicating artery |
| Subarachnoid hemorrhage | 430.1 | ICD10CM | I60.52 | Nontraumatic subarachnoid hemorrhage from left vertebral artery |
| Subarachnoid hemorrhage | 430.1 | ICD10CM | I60.5 | Nontraumatic subarachnoid hemorrhage from vertebral artery |
| Subarachnoid hemorrhage | 430.1 | ICD10CM | I60.4 | Nontraumatic subarachnoid hemorrhage from basilar artery |
| Subarachnoid hemorrhage | 430.1 | ICD10CM | I60.00 | Nontraumatic subarachnoid hemorrhage from unspecified carotid siphon and bifurcation |
| Subarachnoid hemorrhage | 430.1 | ICD10CM | I60.51 | Nontraumatic subarachnoid hemorrhage from right vertebral artery |

| | | | | |
|---|---|---|---|---|
| Subarachnoid hemorrhage | 430.1 | ICD10CM | I60.6 | Nontraumatic subarachnoid hemorrhage from other intracranial arteries |
| Cerebrovascular disease | 433 | ICD9CM | 437.4 | Cerebral arteritis |
| Cerebrovascular disease | 433 | ICD10CM | G46.3 | Brain stem stroke syndrome |
| Cerebrovascular disease | 433 | ICD10CM | I68 | Cerebrovascular disorders in diseases classified elsewhere |
| Cerebrovascular disease | 433 | ICD10CM | G46.5 | Pure motor lacunar syndrome |
| Cerebrovascular disease | 433 | ICD9CM | 437.7 | Transient global amnesia |
| Cerebrovascular disease | 433 | ICD10CM | I68.8 | Other cerebrovascular disorders in diseases classified elsewhere |
| Cerebrovascular disease | 433 | ICD10CM | G46.6 | Pure sensory lacunar syndrome |
| Cerebrovascular disease | 433 | ICD10CM | G46.8 | Other vascular syndromes of brain in cerebrovascular diseases |
| Cerebrovascular disease | 433 | ICD10CM | G46.4 | Cerebellar stroke syndrome |
| Cerebrovascular disease | 433 | ICD9CM | 437 | Other and ill-defined cerebrovascular disease |
| Cerebrovascular disease | 433 | ICD10CM | G45.4 | Transient global amnesia |
| Cerebrovascular disease | 433 | ICD10CM | I67.9 | Cerebrovascular disease, unspecified |
| Cerebrovascular disease | 433 | ICD10CM | I67.7 | Cerebral arteritis, not elsewhere classified |
| Cerebrovascular disease | 433 | ICD10CM | I68.2 | Cerebral arteritis in other diseases classified elsewhere |
| Cerebrovascular disease | 433 | ICD9CM | 437.8 | Other ill-defined cerebrovascular disease |
| Cerebrovascular disease | 433 | ICD9CM | 437.9 | Unspecified cerebrovascular disease |
| Cerebrovascular disease | 433 | ICD10CM | G46.7 | Other lacunar syndromes |
| Cerebrovascular disease | 433 | ICD10CM | I68.0 | Cerebral amyloid angiopathy |
| Cerebrovascular disease | 433 | ICD10CM | I65.03 | Occlusion and stenosis of bilateral vertebral arteries |
| Cerebrovascular disease | 433 | ICD10CM | I65.01 | Occlusion and stenosis of right vertebral artery |
| Cerebrovascular disease | 433 | ICD10CM | I65.23 | Occlusion and stenosis of bilateral carotid arteries |
| Cerebrovascular disease | 433 | ICD9CM | 433.1 | Occlusion and stenosis of carotid artery without mention of cerebral infarction |
| Cerebrovascular disease | 433 | ICD9CM | 433.8 | Occlusion and stenosis of other specified precerebral artery |
| Cerebrovascular disease | 433 | ICD10CM | I65.21 | Occlusion and stenosis of right carotid artery |

| | | | | |
|---|---|---|---|---|
| Cerebrovascular disease | 433 | ICD9CM | 433 | Occlusion and stenosis of precerebral arteries |
| Cerebrovascular disease | 433 | ICD9CM | 433.1 | Occlusion and stenosis of carotid artery |
| Cerebrovascular disease | 433 | ICD10CM | I65.0 | Occlusion and stenosis of vertebral artery |
| Cerebrovascular disease | 433 | ICD9CM | 433.9 | Occlusion and stenosis of unspecified precerebral artery without mention of cerebral infarction |
| Cerebrovascular disease | 433 | ICD10CM | I65.29 | Occlusion and stenosis of unspecified carotid artery |
| Cerebrovascular disease | 433 | ICD9CM | 433.3 | Occlusion and stenosis of multiple and bilateral precerebral arteries without mention of cerebral infarction |
| Cerebrovascular disease | 433 | ICD9CM | 433.9 | Occlusion and stenosis of unspecified precerebral artery |
| Cerebrovascular disease | 433 | ICD9CM | 433.8 | Occlusion and stenosis of other specified precerebral artery without mention of cerebral infarction |
| Cerebrovascular disease | 433 | ICD10CM | I65 | Occlusion and stenosis of precerebral arteries, not resulting in cerebral infarction |
| Cerebrovascular disease | 433 | ICD10CM | I65.02 | Occlusion and stenosis of left vertebral artery |
| Cerebrovascular disease | 433 | ICD9CM | 433 | Occlusion and stenosis of basilar artery without mention of cerebral infarction |
| Cerebrovascular disease | 433 | ICD9CM | 433.2 | Occlusion and stenosis of vertebral artery |
| Cerebrovascular disease | 433 | ICD10CM | I65.8 | Occlusion and stenosis of other precerebral arteries |
| Cerebrovascular disease | 433 | ICD9CM | 433.3 | Occlusion and stenosis of multiple and bilateral precerebral arteries |
| Cerebrovascular disease | 433 | ICD9CM | 433.2 | Occlusion and stenosis of vertebral artery without mention of cerebral infarction |
| Cerebrovascular disease | 433 | ICD10CM | I65.9 | Occlusion and stenosis of unspecified precerebral artery |
| Cerebrovascular disease | 433 | ICD10CM | I65.1 | Occlusion and stenosis of basilar artery |
| Cerebrovascular disease | 433 | ICD10CM | I65.2 | Occlusion and stenosis of carotid artery |
| Cerebrovascular disease | 433 | ICD9CM | 433 | Occlusion and stenosis of basilar artery |
| Cerebrovascular disease | 433 | ICD10CM | I65.09 | Occlusion and stenosis of unspecified vertebral artery |
| Cerebrovascular disease | 433 | ICD10CM | I65.22 | Occlusion and stenosis of left carotid artery |
| Cerebrovascular disease | 433 | ICD10CM | I69.815 | Cognitive social or emotional deficit following other cerebrovascular disease |
| Cerebrovascular disease | 433 | ICD9CM | 438.2 | Hemiplegia/hemiparesis as late effect of cerebrovascular disease |
| Cerebrovascular disease | 433 | ICD9CM | 438.81 | Other late effects of cerebrovascular disease, apraxia |

| | | | | |
|---|---|---|---|---|
| Cerebrovascular disease | 433 | ICD10CM | I69.21 | Cognitive deficits following other nontraumatic intracranial hemorrhage |
| Cerebrovascular disease | 433 | ICD10CM | I69.821 | Dysphasia following other cerebrovascular disease |
| Cerebrovascular disease | 433 | ICD10CM | I69.152 | Hemiplegia and hemiparesis following nontraumatic intracerebral hemorrhage affecting left dominant side |
| Cerebrovascular disease | 433 | ICD10CM | I69.251 | Hemiplegia and hemiparesis following other nontraumatic intracranial hemorrhage affecting right dominant side |
| Cerebrovascular disease | 433 | ICD10CM | I69.169 | Other paralytic syndrome following nontraumatic intracerebral hemorrhage affecting unspecified side |
| Cerebrovascular disease | 433 | ICD10CM | I69.132 | Monoplegia of upper limb following nontraumatic intracerebral hemorrhage affecting left dominant side |
| Cerebrovascular disease | 433 | ICD10CM | I69.021 | Dysphasia following nontraumatic subarachnoid hemorrhage |
| Cerebrovascular disease | 433 | ICD9CM | 438 | Late effects of cerebrovascular disease |
| Cerebrovascular disease | 433 | ICD9CM | 438.1 | Speech and language deficits as late effect of cerebrovascular disease |
| Cerebrovascular disease | 433 | ICD10CM | I69.951 | Hemiplegia and hemiparesis following unspecified cerebrovascular disease affecting right dominant side |
| Cerebrovascular disease | 433 | ICD10CM | I69.342 | Monoplegia of lower limb following cerebral infarction affecting left dominant side |
| Cerebrovascular disease | 433 | ICD9CM | 438.85 | Other late effects of cerebrovascular disease, vertigo |
| Cerebrovascular disease | 433 | ICD10CM | I69.318 | Other symptoms and signs involving cognitive functions following cerebral infarction |
| Cerebrovascular disease | 433 | ICD10CM | I69.83 | Monoplegia of upper limb following other cerebrovascular disease |
| Cerebrovascular disease | 433 | ICD10CM | I69.392 | Facial weakness following cerebral infarction |
| Cerebrovascular disease | 433 | ICD10CM | I69.214 | Frontal lobe and executive function deficit following other nontraumatic intracranial hemorrhage |
| Cerebrovascular disease | 433 | ICD10CM | I69.8 | Sequelae of other cerebrovascular diseases |
| Cerebrovascular disease | 433 | ICD10CM | I69.86 | Other paralytic syndrome following other cerebrovascular disease |
| Cerebrovascular disease | 433 | ICD10CM | I69.292 | Facial weakness following other nontraumatic intracranial hemorrhage |
| Cerebrovascular disease | 433 | ICD10CM | I69.811 | Memory deficit following other cerebrovascular disease |
| Cerebrovascular disease | 433 | ICD10CM | I69.810 | Attention and concentration deficit following other cerebrovascular disease |
| Cerebrovascular disease | 433 | ICD10CM | I69.015 | Cognitive social or emotional deficit following nontraumatic subarachnoid hemorrhage |

| | | | | |
|---|---|---|---|---|
| Cerebrovascular disease | 433 | ICD10CM | I69.111 | Memory deficit following nontraumatic intracerebral hemorrhage |
| Cerebrovascular disease | 433 | ICD10CM | I69.03 | Monoplegia of upper limb following nontraumatic subarachnoid hemorrhage |
| Cerebrovascular disease | 433 | ICD10CM | I69.028 | Other speech and language deficits following nontraumatic subarachnoid hemorrhage |
| Cerebrovascular disease | 433 | ICD10CM | I69.165 | Other paralytic syndrome following nontraumatic intracerebral hemorrhage, bilateral |
| Cerebrovascular disease | 433 | ICD9CM | 438.14 | Late effects of cerebrovascular disease, fluency disorder |
| Cerebrovascular disease | 433 | ICD10CM | I69.211 | Memory deficit following other nontraumatic intracranial hemorrhage |
| Cerebrovascular disease | 433 | ICD9CM | 433.31 | Occlusion and stenosis of multiple and bilateral precerebral arteries with cerebral infarction |
| Cerebrovascular disease | 433 | ICD10CM | I63.10 | Cerebral infarction due to embolism of unspecified precerebral artery |
| Cerebrovascular disease | 433 | ICD10CM | I63.2 | Cerebral infarction due to unspecified occlusion or stenosis of precerebral arteries |
| Cerebrovascular disease | 433 | ICD10CM | I63.211 | Cerebral infarction due to unspecified occlusion or stenosis of right vertebral artery |
| Cerebrovascular disease | 433 | ICD10CM | I63.112 | Cerebral infarction due to embolism of left vertebral artery |
| Cerebrovascular disease | 433 | ICD10CM | I63.019 | Cerebral infarction due to thrombosis of unspecified vertebral artery |
| Cerebrovascular disease | 433 | ICD10CM | I63.29 | Cerebral infarction due to unspecified occlusion or stenosis of other precerebral arteries |
| Cerebrovascular disease | 433 | ICD10CM | I63.012 | Cerebral infarction due to thrombosis of left vertebral artery |
| Cerebrovascular disease | 433 | ICD10CM | I63.13 | Cerebral infarction due to embolism of carotid artery |
| Cerebrovascular disease | 433 | ICD10CM | I63.232 | Cerebral infarction due to unspecified occlusion or stenosis of left carotid arteries |
| Cerebrovascular disease | 433 | ICD10CM | I63.113 | Cerebral infarction due to embolism of bilateral vertebral arteries |
| Cerebrovascular disease | 433 | ICD9CM | 433.21 | Occlusion and stenosis of vertebral artery with cerebral infarction |
| Cerebrovascular disease | 433 | ICD10CM | I63.213 | Cerebral infarction due to unspecified occlusion or stenosis of bilateral vertebral arteries |
| Cerebrovascular disease | 433 | ICD10CM | I63.20 | Cerebral infarction due to unspecified occlusion or stenosis of unspecified precerebral arteries |
| Cerebrovascular disease | 433 | ICD10CM | I63.119 | Cerebral infarction due to embolism of unspecified vertebral artery |
| Cerebrovascular disease | 433 | ICD10CM | I63.23 | Cerebral infarction due to unspecified occlusion or stenosis of carotid arteries |
| Cerebrovascular disease | 433 | ICD10CM | I63.22 | Cerebral infarction due to unspecified occlusion or stenosis of basilar artery |
| Cerebrovascular disease | 433 | ICD10CM | I63.132 | Cerebral infarction due to embolism of left carotid artery |
| Cerebrovascular disease | 433 | ICD10CM | I63.031 | Cerebral infarction due to thrombosis of right carotid artery |

| | | | | |
|---|---|---|---|---|
| Cerebrovascular disease | 433 | ICD10CM | I63.111 | Cerebral infarction due to embolism of right vertebral artery |
| Cerebrovascular disease | 433 | ICD10CM | I63.02 | Cerebral infarction due to thrombosis of basilar artery |
| Cerebrovascular disease | 433 | ICD10CM | I63.233 | Cerebral infarction due to unspecified occlusion or stenosis of bilateral carotid arteries |
| Cerebrovascular disease | 433 | ICD9CM | 433.11 | Occlusion and stenosis of carotid artery with cerebral infarction |
| Cerebrovascular disease | 433 | ICD10CM | I63.139 | Cerebral infarction due to embolism of unspecified carotid artery |
| Cerebrovascular disease | 433 | ICD10CM | I63.011 | Cerebral infarction due to thrombosis of right vertebral artery |
| Cerebrovascular disease | 433 | ICD10CM | I63.032 | Cerebral infarction due to thrombosis of left carotid artery |
| Cerebrovascular disease | 433 | ICD10CM | I63.231 | Cerebral infarction due to unspecified occlusion or stenosis of right carotid arteries |
| Cerebrovascular disease | 433 | ICD10CM | I63.1 | Cerebral infarction due to embolism of precerebral arteries |
| Cerebrovascular disease | 433 | ICD10CM | I63.239 | Cerebral infarction due to unspecified occlusion or stenosis of unspecified carotid artery |
| Cerebrovascular disease | 433 | ICD9CM | 433.01 | Occlusion and stenosis of basilar artery with cerebral infarction |
| Cerebrovascular disease | 433 | ICD10CM | I63.03 | Cerebral infarction due to thrombosis of carotid artery |
| Cerebrovascular disease | 433 | ICD10CM | I63.133 | Cerebral infarction due to embolism of bilateral carotid arteries |
| Cerebrovascular disease | 433 | ICD10CM | I63.039 | Cerebral infarction due to thrombosis of unspecified carotid artery |
| Cerebrovascular disease | 433 | ICD9CM | 433.91 | Occlusion and stenosis of unspecified precerebral artery with cerebral infarction |
| Cerebrovascular disease | 433 | ICD10CM | I63.19 | Cerebral infarction due to embolism of other precerebral artery |
| Cerebrovascular disease | 433 | ICD10CM | I63.00 | Cerebral infarction due to thrombosis of unspecified precerebral artery |
| Cerebrovascular disease | 433 | ICD10CM | I63.11 | Cerebral infarction due to embolism of vertebral artery |
| Cerebrovascular disease | 433 | ICD10CM | I63.219 | Cerebral infarction due to unspecified occlusion or stenosis of unspecified vertebral artery |
| Cerebrovascular disease | 433 | ICD10CM | I63.212 | Cerebral infarction due to unspecified occlusion or stenosis of left vertebral artery |
| Cerebrovascular disease | 433 | ICD10CM | I63.131 | Cerebral infarction due to embolism of right carotid artery |
| Cerebrovascular disease | 433 | ICD9CM | 433.81 | Occlusion and stenosis of other specified precerebral artery with cerebral infarction |
| Cerebrovascular disease | 433 | ICD10CM | I63.21 | Cerebral infarction due to unspecified occlusion or stenosis of vertebral arteries |
| Cerebrovascular disease | 433 | ICD10CM | I63.12 | Cerebral infarction due to embolism of basilar artery |
| Cerebrovascular disease | 433 | ICD10CM | I63.09 | Cerebral infarction due to thrombosis of other precerebral artery |

| | | | | |
|---|---|---|---|---|
| Cerebrovascular disease | 433 | ICD10CM | I63.01 | Cerebral infarction due to thrombosis of vertebral artery |
| Cerebrovascular disease | 433 | ICD10CM | I69.832 | Monoplegia of upper limb following other cerebrovascular disease affecting left dominant side |
| Cerebrovascular disease | 433 | ICD10CM | I69.259 | Hemiplegia and hemiparesis following other nontraumatic intracranial hemorrhage affecting unspecified side |
| Cerebrovascular disease | 433 | ICD10CM | I69.22 | Speech and language deficits following other nontraumatic intracranial hemorrhage |
| Cerebrovascular disease | 433 | ICD10CM | I69.05 | Hemiplegia and hemiparesis following nontraumatic subarachnoid hemorrhage |
| Cerebrovascular disease | 433 | ICD10CM | I69.141 | Monoplegia of lower limb following nontraumatic intracerebral hemorrhage affecting right dominant side |
| Cerebrovascular disease | 433 | ICD10CM | I69.15 | Hemiplegia and hemiparesis following nontraumatic intracerebral hemorrhage |
| Cerebrovascular disease | 433 | ICD10CM | I69.254 | Hemiplegia and hemiparesis following other nontraumatic intracranial hemorrhage affecting left non-dominant side |
| Cerebrovascular disease | 433 | ICD10CM | I69.332 | Monoplegia of upper limb following cerebral infarction affecting left dominant side |
| Cerebrovascular disease | 433 | ICD10CM | I69.915 | Cognitive social or emotional deficit following unspecified cerebrovascular disease |
| Cerebrovascular disease | 433 | ICD10CM | I69.061 | Other paralytic syndrome following nontraumatic subarachnoid hemorrhage affecting right dominant side |
| Cerebrovascular disease | 433 | ICD10CM | I69.069 | Other paralytic syndrome following nontraumatic subarachnoid hemorrhage affecting unspecified side |
| Cerebrovascular disease | 433 | ICD10CM | I69.892 | Facial weakness following other cerebrovascular disease |
| Cerebrovascular disease | 433 | ICD10CM | I69.96 | Other paralytic syndrome following unspecified cerebrovascular disease |
| Cerebrovascular disease | 433 | ICD10CM | I69.9 | Sequelae of unspecified cerebrovascular diseases |
| Cerebrovascular disease | 433 | ICD10CM | I69.122 | Dysarthria following nontraumatic intracerebral hemorrhage |
| Cerebrovascular disease | 433 | ICD10CM | I69.065 | Other paralytic syndrome following nontraumatic subarachnoid hemorrhage, bilateral |
| Cerebrovascular disease | 433 | ICD10CM | I69.998 | Other sequelae following unspecified cerebrovascular disease |
| Cerebrovascular disease | 433 | ICD10CM | I69.965 | Other paralytic syndrome following unspecified cerebrovascular disease, bilateral |
| Cerebrovascular disease | 433 | ICD10CM | I69.913 | Psychomotor deficit following unspecified cerebrovascular disease |
| Cerebrovascular disease | 433 | ICD10CM | I69.891 | Dysphagia following other cerebrovascular disease |
| Cerebrovascular disease | 433 | ICD10CM | I69.162 | Other paralytic syndrome following nontraumatic intracerebral hemorrhage affecting left dominant side |

| | | | | |
|---|---|---|---|---|
| Cerebrovascular disease | 433 | ICD10CM | I69.820 | Aphasia following other cerebrovascular disease |
| Cerebrovascular disease | 433 | ICD10CM | I69.823 | Fluency disorder following other cerebrovascular disease |
| Cerebrovascular disease | 433 | ICD9CM | 438.11 | Late effects of cerebrovascular disease, aphasia |
| Cerebrovascular disease | 433 | ICD10CM | I69.051 | Hemiplegia and hemiparesis following nontraumatic subarachnoid hemorrhage affecting right dominant side |
| Cerebrovascular disease | 433 | ICD10CM | I69.813 | Psychomotor deficit following other cerebrovascular disease |
| Cerebrovascular disease | 433 | ICD10CM | I69.844 | Monoplegia of lower limb following other cerebrovascular disease affecting left non-dominant side |
| Cerebrovascular disease | 433 | ICD10CM | I69.149 | Monoplegia of lower limb following nontraumatic intracerebral hemorrhage affecting unspecified side |
| Cerebrovascular disease | 433 | ICD10CM | I69.013 | Psychomotor deficit following nontraumatic subarachnoid hemorrhage |
| Cerebrovascular disease | 433 | ICD10CM | I69.014 | Frontal lobe and executive function deficit following nontraumatic subarachnoid hemorrhage |
| Cerebrovascular disease | 433 | ICD10CM | I69.131 | Monoplegia of upper limb following nontraumatic intracerebral hemorrhage affecting right dominant side |
| Cerebrovascular disease | 433 | ICD9CM | 438.3 | Late effects of cerebrovascular disease, monoplegia of upper limb affecting unspecified side |
| Cerebrovascular disease | 433 | ICD10CM | I69.993 | Ataxia following unspecified cerebrovascular disease |
| Cerebrovascular disease | 433 | ICD10CM | I69.919 | Unspecified symptoms and signs involving cognitive functions following unspecified cerebrovascular disease |
| Cerebrovascular disease | 433 | ICD10CM | I69.253 | Hemiplegia and hemiparesis following other nontraumatic intracranial hemorrhage affecting right non-dominant side |
| Cerebrovascular disease | 433 | ICD10CM | I69.932 | Monoplegia of upper limb following unspecified cerebrovascular disease affecting left dominant side |
| Cerebrovascular disease | 433 | ICD10CM | I69.154 | Hemiplegia and hemiparesis following nontraumatic intracerebral hemorrhage affecting left non-dominant side |
| Cerebrovascular disease | 433 | ICD10CM | I69.291 | Dysphagia following other nontraumatic intracranial hemorrhage |
| Cerebrovascular disease | 433 | ICD10CM | I69.94 | Monoplegia of lower limb following unspecified cerebrovascular disease |
| Cerebrovascular disease | 433 | ICD10CM | I69.049 | Monoplegia of lower limb following nontraumatic subarachnoid hemorrhage affecting unspecified side |
| Cerebrovascular disease | 433 | ICD10CM | I69.134 | Monoplegia of upper limb following nontraumatic intracerebral hemorrhage affecting left non-dominant side |

| | | | | |
|---|---|---|---|---|
| Cerebrovascular disease | 433 | ICD10CM | I69.81 | Cognitive deficits following other cerebrovascular disease |
| Cerebrovascular disease | 433 | ICD10CM | I69.34 | Monoplegia of lower limb following cerebral infarction |
| Cerebrovascular disease | 433 | ICD10CM | I69.339 | Monoplegia of upper limb following cerebral infarction affecting unspecified side |
| Cerebrovascular disease | 433 | ICD10CM | I69.219 | Unspecified symptoms and signs involving cognitive functions following other nontraumatic intracranial hemorrhage |
| Cerebrovascular disease | 433 | ICD10CM | I69.952 | Hemiplegia and hemiparesis following unspecified cerebrovascular disease affecting left dominant side |
| Cerebrovascular disease | 433 | ICD10CM | I69.341 | Monoplegia of lower limb following cerebral infarction affecting right dominant side |
| Cerebrovascular disease | 433 | ICD10CM | I69.29 | Other sequelae of other nontraumatic intracranial hemorrhage |
| Cerebrovascular disease | 433 | ICD10CM | I69.052 | Hemiplegia and hemiparesis following nontraumatic subarachnoid hemorrhage affecting left dominant side |
| Cerebrovascular disease | 433 | ICD10CM | I69.04 | Monoplegia of lower limb following nontraumatic subarachnoid hemorrhage |
| Cerebrovascular disease | 433 | ICD10CM | I69.953 | Hemiplegia and hemiparesis following unspecified cerebrovascular disease affecting right non-dominant side |
| Cerebrovascular disease | 433 | ICD10CM | I69.928 | Other speech and language deficits following unspecified cerebrovascular disease |
| Cerebrovascular disease | 433 | ICD10CM | I69.054 | Hemiplegia and hemiparesis following nontraumatic subarachnoid hemorrhage affecting left non-dominant side |
| Cerebrovascular disease | 433 | ICD10CM | I69.233 | Monoplegia of upper limb following other nontraumatic intracranial hemorrhage affecting right non-dominant side |
| Cerebrovascular disease | 433 | ICD10CM | I69.933 | Monoplegia of upper limb following unspecified cerebrovascular disease affecting right non-dominant side |
| Cerebrovascular disease | 433 | ICD10CM | I69.949 | Monoplegia of lower limb following unspecified cerebrovascular disease affecting unspecified side |
| Cerebrovascular disease | 433 | ICD10CM | I69.1 | Sequelae of nontraumatic intracerebral hemorrhage |
| Cerebrovascular disease | 433 | ICD10CM | I69.012 | Visuospatial deficit and spatial neglect following nontraumatic subarachnoid hemorrhage |
| Cerebrovascular disease | 433 | ICD9CM | 438.31 | Late effects of cerebrovascular disease, monoplegia of upper limb affecting dominant side |
| Cerebrovascular disease | 433 | ICD10CM | I69.264 | Other paralytic syndrome following other nontraumatic intracranial hemorrhage affecting left non-dominant side |
| Cerebrovascular disease | 433 | ICD10CM | I69.85 | Hemiplegia and hemiparesis following other cerebrovascular disease |

| | | | | |
|---|---|---|---|---|
| Cerebrovascular disease | 433 | ICD10CM | I69.963 | Other paralytic syndrome following unspecified cerebrovascular disease affecting right non-dominant side |
| Cerebrovascular disease | 433 | ICD10CM | I69.210 | Attention and concentration deficit following other nontraumatic intracranial hemorrhage |
| Cerebrovascular disease | 433 | ICD10CM | I69.961 | Other paralytic syndrome following unspecified cerebrovascular disease affecting right dominant side |
| Cerebrovascular disease | 433 | ICD10CM | I69.212 | Visuospatial deficit and spatial neglect following other nontraumatic intracranial hemorrhage |
| Cerebrovascular disease | 433 | ICD10CM | I67.81 | Acute cerebrovascular insufficiency |
| Cerebrovascular disease | 433 | ICD10CM | I67.82 | Cerebral ischemia |
| Cerebrovascular disease | 433 | ICD10CM | I67.89 | Other cerebrovascular disease |
| Cerebrovascular disease | 433 | ICD9CM | 437.1 | Other generalized ischemic cerebrovascular disease |
| Cerebrovascular disease | 433 | ICD10CM | I67.8 | Other specified cerebrovascular diseases |
| Cerebrovascular disease | 433 | ICD10CM | I69.244 | Monoplegia of lower limb following other nontraumatic intracranial hemorrhage affecting left non-dominant side |
| Cerebrovascular disease | 433 | ICD10CM | I69.918 | Other symptoms and signs involving cognitive functions following unspecified cerebrovascular disease |
| Cerebrovascular disease | 433 | ICD10CM | I69.062 | Other paralytic syndrome following nontraumatic subarachnoid hemorrhage affecting left dominant side |
| Cerebrovascular disease | 433 | ICD10CM | I69.922 | Dysarthria following unspecified cerebrovascular disease |
| Cerebrovascular disease | 433 | ICD9CM | 438.4 | Monoplegia of lower limb as late effect of cerebrovascular disease |
| Cerebrovascular disease | 433 | ICD10CM | I69.228 | Other speech and language deficits following other nontraumatic intracranial hemorrhage |
| Cerebrovascular disease | 433 | ICD10CM | I69.92 | Speech and language deficits following unspecified cerebrovascular disease |
| Cerebrovascular disease | 433 | ICD10CM | I69.943 | Monoplegia of lower limb following unspecified cerebrovascular disease affecting right non-dominant side |
| Cerebrovascular disease | 433 | ICD10CM | M47.02 | Vertebral artery compression syndromes |
| Cerebrovascular disease | 433 | ICD9CM | 435.3 | Vertebrobasilar artery syndrome |
| Cerebrovascular disease | 433 | ICD9CM | 435.8 | Other specified transient cerebral ischemias |
| Cerebrovascular disease | 433 | ICD9CM | 435 | Basilar artery syndrome |
| Cerebrovascular disease | 433 | ICD9CM | 435.9 | Unspecified transient cerebral ischemia |

| | | | | |
|---|---|---|---|---|
| Cerebrovascular disease | 433 | ICD10CM | G46.0 | Middle cerebral artery syndrome |
| Cerebrovascular disease | 433 | ICD10CM | I67.2 | Cerebral atherosclerosis |
| Cerebrovascular disease | 433 | ICD9CM | 437 | Cerebral atherosclerosis |
| Cerebrovascular disease | 433 | ICD10CM | Z86.73 | Personal history of transient ischemic attack (TIA), and cerebral infarction without residual deficits |
| Cerebrovascular disease | 433 | ICD10CM | G45.8 | Other transient cerebral ischemic attacks and related syndromes |
| Cerebrovascular disease | 433 | ICD9CM | V12.54 | Personal history of transient ischemic attack (TIA), and cerebral infarction without residual deficits |
| Cerebrovascular disease | 433 | ICD10CM | G45.0 | Vertebro-basilar artery syndrome |
| Cerebrovascular disease | 433 | ICD10CM | G46.2 | Posterior cerebral artery syndrome |
| Cerebrovascular disease | 433 | ICD10CM | I67.6 | Nonpyogenic thrombosis of intracranial venous system |
| Cerebrovascular disease | 433 | ICD10CM | I66.23 | Occlusion and stenosis of bilateral posterior cerebral arteries |
| Cerebrovascular disease | 433 | ICD10CM | I66.13 | Occlusion and stenosis of bilateral anterior cerebral arteries |
| Cerebrovascular disease | 433 | ICD10CM | I66.29 | Occlusion and stenosis of unspecified posterior cerebral artery |
| Cerebrovascular disease | 433 | ICD10CM | I66.19 | Occlusion and stenosis of unspecified anterior cerebral artery |
| Cerebrovascular disease | 433 | ICD10CM | I66.03 | Occlusion and stenosis of bilateral middle cerebral arteries |
| Cerebrovascular disease | 433 | ICD10CM | I66.9 | Occlusion and stenosis of unspecified cerebral artery |
| Cerebrovascular disease | 433 | ICD9CM | 434 | Occlusion of cerebral arteries |
| Cerebrovascular disease | 433 | ICD10CM | I66.11 | Occlusion and stenosis of right anterior cerebral artery |
| Cerebrovascular disease | 433 | ICD10CM | I63 | Cerebral infarction |
| Cerebrovascular disease | 433 | ICD10CM | I66 | Occlusion and stenosis of cerebral arteries, not resulting in cerebral infarction |
| Cerebrovascular disease | 433 | ICD9CM | 434.1 | Cerebral embolism without mention of cerebral infarction |
| Cerebrovascular disease | 433 | ICD10CM | I66.22 | Occlusion and stenosis of left posterior cerebral artery |
| Cerebrovascular disease | 433 | ICD9CM | 434 | Cerebral thrombosis without mention of cerebral infarction |
| Cerebrovascular disease | 433 | ICD10CM | I66.21 | Occlusion and stenosis of right posterior cerebral artery |
| Cerebrovascular disease | 433 | ICD10CM | I66.02 | Occlusion and stenosis of left middle cerebral artery |

| | | | | |
|---|---|---|---|---|
| Cerebrovascular disease | 433 | ICD10CM | I66.8 | Occlusion and stenosis of other cerebral arteries |
| Cerebrovascular disease | 433 | ICD9CM | 437.6 | Nonpyogenic thrombosis of intracranial venous sinus |
| Cerebrovascular disease | 433 | ICD10CM | I66.01 | Occlusion and stenosis of right middle cerebral artery |
| Cerebrovascular disease | 433 | ICD9CM | 434.1 | Cerebral embolism |
| Cerebrovascular disease | 433 | ICD9CM | 434.9 | Cerebral artery occlusion, unspecified |
| Cerebrovascular disease | 433 | ICD10CM | I66.1 | Occlusion and stenosis of anterior cerebral artery |
| Cerebrovascular disease | 433 | ICD10CM | I66.3 | Occlusion and stenosis of cerebellar arteries |
| Cerebrovascular disease | 433 | ICD10CM | I66.2 | Occlusion and stenosis of posterior cerebral artery |
| Cerebrovascular disease | 433 | ICD10CM | I66.12 | Occlusion and stenosis of left anterior cerebral artery |
| Cerebrovascular disease | 433 | ICD10CM | I66.09 | Occlusion and stenosis of unspecified middle cerebral artery |
| Cerebrovascular disease | 433 | ICD9CM | 434 | Cerebral thrombosis |
| Cerebrovascular disease | 433 | ICD10CM | I66.0 | Occlusion and stenosis of middle cerebral artery |
| Cerebrovascular disease | 433 | ICD9CM | 434.9 | Cerebral artery occlusion, unspecified without mention of cerebral infarction |
| Cerebrovascular disease | 433 | ICD9CM | 438.51 | Late effects of cerebrovascular disease, other paralytic syndrome affecting dominant side |
| Cerebrovascular disease | 433 | ICD10CM | I69.090 | Apraxia following nontraumatic subarachnoid hemorrhage |
| Cerebrovascular disease | 433 | ICD10CM | I69.10 | Unspecified sequelae of nontraumatic intracerebral hemorrhage |
| Cerebrovascular disease | 433 | ICD10CM | I69.252 | Hemiplegia and hemiparesis following other nontraumatic intracranial hemorrhage affecting left dominant side |
| Cerebrovascular disease | 433 | ICD9CM | 438.4 | Late effects of cerebrovascular disease, monoplegia of lower limb affecting unspecified side |
| Cerebrovascular disease | 433 | ICD10CM | I69.962 | Other paralytic syndrome following unspecified cerebrovascular disease affecting left dominant side |
| Cerebrovascular disease | 433 | ICD10CM | I69.039 | Monoplegia of upper limb following nontraumatic subarachnoid hemorrhage affecting unspecified side |
| Cerebrovascular disease | 433 | ICD10CM | I69.044 | Monoplegia of lower limb following nontraumatic subarachnoid hemorrhage affecting left non-dominant side |
| Cerebrovascular disease | 433 | ICD10CM | I69.112 | Visuospatial deficit and spatial neglect following nontraumatic intracerebral hemorrhage |
| Cerebrovascular disease | 433 | ICD10CM | I69.115 | Cognitive social or emotional deficit following nontraumatic intracerebral hemorrhage |

| | | | | |
|---|---|---|---|---|
| Cerebrovascular disease | 433 | ICD10CM | I69.098 | Other sequelae following nontraumatic subarachnoid hemorrhage |
| Cerebrovascular disease | 433 | ICD10CM | I69.265 | Other paralytic syndrome following other nontraumatic intracranial hemorrhage, bilateral |
| Cerebrovascular disease | 433 | ICD10CM | I69.911 | Memory deficit following unspecified cerebrovascular disease |
| Cerebrovascular disease | 433 | ICD10CM | I69.910 | Attention and concentration deficit following unspecified cerebrovascular disease |
| Cerebrovascular disease | 433 | ICD9CM | 438.19 | Late effects of cerebrovascular disease, other speech and language deficits |
| Cerebrovascular disease | 433 | ICD10CM | I69 | Sequelae of cerebrovascular disease |
| Cerebrovascular disease | 433 | ICD10CM | I69.923 | Fluency disorder following unspecified cerebrovascular disease |
| Cerebrovascular disease | 433 | ICD9CM | 438 | Late effects of cerebrovascular disease, cognitive deficits |
| Cerebrovascular disease | 433 | ICD10CM | I69.159 | Hemiplegia and hemiparesis following nontraumatic intracerebral hemorrhage affecting unspecified side |
| Cerebrovascular disease | 433 | ICD10CM | I69.161 | Other paralytic syndrome following nontraumatic intracerebral hemorrhage affecting right dominant side |
| Cerebrovascular disease | 433 | ICD10CM | I69.812 | Visuospatial deficit and spatial neglect following other cerebrovascular disease |
| Cerebrovascular disease | 433 | ICD10CM | I69.192 | Facial weakness following nontraumatic intracerebral hemorrhage |
| Cerebrovascular disease | 433 | ICD10CM | I69.198 | Other sequelae of nontraumatic intracerebral hemorrhage |
| Cerebrovascular disease | 433 | ICD10CM | I69.14 | Monoplegia of lower limb following nontraumatic intracerebral hemorrhage |
| Cerebrovascular disease | 433 | ICD10CM | I69.944 | Monoplegia of lower limb following unspecified cerebrovascular disease affecting left non-dominant side |
| Cerebrovascular disease | 433 | ICD10CM | I69.91 | Cognitive deficits following unspecified cerebrovascular disease |
| Cerebrovascular disease | 433 | ICD10CM | I69.992 | Facial weakness following unspecified cerebrovascular disease |
| Cerebrovascular disease | 433 | ICD10CM | I69.959 | Hemiplegia and hemiparesis following unspecified cerebrovascular disease affecting unspecified side |
| Cerebrovascular disease | 433 | ICD10CM | I69.95 | Hemiplegia and hemiparesis following unspecified cerebrovascular disease |
| Cerebrovascular disease | 433 | ICD10CM | I69.364 | Other paralytic syndrome following cerebral infarction affecting left non-dominant side |
| Cerebrovascular disease | 433 | ICD9CM | 437.5 | Moyamoya disease |
| Cerebrovascular disease | 433 | ICD10CM | I67.5 | Moyamoya disease |
| Cerebrovascular disease | 433 | ICD10CM | I63.54 | Cerebral infarction due to unspecified occlusion or stenosis of cerebellar artery |

| | | | | |
|---|---|---|---|---|
| Cerebrovascular disease | 433 | ICD10CM | I63.541 | Cerebral infarction due to unspecified occlusion or stenosis of right cerebellar artery |
| Cerebrovascular disease | 433 | ICD10CM | I63.421 | Cerebral infarction due to embolism of right anterior cerebral artery |
| Cerebrovascular disease | 433 | ICD10CM | I63.522 | Cerebral infarction due to unspecified occlusion or stenosis of left anterior cerebral artery |
| Cerebrovascular disease | 433 | ICD10CM | I63.9 | Cerebral infarction, unspecified |
| Cerebrovascular disease | 433 | ICD10CM | I63.32 | Cerebral infarction due to thrombosis of anterior cerebral artery |
| Cerebrovascular disease | 433 | ICD10CM | I63.439 | Cerebral infarction due to embolism of unspecified posterior cerebral artery |
| Cerebrovascular disease | 433 | ICD10CM | I63.431 | Cerebral infarction due to embolism of right posterior cerebral artery |
| Cerebrovascular disease | 433 | ICD10CM | I63.411 | Cerebral infarction due to embolism of right middle cerebral artery |
| Cerebrovascular disease | 433 | ICD10CM | I63.512 | Cerebral infarction due to unspecified occlusion or stenosis of left middle cerebral artery |
| Cerebrovascular disease | 433 | ICD9CM | 434.11 | Cerebral embolism with cerebral infarction |
| Cerebrovascular disease | 433 | ICD10CM | I63.323 | Cerebral infarction due to thrombosis of bilateral anterior cerebral arteries |
| Cerebrovascular disease | 433 | ICD10CM | I63.531 | Cerebral infarction due to unspecified occlusion or stenosis of right posterior cerebral artery |
| Cerebrovascular disease | 433 | ICD10CM | I63.59 | Cerebral infarction due to unspecified occlusion or stenosis of other cerebral artery |
| Cerebrovascular disease | 433 | ICD10CM | I63.33 | Cerebral infarction due to thrombosis of posterior cerebral artery |
| Cerebrovascular disease | 433 | ICD10CM | I63.52 | Cerebral infarction due to unspecified occlusion or stenosis of anterior cerebral artery |
| Cerebrovascular disease | 433 | ICD10CM | I63.333 | Cerebral infarction due to thrombosis of bilateral posterior cerebral arteries |
| Cerebrovascular disease | 433 | ICD10CM | I63.543 | Cerebral infarction due to unspecified occlusion or stenosis of bilateral cerebellar arteries |
| Cerebrovascular disease | 433 | ICD10CM | I63.30 | Cerebral infarction due to thrombosis of unspecified cerebral artery |
| Cerebrovascular disease | 433 | ICD10CM | I63.539 | Cerebral infarction due to unspecified occlusion or stenosis of unspecified posterior cerebral artery |
| Cerebrovascular disease | 433 | ICD9CM | 434.91 | Cerebral artery occlusion, unspecified with cerebral infarction |
| Cerebrovascular disease | 433 | ICD10CM | I63.432 | Cerebral infarction due to embolism of left posterior cerebral artery |
| Cerebrovascular disease | 433 | ICD10CM | I63.542 | Cerebral infarction due to unspecified occlusion or stenosis of left cerebellar artery |
| Cerebrovascular disease | 433 | ICD10CM | I63.8 | Other cerebral infarction |
| Cerebrovascular disease | 433 | ICD10CM | I63.31 | Cerebral infarction due to thrombosis of middle cerebral artery |

| | | | | |
|---|---|---|---|---|
| Cerebrovascular disease | 433 | ICD10CM | I63.0 | Cerebral infarction due to thrombosis of precerebral arteries |
| Cerebrovascular disease | 433 | ICD10CM | I63.513 | Cerebral infarction due to unspecified occlusion or stenosis of bilateral middle cerebral arteries |
| Cerebrovascular disease | 433 | ICD10CM | I63.321 | Cerebral infarction due to thrombosis of right anterior cerebral artery |
| Cerebrovascular disease | 433 | ICD10CM | I63.429 | Cerebral infarction due to embolism of unspecified anterior cerebral artery |
| Cerebrovascular disease | 433 | ICD10CM | I63.40 | Cerebral infarction due to embolism of unspecified cerebral artery |
| Cerebrovascular disease | 433 | ICD10CM | I63.311 | Cerebral infarction due to thrombosis of right middle cerebral artery |
| Cerebrovascular disease | 433 | ICD10CM | I63.341 | Cerebral infarction due to thrombosis of right cerebellar artery |
| Cerebrovascular disease | 433 | ICD10CM | I63.549 | Cerebral infarction due to unspecified occlusion or stenosis of unspecified cerebellar artery |
| Cerebrovascular disease | 433 | ICD10CM | I63.413 | Cerebral infarction due to embolism of bilateral middle cerebral arteries |
| Cerebrovascular disease | 433 | ICD10CM | I63.322 | Cerebral infarction due to thrombosis of left anterior cerebral artery |
| Cerebrovascular disease | 433 | ICD10CM | I63.41 | Cerebral infarction due to embolism of middle cerebral artery |
| Cerebrovascular disease | 433 | ICD10CM | I63.529 | Cerebral infarction due to unspecified occlusion or stenosis of unspecified anterior cerebral artery |
| Cerebrovascular disease | 433 | ICD10CM | I63.42 | Cerebral infarction due to embolism of anterior cerebral artery |
| Cerebrovascular disease | 433 | ICD10CM | I63.51 | Cerebral infarction due to unspecified occlusion or stenosis of middle cerebral artery |
| Cerebrovascular disease | 433 | ICD10CM | I63.533 | Cerebral infarction due to unspecified occlusion or stenosis of bilateral posterior cerebral arteries |
| Cerebrovascular disease | 433 | ICD10CM | I63.34 | Cerebral infarction due to thrombosis of cerebellar artery |
| Cerebrovascular disease | 433 | ICD10CM | I63.6 | Cerebral infarction due to cerebral venous thrombosis, nonpyogenic |
| Cerebrovascular disease | 433 | ICD9CM | 434.01 | Cerebral thrombosis with cerebral infarction |
| Cerebrovascular disease | 433 | ICD10CM | I63.532 | Cerebral infarction due to unspecified occlusion or stenosis of left posterior cerebral artery |
| Cerebrovascular disease | 433 | ICD10CM | I63.343 | Cerebral infarction due to thrombosis of bilateral cerebellar arteries |
| Cerebrovascular disease | 433 | ICD10CM | I63.329 | Cerebral infarction due to thrombosis of unspecified anterior cerebral artery |
| Cerebrovascular disease | 433 | ICD10CM | I63.511 | Cerebral infarction due to unspecified occlusion or stenosis of right middle cerebral artery |
| Cerebrovascular disease | 433 | ICD10CM | I63.349 | Cerebral infarction due to thrombosis of unspecified cerebellar artery |
| Cerebrovascular disease | 433 | ICD10CM | I63.44 | Cerebral infarction due to embolism of cerebellar artery |

| | | | | |
|---|---|---|---|---|
| Cerebrovascular disease | 433 | ICD10CM | I63.441 | Cerebral infarction due to embolism of right cerebellar artery |
| Cerebrovascular disease | 433 | ICD10CM | I63.422 | Cerebral infarction due to embolism of left anterior cerebral artery |
| Cerebrovascular disease | 433 | ICD10CM | I63.313 | Cerebral infarction due to thrombosis of bilateral middle cerebral arteries |
| Cerebrovascular disease | 433 | ICD10CM | I63.521 | Cerebral infarction due to unspecified occlusion or stenosis of right anterior cerebral artery |
| Cerebrovascular disease | 433 | ICD10CM | I63.49 | Cerebral infarction due to embolism of other cerebral artery |
| Cerebrovascular disease | 433 | ICD10CM | I63.319 | Cerebral infarction due to thrombosis of unspecified middle cerebral artery |
| Cerebrovascular disease | 433 | ICD10CM | I63.332 | Cerebral infarction due to thrombosis of left posterior cerebral artery |
| Cerebrovascular disease | 433 | ICD10CM | I63.423 | Cerebral infarction due to embolism of bilateral anterior cerebral arteries |
| Cerebrovascular disease | 433 | ICD10CM | I63.4 | Cerebral infarction due to embolism of cerebral arteries |
| Cerebrovascular disease | 433 | ICD10CM | I63.523 | Cerebral infarction due to unspecified occlusion or stenosis of bilateral anterior cerebral arteries |
| Cerebrovascular disease | 433 | ICD10CM | I63.53 | Cerebral infarction due to unspecified occlusion or stenosis of posterior cerebral artery |
| Cerebrovascular disease | 433 | ICD10CM | I63.412 | Cerebral infarction due to embolism of left middle cerebral artery |
| Cerebrovascular disease | 433 | ICD10CM | I63.3 | Cerebral infarction due to thrombosis of cerebral arteries |
| Cerebrovascular disease | 433 | ICD10CM | I63.443 | Cerebral infarction due to embolism of bilateral cerebellar arteries |
| Cerebrovascular disease | 433 | ICD10CM | I63.442 | Cerebral infarction due to embolism of left cerebellar artery |
| Cerebrovascular disease | 433 | ICD10CM | I63.39 | Cerebral infarction due to thrombosis of other cerebral artery |
| Cerebrovascular disease | 433 | ICD10CM | I63.342 | Cerebral infarction due to thrombosis of left cerebellar artery |
| Cerebrovascular disease | 433 | ICD10CM | I63.339 | Cerebral infarction due to thrombosis of unspecified posterior cerebral artery |
| Cerebrovascular disease | 433 | ICD10CM | I63.419 | Cerebral infarction due to embolism of unspecified middle cerebral artery |
| Cerebrovascular disease | 433 | ICD10CM | I63.312 | Cerebral infarction due to thrombosis of left middle cerebral artery |
| Cerebrovascular disease | 433 | ICD10CM | I63.50 | Cerebral infarction due to unspecified occlusion or stenosis of unspecified cerebral artery |
| Cerebrovascular disease | 433 | ICD10CM | I63.5 | Cerebral infarction due to unspecified occlusion or stenosis of cerebral arteries |
| Cerebrovascular disease | 433 | ICD10CM | I63.449 | Cerebral infarction due to embolism of unspecified cerebellar artery |
| Cerebrovascular disease | 433 | ICD10CM | I63.433 | Cerebral infarction due to embolism of bilateral posterior cerebral arteries |
| Cerebrovascular disease | 433 | ICD10CM | I63.331 | Cerebral infarction due to thrombosis of right posterior cerebral artery |

| | | | | |
|---|---|---|---|---|
| Cerebrovascular disease | 433 | ICD10CM | I63.43 | Cerebral infarction due to embolism of posterior cerebral artery |
| Cerebrovascular disease | 433 | ICD10CM | I63.519 | Cerebral infarction due to unspecified occlusion or stenosis of unspecified middle cerebral artery |
| Cerebrovascular disease | 433 | ICD9CM | 438.22 | Late effects of cerebrovascular disease, hemiplegia affecting nondominant side |
| Cerebrovascular disease | 433 | ICD10CM | I69.133 | Monoplegia of upper limb following nontraumatic intracerebral hemorrhage affecting right non-dominant side |
| Cerebrovascular disease | 433 | ICD10CM | I69.841 | Monoplegia of lower limb following other cerebrovascular disease affecting right dominant side |
| Cerebrovascular disease | 433 | ICD10CM | I69.143 | Monoplegia of lower limb following nontraumatic intracerebral hemorrhage affecting right non-dominant side |
| Cerebrovascular disease | 433 | ICD9CM | 438.9 | Unspecified late effects of cerebrovascular disease |
| Cerebrovascular disease | 433 | ICD10CM | I69.861 | Other paralytic syndrome following other cerebrovascular disease affecting right dominant side |
| Cerebrovascular disease | 433 | ICD10CM | I69.369 | Other paralytic syndrome following cerebral infarction affecting unspecified side |
| Cerebrovascular disease | 433 | ICD10CM | I69.842 | Monoplegia of lower limb following other cerebrovascular disease affecting left dominant side |
| Cerebrovascular disease | 433 | ICD10CM | I69.193 | Ataxia following nontraumatic intracerebral hemorrhage |
| Cerebrovascular disease | 433 | ICD10CM | I69.11 | Cognitive deficits following nontraumatic intracerebral hemorrhage |
| Cerebrovascular disease | 433 | ICD9CM | 438.2 | Late effects of cerebrovascular disease, hemiplegia affecting unspecified side |
| Cerebrovascular disease | 433 | ICD10CM | I69.20 | Unspecified sequelae of other nontraumatic intracranial hemorrhage |
| Cerebrovascular disease | 433 | ICD10CM | I69.921 | Dysphasia following unspecified cerebrovascular disease |
| Cerebrovascular disease | 433 | ICD10CM | I69.215 | Cognitive social or emotional deficit following other nontraumatic intracranial hemorrhage |
| Cerebrovascular disease | 433 | ICD10CM | I69.852 | Hemiplegia and hemiparesis following other cerebrovascular disease affecting left dominant side |
| Cerebrovascular disease | 433 | ICD10CM | I69.314 | Frontal lobe and executive function deficit following cerebral infarction |
| Cerebrovascular disease | 433 | ICD10CM | I69.839 | Monoplegia of upper limb following other cerebrovascular disease affecting unspecified side |
| Cerebrovascular disease | 433 | ICD9CM | 438.89 | Other late effects of cerebrovascular disease |
| Cerebrovascular disease | 433 | ICD10CM | I69.123 | Fluency disorder following nontraumatic intracerebral hemorrhage |

| | | | | |
|---|---|---|---|---|
| Cerebrovascular disease | 433 | ICD10CM | I69.153 | Hemiplegia and hemiparesis following nontraumatic intracerebral hemorrhage affecting right non-dominant side |
| Cerebrovascular disease | 433 | ICD10CM | I69.853 | Hemiplegia and hemiparesis following other cerebrovascular disease affecting right non-dominant side |
| Cerebrovascular disease | 433 | ICD10CM | I69.121 | Dysphasia following nontraumatic intracerebral hemorrhage |
| Cerebrovascular disease | 433 | ICD10CM | I69.36 | Other paralytic syndrome following cerebral infarction |
| Cerebrovascular disease | 433 | ICD10CM | I69.851 | Hemiplegia and hemiparesis following other cerebrovascular disease affecting right dominant side |
| Cerebrovascular disease | 433 | ICD10CM | I69.398 | Other sequelae of cerebral infarction |
| Cerebrovascular disease | 433 | ICD9CM | 438.3 | Monoplegia of upper limb as late effect of cerebrovascular disease |
| Cerebrovascular disease | 433 | ICD10CM | I69.128 | Other speech and language deficits following nontraumatic intracerebral hemorrhage |
| Cerebrovascular disease | 433 | ICD10CM | I69.113 | Psychomotor deficit following nontraumatic intracerebral hemorrhage |
| Cerebrovascular disease | 433 | ICD10CM | I69.019 | Unspecified symptoms and signs involving cognitive functions following nontraumatic subarachnoid hemorrhage |
| Cerebrovascular disease | 433 | ICD10CM | I69.163 | Other paralytic syndrome following nontraumatic intracerebral hemorrhage affecting right non-dominant side |
| Cerebrovascular disease | 433 | ICD10CM | I69.311 | Memory deficit following cerebral infarction |
| Cerebrovascular disease | 433 | ICD10CM | I69.034 | Monoplegia of upper limb following nontraumatic subarachnoid hemorrhage affecting left non-dominant side |
| Cerebrovascular disease | 433 | ICD10CM | I69.041 | Monoplegia of lower limb following nontraumatic subarachnoid hemorrhage affecting right dominant side |
| Cerebrovascular disease | 433 | ICD10CM | I69.151 | Hemiplegia and hemiparesis following nontraumatic intracerebral hemorrhage affecting right dominant side |
| Cerebrovascular disease | 433 | ICD10CM | I69.293 | Ataxia following other nontraumatic intracranial hemorrhage |
| Cerebrovascular disease | 433 | ICD10CM | I69.12 | Speech and language deficits following nontraumatic intracerebral hemorrhage |
| Cerebrovascular disease | 433 | ICD10CM | I69.043 | Monoplegia of lower limb following nontraumatic subarachnoid hemorrhage affecting right non-dominant side |
| Cerebrovascular disease | 433 | ICD10CM | I69.033 | Monoplegia of upper limb following nontraumatic subarachnoid hemorrhage affecting right non-dominant side |
| Cerebrovascular disease | 433 | ICD10CM | I69.25 | Hemiplegia and hemiparesis following other nontraumatic intracranial hemorrhage |

| | | | | |
|---|---|---|---|---|
| Cerebrovascular disease | 433 | ICD10CM | I69.333 | Monoplegia of upper limb following cerebral infarction affecting right non-dominant side |
| Cerebrovascular disease | 433 | ICD10CM | I69.319 | Unspecified symptoms and signs involving cognitive functions following cerebral infarction |
| Cerebrovascular disease | 433 | ICD10CM | I69.298 | Other sequelae of other nontraumatic intracranial hemorrhage |
| Cerebrovascular disease | 433 | ICD10CM | I69.220 | Aphasia following other nontraumatic intracranial hemorrhage |
| Cerebrovascular disease | 433 | ICD10CM | I69.144 | Monoplegia of lower limb following nontraumatic intracerebral hemorrhage affecting left non-dominant side |
| Cerebrovascular disease | 433 | ICD10CM | I69.931 | Monoplegia of upper limb following unspecified cerebrovascular disease affecting right dominant side |
| Cerebrovascular disease | 433 | ICD10CM | I69.221 | Dysphasia following other nontraumatic intracranial hemorrhage |
| Cerebrovascular disease | 433 | ICD10CM | I69.893 | Ataxia following other cerebrovascular disease |
| Cerebrovascular disease | 433 | ICD10CM | I69.092 | Facial weakness following nontraumatic subarachnoid hemorrhage |
| Cerebrovascular disease | 433 | ICD9CM | 438.52 | Late effects of cerebrovascular disease, other paralytic syndrome affecting nondominant side |
| Cerebrovascular disease | 433 | ICD10CM | I69.0 | Sequelae of nontraumatic subarachnoid hemorrhage |
| Cerebrovascular disease | 433 | ICD10CM | I69.218 | Other symptoms and signs involving cognitive functions following other nontraumatic intracranial hemorrhage |
| Cerebrovascular disease | 433 | ICD10CM | I69.89 | Other sequelae of other cerebrovascular disease |
| Cerebrovascular disease | 433 | ICD9CM | 346.63 | Persistent migraine aura with cerebral infarction, with intractable migraine, so stated, with status migrainosus |
| Cerebrovascular disease | 433 | ICD9CM | 346.62 | Persistent migraine aura with cerebral infarction, without mention of intractable migraine with status migrainosus |
| Cerebrovascular disease | 433 | ICD10CM | I69.822 | Dysarthria following other cerebrovascular disease |
| Cerebrovascular disease | 433 | ICD10CM | I69.359 | Hemiplegia and hemiparesis following cerebral infarction affecting unspecified side |
| Cerebrovascular disease | 433 | ICD10CM | I69.320 | Aphasia following cerebral infarction |
| Cerebrovascular disease | 433 | ICD10CM | I69.814 | Frontal lobe and executive function deficit following other cerebrovascular disease |
| Cerebrovascular disease | 433 | ICD10CM | I69.142 | Monoplegia of lower limb following nontraumatic intracerebral hemorrhage affecting left dominant side |
| Cerebrovascular disease | 433 | ICD10CM | I69.13 | Monoplegia of upper limb following nontraumatic intracerebral hemorrhage |
| Cerebrovascular disease | 433 | ICD10CM | I69.032 | Monoplegia of upper limb following nontraumatic subarachnoid hemorrhage affecting left dominant side |

| | | | | |
|---|---|---|---|---|
| Cerebrovascular disease | 433 | ICD10CM | I69.018 | Other symptoms and signs involving cognitive functions following nontraumatic subarachnoid hemorrhage |
| Cerebrovascular disease | 433 | ICD10CM | I69.362 | Other paralytic syndrome following cerebral infarction affecting left dominant side |
| Cerebrovascular disease | 433 | ICD10CM | I69.093 | Ataxia following nontraumatic subarachnoid hemorrhage |
| Cerebrovascular disease | 433 | ICD10CM | I69.00 | Unspecified sequelae of nontraumatic subarachnoid hemorrhage |
| Cerebrovascular disease | 433 | ICD10CM | I69.334 | Monoplegia of upper limb following cerebral infarction affecting left non-dominant side |
| Cerebrovascular disease | 433 | ICD10CM | I69.969 | Other paralytic syndrome following unspecified cerebrovascular disease affecting unspecified side |
| Cerebrovascular disease | 433 | ICD10CM | I69.020 | Aphasia following nontraumatic subarachnoid hemorrhage |
| Cerebrovascular disease | 433 | ICD9CM | 438.32 | Late effects of cerebrovascular disease, monoplegia of upper limb affecting nondominant side |
| Cerebrovascular disease | 433 | ICD9CM | 438.7 | Late effects of cerebrovascular disease, disturbances of vision |
| Cerebrovascular disease | 433 | ICD10CM | I69.31 | Cognitive deficits following cerebral infarction |
| Cerebrovascular disease | 433 | ICD10CM | I69.261 | Other paralytic syndrome following other nontraumatic intracranial hemorrhage affecting right dominant side |
| Cerebrovascular disease | 433 | ICD10CM | I69.352 | Hemiplegia and hemiparesis following cerebral infarction affecting left dominant side |
| Cerebrovascular disease | 433 | ICD10CM | I69.353 | Hemiplegia and hemiparesis following cerebral infarction affecting right non-dominant side |
| Cerebrovascular disease | 433 | ICD10CM | I69.834 | Monoplegia of upper limb following other cerebrovascular disease affecting left non-dominant side |
| Cerebrovascular disease | 433 | ICD10CM | I69.344 | Monoplegia of lower limb following cerebral infarction affecting left non-dominant side |
| Cerebrovascular disease | 433 | ICD9CM | 438.82 | Other late effects of cerebrovascular disease, dysphagia |
| Cerebrovascular disease | 433 | ICD10CM | I69.912 | Visuospatial deficit and spatial neglect following unspecified cerebrovascular disease |
| Cerebrovascular disease | 433 | ICD10CM | I69.2 | Sequelae of other nontraumatic intracranial hemorrhage |
| Cerebrovascular disease | 433 | ICD10CM | I69.315 | Cognitive social or emotional deficit following cerebral infarction |
| Cerebrovascular disease | 433 | ICD10CM | I69.312 | Visuospatial deficit and spatial neglect following cerebral infarction |
| Cerebrovascular disease | 433 | ICD10CM | I69.843 | Monoplegia of lower limb following other cerebrovascular disease affecting right non-dominant side |

| | | | | |
|---|---|---|---|---|
| Cerebrovascular disease | 433 | ICD10CM | I69.833 | Monoplegia of upper limb following other cerebrovascular disease affecting right non-dominant side |
| Cerebrovascular disease | 433 | ICD10CM | I69.831 | Monoplegia of upper limb following other cerebrovascular disease affecting right dominant side |
| Cerebrovascular disease | 433 | ICD10CM | I69.139 | Monoplegia of upper limb following nontraumatic intracerebral hemorrhage affecting unspecified side |
| Cerebrovascular disease | 433 | ICD10CM | I69.053 | Hemiplegia and hemiparesis following nontraumatic subarachnoid hemorrhage affecting right non-dominant side |
| Cerebrovascular disease | 433 | ICD10CM | I69.914 | Frontal lobe and executive function deficit following unspecified cerebrovascular disease |
| Cerebrovascular disease | 433 | ICD10CM | I69.26 | Other paralytic syndrome following other nontraumatic intracranial hemorrhage |
| Cerebrovascular disease | 433 | ICD10CM | I69.022 | Dysarthria following nontraumatic subarachnoid hemorrhage |
| Cerebrovascular disease | 433 | ICD10CM | I69.031 | Monoplegia of upper limb following nontraumatic subarachnoid hemorrhage affecting right dominant side |
| Cerebrovascular disease | 433 | ICD9CM | 438.12 | Late effects of cerebrovascular disease, dysphasia |
| Cerebrovascular disease | 433 | ICD10CM | I69.954 | Hemiplegia and hemiparesis following unspecified cerebrovascular disease affecting left non-dominant side |
| Cerebrovascular disease | 433 | ICD10CM | I69.243 | Monoplegia of lower limb following other nontraumatic intracranial hemorrhage affecting right non-dominant side |
| Cerebrovascular disease | 433 | ICD10CM | I69.828 | Other speech and language deficits following other cerebrovascular disease |
| Cerebrovascular disease | 433 | ICD10CM | I69.393 | Ataxia following cerebral infarction |
| Cerebrovascular disease | 433 | ICD10CM | I69.80 | Unspecified sequelae of other cerebrovascular disease |
| Cerebrovascular disease | 433 | ICD10CM | I69.361 | Other paralytic syndrome following cerebral infarction affecting right dominant side |
| Cerebrovascular disease | 433 | ICD10CM | I69.321 | Dysphasia following cerebral infarction |
| Cerebrovascular disease | 433 | ICD10CM | I69.222 | Dysarthria following other nontraumatic intracranial hemorrhage |
| Cerebrovascular disease | 433 | ICD9CM | 438.5 | Late effects of cerebrovascular disease, other paralytic syndrome affecting unspecified side |
| Cerebrovascular disease | 433 | ICD10CM | I69.091 | Dysphagia following nontraumatic subarachnoid hemorrhage |
| Cerebrovascular disease | 433 | ICD10CM | I69.322 | Dysarthria following cerebral infarction |
| Cerebrovascular disease | 433 | ICD10CM | I69.323 | Fluency disorder following cerebral infarction |
| Cerebrovascular disease | 433 | ICD9CM | 438.1 | Late effects of cerebrovascular disease, speech and language deficit, unspecified |

| | | | | |
|---|---|---|---|---|
| Cerebrovascular disease | 433 | ICD10CM | I69.82 | Speech and language deficits following other cerebrovascular disease |
| Cerebrovascular disease | 433 | ICD10CM | I69.819 | Unspecified symptoms and signs involving cognitive functions following other cerebrovascular disease |
| Cerebrovascular disease | 433 | ICD10CM | I69.343 | Monoplegia of lower limb following cerebral infarction affecting right non-dominant side |
| Cerebrovascular disease | 433 | ICD10CM | I69.920 | Aphasia following unspecified cerebrovascular disease |
| Cerebrovascular disease | 433 | ICD10CM | I69.269 | Other paralytic syndrome following other nontraumatic intracranial hemorrhage affecting unspecified side |
| Cerebrovascular disease | 433 | ICD10CM | I69.223 | Fluency disorder following other nontraumatic intracranial hemorrhage |
| Cerebrovascular disease | 433 | ICD10CM | I69.290 | Apraxia following other nontraumatic intracranial hemorrhage |
| Cerebrovascular disease | 433 | ICD10CM | I69.063 | Other paralytic syndrome following nontraumatic subarachnoid hemorrhage affecting right non-dominant side |
| Cerebrovascular disease | 433 | ICD10CM | I69.941 | Monoplegia of lower limb following unspecified cerebrovascular disease affecting right dominant side |
| Cerebrovascular disease | 433 | ICD10CM | I69.09 | Other sequelae of nontraumatic subarachnoid hemorrhage |
| Cerebrovascular disease | 433 | ICD9CM | 438.84 | Other late effects of cerebrovascular disease, ataxia |
| Cerebrovascular disease | 433 | ICD10CM | I69.990 | Apraxia following unspecified cerebrovascular disease |
| Cerebrovascular disease | 433 | ICD10CM | I69.991 | Dysphagia following unspecified cerebrovascular disease |
| Cerebrovascular disease | 433 | ICD10CM | I67.841 | Reversible cerebrovascular vasoconstriction syndrome |
| Cerebrovascular disease | 433 | ICD10CM | G45.1 | Carotid artery syndrome (hemispheric) |
| Cerebrovascular disease | 433 | ICD9CM | 435 | Transient cerebral ischemia |
| Cerebrovascular disease | 433 | ICD9CM | 437.3 | Cerebral aneurysm, nonruptured |
| Cerebrovascular disease | 433 | ICD10CM | I67.1 | Cerebral aneurysm, nonruptured |
| Cerebrovascular disease | 433 | ICD9CM | 438.83 | Other late effects of cerebrovascular disease, facial weakness |
| Cerebrovascular disease | 433 | ICD10CM | I69.942 | Monoplegia of lower limb following unspecified cerebrovascular disease affecting left dominant side |
| Cerebrovascular disease | 433 | ICD10CM | I69.234 | Monoplegia of upper limb following other nontraumatic intracranial hemorrhage affecting left non-dominant side |
| Cerebrovascular disease | 433 | ICD10CM | I69.864 | Other paralytic syndrome following other cerebrovascular disease affecting left non-dominant side |

| | | | | |
|---|---|---|---|---|
| Cerebrovascular disease | 433 | ICD10CM | I69.023 | Fluency disorder following nontraumatic subarachnoid hemorrhage |
| Cerebrovascular disease | 433 | ICD10CM | I69.331 | Monoplegia of upper limb following cerebral infarction affecting right dominant side |
| Cerebrovascular disease | 433 | ICD10CM | G43.601 | Persistent migraine aura with cerebral infarction, not intractable, with status migrainosus |
| Cerebrovascular disease | 433 | ICD10CM | G43.611 | Persistent migraine aura with cerebral infarction, intractable, with status migrainosus |
| Cerebrovascular disease | 433 | ICD10CM | G43.619 | Persistent migraine aura with cerebral infarction, intractable, without status migrainosus |
| Cerebrovascular disease | 433 | ICD9CM | 346.61 | Persistent migraine aura with cerebral infarction, with intractable migraine, so stated, without mention of status migrainosus |
| Cerebrovascular disease | 433 | ICD9CM | 346.6 | Persistent migraine aura with cerebral infarction |
| Cerebrovascular disease | 433 | ICD10CM | G43.609 | Persistent migraine aura with cerebral infarction, not intractable, without status migrainosus |
| Cerebrovascular disease | 433 | ICD10CM | I69.93 | Monoplegia of upper limb following unspecified cerebrovascular disease |
| Cerebrovascular disease | 433 | ICD10CM | I69.854 | Hemiplegia and hemiparesis following other cerebrovascular disease affecting left non-dominant side |
| Cerebrovascular disease | 433 | ICD10CM | I69.119 | Unspecified symptoms and signs involving cognitive functions following nontraumatic intracerebral hemorrhage |
| Cerebrovascular disease | 433 | ICD10CM | I69.351 | Hemiplegia and hemiparesis following cerebral infarction affecting right dominant side |
| Cerebrovascular disease | 433 | ICD10CM | I69.39 | Other sequelae of cerebral infarction |
| Cerebrovascular disease | 433 | ICD10CM | I69.16 | Other paralytic syndrome following nontraumatic intracerebral hemorrhage |
| Cerebrovascular disease | 433 | ICD10CM | I69.02 | Speech and language deficits following nontraumatic subarachnoid hemorrhage |
| Cerebrovascular disease | 433 | ICD9CM | 438.21 | Late effects of cerebrovascular disease, hemiplegia affecting dominant side |
| Cerebrovascular disease | 433 | ICD10CM | I69.114 | Frontal lobe and executive function deficit following nontraumatic intracerebral hemorrhage |
| Cerebrovascular disease | 433 | ICD10CM | I69.120 | Aphasia following nontraumatic intracerebral hemorrhage |
| Cerebrovascular disease | 433 | ICD10CM | I69.190 | Apraxia following nontraumatic intracerebral hemorrhage |
| Cerebrovascular disease | 433 | ICD10CM | I67.848 | Other cerebrovascular vasospasm and vasoconstriction |
| Cerebrovascular disease | 433 | ICD10CM | G45.9 | Transient cerebral ischemic attack, unspecified |
| Cerebrovascular disease | 433 | ICD9CM | 435.2 | Subclavian steal syndrome |
| Cerebrovascular disease | 433 | ICD10CM | I69.90 | Unspecified sequelae of unspecified cerebrovascular disease |

| | | | | |
|---|---|---|---|---|
| Cerebrovascular disease | 433 | ICD9CM | 438.5 | Other paralytic syndrome as late effect of cerebrovascular disease |
| Cerebrovascular disease | 433 | ICD10CM | I69.01 | Cognitive deficits following nontraumatic subarachnoid hemorrhage |
| Cerebrovascular disease | 433 | ICD10CM | I69.363 | Other paralytic syndrome following cerebral infarction affecting right non-dominant side |
| Cerebrovascular disease | 433 | ICD10CM | I69.865 | Other paralytic syndrome following other cerebrovascular disease, bilateral |
| Cerebrovascular disease | 433 | ICD9CM | 438.42 | Late effects of cerebrovascular disease, monoplegia of lower limb affecting nondominant side |
| Cerebrovascular disease | 433 | ICD10CM | I69.263 | Other paralytic syndrome following other nontraumatic intracranial hemorrhage affecting right non-dominant side |
| Cerebrovascular disease | 433 | ICD10CM | I69.390 | Apraxia following cerebral infarction |
| Cerebrovascular disease | 433 | ICD10CM | I69.191 | Dysphagia following nontraumatic intracerebral hemorrhage |
| Cerebrovascular disease | 433 | ICD9CM | 435.1 | Vertebral artery syndrome |
| Cerebrovascular disease | 433 | ICD10CM | G45.2 | Multiple and bilateral precerebral artery syndromes |
| Cerebrovascular disease | 433 | ICD10CM | G46.1 | Anterior cerebral artery syndrome |
| Cerebrovascular disease | 433 | ICD9CM | 346.6 | Persistent migraine aura with cerebral infarction, without mention of intractable migraine without mention of status migrainosus |
| Cerebrovascular disease | 433 | ICD10CM | I69.19 | Other sequelae of nontraumatic intracerebral hemorrhage |
| Cerebrovascular disease | 433 | ICD10CM | I69.365 | Other paralytic syndrome following cerebral infarction, bilateral |
| Cerebrovascular disease | 433 | ICD10CM | I69.328 | Other speech and language deficits following cerebral infarction |
| Cerebrovascular disease | 433 | ICD10CM | I69.24 | Monoplegia of lower limb following other nontraumatic intracranial hemorrhage |
| Cerebrovascular disease | 433 | ICD10CM | I69.313 | Psychomotor deficit following cerebral infarction |
| Cerebrovascular disease | 433 | ICD10CM | I69.164 | Other paralytic syndrome following nontraumatic intracerebral hemorrhage affecting left non-dominant side |
| Cerebrovascular disease | 433 | ICD10CM | I69.3 | Sequelae of cerebral infarction |
| Cerebrovascular disease | 433 | ICD10CM | I69.934 | Monoplegia of upper limb following unspecified cerebrovascular disease affecting left non-dominant side |
| Cerebrovascular disease | 433 | ICD10CM | I69.35 | Hemiplegia and hemiparesis following cerebral infarction |
| Cerebrovascular disease | 433 | ICD10CM | I69.354 | Hemiplegia and hemiparesis following cerebral infarction affecting left non-dominant side |
| Cerebrovascular disease | 433 | ICD9CM | 436 | Acute, but ill-defined, cerebrovascular disease |

| | | | | |
|---|---|---|---|---|
| Cerebrovascular disease | 433 | ICD10CM | G43.61 | Persistent migraine aura with cerebral infarction, intractable |
| Cerebrovascular disease | 433 | ICD10CM | I69.110 | Attention and concentration deficit following nontraumatic intracerebral hemorrhage |
| Cerebrovascular disease | 433 | ICD10CM | G43.60 | Persistent migraine aura with cerebral infarction, not intractable |
| Cerebrovascular disease | 433 | ICD9CM | 438.8 | Other late effects of cerebrovascular disease |
| Cerebrovascular disease | 433 | ICD10CM | I69.964 | Other paralytic syndrome following unspecified cerebrovascular disease affecting left non-dominant side |
| Cerebrovascular disease | 433 | ICD10CM | I69.213 | Psychomotor deficit following other nontraumatic intracranial hemorrhage |
| Cerebrovascular disease | 433 | ICD10CM | I69.863 | Other paralytic syndrome following other cerebrovascular disease affecting right non-dominant side |
| Cerebrovascular disease | 433 | ICD10CM | I69.064 | Other paralytic syndrome following nontraumatic subarachnoid hemorrhage affecting left non-dominant side |
| Cerebrovascular disease | 433 | ICD10CM | I69.310 | Attention and concentration deficit following cerebral infarction |
| Cerebrovascular disease | 433 | ICD10CM | I69.84 | Monoplegia of lower limb following other cerebrovascular disease |
| Cerebrovascular disease | 433 | ICD10CM | I69.249 | Monoplegia of lower limb following other nontraumatic intracranial hemorrhage affecting unspecified side |
| Cerebrovascular disease | 433 | ICD10CM | I69.391 | Dysphagia following cerebral infarction |
| Cerebrovascular disease | 433 | ICD10CM | I69.849 | Monoplegia of lower limb following other cerebrovascular disease affecting unspecified side |
| Cerebrovascular disease | 433 | ICD10CM | I69.010 | Attention and concentration deficit following nontraumatic subarachnoid hemorrhage |
| Cerebrovascular disease | 433 | ICD10CM | I69.349 | Monoplegia of lower limb following cerebral infarction affecting unspecified side |
| Cerebrovascular disease | 433 | ICD10CM | I69.118 | Other symptoms and signs involving cognitive functions following nontraumatic intracerebral hemorrhage |
| Cerebrovascular disease | 433 | ICD10CM | I69.262 | Other paralytic syndrome following other nontraumatic intracranial hemorrhage affecting left dominant side |
| Cerebrovascular disease | 433 | ICD10CM | I69.239 | Monoplegia of upper limb following other nontraumatic intracranial hemorrhage affecting unspecified side |
| Cerebrovascular disease | 433 | ICD9CM | 438.6 | Late effects of cerebrovascular disease, alterations of sensations |
| Cerebrovascular disease | 433 | ICD9CM | 438.41 | Late effects of cerebrovascular disease, monoplegia of lower limb affecting dominant side |
| Cerebrovascular disease | 433 | ICD9CM | 438.53 | Late effects of cerebrovascular disease, other paralytic syndrome, bilateral |

| | | | | |
|---|---|---|---|---|
| Cerebrovascular disease | 433 | ICD10CM | I69.23 | Monoplegia of upper limb following other nontraumatic intracranial hemorrhage |
| Cerebrovascular disease | 433 | ICD10CM | I69.818 | Other symptoms and signs involving cognitive functions following other cerebrovascular disease |
| Cerebrovascular disease | 433 | ICD10CM | I69.042 | Monoplegia of lower limb following nontraumatic subarachnoid hemorrhage affecting left dominant side |
| Cerebrovascular disease | 433 | ICD10CM | I69.890 | Apraxia following other cerebrovascular disease |
| Cerebrovascular disease | 433 | ICD10CM | I69.99 | Other sequelae of unspecified cerebrovascular disease |
| Cerebrovascular disease | 433 | ICD10CM | I69.869 | Other paralytic syndrome following other cerebrovascular disease affecting unspecified side |
| Cerebrovascular disease | 433 | ICD9CM | 438.13 | Late effects of cerebrovascular disease, dysarthria |
| Cerebrovascular disease | 433 | ICD10CM | I69.32 | Speech and language deficits following cerebral infarction |
| Cerebrovascular disease | 433 | ICD10CM | I69.059 | Hemiplegia and hemiparesis following nontraumatic subarachnoid hemorrhage affecting unspecified side |
| Cerebrovascular disease | 433 | ICD10CM | G43.6 | Persistent migraine aura with cerebral infarction |
| Cerebrovascular disease | 433 | ICD10CM | I69.011 | Memory deficit following nontraumatic subarachnoid hemorrhage |
| Cerebrovascular disease | 433 | ICD10CM | I69.06 | Other paralytic syndrome following nontraumatic subarachnoid hemorrhage |
| Cerebrovascular disease | 433 | ICD10CM | I69.241 | Monoplegia of lower limb following other nontraumatic intracranial hemorrhage affecting right dominant side |
| Cerebrovascular disease | 433 | ICD10CM | I69.898 | Other sequelae of other cerebrovascular disease |
| Cerebrovascular disease | 433 | ICD10CM | I69.232 | Monoplegia of upper limb following other nontraumatic intracranial hemorrhage affecting left dominant side |
| Cerebrovascular disease | 433 | ICD10CM | I69.231 | Monoplegia of upper limb following other nontraumatic intracranial hemorrhage affecting right dominant side |
| Cerebrovascular disease | 433 | ICD10CM | I69.939 | Monoplegia of upper limb following unspecified cerebrovascular disease affecting unspecified side |
| Cerebrovascular disease | 433 | ICD10CM | I69.30 | Unspecified sequelae of cerebral infarction |
| Cerebrovascular disease | 433 | ICD10CM | I69.242 | Monoplegia of lower limb following other nontraumatic intracranial hemorrhage affecting left dominant side |
| Cerebrovascular disease | 433 | ICD10CM | I69.33 | Monoplegia of upper limb following cerebral infarction |
| Cerebrovascular disease | 433 | ICD10CM | I69.859 | Hemiplegia and hemiparesis following other cerebrovascular disease affecting unspecified side |

| | | | | | |
|---|---|---|---|---|---|
| | Cerebrovascular disease | 433 | ICD10CM | I69.862 | Other paralytic syndrome following other cerebrovascular disease affecting left dominant side |
| | Cerebral aneurysm | 433.5 | ICD9CM | 437.3 | Cerebral aneurysm, nonruptured |
| | Cerebral aneurysm | 433.5 | ICD10CM | I67.1 | Cerebral aneurysm, nonruptured |
| | Peripheral vascular disease | 443 | ICD9CM | 443 | Other peripheral vascular disease |
| | Peripheral vascular disease | 443 | ICD10CM | I73.01 | Raynaud's syndrome with gangrene |
| | Peripheral vascular disease | 443 | ICD10CM | I73.00 | Raynaud's syndrome without gangrene |
| | Peripheral vascular disease | 443 | ICD10CM | I73.0 | Raynaud's syndrome |
| | Peripheral vascular disease | 443 | ICD9CM | 443 | Raynaud's syndrome |
| | Peripheral vascular disease | 443 | ICD10CM | I73.8 | Other specified peripheral vascular diseases |
| | Peripheral vascular disease | 443 | ICD9CM | 443.82 | Erythromelalgia |
| | Peripheral vascular disease | 443 | ICD10CM | I73.81 | Erythromelalgia |
| | Peripheral vascular disease | 443 | ICD10CM | I73.89 | Other specified peripheral vascular diseases |
| | Peripheral vascular disease | 443 | ICD10CM | I73 | Other peripheral vascular diseases |
| | Peripheral vascular disease | 443 | ICD9CM | 443.81 | Peripheral angiopathy in diseases classified elsewhere |
| | Peripheral vascular disease | 443 | ICD10CM | I79.1 | Aortitis in diseases classified elsewhere |
| | Peripheral vascular disease | 443 | ICD10CM | I79.8 | Other disorders of arteries, arterioles and capillaries in diseases classified elsewhere |
| | Peripheral vascular disease | 443 | ICD10CM | I73.9 | Peripheral vascular disease, unspecified |
| | Peripheral vascular disease | 443 | ICD9CM | 443.89 | Other specified peripheral vascular diseases |
| | Peripheral vascular disease | 443 | ICD9CM | 443.8 | Other specified peripheral vascular diseases |
| | Peripheral vascular disease | 443 | ICD9CM | 443.9 | Peripheral vascular disease, unspecified |
| **Respiratory System** | Asthma | 495 | ICD9CM | 493 | Asthma |
| | Asthma | 495 | ICD10CM | J45.998 | Other asthma |
| | Asthma | 495 | ICD10CM | J45 | Asthma |
| | Asthma | 495 | ICD10CM | J45.40 | Moderate persistent asthma, uncomplicated |
| | Asthma | 495 | ICD9CM | 493.9 | Asthma, unspecified |
| | Asthma | 495 | ICD9CM | 493 | Extrinsic asthma, unspecified |
| | Asthma | 495 | ICD9CM | 493.8 | Other forms of asthma |
| | Asthma | 495 | ICD9CM | 493.82 | Cough variant asthma |
| | Asthma | 495 | ICD10CM | J45.909 | Unspecified asthma, uncomplicated |

| | | | | |
|---|---|---|---|---|
| Asthma | 495 | ICD10CM | J45.30 | Mild persistent asthma, uncomplicated |
| Asthma | 495 | ICD10CM | J45.90 | Unspecified asthma |
| Asthma | 495 | ICD9CM | 493.1 | Intrinsic asthma, unspecified |
| Asthma | 495 | ICD10CM | J45.50 | Severe persistent asthma, uncomplicated |
| Asthma | 495 | ICD10CM | J45.20 | Mild intermittent asthma, uncomplicated |
| Asthma | 495 | ICD10CM | J45.991 | Cough variant asthma |
| Asthma | 495 | ICD9CM | 493 | Extrinsic asthma |
| Asthma | 495 | ICD9CM | 493.9 | Asthma, unspecified type, unspecified |
| Asthma | 495 | ICD9CM | 493.1 | Intrinsic asthma |
| Asthma | 495 | ICD9CM | 493.2 | Chronic obstructive asthma, unspecified |
| Asthma | 495 | ICD9CM | 493.2 | Chronic obstructive asthma |
| Asthma | 495 | ICD10CM | J45.21 | Mild intermittent asthma with (acute) exacerbation |
| Asthma | 495 | ICD9CM | 493.01 | Extrinsic asthma with status asthmaticus |
| Asthma | 495 | ICD9CM | 493.21 | Chronic obstructive asthma with status asthmaticus |
| Asthma | 495 | ICD9CM | 493.22 | Chronic obstructive asthma with (acute) exacerbation |
| Asthma | 495 | ICD10CM | J45.31 | Mild persistent asthma with (acute) exacerbation |
| Asthma | 495 | ICD9CM | 493.81 | Exercise induced bronchospasm |
| Asthma | 495 | ICD9CM | 493.91 | Asthma, unspecified type, with status asthmaticus |
| Asthma | 495 | ICD10CM | J45.990 | Exercise induced bronchospasm |
| Asthma | 495 | ICD10CM | J45.22 | Mild intermittent asthma with status asthmaticus |
| Asthma | 495 | ICD9CM | 493.02 | Extrinsic asthma with (acute) exacerbation |
| Asthma | 495 | ICD10CM | J45.41 | Moderate persistent asthma with (acute) exacerbation |
| Asthma | 495 | ICD10CM | J45.42 | Moderate persistent asthma with status asthmaticus |
| Asthma | 495 | ICD9CM | 493.11 | Intrinsic asthma with status asthmaticus |
| Asthma | 495 | ICD9CM | 493.12 | Intrinsic asthma with (acute) exacerbation |
| Asthma | 495 | ICD10CM | J45.32 | Mild persistent asthma with status asthmaticus |
| Asthma | 495 | ICD10CM | J45.51 | Severe persistent asthma with (acute) exacerbation |
| Asthma | 495 | ICD10CM | J45.902 | Unspecified asthma with status asthmaticus |
| Asthma | 495 | ICD9CM | 493.92 | Asthma, unspecified type, with (acute) exacerbation |
| Asthma | 495 | ICD10CM | J45.901 | Unspecified asthma with (acute) exacerbation |
| Asthma | 495 | ICD10CM | J45.52 | Severe persistent asthma with status asthmaticus |
| Obstructive chronic bronchitis | 496.21 | ICD10CM | J44.0 | Chronic obstructive pulmonary disease with (acute) lower respiratory infection |
| Obstructive chronic bronchitis | 496.21 | ICD9CM | 491.22 | Obstructive chronic bronchitis with acute bronchitis |

| | | | | | |
|---|---|---|---|---|---|
| | Obstructive chronic bronchitis | 496.21 | ICD9CM | 491.2 | Obstructive chronic bronchitis |
| | Obstructive chronic bronchitis | 496.21 | ICD10CM | J44.89 | Other specified chronic obstructive pulmonary disease |
| | Obstructive chronic bronchitis | 496.21 | ICD9CM | 491.21 | Obstructive chronic bronchitis with (acute) exacerbation |
| | Obstructive chronic bronchitis | 496.21 | ICD10CM | J44.1 | Chronic obstructive pulmonary disease with (acute) exacerbation |
| | Obstructive chronic bronchitis | 496.21 | ICD9CM | 491.2 | Obstructive chronic bronchitis without exacerbation |
| | Obstructive chronic bronchitis | 496.21 | ICD10CM | J44.81 | Bronchiolitis obliterans and bronchiolitis obliterans syndrome |
| | Obstructive chronic bronchitis | 496.21 | ICD10CM | J44 | Other chronic obstructive pulmonary disease |
| **Digestive system** | GERD | 530.11 | ICD10CM | K21.9 | Gastro-esophageal reflux disease without esophagitis |
| | GERD | 530.11 | ICD10CM | K21 | Gastro-esophageal reflux disease |
| | GERD | 530.11 | ICD9CM | 530.81 | Esophageal reflux |
| | Inflammatory bowel disease and other gastroenteritis and colitis | 555 | ICD9CM | 555 | Regional enteritis |
| | Inflammatory bowel disease and other gastroenteritis and colitis | 555 | ICD10CM | K50.811 | Crohn's disease of both small and large intestine with rectal bleeding |
| | Inflammatory bowel disease and other gastroenteritis and colitis | 555 | ICD10CM | K50.111 | Crohn's disease of large intestine with rectal bleeding |
| | Inflammatory bowel disease and other gastroenteritis and colitis | 555 | ICD10CM | K50.113 | Crohn's disease of large intestine with fistula |
| | Inflammatory bowel disease and other gastroenteritis and colitis | 555 | ICD10CM | K50.118 | Crohn's disease of large intestine with other complication |
| | Inflammatory bowel disease and other gastroenteritis and colitis | 555 | ICD9CM | 555 | Regional enteritis of small intestine |
| | Inflammatory bowel disease and other gastroenteritis and colitis | 555 | ICD10CM | K50.014 | Crohn's disease of small intestine with abscess |
| | Inflammatory bowel disease and other gastroenteritis and colitis | 555 | ICD10CM | K50.112 | Crohn's disease of large intestine with intestinal obstruction |

| | | | | |
|---|---|---|---|---|
| Inflammatory bowel disease and other gastroenteritis and colitis | 555 | ICD9CM | 555.1 | Regional enteritis of large intestine |
| Inflammatory bowel disease and other gastroenteritis and colitis | 555 | ICD10CM | K50.914 | Crohn's disease, unspecified, with abscess |
| Inflammatory bowel disease and other gastroenteritis and colitis | 555 | ICD10CM | K50.812 | Crohn's disease of both small and large intestine with intestinal obstruction |
| Inflammatory bowel disease and other gastroenteritis and colitis | 555 | ICD10CM | K50.818 | Crohn's disease of both small and large intestine with other complication |
| Inflammatory bowel disease and other gastroenteritis and colitis | 555 | ICD10CM | K50.10 | Crohn's disease of large intestine without complications |
| Inflammatory bowel disease and other gastroenteritis and colitis | 555 | ICD9CM | 555.9 | Regional enteritis of unspecified site |
| Inflammatory bowel disease and other gastroenteritis and colitis | 555 | ICD10CM | K50.90 | Crohn's disease, unspecified, without complications |
| Inflammatory bowel disease and other gastroenteritis and colitis | 555 | ICD10CM | K50.80 | Crohn's disease of both small and large intestine without complications |
| Inflammatory bowel disease and other gastroenteritis and colitis | 555 | ICD10CM | K50.119 | Crohn's disease of large intestine with unspecified complications |
| Inflammatory bowel disease and other gastroenteritis and colitis | 555 | ICD10CM | K50.00 | Crohn's disease of small intestine without complications |
| Inflammatory bowel disease and other gastroenteritis and colitis | 555 | ICD10CM | K50.019 | Crohn's disease of small intestine with unspecified complications |
| Inflammatory bowel disease and other gastroenteritis and colitis | 555 | ICD10CM | K50.011 | Crohn's disease of small intestine with rectal bleeding |
| Inflammatory bowel disease and other gastroenteritis and colitis | 555 | ICD10CM | K50.813 | Crohn's disease of both small and large intestine with fistula |

| | | | | |
|---|---|---|---|---|
| Inflammatory bowel disease and other gastroenteritis and colitis | 555 | ICD10CM | K50.9 | Crohn's disease, unspecified |
| Inflammatory bowel disease and other gastroenteritis and colitis | 555 | ICD10CM | K50.012 | Crohn's disease of small intestine with intestinal obstruction |
| Inflammatory bowel disease and other gastroenteritis and colitis | 555 | ICD10CM | K50.913 | Crohn's disease, unspecified, with fistula |
| Inflammatory bowel disease and other gastroenteritis and colitis | 555 | ICD10CM | K50.0 | Crohn's disease of small intestine |
| Inflammatory bowel disease and other gastroenteritis and colitis | 555 | ICD10CM | K50.819 | Crohn's disease of both small and large intestine with unspecified complications |
| Inflammatory bowel disease and other gastroenteritis and colitis | 555 | ICD10CM | K50.918 | Crohn's disease, unspecified, with other complication |
| Inflammatory bowel disease and other gastroenteritis and colitis | 555 | ICD10CM | K50.919 | Crohn's disease, unspecified, with unspecified complications |
| Inflammatory bowel disease and other gastroenteritis and colitis | 555 | ICD9CM | 555.2 | Regional enteritis of small intestine with large intestine |
| Inflammatory bowel disease and other gastroenteritis and colitis | 555 | ICD10CM | K50.912 | Crohn's disease, unspecified, with intestinal obstruction |
| Inflammatory bowel disease and other gastroenteritis and colitis | 555 | ICD10CM | K50.013 | Crohn's disease of small intestine with fistula |
| Inflammatory bowel disease and other gastroenteritis and colitis | 555 | ICD10CM | K50.114 | Crohn's disease of large intestine with abscess |
| Inflammatory bowel disease and other gastroenteritis and colitis | 555 | ICD10CM | K50.814 | Crohn's disease of both small and large intestine with abscess |
| Inflammatory bowel disease and other gastroenteritis and colitis | 555 | ICD10CM | K50.1 | Crohn's disease of large intestine |

| | | | | |
|---|---|---|---|---|
| Inflammatory bowel disease and other gastroenteritis and colitis | 555 | ICD10CM | K50.911 | Crohn's disease, unspecified, with rectal bleeding |
| Inflammatory bowel disease and other gastroenteritis and colitis | 555 | ICD10CM | K50 | Crohn's disease [regional enteritis] |
| Inflammatory bowel disease and other gastroenteritis and colitis | 555 | ICD10CM | K50.018 | Crohn's disease of small intestine with other complication |
| Inflammatory bowel disease and other gastroenteritis and colitis | 555 | ICD10CM | K51.811 | Other ulcerative colitis with rectal bleeding |
| Inflammatory bowel disease and other gastroenteritis and colitis | 555 | ICD10CM | K51.8 | Other ulcerative colitis |
| Inflammatory bowel disease and other gastroenteritis and colitis | 555 | ICD10CM | K51.318 | Ulcerative (chronic) rectosigmoiditis with other complication |
| Inflammatory bowel disease and other gastroenteritis and colitis | 555 | ICD9CM | 556 | Ulcerative colitis |
| Inflammatory bowel disease and other gastroenteritis and colitis | 555 | ICD9CM | 556.5 | Left-sided ulcerative (chronic) colitis |
| Inflammatory bowel disease and other gastroenteritis and colitis | 555 | ICD10CM | K51.518 | Left sided colitis with other complication |
| Inflammatory bowel disease and other gastroenteritis and colitis | 555 | ICD10CM | K51.50 | Left sided colitis without complications |
| Inflammatory bowel disease and other gastroenteritis and colitis | 555 | ICD10CM | K51.511 | Left sided colitis with rectal bleeding |
| Inflammatory bowel disease and other gastroenteritis and colitis | 555 | ICD10CM | K51.2 | Ulcerative (chronic) proctitis |
| Inflammatory bowel disease and other gastroenteritis and colitis | 555 | ICD10CM | K51.20 | Ulcerative (chronic) proctitis without complications |

| | | | | |
|---|---|---|---|---|
| Inflammatory bowel disease and other gastroenteritis and colitis | 555 | ICD9CM | 556 | Ulcerative (chronic) enterocolitis |
| Inflammatory bowel disease and other gastroenteritis and colitis | 555 | ICD10CM | K51.519 | Left sided colitis with unspecified complications |
| Inflammatory bowel disease and other gastroenteritis and colitis | 555 | ICD10CM | K51.212 | Ulcerative (chronic) proctitis with intestinal obstruction |
| Inflammatory bowel disease and other gastroenteritis and colitis | 555 | ICD9CM | 556.6 | Universal ulcerative (chronic) colitis |
| Inflammatory bowel disease and other gastroenteritis and colitis | 555 | ICD10CM | K51.218 | Ulcerative (chronic) proctitis with other complication |
| Inflammatory bowel disease and other gastroenteritis and colitis | 555 | ICD10CM | K51.013 | Ulcerative (chronic) pancolitis with fistula |
| Inflammatory bowel disease and other gastroenteritis and colitis | 555 | ICD10CM | K51.213 | Ulcerative (chronic) proctitis with fistula |
| Inflammatory bowel disease and other gastroenteritis and colitis | 555 | ICD10CM | K51.219 | Ulcerative (chronic) proctitis with unspecified complications |
| Inflammatory bowel disease and other gastroenteritis and colitis | 555 | ICD10CM | K51.214 | Ulcerative (chronic) proctitis with abscess |
| Inflammatory bowel disease and other gastroenteritis and colitis | 555 | ICD10CM | K51.011 | Ulcerative (chronic) pancolitis with rectal bleeding |
| Inflammatory bowel disease and other gastroenteritis and colitis | 555 | ICD10CM | K51.0 | Ulcerative (chronic) pancolitis |
| Inflammatory bowel disease and other gastroenteritis and colitis | 555 | ICD10CM | K51.514 | Left sided colitis with abscess |
| Inflammatory bowel disease and other gastroenteritis and colitis | 555 | ICD10CM | K51.019 | Ulcerative (chronic) pancolitis with unspecified complications |

| | | | | |
|---|---|---|---|---|
| Inflammatory bowel disease and other gastroenteritis and colitis | 555 | ICD10CM | K51.512 | Left sided colitis with intestinal obstruction |
| Inflammatory bowel disease and other gastroenteritis and colitis | 555 | ICD10CM | K51.513 | Left sided colitis with fistula |
| Inflammatory bowel disease and other gastroenteritis and colitis | 555 | ICD10CM | K51.014 | Ulcerative (chronic) pancolitis with abscess |
| Inflammatory bowel disease and other gastroenteritis and colitis | 555 | ICD10CM | K51.012 | Ulcerative (chronic) pancolitis with intestinal obstruction |
| Inflammatory bowel disease and other gastroenteritis and colitis | 555 | ICD10CM | K51.211 | Ulcerative (chronic) proctitis with rectal bleeding |
| Inflammatory bowel disease and other gastroenteritis and colitis | 555 | ICD10CM | K51.00 | Ulcerative (chronic) pancolitis without complications |
| Inflammatory bowel disease and other gastroenteritis and colitis | 555 | ICD9CM | 556.1 | Ulcerative (chronic) ileocolitis |
| Inflammatory bowel disease and other gastroenteritis and colitis | 555 | ICD10CM | K51.018 | Ulcerative (chronic) pancolitis with other complication |
| Inflammatory bowel disease and other gastroenteritis and colitis | 555 | ICD9CM | 556.2 | Ulcerative (chronic) proctitis |
| Inflammatory bowel disease and other gastroenteritis and colitis | 555 | ICD10CM | K51.918 | Ulcerative colitis, unspecified with other complication |
| Inflammatory bowel disease and other gastroenteritis and colitis | 555 | ICD10CM | K51.319 | Ulcerative (chronic) rectosigmoiditis with unspecified complications |
| Inflammatory bowel disease and other gastroenteritis and colitis | 555 | ICD10CM | K51.3 | Ulcerative (chronic) rectosigmoiditis |
| Inflammatory bowel disease and other gastroenteritis and colitis | 555 | ICD10CM | K51.814 | Other ulcerative colitis with abscess |

| | | | | |
|---|---|---|---|---|
| Inflammatory bowel disease and other gastroenteritis and colitis | 555 | ICD10CM | K51.914 | Ulcerative colitis, unspecified with abscess |
| Inflammatory bowel disease and other gastroenteritis and colitis | 555 | ICD10CM | K51.919 | Ulcerative colitis, unspecified with unspecified complications |
| Inflammatory bowel disease and other gastroenteritis and colitis | 555 | ICD10CM | K51.80 | Other ulcerative colitis without complications |
| Inflammatory bowel disease and other gastroenteritis and colitis | 555 | ICD10CM | K51.4 | Inflammatory polyps of colon |
| Inflammatory bowel disease and other gastroenteritis and colitis | 555 | ICD10CM | K51.411 | Inflammatory polyps of colon with rectal bleeding |
| Inflammatory bowel disease and other gastroenteritis and colitis | 555 | ICD10CM | K51.913 | Ulcerative colitis, unspecified with fistula |
| Inflammatory bowel disease and other gastroenteritis and colitis | 555 | ICD10CM | K51.312 | Ulcerative (chronic) rectosigmoiditis with intestinal obstruction |
| Inflammatory bowel disease and other gastroenteritis and colitis | 555 | ICD10CM | K51.414 | Inflammatory polyps of colon with abscess |
| Inflammatory bowel disease and other gastroenteritis and colitis | 555 | ICD10CM | K51.412 | Inflammatory polyps of colon with intestinal obstruction |
| Inflammatory bowel disease and other gastroenteritis and colitis | 555 | ICD10CM | K51.912 | Ulcerative colitis, unspecified with intestinal obstruction |
| Inflammatory bowel disease and other gastroenteritis and colitis | 555 | ICD10CM | K51.413 | Inflammatory polyps of colon with fistula |
| Inflammatory bowel disease and other gastroenteritis and colitis | 555 | ICD10CM | K51.812 | Other ulcerative colitis with intestinal obstruction |
| Inflammatory bowel disease and other gastroenteritis and colitis | 555 | ICD10CM | K51.813 | Other ulcerative colitis with fistula |

| | | | | |
|---|---|---|---|---|
| Inflammatory bowel disease and other gastroenteritis and colitis | 555 | ICD10CM | K51.419 | Inflammatory polyps of colon with unspecified complications |
| Inflammatory bowel disease and other gastroenteritis and colitis | 555 | ICD10CM | K51 | Ulcerative colitis |
| Inflammatory bowel disease and other gastroenteritis and colitis | 555 | ICD9CM | 556.9 | Ulcerative colitis, unspecified |
| Inflammatory bowel disease and other gastroenteritis and colitis | 555 | ICD10CM | K51.314 | Ulcerative (chronic) rectosigmoiditis with abscess |
| Inflammatory bowel disease and other gastroenteritis and colitis | 555 | ICD9CM | 556.4 | Pseudopolyposis of colon |
| Inflammatory bowel disease and other gastroenteritis and colitis | 555 | ICD10CM | K51.30 | Ulcerative (chronic) rectosigmoiditis without complications |
| Inflammatory bowel disease and other gastroenteritis and colitis | 555 | ICD10CM | K51.313 | Ulcerative (chronic) rectosigmoiditis with fistula |
| Inflammatory bowel disease and other gastroenteritis and colitis | 555 | ICD10CM | K51.9 | Ulcerative colitis, unspecified |
| Inflammatory bowel disease and other gastroenteritis and colitis | 555 | ICD10CM | K51.40 | Inflammatory polyps of colon without complications |
| Inflammatory bowel disease and other gastroenteritis and colitis | 555 | ICD10CM | K51.311 | Ulcerative (chronic) rectosigmoiditis with rectal bleeding |
| Inflammatory bowel disease and other gastroenteritis and colitis | 555 | ICD9CM | 556.8 | Other ulcerative colitis |
| Inflammatory bowel disease and other gastroenteritis and colitis | 555 | ICD9CM | 556.3 | Ulcerative (chronic) proctosigmoiditis |
| Inflammatory bowel disease and other gastroenteritis and colitis | 555 | ICD10CM | K51.90 | Ulcerative colitis, unspecified, without complications |

| | | | | |
|---|---|---|---|---|
| Inflammatory bowel disease and other gastroenteritis and colitis | 555 | ICD10CM | K51.911 | Ulcerative colitis, unspecified with rectal bleeding |
| Inflammatory bowel disease and other gastroenteritis and colitis | 555 | ICD10CM | K51.819 | Other ulcerative colitis with unspecified complications |
| Inflammatory bowel disease and other gastroenteritis and colitis | 555 | ICD10CM | K51.818 | Other ulcerative colitis with other complication |
| Inflammatory bowel disease and other gastroenteritis and colitis | 555 | ICD10CM | K51.418 | Inflammatory polyps of colon with other complication |
| Chronic liver disease and cirrhosis | 571 | ICD9CM | 571 | Chronic liver disease and cirrhosis |
| Chronic liver disease and cirrhosis | 571 | ICD10CM | K75.81 | Nonalcoholic steatohepatitis (NASH) |
| Chronic liver disease and cirrhosis | 571 | ICD9CM | 571.8 | Other chronic nonalcoholic liver disease |
| Chronic liver disease and cirrhosis | 571 | ICD10CM | K76.82 | Hepatic encephalopathy |
| Chronic liver disease and cirrhosis | 571 | ICD10CM | K76.8 | Other specified diseases of liver |
| Chronic liver disease and cirrhosis | 571 | ICD10CM | K74.1 | Hepatic sclerosis |
| Chronic liver disease and cirrhosis | 571 | ICD10CM | K76.89 | Other specified diseases of liver |
| Chronic liver disease and cirrhosis | 571 | ICD10CM | K76.0 | Fatty (change of) liver, not elsewhere classified |
| Chronic liver disease and cirrhosis | 571 | ICD10CM | K74.2 | Hepatic fibrosis with hepatic sclerosis |
| Chronic liver disease and cirrhosis | 571 | ICD9CM | 571.9 | Unspecified chronic liver disease without mention of alcohol |
| Chronic liver disease and cirrhosis | 571 | ICD10CM | K72.9 | Hepatic failure, unspecified |
| Chronic liver disease and cirrhosis | 571 | ICD10CM | K70.41 | Alcoholic hepatic failure with coma |
| Chronic liver disease and cirrhosis | 571 | ICD10CM | K75.1 | Phlebitis of portal vein |
| Chronic liver disease and cirrhosis | 571 | ICD10CM | K72.91 | Hepatic failure, unspecified with coma |
| Chronic liver disease and cirrhosis | 571 | ICD9CM | 572 | Abscess of liver |
| Chronic liver disease and cirrhosis | 571 | ICD10CM | K71.11 | Toxic liver disease with hepatic necrosis, with coma |
| Chronic liver disease and cirrhosis | 571 | ICD9CM | 572.4 | Hepatorenal syndrome |
| Chronic liver disease and cirrhosis | 571 | ICD10CM | K72.10 | Chronic hepatic failure without coma |

| | | | | |
|---|---|---|---|---|
| Chronic liver disease and cirrhosis | 571 | ICD9CM | 572 | Liver abscess and sequelae of chronic liver disease |
| Chronic liver disease and cirrhosis | 571 | ICD9CM | 572.3 | Portal hypertension |
| Chronic liver disease and cirrhosis | 571 | ICD10CM | K74.6 | Other and unspecified cirrhosis of liver |
| Chronic liver disease and cirrhosis | 571 | ICD10CM | K74.0 | Hepatic fibrosis |
| Chronic liver disease and cirrhosis | 571 | ICD10CM | K74.60 | Unspecified cirrhosis of liver |
| Chronic liver disease and cirrhosis | 571 | ICD9CM | 571.5 | Cirrhosis of liver without mention of alcohol |
| Chronic liver disease and cirrhosis | 571 | ICD10CM | K74.69 | Other cirrhosis of liver |
| Chronic liver disease and cirrhosis | 571 | ICD10CM | K74.02 | Hepatic fibrosis, advanced fibrosis |
| Chronic liver disease and cirrhosis | 571 | ICD10CM | K74.01 | Hepatic fibrosis, early fibrosis |
| Chronic liver disease and cirrhosis | 571 | ICD10CM | K74.00 | Hepatic fibrosis, unspecified |
| Chronic liver disease and cirrhosis | 571 | ICD9CM | 572.2 | Hepatic encephalopathy |
| Chronic liver disease and cirrhosis | 571 | ICD10CM | K72 | Hepatic failure, not elsewhere classified |
| Chronic liver disease and cirrhosis | 571 | ICD10CM | K76.7 | Hepatorenal syndrome |
| Chronic liver disease and cirrhosis | 571 | ICD10CM | K72.1 | Chronic hepatic failure |
| Chronic liver disease and cirrhosis | 571 | ICD9CM | 572.1 | Portal pyemia |
| Chronic liver disease and cirrhosis | 571 | ICD9CM | 572.8 | Other sequelae of chronic liver disease |
| Chronic liver disease and cirrhosis | 571 | ICD10CM | K72.90 | Hepatic failure, unspecified without coma |
| Chronic liver disease and cirrhosis | 571 | ICD10CM | K74.3 | Primary biliary cirrhosis |
| Chronic liver disease and cirrhosis | 571 | ICD10CM | K74.5 | Biliary cirrhosis, unspecified |
| Chronic liver disease and cirrhosis | 571 | ICD9CM | 571.6 | Biliary cirrhosis |
| Chronic liver disease and cirrhosis | 571 | ICD10CM | K74.4 | Secondary biliary cirrhosis |
| Chronic liver disease and cirrhosis | 571 | ICD10CM | K72.11 | Chronic hepatic failure with coma |
| Chronic liver disease and cirrhosis | 571 | ICD10CM | K70.4 | Alcoholic hepatic failure |
| Chronic liver disease and cirrhosis | 571 | ICD10CM | K76.6 | Portal hypertension |
| Chronic liver disease and cirrhosis | 571 | ICD10CM | K72.01 | Acute and subacute hepatic failure with coma |

| | | | | | |
|---|---|---|---|---|---|
| | Chronic liver disease and cirrhosis | 571 | ICD10CM | K75.0 | Abscess of liver |
| **Genitourinary system** | Chronic renal failure [CKD] | 585.3 | ICD10CM | N18.9 | Chronic kidney disease, unspecified |
| | Chronic renal failure [CKD] | 585.3 | ICD10CM | N18 | Chronic kidney disease (CKD) |
| | Chronic renal failure [CKD] | 585.3 | ICD9CM | 585.9 | Chronic kidney disease, unspecified |
| | Chronic renal failure [CKD] | 585.3 | ICD10CM | Z49.02 | Encounter for fitting and adjustment of peritoneal dialysis catheter |
| | Chronic renal failure [CKD] | 585.3 | ICD10CM | Z49.3 | Encounter for adequacy testing for dialysis |
| | Chronic renal failure [CKD] | 585.3 | ICD10CM | Z91.110 | Patient's noncompliance with dietary regimen due to financial hardship |
| | Chronic renal failure [CKD] | 585.3 | ICD10CM | Z91.141 | Patient's other noncompliance with medication regimen due to financial hardship |
| | Chronic renal failure [CKD] | 585.3 | ICD9CM | V45.11 | Renal dialysis status |
| | Chronic renal failure [CKD] | 585.3 | ICD10CM | Z99.2 | Dependence on renal dialysis |
| | Chronic renal failure [CKD] | 585.3 | ICD9CM | E879.1 | Kidney dialysis as the cause of abnormal reaction of patient, or of later complication, without mention of misadventure at time of procedure |
| | Chronic renal failure [CKD] | 585.3 | ICD10CM | Y84.1 | Kidney dialysis as the cause of abnormal reaction of the patient, or of later complication, without mention of misadventure at the time of the procedure |
| | Chronic renal failure [CKD] | 585.3 | ICD9CM | 585 | Chronic kidney disease (CKD) * |
| | Chronic renal failure [CKD] | 585.3 | ICD10CM | Z49.31 | Encounter for adequacy testing for hemodialysis |
| | Chronic renal failure [CKD] | 585.3 | ICD9CM | V56.31 | Encounter for adequacy testing for hemodialysis |
| | Chronic renal failure [CKD] | 585.3 | ICD9CM | V56.2 | Fitting and adjustment of peritoneal dialysis catheter |
| | Chronic renal failure [CKD] | 585.3 | ICD10CM | R88.0 | Cloudy (hemodialysis) (peritoneal) dialysis effluent |
| | Chronic renal failure [CKD] | 585.3 | ICD9CM | E874.2 | Mechanical failure of instrument or apparatus during kidney dialysis and other perfusion |
| | Chronic renal failure [CKD] | 585.3 | ICD9CM | 585 | Chronic kidney disease (CKD) |
| | Chronic renal failure [CKD] | 585.3 | ICD9CM | 792.5 | Cloudy (hemodialysis) (peritoneal) dialysis effluent |
| | Chronic renal failure [CKD] | 585.3 | ICD9CM | E872.2 | Failure of sterile precautions during kidney dialysis and other perfusion |
| | Chronic renal failure [CKD] | 585.3 | ICD9CM | V45.12 | Noncompliance with renal dialysis |
| | Chronic renal failure [CKD] | 585.3 | ICD9CM | E870.2 | Accidental cut, puncture, perforation or hemorrhage during kidney dialysis or other perfusion |

| | | | | |
|---|---|---|---|---|
| Chronic renal failure [CKD] | 585.3 | ICD9CM | V45.1 | Postsurgical renal dialysis status |
| Chronic renal failure [CKD] | 585.3 | ICD10CM | Z91.158 | Patient's noncompliance with renal dialysis for other reason |
| Chronic renal failure [CKD] | 585.3 | ICD10CM | Y62.2 | Failure of sterile precautions during kidney dialysis and other perfusion |
| Chronic renal failure [CKD] | 585.3 | ICD10CM | Z91.119 | Patient's noncompliance with dietary regimen due to unspecified reason |
| Chronic renal failure [CKD] | 585.3 | ICD10CM | Z49.32 | Encounter for adequacy testing for peritoneal dialysis |
| Chronic renal failure [CKD] | 585.3 | ICD9CM | 996.73 | Other complications due to renal dialysis device, implant, and graft |
| Chronic renal failure [CKD] | 585.3 | ICD9CM | V56.32 | Encounter for adequacy testing for peritoneal dialysis |
| Chronic renal failure [CKD] | 585.3 | ICD9CM | V56.1 | Fitting and adjustment of extracorporeal dialysis catheter |
| Chronic renal failure [CKD] | 585.3 | ICD10CM | Z91.151 | Patient's noncompliance with renal dialysis due to financial hardship |
| Chronic renal failure [CKD] | 585.3 | ICD9CM | V56 | Encounter for dialysis and dialysis catheter care |
| Chronic renal failure [CKD] | 585.3 | ICD10CM | Z49.01 | Encounter for fitting and adjustment of extracorporeal dialysis catheter |
| Chronic renal failure [CKD] | 585.3 | ICD9CM | 585.4 | Chronic kidney disease, Stage IV (severe) |
| Chronic renal failure [CKD] | 585.3 | ICD10CM | N18.4 | Chronic kidney disease, stage 4 (severe) |
| Chronic renal failure [CKD] | 585.3 | ICD10CM | Z49.0 | Preparatory care for renal dialysis |
| Chronic renal failure [CKD] | 585.3 | ICD10CM | Z49 | Encounter for care involving renal dialysis |
| Chronic renal failure [CKD] | 585.3 | ICD10CM | Z91.198 | Patient's noncompliance with other medical treatment and regimen for other reason |
| Chronic renal failure [CKD] | 585.3 | ICD9CM | V56.0 | Encounter for extracorporeal dialysis |
| Chronic renal failure [CKD] | 585.3 | ICD10CM | Z91.148 | Patient's other noncompliance with medication regimen for other reason |
| Chronic renal failure [CKD] | 585.3 | ICD9CM | 585.5 | Chronic kidney disease, Stage V |
| Chronic renal failure [CKD] | 585.3 | ICD9CM | V56.8 | Encounter for other dialysis |
| Chronic renal failure [CKD] | 585.3 | ICD9CM | E871.2 | Foreign object left in body during kidney dialysis or other perfusion |
| Chronic renal failure [CKD] | 585.3 | ICD10CM | Z91.199 | Patient's noncompliance with other medical treatment and regimen due to unspecified reason |
| Chronic renal failure [CKD] | 585.3 | ICD10CM | Z91.190 | Patient's noncompliance with other medical treatment and regimen due to financial hardship |
| Chronic renal failure [CKD] | 585.3 | ICD10CM | N18.5 | Chronic kidney disease, stage 5 |
| Chronic renal failure [CKD] | 585.3 | ICD9CM | 585.6 | End stage renal disease |

| | | | | | |
|---|---|---|---|---|---|
| | Chronic renal failure [CKD] | 585.3 | ICD10CM | Z91.118 | Patient's noncompliance with dietary regimen for other reason |
| | Chronic renal failure [CKD] | 585.3 | ICD10CM | N18.6 | End stage renal disease |
| | Chronic renal failure [CKD] | 585.3 | ICD9CM | 585.3 | Chronic kidney disease, Stage III (moderate) |
| | Chronic renal failure [CKD] | 585.3 | ICD10CM | Y62.0 | Failure of sterile precautions during surgical operation |
| | Chronic renal failure [CKD] | 585.3 | ICD10CM | N18.3 | Chronic kidney disease, stage 3 (moderate) |
| **Musculoskeletal** | Osteoarthrosis | 740 | ICD9CM | V13.4 | Personal history of arthritis |
| | Osteoarthrosis | 740 | ICD9CM | 715.08 | Osteoarthrosis, generalized * |
| | Osteoarthrosis | 740 | ICD9CM | 715.05 | Osteoarthrosis, generalized * |
| | Osteoarthrosis | 740 | ICD9CM | 715.99 | Osteoarthrosis, unspecified whether generalized or localized * |
| | Osteoarthrosis | 740 | ICD9CM | 715 | Osteoarthrosis and allied disorders |
| | Osteoarthrosis | 740 | ICD9CM | 715.06 | Osteoarthrosis, generalized * |
| | Osteoarthrosis | 740 | ICD9CM | 715.36 | Osteoarthrosis, localized, not specified whether primary or secondary, lower leg |
| | Osteoarthrosis | 740 | ICD10CM | M17.9 | Osteoarthritis of knee, unspecified |
| | Osteoarthrosis | 740 | ICD9CM | 715.34 | Osteoarthrosis, localized, not specified whether primary or secondary, hand |
| | Osteoarthrosis | 740 | ICD10CM | M18.9 | Osteoarthritis of first carpometacarpal joint, unspecified |
| | Osteoarthrosis | 740 | ICD9CM | 715.35 | Osteoarthrosis, localized, not specified whether primary or secondary, pelvic region and thigh |
| | Osteoarthrosis | 740 | ICD9CM | 715.38 | Osteoarthrosis, localized, not specified whether primary or secondary, other specified sites |
| | Osteoarthrosis | 740 | ICD9CM | 715.32 | Osteoarthrosis, localized, not specified whether primary or secondary, upper arm |
| | Osteoarthrosis | 740 | ICD9CM | 715.33 | Osteoarthrosis, localized, not specified whether primary or secondary, forearm |
| | Osteoarthrosis | 740 | ICD9CM | 715.3 | Osteoarthrosis, localized, not specified whether primary or secondary, site unspecified |
| | Osteoarthrosis | 740 | ICD9CM | 715.31 | Osteoarthrosis, localized, not specified whether primary or secondary, shoulder region |
| | Osteoarthrosis | 740 | ICD9CM | 715.37 | Osteoarthrosis, localized, not specified whether primary or secondary, ankle and foot |
| | Osteoarthrosis | 740 | ICD10CM | M16.9 | Osteoarthritis of hip, unspecified |
| | Osteoarthrosis | 740 | ICD9CM | 715.3 | Osteoarthrosis, localized, not specified whether primary or secondary |
| | Osteoarthrosis | 740 | ICD9CM | 715.9 | Osteoarthrosis, unspecified whether generalized or localized |
| | Osteoarthrosis | 740 | ICD9CM | 715.93 | Osteoarthrosis, unspecified whether generalized or localized, forearm |
| | Osteoarthrosis | 740 | ICD9CM | 715.9 | Osteoarthrosis, unspecified whether generalized or localized, site unspecified |
| | Osteoarthrosis | 740 | ICD10CM | M19.90 | Unspecified osteoarthritis, unspecified site |

| | | | | |
|---|---|---|---|---|
| Osteoarthrosis | 740 | ICD10CM | M17 | Osteoarthritis of knee |
| Osteoarthrosis | 740 | ICD9CM | 715.92 | Osteoarthrosis, unspecified whether generalized or localized, upper arm |
| Osteoarthrosis | 740 | ICD10CM | M15.9 | Polyosteoarthritis, unspecified |
| Osteoarthrosis | 740 | ICD9CM | 715.97 | Osteoarthrosis, unspecified whether generalized or localized, ankle and foot |
| Osteoarthrosis | 740 | ICD9CM | 715.98 | Osteoarthrosis, unspecified whether generalized or localized, other specified sites |
| Osteoarthrosis | 740 | ICD9CM | 715.96 | Osteoarthrosis, unspecified whether generalized or localized, lower leg |
| Osteoarthrosis | 740 | ICD10CM | M18.31 | Unilateral post-traumatic osteoarthritis of first carpometacarpal joint, right hand |
| Osteoarthrosis | 740 | ICD10CM | M19.91 | Primary osteoarthritis, unspecified site |
| Osteoarthrosis | 740 | ICD10CM | M19.079 | Primary osteoarthritis, unspecified ankle and foot |
| Osteoarthrosis | 740 | ICD10CM | M19.02 | Primary osteoarthritis, elbow |
| Osteoarthrosis | 740 | ICD10CM | M18.0 | Bilateral primary osteoarthritis of first carpometacarpal joints |
| Osteoarthrosis | 740 | ICD9CM | 715.13 | Osteoarthrosis, localized, primary, forearm |
| Osteoarthrosis | 740 | ICD10CM | M17.10 | Unilateral primary osteoarthritis, unspecified knee |
| Osteoarthrosis | 740 | ICD10CM | M16.11 | Unilateral primary osteoarthritis, right hip |
| Osteoarthrosis | 740 | ICD10CM | M19.071 | Primary osteoarthritis, right ankle and foot |
| Osteoarthrosis | 740 | ICD10CM | M18.11 | Unilateral primary osteoarthritis of first carpometacarpal joint, right hand |
| Osteoarthrosis | 740 | ICD10CM | M19.021 | Primary osteoarthritis, right elbow |
| Osteoarthrosis | 740 | ICD10CM | M19.032 | Primary osteoarthritis, left wrist |
| Osteoarthrosis | 740 | ICD10CM | M19.049 | Primary osteoarthritis, unspecified hand |
| Osteoarthrosis | 740 | ICD10CM | M19.031 | Primary osteoarthritis, right wrist |
| Osteoarthrosis | 740 | ICD10CM | M17.1 | Unilateral primary osteoarthritis of knee |
| Osteoarthrosis | 740 | ICD9CM | 715.12 | Osteoarthrosis, localized, primary, upper arm |
| Osteoarthrosis | 740 | ICD9CM | 715.16 | Osteoarthrosis, localized, primary, lower leg |
| Osteoarthrosis | 740 | ICD10CM | M19.042 | Primary osteoarthritis, left hand |
| Osteoarthrosis | 740 | ICD10CM | M19.011 | Primary osteoarthritis, right shoulder |
| Osteoarthrosis | 740 | ICD10CM | M18.10 | Unilateral primary osteoarthritis of first carpometacarpal joint, unspecified hand |
| Osteoarthrosis | 740 | ICD10CM | M19.0 | Primary osteoarthritis of other joints |
| Osteoarthrosis | 740 | ICD10CM | M19.012 | Primary osteoarthritis, left shoulder |
| Osteoarthrosis | 740 | ICD10CM | M18.12 | Unilateral primary osteoarthritis of first carpometacarpal joint, left hand |
| Osteoarthrosis | 740 | ICD10CM | M19.019 | Primary osteoarthritis, unspecified shoulder |
| Osteoarthrosis | 740 | ICD9CM | 715.15 | Osteoarthrosis, localized, primary, pelvic region and thigh |
| Osteoarthrosis | 740 | ICD10CM | M19.039 | Primary osteoarthritis, unspecified wrist |

| | | | | |
|---|---|---|---|---|
| Osteoarthrosis | 740 | ICD9CM | 715.18 | Osteoarthrosis, localized, primary, other specified sites |
| Osteoarthrosis | 740 | ICD10CM | M18.1 | Unilateral primary osteoarthritis of first carpometacarpal joint |
| Osteoarthrosis | 740 | ICD10CM | M19.022 | Primary osteoarthritis, left elbow |
| Osteoarthrosis | 740 | ICD10CM | M19.07 | Primary osteoarthritis ankle and foot |
| Osteoarthrosis | 740 | ICD10CM | M16.10 | Unilateral primary osteoarthritis, unspecified hip |
| Osteoarthrosis | 740 | ICD10CM | M19.072 | Primary osteoarthritis, left ankle and foot |
| Osteoarthrosis | 740 | ICD10CM | M16.1 | Unilateral primary osteoarthritis of hip |
| Osteoarthrosis | 740 | ICD10CM | M16.12 | Unilateral primary osteoarthritis, left hip |
| Osteoarthrosis | 740 | ICD9CM | 715.17 | Osteoarthrosis, localized, primary, ankle and foot |
| Osteoarthrosis | 740 | ICD10CM | M19.03 | Primary osteoarthritis, wrist |
| Osteoarthrosis | 740 | ICD10CM | M17.11 | Unilateral primary osteoarthritis, right knee |
| Osteoarthrosis | 740 | ICD10CM | M16.0 | Bilateral primary osteoarthritis of hip |
| Osteoarthrosis | 740 | ICD10CM | M19.04 | Primary osteoarthritis, hand |
| Osteoarthrosis | 740 | ICD9CM | 715.1 | Osteoarthrosis, localized, primary, site unspecified |
| Osteoarthrosis | 740 | ICD10CM | M17.12 | Unilateral primary osteoarthritis, left knee |
| Osteoarthrosis | 740 | ICD10CM | M19.029 | Primary osteoarthritis, unspecified elbow |
| Osteoarthrosis | 740 | ICD10CM | M19.09 | Primary osteoarthritis, other specified site |
| Osteoarthrosis | 740 | ICD9CM | 715.11 | Osteoarthrosis, localized, primary, shoulder region |
| Osteoarthrosis | 740 | ICD10CM | M19.01 | Primary osteoarthritis, shoulder |
| Osteoarthrosis | 740 | ICD10CM | M17.0 | Bilateral primary osteoarthritis of knee |
| Osteoarthrosis | 740 | ICD9CM | 715.14 | Osteoarthrosis, localized, primary, hand |
| Osteoarthrosis | 740 | ICD10CM | M19.041 | Primary osteoarthritis, right hand |
| Osteoarthrosis | 740 | ICD9CM | 715.1 | Osteoarthrosis, localized, primary |
| Osteoarthrosis | 740 | ICD9CM | 715.24 | Osteoarthrosis, localized, secondary, hand |
| Osteoarthrosis | 740 | ICD10CM | M19.19 | Post-traumatic osteoarthritis, other specified site |
| Osteoarthrosis | 740 | ICD10CM | M16.3 | Unilateral osteoarthritis resulting from hip dysplasia |
| Osteoarthrosis | 740 | ICD10CM | M19.29 | Secondary osteoarthritis, other specified site |
| Osteoarthrosis | 740 | ICD10CM | M19.149 | Post-traumatic osteoarthritis, unspecified hand |
| Osteoarthrosis | 740 | ICD10CM | M19.211 | Secondary osteoarthritis, right shoulder |
| Osteoarthrosis | 740 | ICD9CM | 715.26 | Osteoarthrosis, localized, secondary, lower leg |
| Osteoarthrosis | 740 | ICD9CM | 715.25 | Osteoarthrosis, localized, secondary, pelvic region and thigh |
| Osteoarthrosis | 740 | ICD10CM | M19.21 | Secondary osteoarthritis, shoulder |
| Osteoarthrosis | 740 | ICD10CM | M18.2 | Bilateral post-traumatic osteoarthritis of first carpometacarpal joints |
| Osteoarthrosis | 740 | ICD10CM | M19.142 | Post-traumatic osteoarthritis, left hand |
| Osteoarthrosis | 740 | ICD10CM | M19.242 | Secondary osteoarthritis, left hand |

| | | | | |
|---|---|---|---|---|
| Osteoarthrosis | 740 | ICD10CM | M17.31 | Unilateral post-traumatic osteoarthritis, right knee |
| Osteoarthrosis | 740 | ICD10CM | M19.231 | Secondary osteoarthritis, right wrist |
| Osteoarthrosis | 740 | ICD10CM | M15.2 | Bouchard's nodes (with arthropathy) |
| Osteoarthrosis | 740 | ICD9CM | 715 | Osteoarthrosis, generalized |
| Osteoarthrosis | 740 | ICD9CM | 715 | Osteoarthrosis, generalized, site unspecified |
| Osteoarthrosis | 740 | ICD10CM | M15.0 | Primary generalized (osteo)arthritis |
| Osteoarthrosis | 740 | ICD9CM | 715.09 | Osteoarthrosis, generalized, multiple sites |
| Osteoarthrosis | 740 | ICD10CM | M15.1 | Heberden's nodes (with arthropathy) |
| Osteoarthrosis | 740 | ICD9CM | 715.04 | Osteoarthrosis, generalized, hand |
| Osteoarthrosis | 740 | ICD10CM | M19.12 | Post-traumatic osteoarthritis, elbow |
| Osteoarthrosis | 740 | ICD10CM | M19.179 | Post-traumatic osteoarthritis, unspecified ankle and foot |
| Osteoarthrosis | 740 | ICD10CM | M19.229 | Secondary osteoarthritis, unspecified elbow |
| Osteoarthrosis | 740 | ICD10CM | M17.5 | Other unilateral secondary osteoarthritis of knee |
| Osteoarthrosis | 740 | ICD10CM | M19.22 | Secondary osteoarthritis, elbow |
| Osteoarthrosis | 740 | ICD10CM | M17.4 | Other bilateral secondary osteoarthritis of knee |
| Osteoarthrosis | 740 | ICD10CM | M17.32 | Unilateral post-traumatic osteoarthritis, left knee |
| Osteoarthrosis | 740 | ICD10CM | M16.4 | Bilateral post-traumatic osteoarthritis of hip |
| Osteoarthrosis | 740 | ICD10CM | M18.4 | Other bilateral secondary osteoarthritis of first carpometacarpal joints |
| Osteoarthrosis | 740 | ICD9CM | 715.89 | Osteoarthrosis involving, or with mention of more than one site, but not specified as generalized, multiple sites |
| Osteoarthrosis | 740 | ICD10CM | M15.3 | Secondary multiple arthritis |
| Osteoarthrosis | 740 | ICD10CM | M15.8 | Other polyosteoarthritis |
| Osteoarthrosis | 740 | ICD9CM | 715.8 | Osteoarthrosis involving, or with mention of more than one site, but not specified as generalized, site unspecified |
| Osteoarthrosis | 740 | ICD9CM | 715.8 | Osteoarthrosis involving or with mention of more than one site, but not specified as generalized |
| Osteoarthrosis | 740 | ICD10CM | M15.4 | Erosive (osteo)arthritis |
| Osteoarthrosis | 740 | ICD9CM | 715.2 | Osteoarthrosis, localized, secondary |
| Osteoarthrosis | 740 | ICD9CM | 715.27 | Osteoarthrosis, localized, secondary, ankle and foot |
| Osteoarthrosis | 740 | ICD10CM | M19.139 | Post-traumatic osteoarthritis, unspecified wrist |
| Osteoarthrosis | 740 | ICD10CM | M16.52 | Unilateral post-traumatic osteoarthritis, left hip |
| Osteoarthrosis | 740 | ICD10CM | M18.3 | Unilateral post-traumatic osteoarthritis of first carpometacarpal joint |
| Osteoarthrosis | 740 | ICD10CM | M16.32 | Unilateral osteoarthritis resulting from hip dysplasia, left hip |
| Osteoarthrosis | 740 | ICD10CM | M19.239 | Secondary osteoarthritis, unspecified wrist |
| Osteoarthrosis | 740 | ICD10CM | M19.9 | Osteoarthritis, unspecified site |

| | | | | |
|---|---|---|---|---|
| Osteoarthrosis | 740 | ICD9CM | 715.94 | Osteoarthrosis, unspecified whether generalized or localized, hand |
| Osteoarthrosis | 740 | ICD9CM | 715.95 | Osteoarthrosis, unspecified whether generalized or localized, pelvic region and thigh |
| Osteoarthrosis | 740 | ICD9CM | 715.91 | Osteoarthrosis, unspecified whether generalized or localized, shoulder region |
| Osteoarthrosis | 740 | ICD10CM | M19.132 | Post-traumatic osteoarthritis, left wrist |
| Osteoarthrosis | 740 | ICD9CM | 715.2 | Osteoarthrosis, localized, secondary, site unspecified |
| Osteoarthrosis | 740 | ICD10CM | M18.5 | Other unilateral secondary osteoarthritis of first carpometacarpal joint |
| Osteoarthrosis | 740 | ICD10CM | M19.93 | Secondary osteoarthritis, unspecified site |
| Osteoarthrosis | 740 | ICD10CM | M18.30 | Unilateral post-traumatic osteoarthritis of first carpometacarpal joint, unspecified hand |
| Osteoarthrosis | 740 | ICD10CM | M16.6 | Other bilateral secondary osteoarthritis of hip |
| Osteoarthrosis | 740 | ICD10CM | M19.272 | Secondary osteoarthritis, left ankle and foot |
| Osteoarthrosis | 740 | ICD10CM | M19.119 | Post-traumatic osteoarthritis, unspecified shoulder |
| Osteoarthrosis | 740 | ICD9CM | 715.21 | Osteoarthrosis, localized, secondary, shoulder region |
| Osteoarthrosis | 740 | ICD10CM | M19.23 | Secondary osteoarthritis, wrist |
| Osteoarthrosis | 740 | ICD10CM | M19.219 | Secondary osteoarthritis, unspecified shoulder |
| Osteoarthrosis | 740 | ICD10CM | M19.17 | Post-traumatic osteoarthritis, ankle and foot |
| Osteoarthrosis | 740 | ICD10CM | M19.24 | Secondary osteoarthritis, hand |
| Osteoarthrosis | 740 | ICD10CM | M18.50 | Other unilateral secondary osteoarthritis of first carpometacarpal joint, unspecified hand |
| Osteoarthrosis | 740 | ICD10CM | M17.30 | Unilateral post-traumatic osteoarthritis, unspecified knee |
| Osteoarthrosis | 740 | ICD10CM | M19.112 | Post-traumatic osteoarthritis, left shoulder |
| Osteoarthrosis | 740 | ICD10CM | M16.2 | Bilateral osteoarthritis resulting from hip dysplasia |
| Osteoarthrosis | 740 | ICD10CM | M19.141 | Post-traumatic osteoarthritis, right hand |
| Osteoarthrosis | 740 | ICD9CM | 715.22 | Osteoarthrosis, localized, secondary, upper arm |
| Osteoarthrosis | 740 | ICD10CM | M19.129 | Post-traumatic osteoarthritis, unspecified elbow |
| Osteoarthrosis | 740 | ICD10CM | M19.232 | Secondary osteoarthritis, left wrist |
| Osteoarthrosis | 740 | ICD10CM | M19.14 | Post-traumatic osteoarthritis, hand |
| Osteoarthrosis | 740 | ICD10CM | M16.50 | Unilateral post-traumatic osteoarthritis, unspecified hip |
| Osteoarthrosis | 740 | ICD10CM | M19.221 | Secondary osteoarthritis, right elbow |
| Osteoarthrosis | 740 | ICD10CM | M19.131 | Post-traumatic osteoarthritis, right wrist |
| Osteoarthrosis | 740 | ICD10CM | M19.13 | Post-traumatic osteoarthritis, wrist |
| Osteoarthrosis | 740 | ICD10CM | M18.32 | Unilateral post-traumatic osteoarthritis of first carpometacarpal joint, left hand |
| Osteoarthrosis | 740 | ICD9CM | 715.23 | Osteoarthrosis, localized, secondary, forearm |

| | | | | |
|---|---|---|---|---|
| Osteoarthrosis | 740 | ICD10CM | M16.51 | Unilateral post-traumatic osteoarthritis, right hip |
| Osteoarthrosis | 740 | ICD10CM | M16.7 | Other unilateral secondary osteoarthritis of hip |
| Osteoarthrosis | 740 | ICD10CM | M19.222 | Secondary osteoarthritis, left elbow |
| Osteoarthrosis | 740 | ICD10CM | M16.5 | Unilateral post-traumatic osteoarthritis of hip |
| Osteoarthrosis | 740 | ICD10CM | M18.51 | Other unilateral secondary osteoarthritis of first carpometacarpal joint, right hand |
| Osteoarthrosis | 740 | ICD10CM | M19.11 | Post-traumatic osteoarthritis, shoulder |
| Osteoarthrosis | 740 | ICD10CM | M19.279 | Secondary osteoarthritis, unspecified ankle and foot |
| Osteoarthrosis | 740 | ICD10CM | M19.1 | Post-traumatic osteoarthritis of other joints |
| Osteoarthrosis | 740 | ICD10CM | M19.249 | Secondary osteoarthritis, unspecified hand |
| Osteoarthrosis | 740 | ICD10CM | M19.171 | Post-traumatic osteoarthritis, right ankle and foot |
| Osteoarthrosis | 740 | ICD10CM | M17.3 | Unilateral post-traumatic osteoarthritis of knee |
| Osteoarthrosis | 740 | ICD9CM | 715.28 | Osteoarthrosis, localized, secondary, other specified sites |
| Osteoarthrosis | 740 | ICD10CM | M19.241 | Secondary osteoarthritis, right hand |
| Osteoarthrosis | 740 | ICD10CM | M19.271 | Secondary osteoarthritis, right ankle and foot |
| Osteoarthrosis | 740 | ICD10CM | M19.27 | Secondary osteoarthritis, ankle and foot |
| Osteoarthrosis | 740 | ICD10CM | M16.31 | Unilateral osteoarthritis resulting from hip dysplasia, right hip |
| Osteoarthrosis | 740 | ICD10CM | M19.92 | Post-traumatic osteoarthritis, unspecified site |
| Osteoarthrosis | 740 | ICD10CM | M19.121 | Post-traumatic osteoarthritis, right elbow |
| Osteoarthrosis | 740 | ICD10CM | M17.2 | Bilateral post-traumatic osteoarthritis of knee |
| Osteoarthrosis | 740 | ICD10CM | M19.122 | Post-traumatic osteoarthritis, left elbow |
| Osteoarthrosis | 740 | ICD10CM | M19.212 | Secondary osteoarthritis, left shoulder |
| Osteoarthrosis | 740 | ICD10CM | M19.172 | Post-traumatic osteoarthritis, left ankle and foot |
| Osteoarthrosis | 740 | ICD10CM | M16.30 | Unilateral osteoarthritis resulting from hip dysplasia, unspecified hip |
| Osteoarthrosis | 740 | ICD10CM | M18.52 | Other unilateral secondary osteoarthritis of first carpometacarpal joint, left hand |
| Osteoarthrosis | 740 | ICD10CM | M19.2 | Secondary osteoarthritis of other joints |
| Osteoarthrosis | 740 | ICD10CM | M19.111 | Post-traumatic osteoarthritis, right shoulder |
| Back pain | 760 | ICD9CM | 724.5 | Backache, unspecified |
| Back pain | 760 | ICD9CM | 724.2 | Lumbago |
| Back pain | 760 | ICD10CM | M54.89 | Other dorsalgia |
| Back pain | 760 | ICD9CM | 724.1 | Pain in thoracic spine |
| Back pain | 760 | ICD10CM | M54.51 | Vertebrogenic low back pain |
| Back pain | 760 | ICD10CM | M54 | Dorsalgia |
| Back pain | 760 | ICD9CM | 724.2 | Lumbago * |
| Back pain | 760 | ICD10CM | M54.5 | Low back pain |
| Back pain | 760 | ICD10CM | M54.50 | Low back pain, unspecified |

| | | | | |
|---|---|---|---|---|
| Back pain | 760 | ICD10CM | M54.9 | Dorsalgia, unspecified |
| Back pain | 760 | ICD10CM | M54.59 | Other low back pain |
| Back pain | 760 | ICD10CM | M54.6 | Pain in thoracic spine |

#### Table S2. GWAS summary statistics used for calculating PRS

| Disease category | Trait | Abbr. | Phe Code | phenotyping | Case | Control | n | Ancestry | Genome build | Author | Year | Source | Used for PAF/DALY analysis |
|---|---|---|---|---|---|---|---|---|---|---|---|---|---|
| **Neoplasms** | Cancer of esophagus | EC | 150 | Esophageal cancer (PheCode 150) | 1,388 | 159,201 | 160,589 | EAS | GRCh37 | Sakaue | 2021 | BBJ1 | No |
| | | EC | | Esophageal cancer (PheCode 150) | 2,386 | 634,509 | 636,895 | EUR | GRCh37 | Sakaue | 2021 | GCST90018841 | No |
| | | EC | | Phecode 150 : Cancer of esophagus | 1,460 | 286,102 | 287,562 | EAS | GRCh38 | Yang | 2025 | TPMI | Yes |
| | Cancer of stomach | SC | 151 | ICD10 C16.9: Malignant neoplasm of stomach, unspecified | 145 | 456,203 | 456,348 | EUR | GRCh37 | Jiang | 2021 | GCST90043847 | No |
| | | SC | | Gastric caner (PheCode 151) | 8,950 | 634,288 | 643,238 | MULTI | GRCh37 | Sakaue | 2021 | GCST90018849 | Yes |
| | | SC | | Phecode 151: Cancer of stomach | 2,618 | 284,931 | 287,549 | EAS | GRCh38 | Yang | 2025 | TPMI | No |
| | Colorectal cancer | CRC | 153 | ICD10 C20: Malignant neoplasm of rectum | 1,473 | 456,348 | 457,821 | EUR | GRCh37 | Jiang | 2021 | GCST90043857 | No |
| | | CRC | | PheCode 153: colorectal cancer | 8,545 | 231,444 | 239,989 | EAS | GRCh37 | Nam | 2022 | KoGES+BBJ | No |
| | | CRC | | Phecode 153: Colorectal cancer | 7,733 | 302,768 | 310,501 | EAS | GRCh38 | Yang | 2025 | TPMI | Yes |
| | Cancer of bronchus; lung | LC | 165 | PheCode 165.1: Cancer of bronchus; lung | 2,120 | 454,228 | 456,348 | EUR | GRCh37 | Jiang | 2021 | GCST90041821 | No |
| | | LC | | Self-reported: lung cancer + PheCode 165.1: Cancer of bronchus; lung | 29,266 | 56,450 | 85,716 | EUR | GRCh37 | McKay | 2017 | GCST004748 | No |
| | | LC | | PheCode 165.1: Cancer of bronchus; lung | 4,489 | 246,535 | 251,024 | EAS | GRCh37 | Nam | 2022 | KoGES+BBJ | No |
| | | LC | | PheCode 165.1: Cancer of bronchus; lung | 8,235 | 663,294 | 671,529 | MULTI | GRCh37 | Sakaue | 2021 | GCST90018875 | No |
| | | LC | | PheCode 165.1: Cancer of bronchus lung | 6,364 | 335,748 | 342,112 | EAS | GRCh38 | Yang | 2025 | TPMI | Yes |
| | Skin cancer | SKC | 172 | Skin cancer (PheCode 172.11/172.2) | 154 | 178,572 | 178,726 | EAS | GRCh37 | Sakaue | 2021 | BBJ1 | Yes |
| | | SKC | | Phecode 172: Skin cancer | 1,715 | 341,372 | 343,087 | EAS | GRCh38 | Yang | 2025 | TPMI | No |

| | | | | | | | | | | | | |
|---|---|---|---|---|---|---|---|---|---|---|---|---|
| Melanomas of skin | SKC | | Melanomas of skin (PheCode 172.11) | 2,824 | 453,524 | 456,348 | EUR | GRCh37 | Jiang | 2021 | GCST90041829 | Yes |
| Breast cancer | BRC | 174 | Breast Cancer Association Consortium (BCAC) and Discovery, Biology and Risk of Inherited Variants in Breast Cancer Consortium (DRIVE) | 76,192 | 63,082 | 139,274 | EUR | GRCh37 | Michailidou | 2017 | GCST004988 | Yes |
| | BRC | | Malignant neoplasm of breast, excluding all Neoplasms (controls excluding all Neoplasms) | 24,270 | 222,078 | 246,348 | EUR | GRCh38 | Kurki | 2024 | Finngen | No |
| | BRC | | Breast cancer (PheCode 174) | 6,749 | 119,131 | 125,880 | EAS | GRCh37 | Nam | 2022 | KoGES+BBJ | No |
| | BRC | | Phecode 174 Breast cancer | 12,566 | 313,661 | 326,227 | EAS | GRCh38 | Yang | 2025 | TPMI | No |
| | BRC1 | | 174.1 Breast cancer female | 11,413 | 162,431 | 173,844 | EAS | GRCh38 | Yang | 2025 | TPMI | No |
| Cervical cancer | CC | 180 | ICD-O-3 codes C530-C539 (excluding 9050-9055, 9140, 9590-9993) | 6,563 | 410,350 | 416,913 | EUR | GRCh37 | Rashkin | 2020 | GCST90011816 | No |
| | CC | | Cervical cancer (PheCode 180.1) | 1,659 | 381,902 | 383,561 | EUR | GRCh37 | Zhou | 2018 | GCST90435606 | No |
| | CC | | Cervical cancer (Phecode 180.1) | 1,876 | 298,863 | 300,739 | MULTI | GRCh37 | Sakaue | 2021 | GCST90018817 | No |
| | CC1 | | Phecode 180.1: Cervical cancer | 1,423 | 158,528 | 159,951 | EAS | GRCh38 | Yang | 2025 | TPMI | No |
| | CC | | 180 Cervical cancer and dysplasia | 4,366 | 158,528 | 162,894 | EAS | GRCh38 | Yang | 2025 | TPMI | Yes |
| Malignant neoplasm of ovary | OC | 184 | Ovarian cancer (PheCode 184.11) | 843 | 60,614 | 61,457 | EAS | GRCh37 | Sakaue | 2021 | GCST90018668 | Yes |
| | OC | | OCAC (Ovarian Cancer Association Consortium) + BCAC (Breast Cancer Association Consortium) | 23,394 | 105,724 | 129,118 | EUR | GRCh38 | Eileen | 2024 | GCST90244167 | No |
| | OC | | 184.1 Malignant neoplasm of ovary and other uterine adnexa | 1,847 | 166,354 | 168,201 | EAS | GRCh38 | Yang | 2025 | TPMI | No |
| Cancer of prostate | PC | 185 | OncoArray project | 79,148 | 61,106 | 140,254 | EUR | GRCh37 | Schumacher | 2018 | GCST006085 | Yes |
| | PC | | Malignant neoplasm of prostate, excluding all Neoplasms | 20,368 | 156,671 | 177,039 | EUR | GRCh38 | Kurki | 2024 | Finngen | No |

| | | | | | | | | | | | | | |
|---|---|---|---|---|---|---|---|---|---|---|---|---|---|
| | | | | (controls excluding all Neoplasms) | | | | | | | | | |
| | | PC | | Phecode 185: Cancer of prostate | 5,385 | 112,559 | 117,944 | EAS | GRCh38 | Yang | 2025 | TPMI | No |
| **Metabolic** | Type 2 diabetes | DM2 | 250 | ICD-10 E11.9 · Type 2 diabetes mellitus without complications | 714 | 455,634 | 456,348 | EUR | GRCh37 | Jiang | 2021 | GCST90043638 | No |
| | | DM2 | | ICD-10 E11.9 · Type 2 diabetes mellitus without complications + self report | 428,452 | 2,107,149 | 2,535,601 | MULTI | GRCh37 | Suzuki | 2024 | Suzuki et al 2024 | Yes |
| | | DM2 | | Type 2 diabetes (T2D) (PheCode 250.2) | 50,466 | 199,159 | 249,625 | EAS | GRCh37 | Nam | 2022 | KoGES | No |
| | | DM2 | | Phecode 250.2: Type 2 diabetes | 59,289 | 269,125 | 328,414 | EAS | GRCh38 | Yang | 2025 | TPMI | No |
| | Hypercholesterolemia | HPC | 272 | Hypercholesterolemia (PheCode 272.11) | 33,242 | 373,034 | 406,276 | EUR | GRCh37 | Zhou | 2018 | GCST90435755 | No |
| | | HPC | | Phecode 272.11: Hypercholesterolemia | 14,083 | 242,117 | 256,200 | EAS | GRCh38 | Yang | 2025 | TPMI | Yes |
| | Gout | GT | 274 | Phecode: 274.1 | 24,278 | 1,027,375 | 1,051,653 | EUR | GRCh38 | Zhou | 2022 | GCST90399701 | No |
| | | GT | | Phecode: 274.1 | 10,425 | 338,934 | 349,359 | EAS | GRCh38 | Zhou | 2022 | GCST90399700 | Yes |
| | | GT | | Phecode 274.1: Gout | 24,411 | 315,107 | 339,518 | EAS | GRCh38 | Yang | 2025 | TPMI | No |
| | Obesity | OB | 278 | ICD-10 E66, E66.01, E66.1, E66.2, E66.8, E66.9 | 31,499 | 468,693 | 500,192 | EUR | GRCh38 | Kurki | 2024 | Finngen | Yes |
| | | OB | | Phecode 278.1: Obesity | 4,576 | 336,405 | 340,981 | EAS | GRCh38 | Yang | 2025 | TPMI | No |
| **Mental Disorders** | Alzheimer's disease | AD | 290 | ICD-10 G30, F00, ICD-9 3310, ICD-8 29010 | 71,880 | 383,378 | 455,258 | EUR | GRCh38 | Mars | 2022 | PGS002753 | No |
| | | AD | | ICD-10 | 85,934 | 401,577 | 487,511 | EUR | GRCh38 | Bellenguez | 2022 | GCST90027158 | No |
| | | AD | | Phecode 290.11: Alzheimers disease | 2,828 | 320,866 | 323,694 | EAS | GRCh38 | Yang | 2025 | TPMI | Yes |
| | Schizophrenia | SCH | 295 | Schizophrenia (PheCode 295.1) | 367 | 455,981 | 456,348 | EUR | GRCh37 | Jiang | 2021 | GCST90043704 | No |
| | | SCH | | ICD-10 F20. 9: Schizophrenia | 11,602 | 36,921 | 48,523 | EUR | GRCh37 | Pedersen | 2023 | GCST90275156 | No |
| | | SCH | | Schizophrenia (PheCode 295.1) | 22,778 | 35,362 | 58,140 | EAS | GRCh37 | Lam | 2019 | PGC | Yes |
| | Depression | DEP | 296 | depression (PheCode 296.2) | 836 | 177,794 | 178,630 | EAS | GRCh37 | Sakaue | 2021 | BBJ1 | No |

| | | | | | | | | | | | | | |
|---|---|---|---|---|---|---|---|---|---|---|---|---|---|
| | | DEP | | ICD10 F32 F33 ICD9 2961 2968 3004 | 59,333 | 434,831 | 494,164 | EUR | GRCh38 | Kurki | 2024 | Finngen | No |
| | | DEP | | ICD-10 F32. 9: Depression + structured clinical interviews + medical care records + symptom questionnaires + self-reported surveys | 88,316 | 902,757 | 991,073 | MULTI | GRCh37 | Meng | 2024 | PGC | Yes |
| | | DEP | | Phecode 296.2: Depression | 7,697 | 266,225 | 273,922 | EAS | GRCh38 | Yang | 2025 | TPMI | No |
| | Generalized anxiety disorder | ANX | 300 | ICD-10 F40.2: Specific (isolated) phobias | 1,809 | 500,672 | 502,481 | EUR | GRCh37 | Brasher | 2023 | GCST90225524 | No |
| | | ANX | | panic disorder | 2,248 | 7,992 | 10,240 | EUR | GRCh37 | Forstner | 2021 | PGC | No |
| | | GAD | | Phecode 300.11: Generalized anxiety disorder | 6,561 | 266,225 | 272,786 | EAS | GRCh38 | Yang | 2025 | TPMI | Yes |
| | Alcoholism | ALC | 317 | Alcoholism (PheCode 317.1) | 588 | 455,760 | 456,348 | EUR | GRCh37 | Jiang | 2021 | GCST90043727 | No |
| | | ALC | | ICD10 F10.2 ICD9 303 | 12,599 | 468,475 | 481,074 | EUR | GRCh38 | Kurki | 2024 | Finngen | No |
| | | ALC | | Phecode 317.1: Alcoholism | 1,368 | 337,684 | 339,052 | EAS | GRCh38 | Yang | 2025 | TPMI | Yes |
| **Neurological** | Migraine | MG | 340 | ICD-10 G43. 909: Migraine | 26,052 | 487,214 | 513,266 | EUR | GRCh37 | Meyers | 2023 | GCST90271641 | No |
| | | MG | | ICD10 G43 ICD9 346 | 26,894 | 374,605 | 401,499 | EUR | GRCh38 | Kurki | 2024 | Finngen | No |
| | | MG | | Phecode 340: Migraine | 9,828 | 313,133 | 322,961 | EAS | GRCh38 | Yang | 2025 | TPMI | Yes |
| | | TH | | Phecode 306.9: Tension headache | 2,659 | 266,225 | 268,884 | EAS | GRCh38 | Yang | 2025 | TPMI | No |
| | Epilepsy | EP | 345 | The International League Against Epilepsy terminology | 27,559 | 42,436 | 69,995 | EUR | GRCh37 | ILAE | 2023 | GCST90271611 | No |
| | | EP | | ICD10 G40 ICD9 345 | 15,645 | 374,605 | 390,250 | EUR | GRCh38 | Kurki | 2024 | Finngen | No |
| | | EP | | Epilepsy (PheCode 345.11) | 2,466 | 175,788 | 178,254 | EAS | GRCh37 | Sakaue | 2021 | BBJ1 | No |
| | | EP | | Phecode 345.1: Epilepsy | 2,067 | 313,497 | 315,564 | EAS | GRCh38 | Yang | 2025 | TPMI | Yes |
| | Genetic generalized epilepsy | GGE | 345 | The International League Against Epilepsy terminology | 6,952 | 42,436 | 49,388 | EUR | GRCh37 | ILAE | 2023 | Pubmed ID: 37653029 | Yes |
| | | GGE | | The International League Against Epilepsy terminology | 7,407 | 52,538 | 59,945 | MULTI | GRCh37 | ILAE | 2023 | Pubmed ID: 37653029 | no |

| | | | | | | | | | | | | | |
|---|---|---|---|---|---|---|---|---|---|---|---|---|---|
| **Sense Organs** | Hearing loss | HL | 389 | Hearing loss (PheCode 389) | 585 | 455,763 | 456,348 | EUR | GRCh37 | Jiang | 2021 | GCST90043836 | No |
| | | HL | | Hearing loss (PheCode 389) | 3,400 | 175,326 | 178,726 | EAS | GRCh37 | Sakaue | 2021 | GCST90018637 | No |
| | | HL | | Hearing loss (PheCode 389) | 287,197 | 285,215 | 572,412 | MULTI | GRCh37 | Verma | 2024 | GCST90480117 | No |
| | | HL | | Phecode 389: Hearing loss | 18,004 | 313,340 | 331,344 | EAS | GRCh38 | Yang | 2025 | TPMI | Yes |
| **Circulatory System** | Hypertension | HT | 401 | PheCode 401.1 hypertension | 14,834 | 57,395 | 72,229 | EAS | GRCh37 | Nam | 2022 | KoGES | No |
| | | HT | | Hypertension (PheCode 401) | 456,409 | 151,853 | 608,262 | MULTI | GRCh37 | Verma | 2024 | GCST90480128 | No |
| | | HT | | Phecode 401: Hypertension | 85,661 | 248,898 | 334,559 | EAS | GRCh38 | Yang | 2025 | TPMI | Yes |
| | | HT1 | | 401.1 Essential hypertension | 68,314 | 248,898 | 317,212 | EAS | GRCh38 | Yang | 2025 | TPMI | No |
| | Ischemic Heart Disease | CAD | 411 | ICD-10 I25.10: Atherosclerotic heart disease of native coronary artery without angina pectoris | 210,842 | 1,167,328 | 1,378,170 | MULTI | GRCh37 | Aragam | 2022 | GCST90132315 | No |
| | | CAD | | Ischemic heart disease (PheCode 411) | 31,355 | 377,103 | 408,458 | EUR | GRCh37 | Zhou | 2019 | GCST008370 | Yes |
| | | CAD | | Ischemic heart disease (PheCode 411) | 32,512 | 146,214 | 178,726 | EAS | GRCh37 | Kanai | 2021 | BBJ2 | No |
| | | CAD | | Phecode 411.4: Coronary atherosclerosis | 18,436 | 297,646 | 316,082 | EAS | GRCh38 | Yang | 2025 | TPMI | No |
| | Myocardial infarction | MI | 411 | Myocardial infarction (PheCode 411.2) | 61,000 | 577,000 | 638,000 | MULTI | GRCh37 | Hartiala | 2021 | Hartiala et al 2021 | Yes |
| | | MI | | Myocardial infarction (PheCode 411.2) | 14,992 | 146,214 | 161,206 | EAS | GRCh37 | Sakaue | 2021 | GCST90018657 | No |
| | | MI | | Myocardial infarction (PheCode 411.2) | 8,528 | 447,820 | 456,348 | EUR | GRCh37 | Jiang | 2021 | GCST90043954 | No |
| | | MI | | Myocardial infarction (PheCode 411.2) | 11,703 | 444,730 | 456,433 | EUR | GRCh37 | Zhou | 2018 | GCST90436073 | No |
| | | MI | | ICD-10 I21 I22, ICD-9 410, ICD-8 410 | 31,666 | 416,171 | 447,837 | EUR | GRCh38 | Kurki | 2024 | Finngen | No |
| | | MI | | Phecode 411.2: Myocardial infarction | 4,062 | 298,735 | 302,797 | EAS | GRCh38 | Yang | 2025 | TPMI | No |
| | Atrial fibrillation | AF | 427 | ICD-10: Unspecified atrial fibrillation | 77,690 | 1,167,040 | 1,244,730 | MULTI | GRCh37 | Miyazawa | 2023 | Miyazawa et al 2023 | Yes |

| | | | | | | | | | | | | |
|---|---|---|---|---|---|---|---|---|---|---|---|---|
| | AF | | ICD-10 code I48 and ICD-9 code 427.3/427.31 (HUNT, deCODE, MGI) | 60,620 | 970,216 | 1,030,836 | EUR | GRCh37 | Nielsen | 2018 | GCST006414 | No |
| | AF | | Phecode 427.21: Atrial fibrillation | 7,931 | 304,431 | 312,362 | EAS | GRCh38 | Yang | 2025 | TPMI | No |
| Heart failure NOS | HF | 428 | ICD-10: Heart failure | 115,150 | 1,550,331 | 1,665,481 | MULTI | GRCh37 | Levin | 2022 | Levin et al 2022 | Yes |
| | HF | | Phecode 428.2: Heart failure NOS | 8,079 | 312,910 | 320,989 | EAS | GRCh38 | Yang | 2025 | TPMI | No |
| Intracranial hemorrhage | ST | 430 | ICD10 I64 Stroke, not specified as haemorrhage or infarction. | 110,182 | 1,393,716 | 1,503,898 | EUR | GRCh37 | Mishra | 2022 | GCST90104534 | No |
| | ST | | ICD10 I64 Stroke, not specified as haemorrhage or infarction. | 23,345 | 245,585 | 268,930 | EAS | GRCh38 | Zhou | 2022 | GCST90399731 | Yes |
| | IH | | Phecode 430: Intracranial hemorrhage | 2,642 | 304,660 | 307,302 | EAS | GRCh38 | Yang | 2025 | TPMI | No |
| Subarachnoid hemorrhage | SAH | 430 | Subarachnoid hemorrhage (PheCode 430.1) | 5,140 | 71,952 | 77,092 | MULTI | GRCh37 | Bakker | 2020 | Bakker et al 2020 | No |
| | SAH | | Subarachnoid hemorrhage (PheCode 430.1) | 832 | 455,516 | 456,348 | EUR | GRCh37 | Jiang | 2021 | GCST90043993 | Yes |
| | SAH | | Subarachnoid hemorrhage (PheCode 430.1) | 1,203 | 152,022 | 153,225 | EAS | GRCh37 | Sakaue | 2021 | GCST90018703 | No |
| Cerebrovascular disease | CVD | 433 | Cerebrovascular disease (PheCode 433) | 475 | 455,873 | 456,348 | EUR | GRCh37 | Jiang | 2021 | GCST90044004 | No |
| | CVD | | Cerebrovascular disease (PheCode 433) | 93,615 | 503,513 | 597,128 | MULTI | GRCh37 | Verma | 2024 | GCST90480195 | Yes |
| | CVD | | Phecode 433: Cerebrovascular disease | 30,664 | 304,660 | 335,324 | EAS | GRCh38 | Yang | 2025 | TPMI | No |
| Cerebral aneurysm | CA | 434 | Cerebral aneurysm (PheCode 433.5) | 4,077 | 624,760 | 628,837 | MULTI | GRCh37 | Sakaue | 2021 | GCST90018815 | No |
| | CA | | Cerebral aneurysm (PheCode 433.5) | 290 | 456,058 | 456,348 | EUR | GRCh37 | Jiang | 2021 | GCST90044003 | No |
| | CA | | Cerebral aneurysm (PheCode 433.5) | 3,132 | 152,022 | 155,154 | EAS | GRCh37 | Sakaue | 2021 | GCST90018595 | Yes |
| Peripheral vascular disease | PAD | 443 | Peripheral vascular disease (PheCode 443) | 11,226 | 649,565 | 660,791 | MULTI | GRCh37 | Sakaue | 2021 | Sakaue et al 2022 | Yes |

| | | | | | | | | | | | | | |
|---|---|---|---|---|---|---|---|---|---|---|---|---|---|
| **Respiratory System** | Asthma | AS | 495 | Phecode: 495 | 121,940 | 1,254,131 | 1,376,071 | EUR | GRCh38 | Zhou | 2022 | GCST90399686 | No |
| | | AS | | Phecode: 495 | 18,549 | 322,655 | 341,204 | EAS | GRCh38 | Zhou | 2022 | GCST90399685 | Yes |
| | | AS | | Phecode 495: Asthma | 20,598 | 297,628 | 318,226 | EAS | GRCh38 | Yang | 2025 | TPMI | No |
| | Obstructive chronic bronchitis | COPD | 496 | Phecode: 496 | 58,559 | 937,358 | 995,917 | EUR | GRCh38 | Zhou | 2022 | GCST90399694 | No |
| | | COPD | | Phecode: 496 | 19,044 | 310,689 | 329,733 | EAS | GRCh38 | Zhou | 2022 | GCST90399693 | Yes |
| | | CAO | | Phecode 496: Chronic airway obstruction | 17,930 | 297,628 | 315,558 | EAS | GRCh38 | Yang | 2025 | TPMI | No |
| | | COPD | | Phecode 496.21 Obstructive chronic bronchitis (COPD) | 8,689 | 297,628 | 306,317 | EAS | GRCh38 | Yang | 2025 | TPMI | No |
| **Digestive system** | GERD | GERD | 530 | Gastroesophageal reflux disease (PheCode 530.11) | 33,905 | 611,812 | 645,717 | MULTI | GRCh37 | Sakaue | 2021 | GCST90018848 | Yes |
| | | GERD | | 530.11 GERD | 47,138 | 271,934 | 319,072 | EAS | GRCh38 | Yang | 2025 | TPMI | No |
| | Inflammatory bowel disease and other gastroenteritis and colitis | IBD | 555 | The European Evidence-based Consensus or the ECCO-ESGAR Guideline for Diagnostic Assessment in IBD | 2,599 | 552,343 | 554,942 | EUR | GRCh37 | Zheng | 2023 | GCST90204144 | No |
| | | IBD | | The European Evidence-based Consensus or the ECCO-ESGAR Guideline for Diagnostic Assessment in IBD | 14,393 | 15,456 | 29,849 | EAS | GRCh37 | Liu | 2023 | GCST90432182 | Yes |
| | Chronic liver disease and cirrhosis | LD | 571 | Alcoholic liver damage (PheCode 317.11) | 451 | 455,897 | 456,348 | EUR | GRCh37 | Jiang | 2021 | GCST90043726 | No |
| | | LD | | ICD-10: K76.9 · Liver disease | 8,021 | 16,489 | 24,510 | EUR | GRCh37 | Cordell | 2021 | GCST90061440 | No |
| | | LD | | ICD10 K70-K77, K77.0*B96.8 | 15,135 | 485,213 | 500,348 | EUR | GRCh38 | Kurki | 2024 | Finngen | No |
| | | LD | | Phecode 571: Chronic liver disease and cirrhosis | 34,960 | 291,169 | 326,129 | EAS | GRCh38 | Yang | 2025 | TPMI | Yes |
| **Genitourinary system** | Chronic renal failure [CKD] | CKD | 585 | Chronic renal failure [CKD] (PheCode 585.3) | 562 | 455,786 | 456,348 | EUR | GRCh37 | Jiang | 2021 | GCST90044223 | No |
| | | CKD | | Chronic renal failure [CKD] (PheCode 585.3) | 2,117 | 174,345 | 176,462 | EAS | GRCh37 | Sakaue | 2021 | GCST90018602 | No |

| | | | | | | | | | | | | | |
|---|---|---|---|---|---|---|---|---|---|---|---|---|---|
| | | CKD | | Chronic renal failure [CKD] (PheCode 585.3) | 10,404 | 648,916 | 659,320 | MULTI | GRCh37 | Sakaue | 2021 | GCST90018822 | No |
| | | CKD | | Phecode 585.3: Chronic renal failure CKD | 25,906 | 294,706 | 320,612 | EAS | GRCh38 | Yang | 2025 | TPMI | Yes |
| **Musculoskeletal** | Osteoarthrosis | OA | 740 | UKBB: Self-reported and hospital diganosed OA - all OA | 77,052 | 378,169 | 455,221 | EUR | GRCh37 | Tachmazidou | 2019 | GCST007093 | No |
| | | OA | | Osteoarthritis of the hip or knee (meta-analysis containing many definitions) | 213,839 | 1,080,482 | 1,294,321 | MULTI | GRCh37 | Hatzikotoulas | 2025 | GCST90566799 | Yes |
| | | OA | | Phecode 740: Osteoarthrosis | 39,910 | 291,025 | 330,935 | EAS | GRCh38 | Yang | 2025 | TPMI | No |
| | Back pain | BP | 760 | Back pain (PheCode 760) | 1,732 | 176,994 | 178,726 | EAS | GRCh37 | Sakaue | 2021 | BBJ1 | No |
| | | BP | | Pain type(s) experienced in last month: Back pain (UKB data field 6159_4) | 116,509 | 338,763 | 455,272 | EUR | GRCh37 | Jiang | 2021 | GCST90044406 | Yes |
| | | BP | | Phecode 760: Back pain | 17,965 | 314,381 | 332,346 | EAS | GRCh38 | Yang | 2025 | TPMI | No |

#### Table S3. Definition of MRFs

| MRF | Abbreviation | Definition from GBD |
|---|---|---|
| Air pollution | Air pollution | Outdoor airpollution, ozone and nitrogen dioxide pollution and household air pollution |
| Dietary risks | Dietary risks | 15 dietary risks including calcium, fiber, fruits, legumes, milk, nuts & seeds, seafood omega-3 acids, processed meat, polyunsaturated fatty acids, red meat, sodium, sugar-sweetedned beverages, trans fatty acids, vegetables, whole grains |
| Drug use | Drug use | Exposure to drug use |
| High alcohol use | High alcohol use | Grams per day of pure alcohol consumed exceeding TMREL among current drinkers |
| High body-mass index | High BMI | >20-25 kg/m2 |
| High fasting plasma glucose | High FPG | > 4.8-5.4 mmol/L |
| High LDL cholesterol | High LDL | > 0.7-1.3 mmol/L |
| High systolic blood pressure | High SBP | > 110-115 mmHg |
| Intimate partner violence | Intimate partner violence | Ever experienced one or more acts of physical and/or sexual violence by a current or former intimate partner since the age of 15 years. |
| Kidney dysfunction | Kidney dysfunction | None/Stage 1-2/Stage 3/Stage 4/Stage 5 CKD |
| Low physical activity | Low physical activity | < 3000-4500 METs-minutes/week |
| Non-optimal temperature | Non-optimal temperature | Same day exposure to ambient temperature that is either warmer or colder than the location-specific TMREL |
| Occupational risks | Occupational risks | Occupational asbestos, asthmagens, carcinogens, ergonomic factors, injuries, noise and particulates |
| Other environmental risks | Other environmental risks | Residential radon and lead exposure |
| Sexual violence against children and bullying | SVCB | Any unwanted contact sexual act experienced by individuals aged 15 years or younger with the perpetrator being five or more years older than the victim and bullying victimisation by peers |
| Tobacco | Tobacco | Current use of any tobacco or former smokers who have quit all tobacco products for at least six months |
| Unsafe sex | Unsafe sex | The absence of disease transmission due to sexual contact |

#### Table S4. Disease prevalence, age of cases and controls and age of onset

| | | | Cases | | | Controls | | | Age of onset | | All-Cohort Prevalence | | |
|---|---|---|---|---|---|---|---|---|---|---|---|---|---|
| Category | Diseases | Phecode | Number | Mean age | SD age | Number | Mean age | SD age | Mean age | SD age | All | Male | Female |
| Neoplasms | Cancer of esophagus | 150 | 176 | 64.33 | 7.17 | 113,578 | 55.37 | 11.96 | 47.72 | 11.35 | 0.12% | 0.09% | 0.14% |
| | Cancer of stomach | 151 | 367 | 65.63 | 8.77 | 118,220 | 55.47 | 11.98 | 58.56 | 10.11 | 0.26% | 0.28% | 0.24% |
| | Colorectal cancer | 153 | 2,196 | 63.57 | 9.81 | 111,878 | 55.05 | 11.94 | 52.88 | 11.98 | 1.54% | 1.75% | 1.42% |
| | Cancer of bronchus; lung | 165.1 | 1,963 | 64.36 | 9.23 | 134,080 | 55.99 | 11.90 | 58.05 | 10.79 | 1.38% | 1.41% | 1.36% |
| | Skin cancer | 172 | 491 | 63.47 | 10.99 | 141,478 | 56.21 | 11.91 | 55.35 | 13.72 | 0.34% | 0.38% | 0.33% |
| | Melanomas of skin | 172.11 | 102 | 59.91 | 12.32 | 141,478 | 56.21 | 11.91 | 49.97 | 15.48 | 0.07% | 0.06% | 0.08% |
| | Breast cancer | 174 | 6,803 | 59.11 | 10.20 | 103,905 | 55.48 | 12.31 | 48.15 | 11.02 | 4.77% | 0.10% | 7.44% |
| | Cervical cancer and dysplasia | 180 | 7,167 | 57.82 | 10.21 | 94,729 | 55.68 | 12.62 | 45.73 | 10.31 | 5.03% | 0.00% | 7.89% |
| | Cervical cancer | 180.1 | 321 | 60.17 | 10.41 | 94,729 | 55.68 | 12.62 | 47.35 | 11.43 | 0.23% | 0.00% | 0.35% |
| | Malignant neoplasm of ovary | 184.11 | 542 | 57.76 | 10.60 | 113,282 | 56.40 | 12.20 | 47.36 | 12.56 | 0.38% | 0.00% | 0.60% |
| | Cancer of prostate | 185 | 851 | 69.86 | 6.50 | 119,857 | 55.39 | 11.74 | 63.65 | 8.37 | 0.60% | 1.65% | 0.00% |
| Metabolic | Type 2 diabetes | 250.2 | 33,364 | 62.52 | 10.06 | 99,746 | 53.85 | 11.75 | 51.54 | 11.13 | 23.42% | 25.06% | 22.48% |
| | Hypercholesterolemia | 272.11 | 19,388 | 63.14 | 9.59 | 68,903 | 51.65 | 11.49 | 53.38 | 10.04 | 13.61% | 12.38% | 14.31% |
| | Gout | 274.1 | 25,756 | 60.73 | 10.97 | 116,386 | 55.23 | 11.88 | 46.49 | 11.94 | 18.08% | 29.10% | 11.81% |
| | Obesity | 278.1 | 5,019 | 52.98 | 11.59 | 134,513 | 56.37 | 11.91 | 43.66 | 11.86 | 3.52% | 2.99% | 3.82% |
| Mental Disorders | Alzheimer's disease | 290.11 | 324 | 71.86 | 6.11 | 127,824 | 55.70 | 11.77 | 69.14 | 5.58 | 0.23% | 0.21% | 0.24% |
| | Schizophrenia | 295.1 | 1,066 | 52.22 | 10.81 | 64,000 | 53.91 | 12.02 | 37.00 | 12.44 | 0.75% | 0.86% | 0.69% |
| | Depression | 296.2 | 12,526 | 57.76 | 11.46 | 64,000 | 53.91 | 12.02 | 44.75 | 12.76 | 8.79% | 6.29% | 10.22% |
| | Generalized anxiety disorder | 300.11 | 14,591 | 59.61 | 11.01 | 61,902 | 53.85 | 11.99 | 51.18 | 12.09 | 10.24% | 7.04% | 12.06% |
| | Alcoholism | 317.1 | 2,241 | 58.61 | 10.62 | 126,960 | 56.10 | 11.96 | 45.28 | 11.68 | 1.57% | 2.88% | 0.83% |
| Neurological | Migraine | 340 | 15,188 | 55.88 | 11.44 | 89,933 | 56.30 | 11.89 | 42.96 | 12.24 | 10.66% | 6.25% | 13.17% |
| | Epilepsy | 345.1 | 1,252 | 56.79 | 12.61 | 117,831 | 55.51 | 11.83 | 45.85 | 15.57 | 0.88% | 1.04% | 0.79% |

| | | | | | | | | | | | | | |
|---|---|---|---|---|---|---|---|---|---|---|---|---|---|
| | Genetic generalized epilepsy | 345.11 | 423 | 54.23 | 12.35 | 122,934 | 55.62 | 11.84 | 41.01 | 14.88 | 0.30% | 0.40% | 0.24% |
| **Sense Organs** | Hearing loss | 389 | 33,751 | 59.64 | 11.44 | 101,101 | 54.92 | 11.82 | 48.27 | 12.76 | 23.69% | 22.34% | 24.46% |
| **Circulatory System** | Hypertension | 401 | 46,783 | 62.59 | 9.96 | 89,368 | 52.72 | 11.42 | 50.92 | 10.10 | 32.83% | 39.60% | 28.99% |
| | Ischemic Heart Disease | 411 | 32,049 | 63.52 | 9.66 | 108,118 | 54.06 | 11.66 | 51.70 | 10.49 | 22.49% | 25.88% | 20.57% |
| | Myocardial infarction | 411.2 | 1,980 | 65.36 | 9.18 | 108,120 | 54.06 | 11.66 | 56.37 | 10.71 | 1.39% | 2.62% | 0.69% |
| | Atrial fibrillation | 427.21 | 2,064 | 66.91 | 8.60 | 98,731 | 54.99 | 11.82 | 59.61 | 10.57 | 1.45% | 2.13% | 1.06% |
| | Heart failure NOS | 428.2 | 958 | 65.40 | 9.49 | 137,471 | 55.94 | 11.86 | 51.80 | 10.44 | 0.67% | 0.70% | 0.66% |
| | Intracranial hemorrhage | 430 | 2,069 | 61.94 | 11.20 | 123,283 | 54.99 | 11.73 | 52.31 | 14.13 | 1.45% | 1.82% | 1.24% |
| | Subarachnoid hemorrhage | 430.1 | 627 | 60.70 | 11.10 | 123,287 | 54.99 | 11.73 | 50.63 | 13.45 | 0.44% | 0.46% | 0.43% |
| | Cerebrovascular disease | 433 | 18,196 | 64.48 | 9.59 | 123,288 | 54.99 | 11.73 | 55.07 | 11.02 | 12.77% | 13.36% | 12.43% |
| | Cerebral aneurysm | 433.5 | 500 | 62.25 | 10.10 | 123,290 | 54.99 | 11.73 | 55.99 | 11.23 | 0.35% | 0.28% | 0.39% |
| | Peripheral vascular disease | 443 | 6,401 | 63.38 | 10.12 | 129,439 | 55.54 | 11.84 | 52.81 | 11.64 | 4.49% | 3.97% | 4.79% |
| **Respiratory System** | Asthma | 495 | 29,028 | 57.62 | 12.40 | 80,366 | 54.65 | 11.60 | 44.62 | 14.25 | 20.37% | 18.10% | 21.67% |
| | Obstructive chronic bronchitis | 496.21 | 3,845 | 64.16 | 10.16 | 80,365 | 54.65 | 11.60 | 51.30 | 12.21 | 2.70% | 3.63% | 2.17% |
| **Digestive system** | GERD | 530.11 | 21,954 | 58.30 | 11.40 | 73,870 | 54.82 | 12.07 | 52.01 | 11.84 | 15.41% | 13.88% | 16.28% |
| | Inflammatory bowel disease and other gastroenteritis and colitis | 555 | 9,166 | 55.01 | 12.27 | 17,367 | 58.04 | 10.76 | 39.23 | 13.46 | 6.43% | 6.56% | 6.36% |
| | Chronic liver disease and cirrhosis | 571 | 17,267 | 59.91 | 10.83 | 103,395 | 55.22 | 11.97 | 48.17 | 12.17 | 12.12% | 14.89% | 10.54% |
| **Genitourinary system** | Chronic renal failure [CKD] | 585.3 | 7,058 | 64.57 | 9.91 | 107,826 | 54.92 | 11.82 | 57.40 | 11.47 | 4.95% | 6.77% | 3.92% |
| **Musculoskeletal** | Osteoarthrosis | 740 | 74,180 | 61.07 | 10.55 | 68,300 | 50.99 | 11.05 | 48.42 | 10.96 | 52.06% | 47.18% | 54.84% |
| | Back pain | 760 | 87,199 | 57.88 | 11.52 | 55,281 | 53.64 | 12.05 | 43.04 | 12.08 | 61.20% | 58.55% | 62.71% |

**Table S5. PRSe effects (top 10%) across all high-burden diseases**

| Category | Disease | PheCode | Effect | SE | HR | 95CI lower | 95CI upper | P_Value |
|---|---|---|---|---|---|---|---|---|
| **Neoplasms** | Cancer of esophagus | 150 | 0.30 | 0.22 | 1.35 | 0.87 | 2.09 | 1.82E-01 |
| | Cancer of stomach | 151 | 0.26 | 0.16 | 1.30 | 0.96 | 1.77 | 9.11E-02 |
| | Colorectal cancer | 153 | 0.28 | 0.06 | 1.33 | 1.17 | 1.51 | 9.54E-06 |
| | Cancer of bronchus; lung | 165.1 | 0.48 | 0.06 | 1.61 | 1.42 | 1.82 | 4.18E-14 |
| | Skin cancer | 172 | 0.20 | 0.14 | 1.22 | 0.93 | 1.60 | 1.52E-01 |
| | Melanomas of skin | 172.11 | -0.10 | 0.16 | 0.91 | 0.67 | 1.23 | 5.34E-01 |
| | Breast cancer | 174 | 0.29 | 0.04 | 1.34 | 1.25 | 1.44 | 1.11E-15 |
| | Cervical cancer and dysplasia | 180 | 0.14 | 0.04 | 1.15 | 1.07 | 1.24 | 2.15E-04 |
| | Cervical cancer | 180.1 | 0.32 | 0.17 | 1.37 | 0.99 | 1.90 | 5.71E-02 |
| | Malignant neoplasm of ovary | 184.11 | 0.31 | 0.13 | 1.37 | 1.06 | 1.76 | 1.46E-02 |
| | Cancer of prostate | 185 | 0.80 | 0.09 | 2.23 | 1.88 | 2.65 | 4.12E-20 |
| **Metabolic** | Type 2 diabetes | 250.2 | 0.76 | 0.01 | 2.13 | 2.07 | 2.19 | 0.00E+00 |
| | Hypercholesterolemia | 272.11 | 0.38 | 0.02 | 1.46 | 1.41 | 1.53 | 7.80E-73 |
| | Gout | 274.1 | 0.88 | 0.02 | 2.41 | 2.33 | 2.49 | 0.00E+00 |
| | Obesity | 278.1 | 0.49 | 0.04 | 1.63 | 1.51 | 1.76 | 1.06E-35 |
| **Mental Disorders** | Alzheimer's disease | 290.11 | 0.60 | 0.15 | 1.83 | 1.37 | 2.45 | 5.10E-05 |
| | Schizophrenia | 295.1 | 0.76 | 0.08 | 2.15 | 1.84 | 2.50 | 1.24E-22 |
| | Depression | 296.2 | 0.44 | 0.03 | 1.55 | 1.47 | 1.63 | 1.45E-64 |
| | Generalized anxiety disorder | 300.11 | 0.15 | 0.03 | 1.16 | 1.10 | 1.22 | 1.24E-08 |
| | Alcoholism | 317.1 | 0.29 | 0.06 | 1.34 | 1.18 | 1.52 | 3.34E-06 |
| **Neurological** | Migraine | 340 | 0.24 | 0.02 | 1.28 | 1.22 | 1.34 | 1.00E-22 |
| | Epilepsy | 345.1 | 0.08 | 0.09 | 1.09 | 0.91 | 1.30 | 3.54E-01 |
| | Genetic generalized epilepsy | 345.11 | 0.17 | 0.15 | 1.18 | 0.88 | 1.59 | 2.72E-01 |
| **Sense Organs** | Hearing loss | 389 | 0.07 | 0.02 | 1.07 | 1.03 | 1.11 | 1.49E-04 |
| **Circulatory System** | Hypertension | 401 | 0.59 | 0.01 | 1.81 | 1.76 | 1.86 | 0.00E+00 |
| | Ischemic Heart Disease | 411 | 0.13 | 0.02 | 1.14 | 1.10 | 1.18 | 8.50E-13 |
| | Myocardial infarction | 411.2 | 0.60 | 0.06 | 1.82 | 1.61 | 2.05 | 4.32E-22 |
| | Atrial fibrillation | 427.21 | 1.27 | 0.05 | 3.56 | 3.23 | 3.92 | 3.00E-146 |
| | Heart failure NOS | 428.2 | 0.28 | 0.10 | 1.33 | 1.10 | 1.61 | 3.54E-03 |
| | Intracranial hemorrhage | 430 | 0.44 | 0.06 | 1.56 | 1.38 | 1.76 | 1.51E-12 |
| | Subarachnoid hemorrhage | 430.1 | 0.21 | 0.12 | 1.23 | 0.96 | 1.56 | 9.53E-02 |
| | Cerebrovascular disease | 433 | 0.14 | 0.02 | 1.15 | 1.10 | 1.20 | 4.55E-09 |
| | Cerebral aneurysm | 433.5 | 0.21 | 0.14 | 1.23 | 0.94 | 1.61 | 1.32E-01 |
| | Peripheral vascular disease | 443 | 0.10 | 0.04 | 1.11 | 1.03 | 1.20 | 8.99E-03 |
| **Respiratory System** | Asthma | 495 | 0.49 | 0.02 | 1.64 | 1.58 | 1.69 | 1.65E-186 |
| | Obstructive chronic bronchitis | 496.21 | 0.35 | 0.05 | 1.43 | 1.30 | 1.56 | 5.82E-14 |

| | | | | | | | | |
|---|---|---|---|---|---|---|---|---|
| **Digestive system** | GERD | 530.11 | 0.16 | 0.02 | 1.17 | 1.12 | 1.22 | 1.06E-13 |
| | Inflammatory bowel disease and other gastroenteritis and colitis | 555 | 0.04 | 0.03 | 1.04 | 0.97 | 1.11 | 2.34E-01 |
| | Chronic liver disease and cirrhosis | 571 | 0.18 | 0.02 | 1.20 | 1.15 | 1.26 | 1.05E-14 |
| **Genitourinary system** | Chronic renal failure [CKD] | 585.3 | 0.24 | 0.04 | 1.27 | 1.18 | 1.37 | 4.03E-11 |
| **Musculoskeletal** | Osteoarthrosis | 740 | 0.16 | 0.01 | 1.17 | 1.14 | 1.20 | 2.94E-39 |
| | Back pain | 760 | 0.10 | 0.01 | 1.10 | 1.08 | 1.12 | 6.07E-18 |

#### Table S6. Sex-specific PRSe effects (top 10%) across all high-burden diseases

| | | | Male | | | | | | Female | | | | | |
|---|---|---|---|---|---|---|---|---|---|---|---|---|---|---|
| Category | Disease | PheCode | Effect | SE | HR | 95CI lower | 95CI upper | P_Value | Effect | SE | HR | 95CI lower | 95CI upper | P_Value |
| **Neoplasms** | Cancer of esophagus | 150 | 0.04 | 0.48 | 1.04 | 0.41 | 2.68 | 9.27E-01 | 0.36 | 0.25 | 1.43 | 0.87 | 2.35 | 1.58E-01 |
| | Cancer of stomach | 151 | 0.15 | 0.26 | 1.16 | 0.70 | 1.93 | 5.63E-01 | 0.33 | 0.20 | 1.39 | 0.94 | 2.04 | 9.53E-02 |
| | Colorectal cancer | 153 | 0.38 | 0.10 | 1.46 | 1.21 | 1.77 | 8.08E-05 | 0.21 | 0.09 | 1.24 | 1.04 | 1.46 | 1.42E-02 |
| | Cancer of bronchus; lung | 165.1 | 0.19 | 0.11 | 1.20 | 0.96 | 1.51 | 1.07E-01 | 0.60 | 0.08 | 1.82 | 1.57 | 2.12 | 4.40E-15 |
| | Skin cancer | 172 | 0.39 | 0.20 | 1.48 | 0.99 | 2.20 | 5.33E-02 | 0.00 | 0.19 | 1.00 | 0.69 | 1.47 | 9.81E-01 |
| | Melanomas of skin | 172.11 | -0.18 | 0.26 | 0.83 | 0.50 | 1.39 | 4.84E-01 | -0.04 | 0.20 | 0.96 | 0.66 | 1.41 | 8.51E-01 |
| | Breast cancer | 174 | 0.36 | 0.41 | 1.44 | 0.65 | 3.20 | 3.72E-01 | 0.30 | 0.04 | 1.34 | 1.25 | 1.44 | 4.82E-16 |
| | Cervical cancer and dysplasia | 180 | NA | NA | NA | NA | NA | NA | 0.14 | 0.04 | 1.15 | 1.07 | 1.24 | 2.10E-04 |
| | Cervical cancer | 180.1 | NA | NA | NA | NA | NA | NA | 0.27 | 0.17 | 1.31 | 0.94 | 1.82 | 1.09E-01 |
| | Malignant neoplasm of ovary | 184.11 | NA | NA | NA | NA | NA | NA | 0.32 | 0.13 | 1.38 | 1.08 | 1.77 | 1.07E-02 |
| | Cancer of prostate | 185 | 0.81 | 0.09 | 2.24 | 1.89 | 2.66 | 2.80E-20 | NA | NA | NA | NA | NA | NA |
| **Metabolic** | Type 2 diabetes | 250.2 | 0.82 | 0.02 | 2.26 | 2.16 | 2.37 | 5.38E-262 | 0.72 | 0.02 | 2.06 | 1.98 | 2.14 | 0E+00 |
| | Hypercholesterolemia | 272.11 | 0.43 | 0.04 | 1.53 | 1.43 | 1.65 | 4.42E-32 | 0.36 | 0.03 | 1.43 | 1.36 | 1.51 | 2.70E-43 |
| | Gout | 274.1 | 1.24 | 0.02 | 3.46 | 3.32 | 3.60 | 0.00E+00 | 0.32 | 0.03 | 1.37 | 1.30 | 1.45 | 2.97E-28 |
| | Obesity | 278.1 | 0.43 | 0.07 | 1.54 | 1.34 | 1.77 | 1.82E-09 | 0.51 | 0.05 | 1.66 | 1.52 | 1.82 | 1.88E-27 |
| **Mental Disorders** | Alzheimer's disease | 290.11 | 0.35 | 0.28 | 1.42 | 0.82 | 2.46 | 2.08E-01 | 0.73 | 0.18 | 2.07 | 1.46 | 2.94 | 4.08E-05 |
| | Schizophrenia | 295.1 | 0.79 | 0.12 | 2.21 | 1.75 | 2.79 | 3.83E-11 | 0.73 | 0.10 | 2.08 | 1.70 | 2.55 | 1.15E-12 |
| | Depression | 296.2 | 0.42 | 0.05 | 1.52 | 1.38 | 1.68 | 7.66E-17 | 0.44 | 0.03 | 1.56 | 1.47 | 1.65 | 3.92E-49 |
| | Generalized anxiety disorder | 300.11 | 0.20 | 0.05 | 1.23 | 1.11 | 1.36 | 7.17E-05 | 0.14 | 0.03 | 1.14 | 1.08 | 1.22 | 8.48E-06 |
| | Alcoholism | 317.1 | 0.36 | 0.08 | 1.43 | 1.23 | 1.66 | 2.55E-06 | 0.15 | 0.11 | 1.16 | 0.93 | 1.45 | 1.96E-01 |
| **Neurological** | Migraine | 340 | 0.20 | 0.05 | 1.22 | 1.09 | 1.35 | 3.43E-04 | 0.26 | 0.03 | 1.29 | 1.22 | 1.37 | 2.85E-20 |
| | Epilepsy | 345.1 | 0.00 | 0.14 | 1.00 | 0.75 | 1.32 | 9.86E-01 | 0.14 | 0.12 | 1.15 | 0.92 | 1.46 | 2.23E-01 |
| | Genetic generalized epilepsy | 345.11 | 0.18 | 0.22 | 1.20 | 0.78 | 1.83 | 4.05E-01 | 0.17 | 0.21 | 1.18 | 0.78 | 1.79 | 4.37E-01 |
| **Sense Organs** | Hearing loss | 389 | 0.11 | 0.03 | 1.12 | 1.06 | 1.19 | 1.16E-04 | 0.04 | 0.02 | 1.04 | 1.00 | 1.09 | 5.89E-02 |
| **Circulatory System** | Hypertension | 401 | 0.60 | 0.02 | 1.82 | 1.75 | 1.89 | 1.83E-185 | 0.59 | 0.02 | 1.81 | 1.75 | 1.88 | 1.40E-243 |
| | Ischemic Heart Disease | 411 | 0.17 | 0.03 | 1.19 | 1.12 | 1.25 | 4.34E-10 | 0.10 | 0.02 | 1.10 | 1.05 | 1.15 | 4.18E-05 |
| | Myocardial infarction | 411.2 | 0.65 | 0.07 | 1.91 | 1.65 | 2.20 | 1.33E-18 | 0.49 | 0.11 | 1.64 | 1.31 | 2.04 | 1.32E-05 |

| | | | | | | | | | | | | | | |
|---|---|---|---|---|---|---|---|---|---|---|---|---|---|---|
| | Atrial fibrillation | 427.21 | 1.36 | 0.07 | 3.90 | 3.42 | 4.45 | 8.38E-93 | 1.17 | 0.07 | 3.23 | 2.80 | 3.73 | 1.40E-57 |
| | Heart failure NOS | 428.2 | 0.32 | 0.16 | 1.38 | 1.02 | 1.88 | 3.98E-02 | 0.27 | 0.13 | 1.31 | 1.02 | 1.67 | 3.22E-02 |
| | Intracranial hemorrhage | 430 | 0.62 | 0.09 | 1.85 | 1.56 | 2.20 | 2.99E-12 | 0.29 | 0.09 | 1.34 | 1.13 | 1.60 | 9.47E-04 |
| | Subarachnoid hemorrhage | 430.1 | -0.02 | 0.22 | 0.98 | 0.64 | 1.50 | 9.20E-01 | 0.33 | 0.15 | 1.39 | 1.03 | 1.86 | 2.87E-02 |
| | Cerebrovascular disease | 433 | 0.14 | 0.04 | 1.15 | 1.06 | 1.24 | 3.82E-04 | 0.13 | 0.03 | 1.14 | 1.08 | 1.21 | 8.24E-06 |
| | Cerebral aneurysm | 433.5 | 0.14 | 0.27 | 1.15 | 0.68 | 1.93 | 6.07E-01 | 0.27 | 0.16 | 1.31 | 0.96 | 1.79 | 9.44E-02 |
| | Peripheral vascular disease | 443 | 0.23 | 0.07 | 1.26 | 1.10 | 1.43 | 8.20E-04 | 0.03 | 0.05 | 1.03 | 0.94 | 1.14 | 4.95E-01 |
| **Respiratory System** | Asthma | 495 | 0.48 | 0.03 | 1.61 | 1.52 | 1.71 | 2.10E-57 | 0.50 | 0.02 | 1.64 | 1.58 | 1.71 | 1.64E-128 |
| | Obstructive chronic bronchitis | 496.21 | 0.45 | 0.07 | 1.57 | 1.38 | 1.78 | 6.18E-12 | 0.26 | 0.07 | 1.29 | 1.13 | 1.48 | 1.85E-04 |
| **Digestive system** | GERD | 530.11 | 0.19 | 0.04 | 1.21 | 1.12 | 1.30 | 3.21E-07 | 0.14 | 0.03 | 1.15 | 1.09 | 1.21 | 6.75E-08 |
| | Inflammatory bowel disease and other gastroenteritis and colitis | 555 | 0.11 | 0.06 | 1.11 | 1.00 | 1.24 | 5.43E-02 | -0.01 | 0.04 | 0.99 | 0.91 | 1.08 | 8.52E-01 |
| | Chronic liver disease and cirrhosis | 571 | 0.19 | 0.04 | 1.21 | 1.13 | 1.30 | 6.61E-08 | 0.18 | 0.03 | 1.20 | 1.13 | 1.28 | 1.02E-08 |
| **Genitourinary system** | Chronic renal failure [CKD] | 585.3 | 0.25 | 0.05 | 1.29 | 1.16 | 1.43 | 1.04E-06 | 0.22 | 0.05 | 1.25 | 1.13 | 1.38 | 1.41E-05 |
| **Musculoskeletal** | Osteoarthrosis | 740 | 0.13 | 0.02 | 1.14 | 1.10 | 1.19 | 7.28E-11 | 0.17 | 0.01 | 1.18 | 1.15 | 1.22 | 3.48E-31 |
| | Back pain | 760 | 0.05 | 0.02 | 1.06 | 1.02 | 1.10 | 4.42E-03 | 0.12 | 0.01 | 1.12 | 1.09 | 1.15 | 8.56E-18 |

#### Table S7. Sensitivity analyses of PRSe effects (top 5% and top 1%) across all high-burden diseases

| | | | Top 5% | | | | | | Top 1% | | | | | |
|---|---|---|---|---|---|---|---|---|---|---|---|---|---|---|
| **Category** | **Disease** | **PheCode** | **Effect** | **SE** | **HR** | **95CI lower** | **95CI upper** | **P_Value** | **Effect** | **SE** | **HR** | **95CI lower** | **95CI upper** | **P_Value** |
| **Neoplasms** | Cancer of esophagus | 150 | 0.12 | 0.33 | 1.13 | 0.59 | 2.1304 | 7.17E-01 | -15.02 | 1370.12 | 0.00 | 0.00 | infinite | 9.91E-01 |
| | Cancer of stomach | 151 | 0.17 | 0.23 | 1.19 | 0.76 | 1.87 | 4.52E-01 | -1.33 | 1.00 | 0.27 | 0.04 | 1.89 | 1.85E-01 |
| | Colorectal cancer | 153 | 0.37 | 0.08 | 1.45 | 1.23 | 1.71 | 1.15E-05 | 0.60 | 0.16 | 1.82 | 1.32 | 2.51 | 2.54E-04 |
| | Cancer of bronchus; lung | 165.1 | 0.55 | 0.08 | 1.74 | 1.48 | 2.04 | 1.46E-11 | 0.99 | 0.14 | 2.70 | 2.05 | 3.57 | 2.52E-12 |
| | Skin cancer | 172 | -0.06 | 0.22 | 0.94 | 0.61 | 1.45 | 7.83E-01 | -0.53 | 0.58 | 0.59 | 0.19 | 1.83 | 3.58E-01 |
| | Melanomas of skin | 172.11 | 0.20 | 0.20 | 1.22 | 0.82 | 1.79 | 3.25E-01 | 0.21 | 0.41 | 1.24 | 0.55 | 2.76 | 6.07E-01 |
| | Breast cancer | 174 | 0.30 | 0.05 | 1.35 | 1.22 | 1.50 | 3.50E-09 | 0.48 | 0.10 | 1.61 | 1.33 | 1.95 | 8.14E-07 |
| | Cervical cancer and dysplasia | 180 | 0.02 | 0.05 | 1.02 | 0.92 | 1.13 | 7.30E-01 | 0.17 | 0.11 | 1.18 | 0.95 | 1.46 | 1.26E-01 |
| | Cervical cancer | 180.1 | 0.02 | 0.05 | 1.02 | 0.92 | 1.13 | 7.30E-01 | 0.17 | 0.11 | 1.18 | 0.95 | 1.46 | 1.26E-01 |
| | Malignant neoplasm of ovary | 184.11 | 0.36 | 0.18 | 1.44 | 1.02 | 2.04 | 4.04E-02 | 0.12 | 0.41 | 1.13 | 0.50 | 2.52 | 7.68E-01 |
| | Cancer of prostate | 185 | 0.84 | 0.12 | 2.32 | 1.83 | 2.95 | 6.38E-12 | 0.90 | 0.23 | 2.46 | 1.58 | 3.84 | 7.41E-05 |
| **Metabolic** | Type 2 diabetes | 250.2 | 0.84 | 0.02 | 2.31 | 2.23 | 2.41 | 0.00E+00 | 0.97 | 0.04 | 2.65 | 2.46 | 2.86 | 3.00E-138 |
| | Hypercholesterolemia | 272.11 | 0.40 | 0.03 | 1.49 | 1.41 | 1.58 | 1.28E-42 | 0.56 | 0.06 | 1.75 | 1.56 | 1.97 | 1.73E-20 |
| | Gout | 274.1 | 1.08 | 0.02 | 2.96 | 2.84 | 3.08 | 0.00E+00 | 1.53 | 0.04 | 4.62 | 4.31 | 4.96 | 0.00E+00 |
| | Obesity | 278.1 | 0.57 | 0.05 | 1.77 | 1.60 | 1.97 | 7.84E-28 | 0.88 | 0.09 | 2.40 | 2.00 | 2.89 | 6.83E-21 |
| **Mental Disorders** | Alzheimer's disease | 290.11 | 0.72 | 0.19 | 2.05 | 1.42 | 2.96 | 1.19E-04 | 1.49 | 0.29 | 4.44 | 2.49 | 7.91 | 4.33E-07 |
| | Schizophrenia | 295.1 | 0.78 | 0.10 | 2.17 | 1.77 | 2.67 | 1.18E-13 | 0.99 | 0.20 | 2.69 | 1.84 | 3.94 | 3.83E-07 |
| | Depression | 296.2 | 0.53 | 0.04 | 1.70 | 1.59 | 1.82 | 5.88E-52 | 0.66 | 0.07 | 1.93 | 1.68 | 2.22 | 4.12E-20 |
| | Generalized anxiety disorder | 300.11 | 0.18 | 0.04 | 1.20 | 1.12 | 1.29 | 2.83E-07 | 0.23 | 0.08 | 1.26 | 1.09 | 1.46 | 2.26E-03 |
| | Alcoholism | 317.1 | 0.30 | 0.08 | 1.35 | 1.15 | 1.60 | 3.56E-04 | 0.44 | 0.17 | 1.56 | 1.12 | 2.18 | 9.17E-03 |
| **Neurological** | Migraine | 340 | 0.27 | 0.03 | 1.31 | 1.23 | 1.40 | 5.65E-16 | 0.35 | 0.07 | 1.41 | 1.23 | 1.63 | 1.46E-06 |
| | Epilepsy | 345.1 | -0.11 | 0.14 | 0.89 | 0.68 | 1.17 | 4.16E-01 | -0.45 | 0.35 | 0.64 | 0.32 | 1.28 | 2.04E-01 |
| | Genetic generalized epilepsy | 345.11 | 0.25 | 0.21 | 1.28 | 0.86 | 1.92 | 2.29E-01 | 0.19 | 0.45 | 1.20 | 0.50 | 2.91 | 6.80E-01 |
| **Sense Organs** | Hearing loss | 389 | 0.10 | 0.02 | 1.11 | 1.06 | 1.16 | 1.97E-05 | 0.17 | 0.05 | 1.18 | 1.07 | 1.31 | 1.28E-03 |
| **Circulatory System** | Hypertension | 401 | 0.66 | 0.02 | 1.93 | 1.86 | 2.00 | 6.73E-297 | 0.79 | 0.04 | 2.20 | 2.05 | 2.37 | 6.59E-102 |
| | Ischemic Heart Disease | 411 | 0.16 | 0.03 | 1.18 | 1.12 | 1.23 | 1.16E-10 | 0.22 | 0.05 | 1.24 | 1.12 | 1.37 | 3.42E-05 |

| | | | | | | | | | | | | | | |
|---|---|---|---|---|---|---|---|---|---|---|---|---|---|---|
| | Myocardial infarction | 411.2 | 0.80 | 0.08 | 2.22 | 1.91 | 2.60 | 2.94E-24 | 0.85 | 0.15 | 2.34 | 1.74 | 3.16 | 2.60E-08 |
| | Atrial fibrillation | 427.21 | 1.37 | 0.06 | 3.93 | 3.48 | 4.44 | 2.02E-107 | 1.69 | 0.10 | 5.44 | 4.46 | 6.63 | 1.24E-62 |
| | Heart failure NOS | 428.2 | 0.36 | 0.13 | 1.44 | 1.11 | 1.87 | 6.72E-03 | 0.41 | 0.28 | 1.50 | 0.87 | 2.60 | 1.45E-01 |
| | Intracranial hemorrhage | 430 | 0.48 | 0.09 | 1.62 | 1.37 | 1.93 | 2.84E-08 | 1.18 | 0.13 | 3.27 | 2.52 | 4.25 | 6.24E-19 |
| | Subarachnoid hemorrhage | 430.1 | 0.07 | 0.18 | 1.07 | 0.74 | 1.53 | 7.23E-01 | -0.45 | 0.50 | 0.64 | 0.24 | 1.71 | 3.72E-01 |
| | Cerebrovascular disease | 433 | 0.17 | 0.03 | 1.18 | 1.11 | 1.26 | 4.24E-07 | 0.24 | 0.07 | 1.27 | 1.11 | 1.45 | 4.89E-04 |
| | Cerebral aneurysm | 433.5 | 0.34 | 0.19 | 1.41 | 0.97 | 2.05 | 6.88E-02 | 0.96 | 0.28 | 2.61 | 1.51 | 4.54 | 6.33E-04 |
| | Peripheral vascular disease | 443 | 0.05 | 0.06 | 1.06 | 0.94 | 1.18 | 3.54E-01 | 0.18 | 0.12 | 1.20 | 0.95 | 1.50 | 1.23E-01 |
| **Respiratory System** | Asthma | 495 | 0.62 | 0.02 | 1.85 | 1.77 | 1.93 | 6.19E-171 | 1.06 | 0.04 | 2.88 | 2.67 | 3.11 | 3.19E-161 |
| | Obstructive chronic bronchitis | 496.21 | 0.39 | 0.07 | 1.48 | 1.30 | 1.68 | 3.29E-09 | 0.74 | 0.12 | 2.09 | 1.64 | 2.66 | 1.73E-09 |
| **Digestive system** | GERD | 530.11 | 0.21 | 0.03 | 1.24 | 1.17 | 1.31 | 9.38E-13 | 0.23 | 0.06 | 1.26 | 1.12 | 1.42 | 1.92E-04 |
| | Inflammatory bowel disease and other gastroenteritis and colitis | 555 | 0.08 | 0.05 | 1.08 | 0.99 | 1.19 | 9.63E-02 | 0.15 | 0.10 | 1.16 | 0.96 | 1.41 | 1.28E-01 |
| | Chronic liver disease and cirrhosis | 571 | 0.24 | 0.03 | 1.27 | 1.19 | 1.35 | 2.42E-13 | 0.26 | 0.07 | 1.29 | 1.13 | 1.48 | 1.74E-04 |
| **Genitourinary system** | Chronic renal failure [CKD] | 585.3 | 0.24 | 0.05 | 1.27 | 1.15 | 1.40 | 1.28E-06 | 0.46 | 0.10 | 1.58 | 1.31 | 1.92 | 1.85E-06 |
| **Musculoskeletal** | Osteoarthrosis | 740 | 0.17 | 0.02 | 1.18 | 1.15 | 1.22 | 5.29E-24 | 0.24 | 0.03 | 1.27 | 1.19 | 1.36 | 3.78E-12 |
| | Back pain | 760 | 0.10 | 0.02 | 1.10 | 1.07 | 1.14 | 5.15E-10 | 0.07 | 0.03 | 1.07 | 1.01 | 1.15 | 3.44E-02 |

#### Table S8. Overall and sex-specific PAF associated with PRSe (top 10%) for diseases with significant genetic effects

| | | | All | | | Male | | | Female | | |
|---|---|---|---|---|---|---|---|---|---|---|---|
| **Category** | **Disease** | **PheCode** | **PAF** | **95CI lower** | **95CI upper** | **PAF** | **95CI lower** | **95CI upper** | **PAF** | **95CI lower** | **95CI upper** |
| **Neoplasms** | Colorectal cancer | 153 | 0.03 | 0.02 | 0.05 | 0.04 | 0.02 | 0.07 | 0.02 | 0.00 | 0.05 |
| | Cancer of bronchus; lung | 165.1 | 0.06 | 0.04 | 0.08 | 0.02 | -0.01 | 0.05 | 0.08 | 0.06 | 0.10 |
| | Breast cancer | 174 | 0.03 | 0.02 | 0.04 | 0.04 | -0.06 | 0.16 | 0.03 | 0.02 | 0.04 |
| | Cervical cancer and dysplasia | 180 | 0.01 | 0.01 | 0.02 | NA | NA | NA | 0.01 | 0.01 | 0.02 |
| | Malignant neoplasm of ovary | 184.11 | 0.04 | 0.00 | 0.07 | NA | NA | NA | 0.04 | 0.01 | 0.07 |
| | Cancer of prostate | 185 | 0.11 | 0.08 | 0.14 | 0.11 | 0.08 | 0.14 | NA | NA | NA |
| **Metabolic** | Type 2 diabetes | 250.2 | 0.10 | 0.10 | 0.11 | 0.11 | 0.10 | 0.12 | 0.10 | 0.09 | 0.10 |
| | Hypercholesterolemia | 272.11 | 0.04 | 0.04 | 0.05 | 0.05 | 0.04 | 0.06 | 0.04 | 0.04 | 0.05 |
| | Gout | 274.1 | 0.12 | 0.12 | 0.13 | 0.20 | 0.19 | 0.21 | 0.04 | 0.03 | 0.04 |
| | Obesity | 278.1 | 0.06 | 0.05 | 0.07 | 0.05 | 0.03 | 0.07 | 0.06 | 0.05 | 0.07 |
| **Mental Disorders** | Alzheimer's disease | 290.11 | 0.08 | 0.04 | 0.12 | 0.04 | -0.02 | 0.12 | 0.10 | 0.04 | 0.16 |
| | Schizophrenia | 295.1 | 0.10 | 0.08 | 0.13 | 0.11 | 0.07 | 0.15 | 0.10 | 0.07 | 0.13 |
| | Depression | 296.2 | 0.05 | 0.04 | 0.06 | 0.05 | 0.04 | 0.06 | 0.05 | 0.04 | 0.06 |
| | Generalized anxiety disorder | 300.11 | 0.02 | 0.01 | 0.02 | 0.02 | 0.01 | 0.03 | 0.01 | 0.01 | 0.02 |
| | Alcoholism | 317.1 | 0.03 | 0.02 | 0.05 | 0.04 | 0.03 | 0.06 | 0.02 | -0.01 | 0.04 |
| **Neurological** | Migraine | 340 | 0.03 | 0.02 | 0.03 | 0.02 | 0.01 | 0.03 | 0.03 | 0.02 | 0.04 |
| **Sense Organs** | Hearing loss | 389 | 0.01 | 0.00 | 0.01 | 0.01 | 0.01 | 0.02 | 0.00 | 0.00 | 0.01 |
| **Circulatory System** | Hypertension | 401 | 0.07 | 0.07 | 0.08 | 0.08 | 0.07 | 0.08 | 0.07 | 0.07 | 0.08 |
| | Ischemic Heart Disease | 411 | 0.01 | 0.01 | 0.02 | 0.02 | 0.01 | 0.02 | 0.01 | 0.00 | 0.02 |
| | Myocardial infarction | 411.2 | 0.08 | 0.06 | 0.09 | 0.08 | 0.06 | 0.11 | 0.06 | 0.03 | 0.10 |
| | Atrial fibrillation | 427.21 | 0.20 | 0.18 | 0.23 | 0.22 | 0.20 | 0.26 | 0.18 | 0.15 | 0.21 |
| | Heart failure NOS | 428.2 | 0.03 | 0.01 | 0.06 | 0.04 | 0.00 | 0.08 | 0.03 | 0.00 | 0.06 |

| | | | | | | | | | | | |
|---|---|---|---|---|---|---|---|---|---|---|---|
| | Intracranial hemorrhage | 430 | 0.05 | 0.04 | 0.07 | 0.08 | 0.05 | 0.11 | 0.03 | 0.01 | 0.05 |
| | Cerebrovascular disease | 433 | 0.01 | 0.01 | 0.02 | 0.01 | 0.01 | 0.02 | 0.01 | 0.01 | 0.02 |
| | Peripheral vascular disease | 443 | 0.01 | 0.00 | 0.02 | 0.02 | 0.01 | 0.04 | 0.00 | -0.01 | 0.01 |
| **Respiratory System** | Asthma | 495 | 0.06 | 0.05 | 0.06 | 0.06 | 0.05 | 0.06 | 0.06 | 0.06 | 0.07 |
| | Obstructive chronic bronchitis | 496.21 | 0.04 | 0.03 | 0.05 | 0.05 | 0.03 | 0.07 | 0.03 | 0.01 | 0.05 |
| **Digestive system** | GERD | 530.11 | 0.02 | 0.01 | 0.02 | 0.02 | 0.01 | 0.03 | 0.01 | 0.01 | 0.02 |
| | Chronic liver disease and cirrhosis | 571 | 0.02 | 0.01 | 0.03 | 0.02 | 0.01 | 0.03 | 0.02 | 0.01 | 0.03 |
| **Genitourinary system** | Chronic renal failure [CKD] | 585.3 | 0.03 | 0.02 | 0.04 | 0.03 | 0.02 | 0.04 | 0.02 | 0.01 | 0.04 |
| **Musculoskeletal** | Osteoarthrosis | 740 | 0.02 | 0.01 | 0.02 | 0.01 | 0.01 | 0.02 | 0.02 | 0.01 | 0.02 |
| | Back pain | 760 | 0.01 | 0.01 | 0.01 | 0.01 | 0.00 | 0.01 | 0.01 | 0.01 | 0.02 |

**Table S9. Overall and sex-specific population DALYs (DALYpop) attributable to PRSe (top 10%) for significant diseases with available DALYs estimates**

| | | | All | | | |
|---|---|---|---|---|---|---|
| **Category** | **Disease** | **PheCode** | **Total DALYs** | **DALYpop** | **95CI lower** | **95CI upper** |
| **Neoplasms** | Colorectal cancer | 153 | 1044.26 | 33.32 | 18.62 | 51.61 |
| | Cancer of bronchus; lung | 165.1 | 1110.40 | 64.06 | 44.36 | 83.91 |
| | Breast cancer | 174 | 413.49 | 13.54 | 10.15 | 17.13 |
| | Cervical cancer and dysplasia | 180 | 101.34 | 1.49 | 0.67 | 2.38 |
| | Malignant neoplasm of ovary | 184.11 | 102.77 | 3.64 | 0.40 | 7.13 |
| | Cancer of prostate | 185 | 192.30 | 21.06 | 14.77 | 27.16 |
| **Metabolic** | Type 2 diabetes | 250.2 | 1836.35 | 186.44 | 177.28 | 196.20 |
| | Gout | 274.1 | 55.57 | 6.86 | 6.53 | 7.20 |
| **Mental Disorders** | Alzheimer's disease | 290.11 | 819.55 | 62.85 | 31.89 | 101.99 |
| | Schizophrenia | 295.1 | 281.40 | 28.92 | 21.24 | 35.77 |
| | Depression | 296.2 | 486.71 | 25.28 | 21.83 | 28.77 |
| | Generalized anxiety disorder | 300.11 | 452.57 | 7.15 | 4.79 | 10.04 |
| | Alcoholism | 317.1 | 260.60 | 8.59 | 4.68 | 12.38 |
| **Neurological** | Migraine | 340 | 515.57 | 13.83 | 10.78 | 16.80 |
| **Sense Organs** | Hearing loss | 389 | 925.33 | 6.38 | 2.94 | 9.75 |
| **Circulatory System** | Ischemic Heart Disease | 411 | 1403.97 | 18.98 | 13.10 | 24.35 |
| | Atrial fibrillation | 427.21 | 179.39 | 36.53 | 32.94 | 40.62 |
| | Intracranial hemorrhage | 430 | 1353.39 | 71.56 | 49.97 | 94.74 |
| **Respiratory System** | Asthma | 495 | 251.58 | 15.04 | 13.59 | 16.03 |
| | Obstructive chronic bronchitis | 496.21 | 637.46 | 26.00 | 17.34 | 33.86 |
| **Digestive system** | GERD | 530.11 | 53.57 | 0.91 | 0.62 | 1.16 |
| | Chronic liver disease and cirrhosis | 571 | 729.04 | 14.48 | 10.37 | 18.36 |

| | | | | | | |
|---|---|---|---|---|---|---|
| **Musculoskeletal** | Osteoarthrosis | 740 | 500.12 | 8.28 | 6.95 | 9.67 |
| | Back pain | 760 | 1369.95 | 13.60 | 10.21 | 16.64 |

| | | | **Male** | | | | **Female** | | | |
|---|---|---|---|---|---|---|---|---|---|---|
| **Category** | **Disease** | **PheCode** | **Total DALYs** | **DALYpop** | **95CI lower** | **95CI upper** | **Total DALYs** | **DALYpop** | **95CI lower** | **95CI upper** |
| **Neoplasms** | Colorectal cancer | 153 | 1347.42 | 59.57 | 29.85 | 95.85 | 748.27 | 17.27 | 3.09 | 33.81 |
| | Cancer of bronchus; lung | 165.1 | 1396.48 | 27.83 | -9.18 | 63.40 | 831.08 | 63.21 | 45.87 | 83.95 |
| | Breast cancer | 174 | 9.17 | 0.39 | -0.51 | 1.50 | 808.25 | 26.83 | 19.73 | 34.02 |
| | Cervical cancer and dysplasia | 180 | NA | NA | NA | NA | 330.09 | 4.85 | 2.08 | 7.37 |
| | Malignant neoplasm of ovary | 184.11 | NA | NA | NA | NA | 161.24 | 5.93 | 1.19 | 11.85 |
| | Cancer of prostate | 185 | 389.25 | 42.88 | 30.45 | 54.04 | NA | NA | NA | NA |
| **Metabolic** | Type 2 diabetes | 250.2 | 2016.03 | 226.34 | 208.67 | 242.59 | 1660.92 | 159.01 | 147.90 | 169.40 |
| | Gout | 274.1 | 81.91 | 16.16 | 15.40 | 16.92 | 29.87 | 1.08 | 0.87 | 1.30 |
| **Mental Disorders** | Alzheimer's disease | 290.11 | 552.61 | 22.33 | -13.46 | 65.27 | 1080.18 | 104.61 | 43.34 | 176.16 |
| | Schizophrenia | 295.1 | 311.81 | 33.62 | 21.53 | 47.76 | 251.71 | 24.55 | 16.48 | 33.65 |
| | Depression | 296.2 | 348.53 | 17.32 | 13.04 | 22.64 | 621.62 | 32.82 | 27.80 | 38.21 |
| | Generalized anxiety disorder | 300.11 | 338.01 | 7.48 | 3.05 | 11.32 | 564.41 | 8.06 | 4.11 | 11.56 |
| | Alcoholism | 317.1 | 471.38 | 19.40 | 11.82 | 29.73 | 54.80 | 0.86 | -0.49 | 2.24 |
| **Neurological** | Migraine | 340 | 305.64 | 6.46 | 2.83 | 10.30 | 720.53 | 20.53 | 15.50 | 25.46 |
| **Sense Organs** | Hearing loss | 389 | 900.00 | 10.82 | 5.06 | 16.81 | 950.06 | 4.02 | -0.14 | 8.24 |
| **Circulatory System** | Ischemic Heart Disease | 411 | 1918.92 | 35.27 | 23.47 | 47.00 | 901.20 | 9.10 | 4.48 | 13.81 |
| | Atrial fibrillation | 427.21 | 176.36 | 39.67 | 34.44 | 45.12 | 182.36 | 33.27 | 27.59 | 38.92 |
| | Intracranial hemorrhage | 430 | 1730.52 | 135.53 | 92.24 | 191.48 | 985.18 | 32.61 | 11.06 | 53.81 |
| **Respiratory System** | Asthma | 495 | 219.45 | 12.60 | 10.35 | 14.24 | 282.95 | 17.07 | 15.60 | 19.00 |
| | Obstructive chronic bronchitis | 496.21 | 899.16 | 48.18 | 30.57 | 63.06 | 381.95 | 10.84 | 4.18 | 17.34 |
| **Digestive system** | GERD | 530.11 | 49.37 | 1.01 | 0.58 | 1.46 | 57.66 | 0.86 | 0.53 | 1.20 |

| | | | | | | | | | | |
|---|---|---|---|---|---|---|---|---|---|---|
| | Chronic liver disease and cirrhosis | 571 | 1145.18 | 23.80 | 15.78 | 34.34 | 322.74 | 6.39 | 4.00 | 8.59 |
| **Musculoskeletal** | Osteoarthrosis | 740 | 387.52 | 5.50 | 3.78 | 7.27 | 610.05 | 11.00 | 8.93 | 13.03 |
| | Back pain | 760 | 1202.96 | 6.64 | 1.35 | 11.51 | 1532.99 | 18.82 | 13.92 | 23.71 |

#### Table S10. Sensitivity analyses for PAF of significant diseases (top 5% and top 1%)

| | | | Top 5% | | | Top 1% | | |
|---|---|---|---|---|---|---|---|---|
| **Category** | **Disease** | **PheCode** | **PAF** | **95CI lower** | **95CI upper** | **PAF** | **95CI lower** | **95CI upper** |
| **Neoplasms** | Colorectal cancer | 153 | 0.02 | 0.01 | 0.03 | 0.01 | 0.00 | 0.01 |
| | Cancer of bronchus; lung | 165.1 | 0.04 | 0.02 | 0.05 | 0.02 | 0.01 | 0.02 |
| | Breast cancer | 174 | 0.02 | 0.01 | 0.02 | 0.01 | 0.00 | 0.01 |
| | Cervical cancer and dysplasia | 180 | 0.00 | 0.00 | 0.01 | 0.00 | 0.00 | 0.00 |
| | Malignant neoplasm of ovary | 184.11 | 0.02 | 0.00 | 0.04 | 0.00 | -0.01 | 0.01 |
| | Cancer of prostate | 185 | 0.06 | 0.04 | 0.08 | 0.01 | 0.01 | 0.02 |
| **Metabolic** | Type 2 diabetes | 250.2 | 0.06 | 0.06 | 0.07 | 0.02 | 0.01 | 0.02 |
| | Hypercholesterolemia | 272.11 | 0.02 | 0.02 | 0.03 | 0.01 | 0.01 | 0.01 |
| | Gout | 274.1 | 0.09 | 0.08 | 0.09 | 0.03 | 0.03 | 0.04 |
| | Obesity | 278.1 | 0.04 | 0.03 | 0.05 | 0.01 | 0.01 | 0.02 |
| **Mental Disorders** | Alzheimer's disease | 290.11 | 0.05 | 0.02 | 0.09 | 0.03 | 0.01 | 0.06 |
| | Schizophrenia | 295.1 | 0.06 | 0.04 | 0.08 | 0.02 | 0.01 | 0.03 |
| | Depression | 296.2 | 0.03 | 0.03 | 0.04 | 0.01 | 0.01 | 0.01 |
| | Generalized anxiety disorder | 300.11 | 0.01 | 0.01 | 0.01 | 0.00 | 0.00 | 0.00 |
| | Alcoholism | 317.1 | 0.02 | 0.01 | 0.03 | 0.01 | 0.00 | 0.01 |
| **Neurological** | Migraine | 340 | 0.02 | 0.01 | 0.02 | 0.00 | 0.00 | 0.01 |
| **Sense Organs** | Hearing loss | 389 | 0.01 | 0.00 | 0.01 | 0.00 | 0.00 | 0.00 |
| **Circulatory System** | Hypertension | 401 | 0.04 | 0.04 | 0.05 | 0.01 | 0.01 | 0.01 |
| | Ischemic Heart Disease | 411 | 0.01 | 0.01 | 0.01 | 0.00 | 0.00 | 0.00 |
| | Myocardial infarction | 411.2 | 0.06 | 0.04 | 0.07 | 0.01 | 0.01 | 0.02 |
| | Atrial fibrillation | 427.21 | 0.13 | 0.11 | 0.15 | 0.04 | 0.03 | 0.05 |
| | Heart failure NOS | 428.2 | 0.02 | 0.00 | 0.04 | 0.00 | 0.00 | 0.01 |
| | Intracranial hemorrhage | 430 | 0.03 | 0.02 | 0.05 | 0.02 | 0.01 | 0.03 |
| | Cerebrovascular disease | 433 | 0.01 | 0.01 | 0.01 | 0.00 | 0.00 | 0.00 |
| | Peripheral vascular disease | 443 | 0.00 | 0.00 | 0.01 | 0.00 | 0.00 | 0.00 |
| **Respiratory System** | Asthma | 495 | 0.04 | 0.04 | 0.05 | 0.02 | 0.02 | 0.02 |
| | Obstructive chronic bronchitis | 496.21 | 0.02 | 0.02 | 0.03 | 0.01 | 0.01 | 0.02 |
| **Digestive system** | GERD | 530.11 | 0.01 | 0.01 | 0.02 | 0.00 | 0.00 | 0.00 |
| | Chronic liver disease and cirrhosis | 571 | 0.01 | 0.01 | 0.02 | 0.00 | 0.00 | 0.00 |

| | | | | | | | | |
|---|---|---|---|---|---|---|---|---|
| **Genitourinary system** | Chronic renal failure [CKD] | 585.3 | 0.01 | 0.01 | 0.02 | 0.01 | 0.00 | 0.01 |
| **Musculoskeletal** | Osteoarthrosis | 740 | 0.01 | 0.01 | 0.01 | 0.00 | 0.00 | 0.00 |
| | Back pain | 760 | 0.01 | 0.00 | 0.01 | 0.00 | 0.00 | 0.00 |

**Table S11. Sensitivity analyses for DALYpop of significant diseases with available DALYs estimates (top 5% and top 1%)**

| | | | Top 5% | | | Top 1% | | |
|---|---|---|---|---|---|---|---|---|
| **Category** | **Disease** | **PheCode** | **DALYpop** | **95CI lower** | **95CI upper** | **DALYpop** | **95CI lower** | **95CI upper** |
| **Neoplasms** | Colorectal cancer | 153 | 22.97 | 10.60 | 35.48 | 8.50 | 2.46 | 15.03 |
| | Cancer of bronchus; lung | 165.1 | 39.65 | 25.69 | 54.35 | 18.58 | 10.49 | 26.28 |
| | Breast cancer | 174 | 7.21 | 4.36 | 9.78 | 2.52 | 1.30 | 3.75 |
| | Cervical cancer and dysplasia | 180 | 0.09 | -0.47 | 0.65 | 0.18 | -0.07 | 0.46 |
| | Malignant neoplasm of ovary | 184.11 | 2.21 | -0.41 | 4.43 | 0.13 | -0.66 | 1.11 |
| | Cancer of prostate | 185 | 11.91 | 8.28 | 16.29 | 2.77 | 0.99 | 4.75 |
| **Metabolic** | Type 2 diabetes | 250.2 | 113.25 | 105.73 | 120.46 | 29.82 | 26.39 | 33.77 |
| | Gout | 274.1 | 4.95 | 4.67 | 5.24 | 1.94 | 1.78 | 2.11 |
| **Mental Disorders** | Alzheimer's disease | 290.11 | 40.90 | 19.66 | 77.40 | 27.25 | 7.62 | 47.53 |
| | Schizophrenia | 295.1 | 15.58 | 10.58 | 21.79 | 4.68 | 2.12 | 7.75 |
| | Depression | 296.2 | 16.49 | 13.54 | 18.67 | 4.49 | 3.20 | 5.70 |
| | Generalized anxiety disorder | 300.11 | 4.51 | 2.74 | 6.49 | 1.18 | 0.33 | 2.02 |
| | Alcoholism | 317.1 | 4.52 | 1.81 | 7.31 | 1.45 | 0.25 | 2.83 |
| **Neurological** | Migraine | 340 | 7.91 | 5.94 | 10.12 | 2.12 | 1.16 | 3.16 |
| **Sense Organs** | Hearing loss | 389 | 4.96 | 2.72 | 7.70 | 1.66 | 0.60 | 2.83 |
| **Circulatory System** | Ischemic Heart Disease | 411 | 12.20 | 8.55 | 16.33 | 3.37 | 1.64 | 5.19 |
| | Atrial fibrillation | 427.21 | 22.95 | 20.34 | 26.96 | 7.62 | 5.90 | 9.55 |
| | Intracranial hemorrhage | 430 | 40.91 | 27.04 | 61.34 | 30.04 | 19.57 | 41.34 |
| **Respiratory System** | Asthma | 495 | 10.27 | 9.50 | 11.42 | 4.65 | 4.06 | 5.25 |
| | Obstructive chronic bronchitis | 496.21 | 14.86 | 10.36 | 21.89 | 6.87 | 3.58 | 10.01 |
| **Digestive system** | GERD | 530.11 | 0.63 | 0.46 | 0.81 | 0.14 | 0.06 | 0.22 |
| | Chronic liver disease and cirrhosis | 571 | 9.55 | 6.88 | 12.61 | 2.13 | 0.84 | 3.56 |
| **Musculoskeletal** | Osteoarthrosis | 740 | 4.54 | 3.68 | 5.59 | 1.36 | 0.91 | 1.81 |
| | Back pain | 760 | 6.94 | 4.36 | 9.15 | 1.01 | -0.03 | 2.03 |

**Table S12. Overall and sex-specific individual DALYs (DALYind) attributable to PRSe (top 10%) for significant diseases with available DALYs estimates**

| | | | All | | | Male | | | Female | | |
|---|---|---|---|---|---|---|---|---|---|---|---|
| **Category** | **Disease** | **PheCode** | **DALYind** | **95CI lower** | **95CI upper** | **DALYind** | **95CI lower** | **95CI upper** | **DALYind** | **95CI lower** | **95CI upper** |
| **Neoplasms** | Colorectal cancer | 153 | 0.27 | 0.15 | 0.41 | 0.46 | 0.23 | 0.74 | 0.14 | 0.03 | 0.28 |
| | Cancer of bronchus; lung | 165.1 | 0.51 | 0.36 | 0.67 | 0.21 | -0.07 | 0.49 | 0.53 | 0.38 | 0.70 |
| | Breast cancer | 174 | 0.11 | 0.08 | 0.14 | 0.00 | 0.00 | 0.01 | 0.22 | 0.17 | 0.28 |
| | Cervical cancer and dysplasia | 180 | 0.01 | 0.01 | 0.02 | NA | NA | NA | 0.04 | 0.02 | 0.06 |
| | Malignant neoplasm of ovary | 184.11 | 0.03 | 0.00 | 0.06 | NA | NA | NA | 0.05 | 0.01 | 0.10 |
| | Cancer of prostate | 185 | 0.17 | 0.12 | 0.22 | 0.33 | 0.23 | 0.42 | NA | NA | NA |
| **Metabolic** | Type 2 diabetes | 250.2 | 1.50 | 1.42 | 1.57 | 1.74 | 1.61 | 1.87 | 1.33 | 1.24 | 1.42 |
| | Gout | 274.1 | 0.06 | 0.05 | 0.06 | 0.12 | 0.12 | 0.13 | 0.01 | 0.01 | 0.01 |
| **Mental Disorders** | Alzheimer's disease | 290.11 | 0.50 | 0.26 | 0.82 | 0.17 | -0.10 | 0.50 | 0.88 | 0.36 | 1.48 |
| | Schizophrenia | 295.1 | 0.23 | 0.17 | 0.29 | 0.26 | 0.17 | 0.37 | 0.21 | 0.14 | 0.28 |
| | Depression | 296.2 | 0.20 | 0.18 | 0.23 | 0.13 | 0.10 | 0.17 | 0.27 | 0.23 | 0.32 |
| | Generalized anxiety disorder | 300.11 | 0.06 | 0.04 | 0.08 | 0.06 | 0.02 | 0.09 | 0.07 | 0.03 | 0.10 |
| | Alcoholism | 317.1 | 0.07 | 0.04 | 0.10 | 0.15 | 0.09 | 0.23 | 0.01 | 0.00 | 0.02 |
| **Neurological** | Migraine | 340 | 0.11 | 0.09 | 0.13 | 0.05 | 0.02 | 0.08 | 0.17 | 0.13 | 0.21 |
| **Sense Organs** | Hearing loss | 389 | 0.05 | 0.02 | 0.08 | 0.08 | 0.04 | 0.13 | 0.03 | 0.00 | 0.07 |
| **Circulatory System** | Ischemic Heart Disease | 411 | 0.15 | 0.11 | 0.20 | 0.27 | 0.18 | 0.36 | 0.08 | 0.04 | 0.12 |
| | Atrial fibrillation | 427.21 | 0.29 | 0.26 | 0.33 | 0.31 | 0.26 | 0.35 | 0.28 | 0.23 | 0.33 |
| | Intracranial hemorrhage | 430 | 0.57 | 0.40 | 0.76 | 1.04 | 0.71 | 1.47 | 0.27 | 0.09 | 0.45 |
| **Respiratory System** | Asthma | 495 | 0.12 | 0.11 | 0.13 | 0.10 | 0.08 | 0.11 | 0.14 | 0.13 | 0.16 |
| | Obstructive chronic bronchitis | 496.21 | 0.21 | 0.14 | 0.27 | 0.37 | 0.24 | 0.49 | 0.09 | 0.03 | 0.15 |

| | | | | | | | | | | | |
|---|---|---|---|---|---|---|---|---|---|---|---|
| **Digestive system** | GERD | 530.11 | 0.01 | 0.00 | 0.01 | 0.01 | 0.00 | 0.01 | 0.01 | 0.00 | 0.01 |
| | Chronic liver disease and cirrhosis | 571 | 0.12 | 0.08 | 0.15 | 0.18 | 0.12 | 0.26 | 0.05 | 0.03 | 0.07 |
| **Musculoskeletal** | Osteoarthrosis | 740 | 0.07 | 0.06 | 0.08 | 0.04 | 0.03 | 0.06 | 0.09 | 0.07 | 0.11 |
| | Back pain | 760 | 0.11 | 0.08 | 0.13 | 0.05 | 0.01 | 0.09 | 0.16 | 0.12 | 0.20 |

**Table S13. Sensitivity analyses for DALYind of significant diseases with available DALYs estimates (top 5% and top 1%)**

| | | | Top 5% | | | Top 1% | | |
|---|---|---|---|---|---|---|---|---|
| **Category** | **Disease** | **PheCode** | **DALYind** | **95CI lower** | **95CI upper** | **DALYind** | **95CI lower** | **95CI upper** |
| **Neoplasms** | Colorectal cancer | 153 | 0.37 | 0.17 | 0.57 | 0.68 | 0.20 | 1.21 |
| | Cancer of bronchus; lung | 165.1 | 0.64 | 0.41 | 0.87 | 1.49 | 0.84 | 2.11 |
| | Breast cancer | 174 | 0.12 | 0.07 | 0.16 | 0.20 | 0.10 | 0.30 |
| | Cervical cancer and dysplasia | 180 | 0.00 | -0.01 | 0.01 | 0.01 | -0.01 | 0.04 |
| | Malignant neoplasm of ovary | 184.11 | 0.04 | -0.01 | 0.07 | 0.01 | -0.05 | 0.09 |
| | Cancer of prostate | 185 | 0.19 | 0.13 | 0.26 | 0.22 | 0.08 | 0.38 |
| **Metabolic** | Type 2 diabetes | 250.2 | 1.82 | 1.70 | 1.93 | 2.39 | 2.12 | 2.71 |
| | Gout | 274.1 | 0.08 | 0.08 | 0.08 | 0.16 | 0.14 | 0.17 |
| **Mental Disorders** | Alzheimer's disease | 290.11 | 0.66 | 0.32 | 1.24 | 2.19 | 0.61 | 3.81 |
| | Schizophrenia | 295.1 | 0.25 | 0.17 | 0.35 | 0.38 | 0.17 | 0.62 |
| | Depression | 296.2 | 0.26 | 0.22 | 0.30 | 0.36 | 0.26 | 0.46 |
| | Generalized anxiety disorder | 300.11 | 0.07 | 0.04 | 0.10 | 0.09 | 0.03 | 0.16 |
| | Alcoholism | 317.1 | 0.07 | 0.03 | 0.12 | 0.12 | 0.02 | 0.23 |
| **Neurological** | Migraine | 340 | 0.13 | 0.10 | 0.16 | 0.17 | 0.09 | 0.25 |
| **Sense Organs** | Hearing loss | 389 | 0.08 | 0.04 | 0.12 | 0.13 | 0.05 | 0.23 |
| **Circulatory System** | Ischemic Heart Disease | 411 | 0.20 | 0.14 | 0.26 | 0.27 | 0.13 | 0.42 |
| | Atrial fibrillation | 427.21 | 0.37 | 0.33 | 0.43 | 0.61 | 0.47 | 0.77 |
| | Intracranial hemorrhage | 430 | 0.66 | 0.43 | 0.98 | 2.41 | 1.57 | 3.32 |
| **Respiratory System** | Asthma | 495 | 0.16 | 0.15 | 0.18 | 0.37 | 0.33 | 0.42 |
| | Obstructive chronic bronchitis | 496.21 | 0.24 | 0.17 | 0.35 | 0.55 | 0.29 | 0.80 |
| **Digestive system** | GERD | 530.11 | 0.01 | 0.01 | 0.01 | 0.01 | 0.00 | 0.02 |
| | Chronic liver disease and cirrhosis | 571 | 0.15 | 0.11 | 0.20 | 0.17 | 0.07 | 0.29 |
| **Musculoskeletal** | Osteoarthrosis | 740 | 0.07 | 0.06 | 0.09 | 0.11 | 0.07 | 0.14 |
| | Back pain | 760 | 0.11 | 0.07 | 0.15 | 0.08 | 0.00 | 0.16 |

#### Table S14. Kappa statistics for overlap between PheCode and self-reported (SR) definitions

| | PheCode | | | | | | | | | | | |
|---|---|---|---|---|---|---|---|---|---|---|---|---|
| **Disease** | **Cases** | **Controls** | **Prevalence** | **HR** | **95CI lower** | **95CI upper** | **PAF** | **95CI lower** | **95CI upper** | **indDALYs** | **95CI lower** | **95CI upper** |
| **Breast cancer** | 6,803 | 103,905 | 4.77% | 1.34 | 1.25 | 1.44 | 0.03 | 0.02 | 0.04 | 0.12 | 0.07 | 0.16 |
| **Type 2 diabetes** | 33,364 | 99,746 | 23.42% | 2.13 | 2.07 | 2.19 | 0.10 | 0.10 | 0.11 | 1.82 | 1.70 | 1.93 |
| **Hypercholesterolemia** | 19,388 | 68,903 | 13.61% | 1.46 | 1.41 | 1.53 | 0.04 | 0.04 | 0.05 | | | |
| **Gout** | 25,756 | 116,386 | 18.08% | 2.41 | 2.33 | 2.49 | 0.12 | 0.12 | 0.13 | 0.08 | 0.08 | 0.08 |
| **Depression** | 12,526 | 64,000 | 8.79% | 1.55 | 1.47 | 1.63 | 0.05 | 0.04 | 0.06 | 0.26 | 0.22 | 0.30 |
| **Hypertension** | 46,783 | 89,368 | 32.83% | 1.81 | 1.76 | 1.86 | 0.07 | 0.07 | 0.08 | | | |
| **Ischemic Heart Disease** | 32,049 | 108,118 | 22.49% | 1.14 | 1.10 | 1.18 | 0.01 | 0.01 | 0.02 | 0.20 | 0.14 | 0.26 |
| **Atrial fibrillation** | 2,064 | 98,731 | 1.45% | 3.56 | 3.23 | 3.92 | 0.20 | 0.18 | 0.23 | 0.37 | 0.33 | 0.43 |
| **Intracranial hemorrhage** | 2,069 | 123,283 | 1.45% | 1.56 | 1.38 | 1.76 | 0.05 | 0.04 | 0.07 | 0.66 | 0.43 | 0.98 |
| **Asthma** | 29,028 | 80,366 | 20.37% | 1.64 | 1.58 | 1.69 | 0.06 | 0.05 | 0.06 | 0.16 | 0.15 | 0.18 |

| | Self-reported | | | | | | | | | | | | | | |
|---|---|---|---|---|---|---|---|---|---|---|---|---|---|---|---|
| **Disease** | **Cases** | **Controls** | **Missing** | **Available age of diagnosis** | **Prevalence** | **HR** | **95CI lower** | **95CI upper** | **PAF** | **95CI lower** | **95CI upper** | **indDALYs** | **95CI lower** | **95CI upper** | **Kappa** |
| **Breast cancer** | 415 | 140,326 | 2,345 | 408 | 0.29% | 2.46 | 1.95 | 3.12 | 0.13 | 0.09 | 0.17 | 0.42 | 0.29 | 0.57 | 0.1079 |
| **Type 2 diabetes** | 8,580 | 133,005 | 1,501 | 6,760 | 6.00% | 3.45 | 3.26 | 3.64 | 0.20 | 0.18 | 0.21 | 2.90 | 2.72 | 3.09 | 0.3404 |
| **Hypercholesterolemia** | 13,936 | 126,636 | 2,514 | 9,534 | 9.74% | 1.52 | 1.43 | 1.61 | 0.05 | 0.04 | 0.06 | | | | 0.3210 |
| **Gout** | 6,020 | 134,779 | 2,287 | 4,210 | 4.21% | 10.72 | 10.08 | 11.39 | 0.49 | 0.48 | 0.51 | 0.22 | 0.21 | 0.23 | 0.2986 |
| **Depression** | 5,912 | 134,842 | 2,332 | 4,570 | 4.13% | 0.99 | 0.90 | 1.09 | | | | | | | 0.4162 |
| **Hypertension** | 20,008 | 120,544 | 2,534 | 14,582 | 13.98% | 2.22 | 2.13 | 2.32 | 0.11 | 0.10 | 0.12 | | | | 0.4614 |
| **Ischemic Heart Disease** | 2,430 | 138,129 | 2,332 | 1,997 | 1.70% | 1.52 | 1.34 | 1.72 | 0.05 | 0.03 | 0.07 | 0.56 | 0.36 | 0.75 | 0.1041 |
| **Atrial fibrillation** | 7,578 | 132,980 | 2,528 | 4,986 | 5.30% | 1.45 | 1.34 | 1.58 | 0.04 | 0.03 | 0.05 | 0.06 | 0.05 | 0.08 | 0.3513 |
| **Intracranial hemorrhage** | 1,205 | 139,345 | 2,536 | 1,019 | 0.84% | 5.88 | 5.18 | 6.67 | 0.33 | 0.29 | 0.36 | 3.56 | 3.20 | 3.91 | 0.2325 |
| **Asthma** | 5,761 | 135,275 | 2,050 | 3,276 | 4.03% | 6.46 | 6.03 | 6.93 | 0.35 | 0.34 | 0.37 | 0.71 | 0.68 | 0.75 | 0.1924 |

**Table S15. Overall and sex-specific comparison of PRSe and MRFs for significant diseases with available DALYs estimates**

| | | | | DALYpop | | | PAF | | |
|---|---|---|---|---|---|---|---|---|---|
| **Category** | **Disease** | **PheCode** | **Risk factor** | **All** | **Male** | **Female** | **All** | **Male** | **Female** |
| **Neoplasms** | Colorectal cancer | 153 | Tobacco | 40.86 | 79.35 | 3.28 | 0.04 | 0.06 | 0.00 |
| | | | High alcohol use | 36.72 | 70.49 | 3.74 | 0.04 | 0.05 | 0.00 |
| | | | High fasting plasma glucose | 122.91 | 158.00 | 88.66 | 0.12 | 0.12 | 0.12 |
| | | | High body-mass index | 89.98 | 113.79 | 66.74 | 0.09 | 0.08 | 0.09 |
| | | | Dietary risks | 380.85 | 469.25 | 294.54 | 0.36 | 0.35 | 0.39 |
| | | | Low physical activity | 71.20 | 78.43 | 64.14 | 0.07 | 0.06 | 0.09 |
| | | | PRS top 10% (PRSe) | 33.32 | 59.57 | 17.27 | 0.03 | 0.04 | 0.02 |
| | Cancer of bronchus; lung | 165.1 | Air pollution | 126.81 | 158.24 | 96.13 | 0.11 | 0.11 | 0.12 |
| | | | Other environmental risks | 11.93 | 14.96 | 8.96 | 0.01 | 0.01 | 0.01 |
| | | | Tobacco | 617.60 | 1052.20 | 193.28 | 0.56 | 0.75 | 0.23 |
| | | | High fasting plasma glucose | 65.35 | 81.90 | 49.20 | 0.06 | 0.06 | 0.06 |
| | | | Dietary risks | 26.93 | 35.74 | 18.32 | 0.02 | 0.03 | 0.02 |
| | | | Occupational risks | 117.09 | 175.40 | 60.15 | 0.11 | 0.13 | 0.07 |
| | | | PRS top 10% (PRSe) | 64.06 | 27.83 | 63.21 | 0.06 | 0.02 | 0.08 |
| | Breast cancer | 174 | Tobacco | 40.20 | 0.49 | 78.98 | 0.10 | 0.05 | 0.10 |
| | | | High alcohol use | 3.37 | 0.53 | 6.14 | 0.01 | 0.06 | 0.01 |
| | | | High fasting plasma glucose | 36.59 | NA | 72.32 | 0.09 | 0.00 | 0.09 |
| | | | High body-mass index | 27.91 | NA | 55.17 | 0.07 | 0.00 | 0.07 |
| | | | Dietary risks | 55.72 | 1.24 | 108.92 | 0.13 | 0.13 | 0.13 |
| | | | Low physical activity | 11.02 | NA | 21.78 | 0.03 | 0.00 | 0.03 |
| | | | PRS top 10% (PRSe) | 13.54 | 0.39 | 26.83 | 0.03 | 0.04 | 0.03 |
| | Cervical cancer and dysplasia | 180 | Tobacco | 3.78 | NA | 7.47 | 0.04 | NA | 0.02 |
| | | | Unsafe sex | 101.34 | NA | 200.28 | 1.00 | NA | 0.61 |
| | | | PRS top 10% (PRSe) | 1.49 | NA | 4.85 | 0.01 | NA | 0.01 |
| | Malignant neoplasm of ovary | | High body-mass index | 7.80 | NA | 15.41 | 0.08 | NA | 0.10 |
| | | | Occupational risks | 0.75 | NA | 1.47 | 0.01 | NA | 0.01 |
| | | | PRS top 10% (PRSe) | 3.64 | NA | 5.93 | 0.04 | NA | 0.04 |
| | Cancer of prostate | 185 | Tobacco | 5.56 | 11.26 | | 0.03 | 0.03 | NA |
| | | | High alcohol use | 1.89 | 3.82 | | 0.01 | 0.01 | NA |

| | | | | | | | | | |
|---|---|---|---|---|---|---|---|---|---|
| | | | Dietary risks | -4.13 | -8.36 | | -0.02 | -0.02 | NA |
| | | | PRS top 10% (PRSe) | 21.06 | 42.88 | NA | 0.11 | 0.11 | NA |
| **Metabolic** | Type 2 diabetes | 250.2 | Low physical activity | 179.55 | 169.57 | 189.28 | 0.10 | 0.08 | 0.11 |
| | | | Non-optimal temperature | 46.34 | 54.43 | 38.45 | 0.03 | 0.03 | 0.02 |
| | | | Sexual violence against children and bullying | 33.66 | 36.91 | 30.49 | 0.02 | 0.02 | 0.02 |
| | | | Air pollution | 269.66 | 295.46 | 244.47 | 0.15 | 0.15 | 0.15 |
| | | | Tobacco | 220.57 | 321.03 | 122.49 | 0.12 | 0.16 | 0.07 |
| | | | High alcohol use | 3.20 | 7.09 | -0.60 | 0.00 | 0.00 | 0.00 |
| | | | High fasting plasma glucose | 1836.35 | 2016.03 | 1660.92 | 1.00 | 1.00 | 1.00 |
| | | | High body-mass index | 949.52 | 1056.26 | 845.32 | 0.52 | 0.52 | 0.51 |
| | | | Dietary risks | 479.26 | 567.94 | 392.67 | 0.26 | 0.28 | 0.24 |
| | | | PRS top 10% (PRSe) | 186.44 | 226.34 | 159.01 | 0.10 | 0.11 | 0.10 |
| | Gout | 274.1 | High body-mass index | 19.69 | 28.81 | 10.79 | 0.35 | 0.35 | 0.36 |
| | | | Kidney dysfunction | 13.65 | 19.18 | 8.25 | 0.25 | 0.23 | 0.28 |
| | | | PRS top 10% (PRSe) | 6.86 | 16.16 | 1.08 | 0.12 | 0.20 | 0.04 |
| **Mental Disorders** | Alzheimer's disease | 290.11 | High fasting plasma glucose | 174.69 | 117.99 | 230.04 | 0.21 | 0.21 | 0.21 |
| | | | High body-mass index | 25.43 | 14.73 | 35.88 | 0.03 | 0.03 | 0.03 |
| | | | Air pollution | 230.43 | 154.80 | 304.27 | 0.28 | 0.28 | 0.28 |
| | | | Tobacco | 20.93 | 36.57 | 5.66 | 0.03 | 0.07 | 0.01 |
| | | | High alcohol use | -8.51 | -15.41 | -1.77 | -0.01 | -0.03 | 0.00 |
| | | | PRS top 10% (PRSe) | 62.85 | 22.33 | 104.61 | 0.08 | 0.04 | 0.10 |
| | Schizophrenia | 295.1 | Sexual violence against children and bullying | 62.98 | 69.43 | 56.69 | 0.22 | 0.22 | 0.23 |
| | | | PRS top 10% (PRSe) | 28.92 | 33.62 | 24.55 | 0.10 | 0.11 | 0.10 |
| | Depression | 296.2 | Intimate partner violence | 35.93 | | 71.01 | 0.07 | 0.00 | 0.11 |
| | | | Sexual violence against children and bullying | 41.70 | 38.77 | 44.56 | 0.09 | 0.11 | 0.07 |
| | | | PRS top 10% (PRSe) | 25.28 | 17.32 | 32.82 | 0.05 | 0.05 | 0.05 |
| | Generalized anxiety disorder | 300.11 | Intimate partner violence | 48.65 | | 96.15 | 0.11 | 0.00 | 0.17 |
| | | | Sexual violence against children and bullying | 56.50 | 53.41 | 59.53 | 0.12 | 0.16 | 0.11 |
| | | | PRS top 10% (PRSe) | 7.15 | 7.48 | 8.06 | 0.02 | 0.02 | 0.01 |
| | Alcoholism | 317.1 | High alcohol use | 260.60 | 471.38 | 54.80 | 1.00 | 1.00 | 1.00 |
| | | | Sexual violence against children and bullying | 23.54 | 42.56 | 4.98 | 0.09 | 0.09 | 0.09 |

| | | | | | | | | | |
|---|---|---|---|---|---|---|---|---|---|
| | | | PRS top 10% (PRSe) | 8.59 | 19.40 | 0.86 | 0.03 | 0.04 | 0.02 |
| **Sense Organs** | Hearing loss | 389 | Occupational risks | 148.32 | 177.07 | 120.25 | 0.16 | 0.20 | 0.13 |
| | | | PRS top 10% (PRSe) | 6.38 | 10.82 | 4.02 | 0.01 | 0.01 | 0.00 |
| **Circulatory System** | Ischemic Heart Disease | 411 | Air pollution | 215.96 | 293.64 | 140.11 | 0.15 | 0.15 | 0.16 |
| | | | Other environmental risks | 369.23 | 508.76 | 233.00 | 0.26 | 0.27 | 0.26 |
| | | | Tobacco | 356.90 | 614.63 | 105.27 | 0.25 | 0.32 | 0.12 |
| | | | High alcohol use | -27.48 | -54.10 | -1.49 | -0.02 | -0.03 | 0.00 |
| | | | High fasting plasma glucose | 213.52 | 281.60 | 147.05 | 0.15 | 0.15 | 0.16 |
| | | | High systolic blood pressure | 659.35 | 876.36 | 447.46 | 0.47 | 0.46 | 0.50 |
| | | | High body-mass index | 158.05 | 227.27 | 90.46 | 0.11 | 0.12 | 0.10 |
| | | | Dietary risks | 596.30 | 844.76 | 353.71 | 0.42 | 0.44 | 0.39 |
| | | | Low physical activity | 28.44 | 29.94 | 26.98 | 0.02 | 0.02 | 0.03 |
| | | | Non-optimal temperature | 59.07 | 83.08 | 35.63 | 0.04 | 0.04 | 0.04 |
| | | | Kidney dysfunction | 260.53 | 337.36 | 185.53 | 0.19 | 0.18 | 0.21 |
| | | | High LDL cholesterol | 536.47 | 756.88 | 321.28 | 0.38 | 0.39 | 0.36 |
| | | | PRS top 10% (PRSe) | 18.98 | 35.27 | 9.10 | 0.01 | 0.02 | 0.01 |
| | Atrial fibrillation | 427.21 | Other environmental risks | 3.19 | 3.49 | 2.90 | 0.02 | 0.02 | 0.02 |
| | | | Tobacco | 6.36 | 11.88 | 0.98 | 0.04 | 0.07 | 0.01 |
| | | | High alcohol use | 2.90 | 5.51 | 0.34 | 0.02 | 0.03 | 0.00 |
| | | | High systolic blood pressure | 50.32 | 47.95 | 52.64 | 0.28 | 0.27 | 0.29 |
| | | | High body-mass index | 7.34 | 6.65 | 8.01 | 0.04 | 0.04 | 0.04 |
| | | | Dietary risks | 7.71 | 7.56 | 7.85 | 0.04 | 0.04 | 0.04 |
| | | | PRS top 10% (PRSe) | 36.53 | 39.67 | 33.27 | 0.20 | 0.22 | 0.18 |
| | Intracranial hemorrhage | 430 | Tobacco | 247.67 | 418.14 | 81.22 | 0.18 | 0.24 | 0.08 |
| | | | High alcohol use | 29.12 | 57.16 | 1.75 | 0.02 | 0.03 | 0.00 |
| | | | Kidney dysfunction | 151.16 | 198.18 | 105.26 | 0.11 | 0.11 | 0.11 |
| | | | High LDL cholesterol | 190.13 | 222.06 | 158.97 | 0.14 | 0.13 | 0.16 |
| | | | High fasting plasma glucose | 165.95 | 207.29 | 125.58 | 0.12 | 0.12 | 0.13 |
| | | | High systolic blood pressure | 717.73 | 918.00 | 522.20 | 0.53 | 0.53 | 0.53 |
| | | | High body-mass index | 61.67 | 79.13 | 44.62 | 0.05 | 0.05 | 0.05 |
| | | | Dietary risks | 148.67 | 179.38 | 118.69 | 0.11 | 0.10 | 0.12 |
| | | | Low physical activity | 31.03 | 35.85 | 26.32 | 0.02 | 0.02 | 0.03 |
| | | | Non-optimal temperature | 41.43 | 55.94 | 27.26 | 0.03 | 0.03 | 0.03 |

| | | | | | | | | | |
|---|---|---|---|---|---|---|---|---|---|
| | | | Air pollution | 178.93 | 227.54 | 131.47 | 0.13 | 0.13 | 0.13 |
| | | | Other environmental risks | 53.61 | 73.42 | 34.27 | 0.04 | 0.04 | 0.03 |
| | | | PRS top 10% (PRSe) | 71.56 | 135.53 | 32.61 | 0.05 | 0.08 | 0.03 |
| **Respiratory System** | Asthma | 495 | Sexual violence against children and bullying | 6.14 | 4.81 | 7.43 | 0.02 | 0.02 | 0.03 |
| | | | Air pollution | 4.83 | 5.47 | 4.21 | 0.02 | 0.02 | 0.01 |
| | | | Tobacco | 32.51 | 36.13 | 28.98 | 0.13 | 0.16 | 0.10 |
| | | | High body-mass index | 48.60 | 40.51 | 56.50 | 0.19 | 0.18 | 0.20 |
| | | | Occupational risks | 20.59 | 20.31 | 20.86 | 0.08 | 0.09 | 0.07 |
| | | | PRS top 10% (PRSe) | 15.04 | 12.60 | 17.07 | 0.06 | 0.06 | 0.06 |
| | Obstructive chronic bronchitis | 496.21 | Air pollution | 119.57 | 169.04 | 71.27 | 0.19 | 0.19 | 0.19 |
| | | | Tobacco | 287.89 | 500.67 | 80.15 | 0.45 | 0.56 | 0.21 |
| | | | Occupational risks | 40.69 | 69.41 | 12.65 | 0.06 | 0.08 | 0.03 |
| | | | Non-optimal temperature | 28.45 | 41.89 | 15.33 | 0.04 | 0.05 | 0.04 |
| | | | PRS top 10% (PRSe) | 26.00 | 48.18 | 10.84 | 0.04 | 0.05 | 0.03 |
| **Digestive system** | Chronic liver disease and cirrhosis | 571 | High alcohol use | 191.54 | 363.85 | 23.31 | 0.26 | 0.32 | 0.07 |
| | | | Drug use | 79.40 | 145.32 | 15.03 | 0.11 | 0.13 | 0.05 |
| | | | PRS top 10% (PRSe) | 14.48 | 23.80 | 6.39 | 0.02 | 0.02 | 0.02 |
| **Musculoskeletal** | Osteoarthrosis | 740 | High body-mass index | 113.24 | 77.86 | 147.77 | 0.23 | 0.20 | 0.24 |
| | | | PRS top 10% (PRSe) | 8.28 | 5.50 | 11.00 | 0.02 | 0.01 | 0.02 |
| | Back pain | 760 | Tobacco | 174.17 | 310.32 | 41.23 | 0.13 | 0.26 | 0.03 |
| | | | High body-mass index | 154.31 | 127.77 | 180.21 | 0.11 | 0.11 | 0.12 |
| | | | Occupational risks | 197.77 | 209.03 | 186.79 | 0.14 | 0.17 | 0.12 |
| | | | PRS top 10% (PRSe) | 13.60 | 6.64 | 18.82 | 0.01 | 0.01 | 0.01 |

#### Table S16. Overall and sex-specific PAF of PRSe, joint PAF of MRFs, and joint PAF of all risk factors

| | | | All | | | Male | | | Female | | |
|---|---|---|---|---|---|---|---|---|---|---|---|
| Category | Disease | PheCode | PAF PRS | Joint PAF MRFs | Joint PAF | PAF PRS | Joint PAF MRFs | Joint PAF | PAF PRS | Joint PAF MRFs | Joint PAF |
| **Neoplasms** | Colorectal cancer | 153 | 0.03 | 0.56 | 0.57 | 0.04 | 0.56 | 0.58 | 0.02 | 0.56 | 0.57 |
| | Cancer of bronchus; lung | 165.1 | 0.06 | 0.68 | 0.70 | 0.02 | 0.83 | 0.83 | 0.08 | 0.43 | 0.47 |
| | Breast cancer | 174 | 0.03 | 0.36 | 0.38 | 0.04 | 0.23 | 0.26 | 0.03 | 0.36 | 0.38 |
| | Cervical cancer and dysplasia | 180 | 0.01 | 1.00 | 1.00 | NA | NA | NA | 0.01 | 0.62 | 0.62 |
| | Malignant neoplasm of ovary | 184.11 | 0.04 | 0.08 | 0.12 | NA | NA | NA | 0.04 | 0.10 | 0.14 |
| | Cancer of prostate | 185 | 0.11 | 0.02 | 0.13 | 0.11 | 0.02 | 0.13 | NA | NA | NA |
| **Metabolic** | Type 2 diabetes | 250.2 | 0.10 | 1.00 | 1.00 | 0.11 | 1.00 | 1.00 | 0.10 | 1.00 | 1.00 |
| | Gout | 274.1 | 0.12 | 0.51 | 0.57 | 0.20 | 0.50 | 0.60 | 0.04 | 0.54 | 0.55 |
| **Mental Disorders** | Alzheimer's disease | 290.11 | 0.08 | 0.46 | 0.50 | 0.04 | 0.47 | 0.49 | 0.10 | 0.46 | 0.51 |
| | Schizophrenia | 295.1 | 0.10 | 0.22 | 0.30 | 0.11 | 0.22 | 0.31 | 0.10 | 0.23 | 0.30 |
| | Depression | 296.2 | 0.05 | 0.15 | 0.20 | 0.05 | 0.11 | 0.16 | 0.05 | 0.18 | 0.22 |
| | Generalized anxiety disorder | 300.11 | 0.02 | 0.22 | 0.23 | 0.02 | 0.16 | 0.18 | 0.01 | 0.26 | 0.27 |
| | Alcoholism | 317.1 | 0.03 | 1.00 | 1.00 | 0.04 | 1.00 | 1.00 | 0.02 | 1.00 | 1.00 |
| **Sense Organs** | Hearing loss | 389 | 0.01 | 0.16 | 0.17 | 0.01 | 0.20 | 0.21 | 0.00 | 0.13 | 0.13 |
| **Circulatory System** | Ischemic Heart Disease | 411 | 0.01 | 0.95 | 0.95 | 0.02 | 0.95 | 0.95 | 0.01 | 0.94 | 0.94 |
| | Atrial fibrillation | 427.21 | 0.20 | 0.38 | 0.51 | 0.22 | 0.41 | 0.54 | 0.18 | 0.36 | 0.48 |
| | Intracranial hemorrhage | 430 | 0.05 | 0.83 | 0.84 | 0.08 | 0.84 | 0.85 | 0.03 | 0.81 | 0.82 |
| **Respiratory System** | Asthma | 495 | 0.06 | 0.38 | 0.42 | 0.06 | 0.41 | 0.44 | 0.06 | 0.36 | 0.40 |
| | Obstructive chronic bronchitis | 496.21 | 0.04 | 0.60 | 0.62 | 0.05 | 0.68 | 0.70 | 0.03 | 0.40 | 0.42 |
| **Digestive system** | Chronic liver disease and cirrhosis | 571 | 0.02 | 0.34 | 0.36 | 0.02 | 0.40 | 0.42 | 0.02 | 0.12 | 0.13 |
| **Musculoskeletal** | Osteoarthrosis | 740 | 0.02 | 0.23 | 0.24 | 0.01 | 0.20 | 0.21 | 0.02 | 0.24 | 0.26 |
| | Back pain | 760 | 0.01 | 0.34 | 0.34 | 0.01 | 0.45 | 0.46 | 0.01 | 0.25 | 0.26 |

**Table S17. Trained mixing weights of integrated PRS for healthcare utilization indices**

| PRS name | Hospitilization days | Inpatient visits | ER visits | Outpatient visits | Total visits |
|---|---|---|---|---|---|
| **LC_EAS_Nam_2022_prscs** | 0.00E+00 | 0.00E+00 | 0.00E+00 | 0.00E+00 | 0.00E+00 |
| **LC_EUR_Jiang_2021_prscs** | 0.00E+00 | 1.02E-05 | 1.79E-06 | 0.00E+00 | 0.00E+00 |
| **LC_EUR_McKay_2017_prscs** | 0.00E+00 | 0.00E+00 | 0.00E+00 | 0.00E+00 | 0.00E+00 |
| **LC_MULTI_Sakaue_2021_prscs** | 0.00E+00 | 3.63E-03 | 0.00E+00 | 0.00E+00 | 0.00E+00 |
| **CRC_EAS_Nam_2022_prscs** | -2.39E+00 | 3.17E-02 | 0.00E+00 | 0.00E+00 | 0.00E+00 |
| **CRC_EUR_Jiang_2021_prscs** | 0.00E+00 | -6.67E-06 | 3.58E-05 | 2.73E-04 | 1.29E-04 |
| **SC_EUR_Jiang_2021_prscs** | 0.00E+00 | 0.00E+00 | -6.34E-06 | -1.10E-02 | -7.42E-03 |
| **SC_MULTI_Sakaue_2021_prscs** | 0.00E+00 | 0.00E+00 | 0.00E+00 | 5.63E+00 | 4.51E+00 |
| **BRC_EAS_Nam_2022_prscs** | 1.60E-01 | 0.00E+00 | -7.70E-02 | 2.17E+00 | 9.54E-01 |
| **BRC_EUR_Michailidou_2017_prscs** | 1.66E-01 | 0.00E+00 | 0.00E+00 | 4.53E-01 | 0.00E+00 |
| **BRC_FINN_2024_prscs** | 3.80E+00 | 5.51E-02 | 4.48E-03 | -1.25E+00 | 0.00E+00 |
| **OC_EAS_Sakaue_2021_prscs** | 0.00E+00 | 0.00E+00 | 0.00E+00 | -1.02E+00 | -3.29E-01 |
| **OC_EUR_Eileen_2024_prscs** | 0.00E+00 | 0.00E+00 | 0.00E+00 | 0.00E+00 | 0.00E+00 |
| **CC_EUR_Rashkin_2020_prscs** | 0.00E+00 | 3.37E-02 | 5.96E-02 | 4.15E+00 | 3.72E+00 |
| **CC_MULTI_Sakaue_2021_prscs** | 0.00E+00 | 3.35E-02 | 2.30E-01 | 5.87E+00 | 5.28E+00 |
| **CC_EUR_Zhou_2018_prscs** | 0.00E+00 | 1.40E-01 | 1.53E-01 | 3.28E+00 | 2.71E+00 |
| **PC_EUR_Schumacher_2018_prscs** | 9.13E-02 | 8.96E-02 | 0.00E+00 | 0.00E+00 | 0.00E+00 |
| **PC_FINN_2024_prscs** | 0.00E+00 | 4.27E-02 | 7.12E-02 | 0.00E+00 | 0.00E+00 |
| **SKC_EAS_Sakaue_2021_prscs** | 0.00E+00 | -6.28E-04 | -2.43E-02 | 0.00E+00 | 0.00E+00 |
| **SKC_EUR_Jiang_2021_prscs** | 0.00E+00 | 0.00E+00 | 0.00E+00 | -5.32E-20 | -4.03E-20 |
| **EC_EAS_Sakaue_2021_prscs** | 0.00E+00 | -1.53E-02 | 2.18E-01 | 6.86E+00 | 4.94E+00 |
| **EC_EUR_Sakaue_2021_prscs** | 0.00E+00 | 5.84E-04 | 0.00E+00 | 4.56E+00 | 2.88E+00 |
| **BLC_EUR_Jiang_2021_prscs** | 9.52E+00 | 1.74E-01 | 1.09E-01 | 0.00E+00 | 0.00E+00 |
| **CAD_EAS_Kanai_2021_prscs** | 0.00E+00 | 0.00E+00 | -1.36E-02 | 0.00E+00 | 0.00E+00 |
| **CAD_EUR_Zhou_2018_prscs** | 0.00E+00 | 3.03E-01 | 0.00E+00 | 1.54E+01 | 1.36E+01 |
| **CAD_MULTI_Aragam_2022_prscs** | 0.00E+00 | 0.00E+00 | 0.00E+00 | 0.00E+00 | 0.00E+00 |
| **CAD_Zhou_Kanai_meta_prscsx** | 0.00E+00 | 0.00E+00 | -1.72E-01 | 0.00E+00 | 0.00E+00 |
| **HT_EAS_Nam_2022_prscs** | 0.00E+00 | 0.00E+00 | 0.00E+00 | 1.17E+01 | 1.14E+01 |
| **HT_MULTI_Verma_2024_prscs** | 0.00E+00 | 1.82E-01 | 7.07E-01 | 3.91E+01 | 4.02E+01 |
| **MI_EAS_Sakaue_2021_prscs** | 0.00E+00 | 0.00E+00 | 0.00E+00 | 0.00E+00 | 0.00E+00 |
| **MI_EUR_Jiang_2021_prscs** | -7.38E-35 | 0.00E+00 | 0.00E+00 | -1.20E-33 | -8.48E-34 |
| **MI_EUR_Zhou_2018_prscs** | 0.00E+00 | 0.00E+00 | 0.00E+00 | -1.37E+01 | -1.02E+01 |
| **MI_FINN_2024_prscs** | 0.00E+00 | 0.00E+00 | 0.00E+00 | 2.75E-01 | 0.00E+00 |
| **MI_MULTI_Hartiala_2021_prscs** | 0.00E+00 | 0.00E+00 | 2.36E-01 | 0.00E+00 | 0.00E+00 |
| **MI_Zhou_Sakaue_meta_prscsx** | 0.00E+00 | 0.00E+00 | 0.00E+00 | 0.00E+00 | 0.00E+00 |
| **SAH_EAS_Sakaue_2021_prscs** | 0.00E+00 | 0.00E+00 | 1.17E-01 | 0.00E+00 | 0.00E+00 |

| | | | | | |
|---|---|---|---|---|---|
| **SAH_EUR_Jiang_2021_prscs** | 0.00E+00 | -4.01E-05 | 0.00E+00 | -1.20E-02 | -9.12E-03 |
| **SAH_MULTI_Bakker_2020_prscs** | 0.00E+00 | 0.00E+00 | 2.27E+00 | 1.24E+01 | 1.17E+01 |
| **SAH_Jiang_Sakaue_meta_prscsx** | 0.00E+00 | 1.29E-03 | 0.00E+00 | 3.15E-03 | 0.00E+00 |
| **CVD_EUR_Jiang_2021_prscs** | 0.00E+00 | 0.00E+00 | 8.31E-02 | 1.14E+01 | 1.02E+01 |
| **CVD_MULTI_Verma_2024_prscs** | 0.00E+00 | 5.58E-01 | 1.04E+00 | 1.67E+01 | 1.81E+01 |
| **PAD_MULTI_Sakaue_2021_prscs** | 0.00E+00 | 0.00E+00 | 0.00E+00 | 2.54E+00 | 1.09E+00 |
| **AF_EUR_Nielsen_2018_prscs** | 0.00E+00 | 3.66E-02 | 0.00E+00 | 0.00E+00 | 0.00E+00 |
| **AF_MULTI_Miyazawa_2023_prscs** | 0.00E+00 | 0.00E+00 | 0.00E+00 | -1.13E+00 | -2.65E-01 |
| **HF_MULTI_Levin_2022_prscs** | 0.00E+00 | 1.67E-01 | 1.84E-02 | -1.73E+00 | 0.00E+00 |
| **DM2_EAS_Nam_2022_prscs** | 0.00E+00 | 0.00E+00 | 3.73E-02 | 0.00E+00 | 0.00E+00 |
| **DM2_EUR_Cordell_2021_prscs** | 0.00E+00 | 0.00E+00 | -6.32E-07 | 0.00E+00 | 0.00E+00 |
| **DM2_MULTI_Suzuki_2024_prscs** | 2.07E+00 | 1.61E-01 | 6.54E-02 | 1.29E+01 | 1.30E+01 |
| **DM2_Ge_meta_prscsx** | 0.00E+00 | 0.00E+00 | 0.00E+00 | 5.69E+00 | 5.50E+00 |
| **OB_FINN_2024_prscs** | 1.55E+00 | 4.00E-01 | 5.27E-01 | 3.32E+00 | 3.84E+00 |
| **COPD_EAS_Zhou_2022_prscs** | 0.00E+00 | 3.07E-01 | 6.16E-01 | 2.67E+01 | 2.76E+01 |
| **COPD_EUR_Zhou_2022_prscs** | 0.00E+00 | 3.17E-01 | 1.60E-01 | 1.98E+01 | 2.08E+01 |
| **COPD_Zhou_Zhou_meta_prscsx** | 0.00E+00 | 3.03E-01 | 1.78E+00 | 0.00E+00 | 0.00E+00 |
| **AS_EAS_Zhou_2022_prscs** | 7.42E-01 | 5.64E-01 | 8.61E-01 | 3.11E+01 | 3.17E+01 |
| **AS_EUR_Zhou_2022_prscs** | 0.00E+00 | 1.26E-01 | 1.28E+00 | 4.55E+01 | 4.67E+01 |
| **AS_Zhou_Zhou_meta_prscsx** | 1.91E+00 | 0.00E+00 | 0.00E+00 | 0.00E+00 | 0.00E+00 |
| **MG_EUR_Meyers_2023_prscs** | 0.00E+00 | 0.00E+00 | 0.00E+00 | 3.72E+00 | 2.19E+00 |
| **MG_FINN_2024_prscs** | 0.00E+00 | 0.00E+00 | 1.12E+00 | 2.54E+01 | 2.59E+01 |
| **AD_EUR_Bell_2022_prscs** | 0.00E+00 | 0.00E+00 | 0.00E+00 | -3.04E-01 | 0.00E+00 |
| **AD_EUR_Mars_2022_prscs** | 0.00E+00 | -4.22E-03 | 0.00E+00 | 0.00E+00 | 0.00E+00 |
| **EP_EAS_Sakaue_2021_prscs** | 0.00E+00 | 7.81E-02 | 0.00E+00 | -6.66E+00 | -5.55E+00 |
| **EP_EUR_ILAE_2023_prscs** | 0.00E+00 | 0.00E+00 | 0.00E+00 | 7.83E+00 | 7.49E+00 |
| **EP_FINN_2024_prscs** | 0.00E+00 | 5.24E-02 | 0.00E+00 | 0.00E+00 | 0.00E+00 |
| **EP_MULTI_ILAE_2023_prscs** | 0.00E+00 | 0.00E+00 | 5.92E-02 | 0.00E+00 | 0.00E+00 |
| **GGE_EUR_ILAE_2023_prscs** | -3.31E-01 | 0.00E+00 | -6.69E-02 | 0.00E+00 | 0.00E+00 |
| **GGE_MULTI_ILAE_2023_prscs** | -2.14E+00 | 0.00E+00 | 0.00E+00 | 0.00E+00 | 0.00E+00 |
| **CA_EAS_Sakaue_2021_prscs** | 0.00E+00 | 0.00E+00 | 3.19E-01 | 0.00E+00 | 0.00E+00 |
| **CA_EUR_Jiang_2021_prscs** | -3.65E+00 | 0.00E+00 | 1.53E-01 | 2.89E+00 | 1.88E+00 |
| **GT_EAS_Zhou_2022_prscs** | 0.00E+00 | 4.63E-02 | 3.61E-01 | 2.49E+01 | 2.49E+01 |
| **GT_EUR_Zhou_2022_prscs** | 2.64E+00 | 3.45E-01 | 4.97E-02 | 0.00E+00 | 0.00E+00 |
| **GT_Zhou_Zhou_meta_prscsx** | 0.00E+00 | 0.00E+00 | 6.62E-01 | 0.00E+00 | 0.00E+00 |
| **BP_EAS_Sakaue_2021_prscs** | 0.00E+00 | 0.00E+00 | 0.00E+00 | -6.52E+00 | -5.60E+00 |
| **BP_EUR_Jiang_2021_prscs** | 3.25E-12 | 2.31E-12 | 4.12E-12 | 1.10E-10 | 1.15E-10 |
| **OA_EUR_Tach_2019_prscs** | 0.00E+00 | 0.00E+00 | 0.00E+00 | 0.00E+00 | 0.00E+00 |
| **OA_MULTI_Hat_2025_prscs** | 0.00E+00 | 4.25E-01 | 5.05E-01 | 5.42E+01 | 5.44E+01 |

| | | | | | |
|---|---|---|---|---|---|
| **DEP_EAS_Sakaue_2021_prscs** | 2.98E+00 | 2.12E-01 | 0.00E+00 | 6.11E+00 | 5.62E+00 |
| **DEP_EUR_Meng_2024_prscs** | 0.00E+00 | 0.00E+00 | 0.00E+00 | 0.00E+00 | 0.00E+00 |
| **DEP_FINN_2024_prscs** | 0.00E+00 | 0.00E+00 | 1.04E-01 | 0.00E+00 | 0.00E+00 |
| **DEP_MULTI_Meng_2024_prscs** | 1.47E+01 | 8.72E-01 | 2.92E+00 | 9.49E+01 | 9.87E+01 |
| **ANX_EUR_Brasher_2023_prscs** | 0.00E+00 | 0.00E+00 | 5.93E-01 | 0.00E+00 | 0.00E+00 |
| **ANX_EUR_Forstner_2023_prscs** | 0.00E+00 | 0.00E+00 | 7.64E-02 | 3.47E+00 | 3.38E+00 |
| **SCH_EAS_Lam_2019_prscs** | 7.11E-01 | 0.00E+00 | 5.40E-05 | 1.98E+00 | 1.51E+00 |
| **SCH_EUR_Brasher_2023_prscs** | 3.62E-01 | 1.23E-01 | 2.61E-01 | 0.00E+00 | 0.00E+00 |
| **SCH_EUR_Jiang_2021_prscs** | 0.00E+00 | 0.00E+00 | 0.00E+00 | -2.24E-02 | -1.76E-02 |
| **SCH_Brasher_Lam_meta_prscsx** | 1.85E+00 | 2.80E-04 | 0.00E+00 | -3.97E+00 | -2.85E+00 |
| **ALC_EUR_Jiang_2021_prscs** | 0.00E+00 | 0.00E+00 | 0.00E+00 | 0.00E+00 | 0.00E+00 |
| **ALC_FINN_2024_prscs** | 0.00E+00 | 5.43E-02 | 6.24E-01 | 2.95E+00 | 3.36E+00 |
| **LD_EUR_Cordell_2021_prscs** | 0.00E+00 | 5.30E-08 | 3.88E-07 | 0.00E+00 | 0.00E+00 |
| **LD_EUR_Jiang_2021_prscs** | 0.00E+00 | 0.00E+00 | 0.00E+00 | 0.00E+00 | 0.00E+00 |
| **LD_FINN_2024_prscs** | 0.00E+00 | 0.00E+00 | 0.00E+00 | 1.41E+01 | 1.32E+01 |
| **HL_EAS_Sakaue_2021_prscs** | 0.00E+00 | 0.00E+00 | 3.72E-01 | 9.10E+00 | 8.73E+00 |
| **HL_EUR_Jiang_2021_prscs** | 0.00E+00 | 1.10E-01 | 1.19E-01 | 0.00E+00 | 0.00E+00 |
| **HL_MULTI_Verma_2024_prscs** | 0.00E+00 | 1.86E-01 | 3.37E-01 | 3.00E+01 | 2.98E+01 |
| **CKD_EAS_Sakaue_2021_prscs** | 1.75E+00 | 1.26E-01 | 0.00E+00 | -4.03E+00 | -3.09E+00 |
| **CKD_EUR_Jiang_2021_prscs** | 2.57E+00 | -8.45E-02 | -5.47E-01 | -1.71E+01 | -1.68E+01 |
| **CKD_MULTI_Sakaue_2021_prscs** | 0.00E+00 | 3.50E-01 | 6.40E-02 | -1.07E+01 | -7.67E+00 |
| **IBD_EAS_Liu_2023_prscs** | 0.00E+00 | 3.18E-02 | 0.00E+00 | 3.32E+00 | 3.10E+00 |
| **IBD_EUR_Zheng_2023_prscs** | 0.00E+00 | 0.00E+00 | 1.39E-02 | 4.98E-01 | 0.00E+00 |
| **HPC_EUR_Zhou_2018_prscs** | 1.64E+00 | 4.41E-02 | 0.00E+00 | 1.90E+00 | 1.56E+00 |
| **ST_EAS_Zhou_2022_prscs** | 0.00E+00 | 3.56E-02 | 6.53E-01 | 0.00E+00 | 0.00E+00 |
| **ST_EUR_Mishra_2022_prscs** | 0.00E+00 | 3.91E-01 | 0.00E+00 | 0.00E+00 | 0.00E+00 |
| **ST_Mishra_Zhou_meta_prscsx** | 5.85E+00 | 7.09E-01 | 4.22E-01 | 0.00E+00 | 0.00E+00 |
| **GERD_MULTI_Sakaue_2021_prscs** | 4.01E+00 | 8.30E-01 | 1.80E+00 | 8.26E+01 | 8.52E+01 |

**References**

1. 1000 Genomes Project Consortium, Auton A, Brooks LD, et al. A global reference for human genetic variation. *Nature* 2015; **526**(7571): 68–74.
2. Kuo T, Frith MC, Sese J, Horton P. EAGLE: Explicit alternative genome likelihood evaluator. *BMC Med Genomics* 2018; **11**: 28.
3. Fuchsberger C, Abecasis GR, Hinds DA. minimac2: faster genotype imputation. *Bioinformatics* 2015; **31**(5): 782–784.
